# Use of directed acyclic graphs (DAGs) in applied health research: review and recommendations

**DOI:** 10.1101/2019.12.20.19015511

**Authors:** Peter WG Tennant, Wendy J Harrison, Eleanor J Murray, Kellyn F Arnold, Laurie Berrie, Matthew P Fox, Sarah C Gadd, Claire Keeble, Lynsie R Ranker, Johannes Textor, Georgia D Tomova, Mark S Gilthorpe, George TH Ellison

## Abstract

**Background:** Directed acyclic graphs (DAGs) are an increasingly popular approach for identifying confounding variables that require adjustment when estimating causal effects. This review examined the use of DAGs in applied health research to inform recommendations for improving their transparency and utility in future research.

**Methods:** Original health research articles published during 1999-2017 mentioning “*directed acyclic graphs*” or similar or citing DAGitty were identified from Scopus, Web of Science, Medline, and Embase. Data were extracted on the reporting of: estimands, DAGs, and adjustment sets, alongside the characteristics of each article’s largest DAG.

**Results:** A total of 234 articles were identified that reported using DAGs. A fifth (n=48, 21%) reported their target estimand(s) and half (n=115, 48%) reported the adjustment set(s) implied by their DAG(s).

Two-thirds of the articles (n=144, 62%) made at least one DAG available. Diagrams varied in size but averaged 12 nodes (IQR: 9-16, range: 3-28) and 29 arcs (IQR: 19-42, range: 3-99). The median saturation (i.e. percentage of total possible arcs) was 46% (IQR: 31-67, range: 12-100). 37% (n=53) of the DAGs included unobserved variables, 17% (n=25) included super-nodes (i.e. nodes containing more than one variable, and a 34% (n=49) were arranged so the constituent arcs flowed in a consistent direction.

**Conclusions:** There is substantial variation in the use and reporting of DAGs in applied health research. Although this partly reflects their flexibility, it also highlight some potential areas for improvement. This review hence offers several recommendations to improve the reporting and use of DAGs in future research.

## INTRODUCTION

Estimating causal effects is a key aim of applied health research.^1^ One approach is to conduct a randomised controlled experiment, but practical and ethical constraints mean this is only possible for a limited range of exposures.^2^ Most causal effects must therefore be estimated from observational data; a notoriously difficult task that requires understanding, identifying, and attempting to address the many sources of bias that arise in non-experimental data, including confounding bias, selection bias, and information bias.^3^

Observational studies exploring causal effects are nevertheless extremely common in health and medical research, although their causal aims are rarely described explicitly.^4^ More often, observational studies adopt the language of ‘prediction’ or ‘association’ and report ‘independent’ associations or ‘predictors’ after adjustment for one or more other related variables, typically by including them as covariates in a multivariable regression model. Many approaches are available to assist with deciding which variables to adjust for from a list of potential candidates, including various theory-free statistical criteria and algorithms.^5^ Unfortunately, few of these conventional approaches explicitly consider the role of each variable in relation to the exposure and outcome; and it is often unclear why some variables were chosen for consideration and others not. Without this information, it is very difficult to relate the reported associations to any specific causal effects of interest.^6,7^

Causal inference approaches, such as the Potential Outcomes Framework, promote greater transparency by encouraging observational data scientists to formally define the causal effect(s) they seek (i.e. their ‘estimand’) before they begin their analysis.^8^ Estimating this effect is then informed by external knowledge of the data generating process, and by measuring and adjusting for all mutual causes of the exposure and outcome (i.e. confounders).^9,10^ However, since the true data generating process can never be known, it must be postulated from expert knowledge, relevant theory, and plausible assumptions. ^9,10^

Directed acyclic graphs (DAGs) provide a simple and transparent way for observational data scientists to identify and demonstrate their knowledge, theories, and assumptions about the causal relationships between variables.^11^ The implied adjustment set for accurately estimating a causal effect can then be deduced by inspection or algorithmically, depending on the DAG’s structure and complexity.^12–14^ Although the accuracy of the resulting estimate is contingent on how closely the DAG matches the (true) data generating process, the act of drawing and sharing a DAG makes these assumptions more explicit and open to scrutiny. Despite these benefits, DAGs are relatively rare within the wider setting of applied health research, likely due to lack of awareness and cultural ambivilance.^15,16^

There is also limited practical guidance available on the use and reporting of DAGs in applied research. In 2013, Sauer and VanderWeele offered a list of core considerations, but highlighted a need for a ‘disciplined approach to developing DAGs’.^17^ Ferguson et al (2019) responded by offering a structured protocol to aid with building DAGs,^18^ but there remains little advice on various practical considerations, including the reporting of: estimands of interest, DAGs, implied adjustment set(s) and results thereof, and the spatial arrangement of variables, and the justification for including or omitting dependencies.

This review aims to examine and evaluate the use of DAGs in applied health research to motivate some simple recommendations to improve the transparency and utility of DAGs in future observational research.

## METHODS

### Overview

The review sought to extract, examine, and summarise information on the use, implementation, and reporting of causal diagrams that satisfy the definition of a DAG, as provided below, in observational health research. We were primarily interested in reporting behaviours and technical features of DAG specification that could motivate subsequent recommendations. These included reporting of: estimands of interest, DAGs, implied adjustment sets, other adjustment set(s), and the estimates obtained from these adjustment sets. They also included: the size of each DAG, the inclusion of unobserved variables, the use of ‘**super-nodes**’ (i.e. nodes containing more than one variable), the assumptions and justifications for including or excluding causal relationships, and whether causal relationships were visually drawn in a consistent direction (e.g. top-to-bottom or left-to-right).

### Definitions

**DAGs** are typically non-parametric diagrammatic representations of the assumed data generating process for a set of variables (and measurements thereof) in a specified context. Variables and their measurements are depicted as **nodes** (or vertices) connected by unidirectional **arcs** (or arrows; hence ‘*directed*’) depicting the hypothesised relationships between them. An arc between two nodes denotes the assumed existence and direction of a causal relationship, but does not specify the sign, magnitude, or parametric form. A node cannot be caused by itself (hence ‘*acyclic*’), because no variable can cause itself at an instantaneous moment in time, and the future cannot cause the past.

A **path** is a collection of one or more arcs that connects two nodes. Paths may be either **open** or **closed**; open paths transmit statistical associations, closed paths do not. A **causal path** (or direct path) is one where all constituent arcs flow in the same direction from one node to another. The **total causal effect** of a specified **exposure** (i.e. cause) on a specified **outcome** (i.e. consequence), which together form the **focal relationship**, is the joint effect transmitted through all causal paths connecting the exposure to the outcome. With respect to the focal relationship, a **confounder** is a common cause of both the exposure and the outcome, a **mediator** is caused by the exposure and in turn causes the outcome (i.e. falls on a causal path between the exposure and outcome), and a **competing exposure** is a cause of the outcome that is neither caused by nor causes the exposure. A **direct causal effect** is the effect that does not act through one or more specified mediators. Figure 1 shows the main components of a DAG and the most common types of variable, defined in relation to the focal relationship.

**Figure 1.**
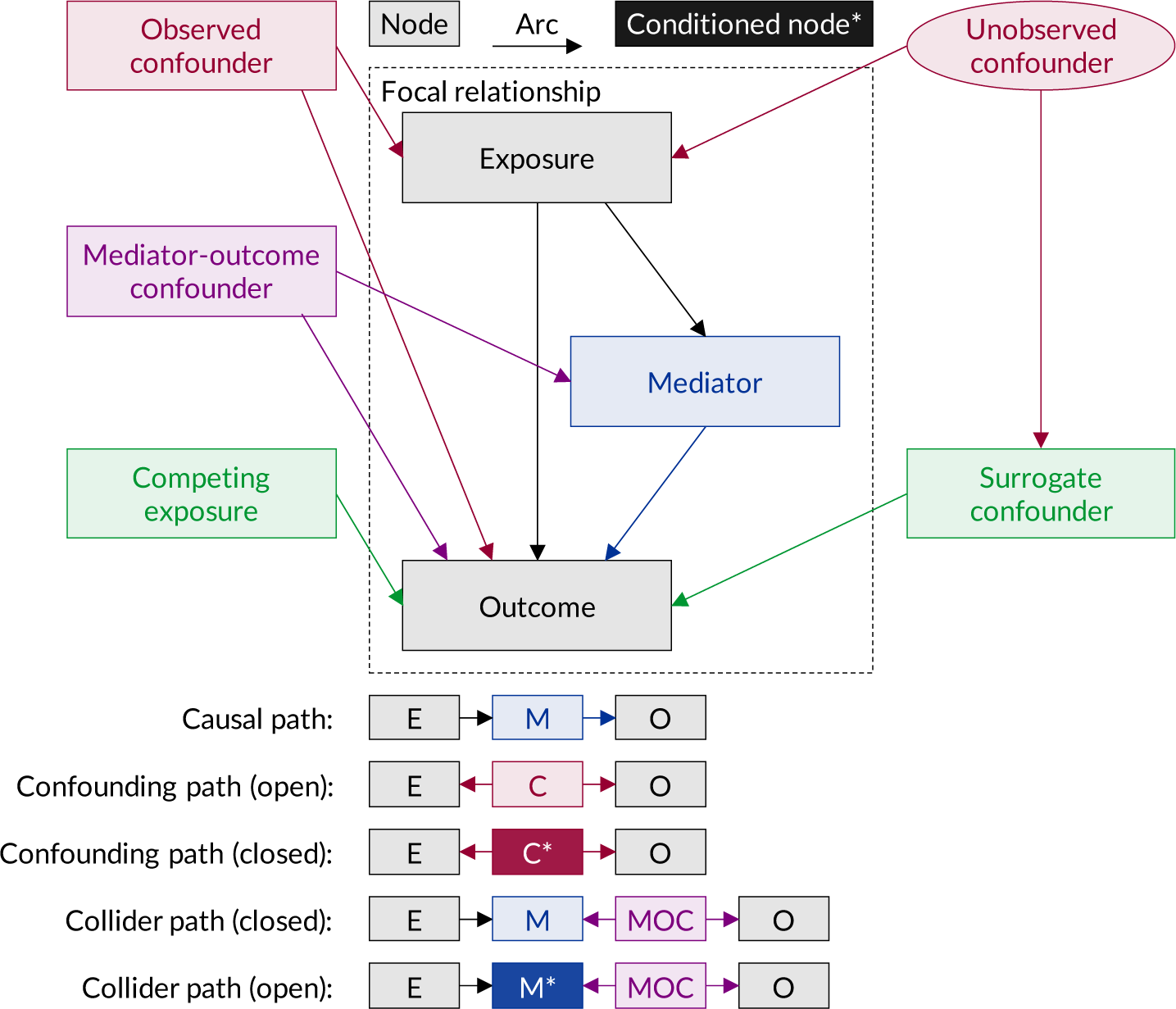
Illustration of the main components of a directed acyclic graph (DAG) and the most common types of contextual variable, defined in reference to the exposure-outcome relationship of interest.

A **confounding path** (or indirect path or backdoor path) between an exposure and an outcome is a path that passes against the flow of causality through one or more **confounders** (Figure 1). When estimating an **estimand** (i.e. the desired causal effect of the exposure on the outcome) from an association, open confounding paths introduce **confounding bias**. This bias can be reduced by closing (or blocking) the confounding path by adjusting for one or more of the nodes on that path, typically by including them as covariates in a multivariable regression model. A **collider path** between an exposure and an outcome is a path that passes against the flow of causality through one or more **colliders**, i.e. nodes that receive two or more arcs (Figure 1). Collider paths are closed and do not transmit statistical associations unless the constituent colliders (or any descendent thereof) are conditioned on. When estimating an estimand from an association, collider paths which have been opened by conditioning on the constituent colliders will introduce **collider bias**. This can occur when estimating direct causal effects if adjustment is made for a mediator that is caused by one or more common causes of the outcome (known as **mediator-outcome confounders**). This is because the mediator in this situation is also a collider on the non-causal path between the exposure and the outcome through the mediator-outcome confounder.^11^

A **sufficient adjustment set** for a particular estimand is any set of variables that, if fully conditioned on, will provide an unbiased estimate for that estimand by *closing* all confounding paths and leaving all causal paths *open*.^19^ Competing exposures are not always included in sufficient adjustment sets, unless they are caused by one or more unobserved confounders for which they can serve as a **surrogate confounder**.^20^

### Search and inclusion criteria

We searched Scopus, Web of Science, Medline, and Embase for articles published between 01 January 1999 and 31 December 2017 inclusive that contained any of the following terms in their title, abstract, keywords, or topic: *‘graphical model theory*’, ‘*directed acyclic graph(s)*’, ‘*causal diagram(s)*’, *‘causal graph(s)*’, or ‘*causal DAG*’. Articles citing DAGitty online or the DAGitty R software packages were also identified.^14,19^ Results were restricted to ‘original articles’ involving human participants (Medline and Embase) indexed within medicine, health, social science, psychology, dentistry, nursing, and related fields.

### Screening and exclusion criteria

Duplicates were identified and removed before PWGT screened the titles and abstracts to determine eligibility. Articles not describing original research (such as teaching articles, review articles and commentaries) were excluded, as were articles not examining health or healthcare in human populations (such as gene ontology and protein hierarchy studies).

### Extraction process

Data were extracted from each article into a standardised database, which was designed, tested, refined, piloted, and further refined before extraction. PWGT, KFA, MSG, WJH, LB, SCG, CK, and GTHE were each assigned a random sample of articles for data extraction. Where information was unclear, individual members of the study team consulted with one or more other members to reach consensus on the appropriate coding. All data were then re-extracted and double-checked by PWGT, GDT, and GTHE. Discordances between the original data extraction and the subsequent re-extraction were recorded and considered preliminary ‘errors’ in data extraction.

### Data of interest

Bibliographic information including author names, journal names, and year of publication were obtained from bibliographic records. The topic(s) of each article was approximated from the indexing categories of the host journal in the 2017 Journal Citation Reports from Clarivate Analytics (https://jcr.clarivate.com/).

Data were extracted on the country of the lead author’s primary affiliation, the number of causal diagrams used and their availability within the manuscript/supplementary materials, and the analytical approach used (noting where mediation analyses or other less conventional methods were used). For the largest diagram available for each article (assessed by number of nodes; or number of arcs if tied), we extracted data on the total number of nodes and total number of arcs, and whether: they were drawn with DAGitty, any unobserved variables were included, any super-nodes were included,^21^ the arcs were drawn to flow in a consistent direction (e.g. left-to-right or top-to-bottom), the diagram included all possible arcs (i.e. was ‘saturated’ or ‘complete’), and citations were used to justify the inclusion or exclusion of arcs. To identify the estimand(s) of interest, we searched and examined occurrences of “*estimand*”, “*effect*”, “*caus*\*”, “*total*”, “*direct*”, “*indirect*”. We inspected whether the implied (sufficient) adjustment set was reported for each estimand or apparent effect of interest, and whether estimates were reported for these sets explicitly. We also examined whether alternative adjustments sets were used and, if so, what approaches were used for covariate selection.

### Errors in DAG data extraction

The probability of ‘error’ in the initially-extracted number of nodes and arcs was explored in relation to the most common diagrammatic features (whether drawn in DAGitty, whether unobserved variables or super-nodes were included, and whether arcs were drawn in a consistent direction), adjusted for number of arcs and nodes by log-linear regression with robust standard errors. Reported probabilities represent model marginal values, with 95% confidence intervals (CI) approximated by the delta-method.

## RESULTS

### Sample description

Figure 2 summarises the derivation of the study sample. A total of 234 eligible articles were identified, including 172 (73.5%) published since the start of 2015 (Figure 3). 230 (98%) articles were written in English and four (2%) were written in German. Brief details of each paper are provided in Supplementary Table 1.

**Figure 2.**
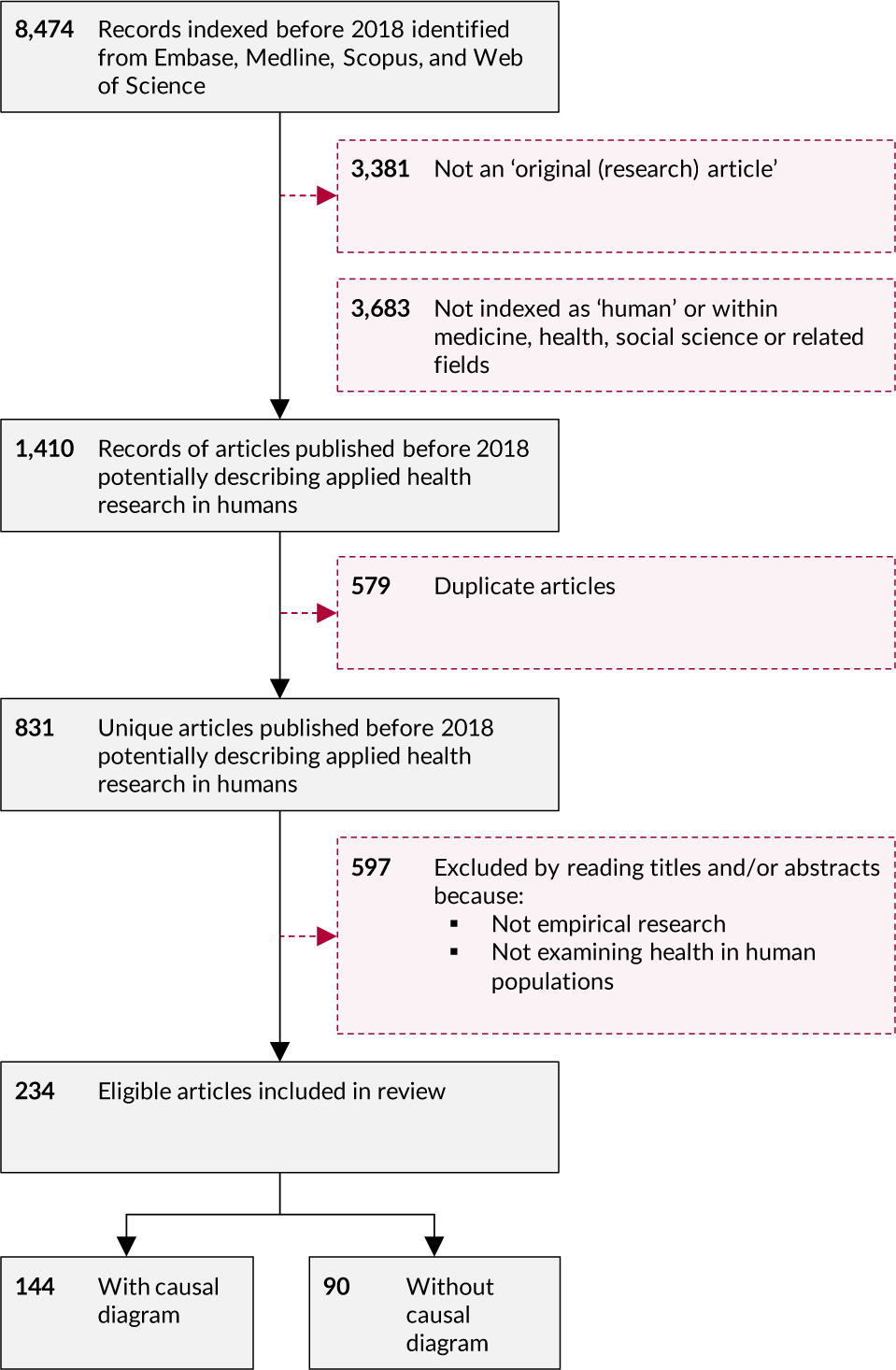
Flow of bibliographic records into the final sample of 234 articles

**Figure 3.**
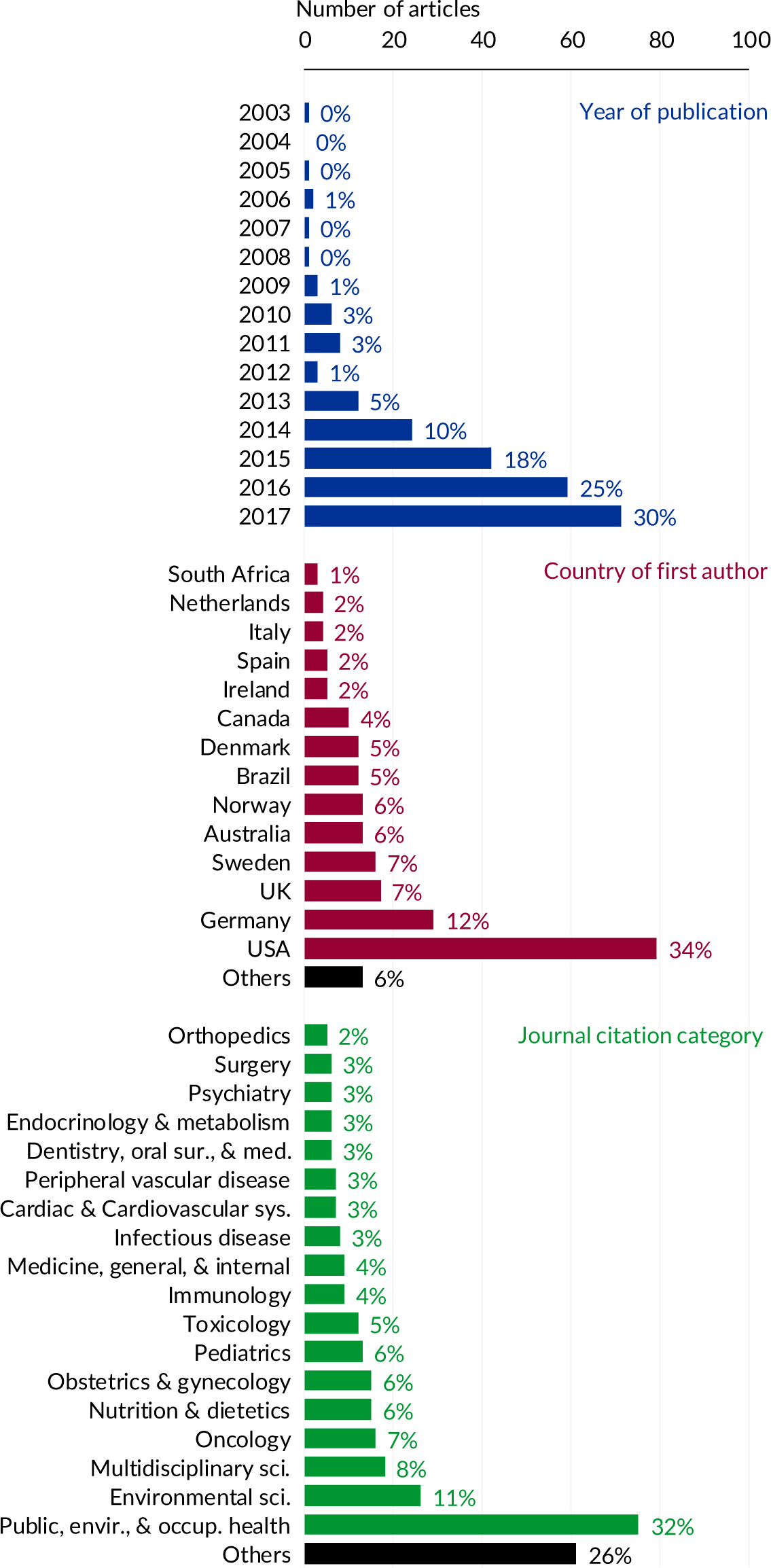
Distribution of the 234 articles included in the review sample, by year of publication, country of first author’s primary affiliation, and journal citation category.

Figure 3 shows the number of articles published by the country of the first author’s primary affiliation. The most prevalent countries were USA (n=79, 34%), Germany (n=29, 12%), UK (n=17, 7%), and Sweden (n=16, 7%). 187 (80%) were led by North American or Northern European authors.

Articles were published in 152 distinct journals. 33 (22%) journals published more than one article, the most appearing in *PLoS One* (n=18), *Environmental Health Perspectives* (n=10), and *Environmental Research* (n=8). These 152 journals covered 50 citation categories, the most prevalent being *Public, Environmental, and Occupational Health* (n=75, 32%), *Environmental Science* (n=26, 11%), and *Multidisciplinary Science* (n=18, 8%) (Figure 3).

### DAG availability, size, and attributes

Causal diagrams were available for 144 (62%) articles. Sixty (42% of those with one or more diagrams available) included the diagram in the manuscript directly, 81 (56%) in supplementary online material, one (<1%) provided the diagram on request (Bliddal et al 2016)^22^ one (<1%) provided a link to the diagram on *www.dagitty.net* (Evandt et al 2017A),^23^ and one (<1%) referred to a diagram from a previous publication (Buchner & Rehfuess 2015).^24^ No diagram was available for 90 (38%) articles, including four (1%) articles that referred to supplementary online materials that were unavailable, one article where the diagram was missing from supplementary material, one (<1%) where the printed figure was incorrect, and one (<1%) that provided invalid weblinks to *www.dagitty.net*.

Of the 144 articles with available diagrams, 116 (81%) included a single diagram and 28 (19%) more than one. Full details of the largest diagram in each of these 144 articles are provided in Supplementary Table 2, and summary details are in Table 1. Diagrams varied substantially in size and complexity, with the number of nodes ranging from 3 to 28 (Median=12, IQR=9-16) and the number of arcs ranging from 3 to 99 (Median=29, IQR=19-42). The median ratio of arcs to nodes was 2.3 (IQR=1.8-3.0, Range=1.0-5.8) and the median saturation (i.e. percentage of total possible arcs) was 46% (IQR=31-67, Range 12-100). Just four (3%) diagrams included all possible arcs (i.e. were ‘saturated’ or ‘complete’).

**Table 1.** Summary information regarding the reporting of estimands and adjustment sets in the 234 included studies, and regarding the reporting and features of the largest diagram in the 144 studies with one or more directed acyclic graph (DAGs).

| **DAG reporting and features<sup>a</sup>** | **N** | **% (N=144)** |  |
| --- | --- | --- | --- |
| DAG available | 144 | 100% |  |
| *Single DAG available* | 116 | 81% |  |
| *Multiple DAGs available* | 28 | 19% |  |
| DAG includes one or more unobserved variables | 53 | 37% |  |
| *DAG includes<sup>1</sup> one or more specific unobserved variables* | 27 | 19% |  |
| *DAG includes<sup>1</sup> one or more generic unobserved variables* | 29 | 20% |  |
| Arcs flow in consistent direction | 49 | 34% |  |
| *Top-to-bottom* | 5 | 3% |  |
| *Left-to-right* | 22 | 15% |  |
| *Corner-to-corner* | 22 | 15% |  |
| Authors provide citations for one or more arcs | 8 | 6% |  |
| **DAG nodes and arcs** | **Median** | **IQR** | **Range** |
| Number of nodes | 12 | 9-16 | 3-28 |
| Number of arcs | 29 | 19-42 | 3-99 |
| Ratio of arcs-to-nodes | 2.3 | 1.8-3.0 | 1.0-5.8 |
| Saturation (%)<sup>b</sup> | 46 | 31-67 | 12-100 |
| **Reporting of estimand(s) and adjustment set(s)** | **N** | **% (N=234)** |  |
| Report one or more estimand(s) of interest | 48 | 21% |  |
| *Report seeking total causal effects* | 18 | 8% |  |
| *Report seeking direct causal effects* | 12 | 5% |  |
| *Report seeking multiple effects* | 18 | 8% |  |
| Report DAG-implied adjustment set(s) | 115 | 49% |  |
| Report results of DAG-implied adjustment set(s) | 101 | 43% |  |
| *Report as primary results* | 95 | 41% |  |
| Report results of other or unclear adjustment set(s) | 171 | 73% |  |
| *Report as primary results* | 159 | 68% |  |
| Use additional statistical criteria for variable selection | 42 | 18% |  |

<sup>a</sup>Details are for the largest diagram reported in each study <sup>b</sup>The saturation percentage represents the proportion of all possible arcs that have been included.

Fifty-three (37%) diagrams included one or more unobserved variable (see <u>Example 1</u> and <u>Example 2</u>); 27 (19%) included one or more specific unobserved variables and 29 (20%) included one or more generic unobserved variables. Twenty-five (17%) diagrams included one or more super-nodes (See <u>Example 3</u>). Forth-nine (34%) diagrams were arranged so that the constituent arcs flowed in a consistent direction (see <u>Example 4</u>, and <u>Example 5</u> for contrast); five (3%) flowed from top-to-bottom, 22 (15%) flowed from left-to-right, and 22 (15%) flowed diagonally from one corner to another. Eight (6%) provided citations to support the inclusion of one or more arcs (see <u>Example 6</u>).

### Errors in DAG data extraction

First round data extraction errors occurred among 56 (39%) DAGs; including two (1%) where the number of nodes were miscounted, 48 (33%) where the number of arcs were miscounted, and six (4%) where both were miscounted. The proportion of errors increased with the number of nodes [4.2% per node (95% CI: 1.5, 7.0)] or arcs [1.6% per arc (95% CI: 1.0, 2.2)]. Conditional on these, the proportion of errors was lower among diagrams drawn in DAGitty [32% (95% CI: 23, 42) vs 50% (95% CI: 36, 64) among diagrams not drawn in DAGitty] or where the arcs flowed consistently from top-to-bottom or left-to-right [30% (12, 47) vs 41% (95% CI: 31, 51) among diagrams without consistent direction] but not where arcs flowed diagonally from corner-to-corner [44% (95% CI: 20, 67)] (Supplementary Table 3). There were negligible differences in the proportion of errors among diagrams that included unobserved variables [42% (95% CI: 28, 56) vs 37% (95% CI: 31, 51) without unobserved variables] or super-nodes [39% (95% CI: 17, 61) vs 39% (95% CI: 31, 48) without super-nodes] (Supplementary Table 3).

### Reporting of estimands and adjustment sets

Of the 234 included articles, 208 (89%) conducted multivariable regression analyses, 13 (6%) conducted mediation analyses, 4 (2%) conducted g-method analyses, and 9 (2%) conducted other or mixed analyses.

Full details regarding the reporting of estimands and adjustment sets are provided in Supplementary Table 4, while summary details are in Table 1. Only 48 (21%) articles explicitly reported one or more causal effect estimand of interest, comprising 18 (8%) that sought total causal effects, 12 (5%) that sought direct causal effects, and 18 (8%) that sought multiple effects. 115 (49%) articles clearly reported the adjustment set(s) implied by their DAG(s), including 90 (38%) who specifically stated one or more sufficient adjustment set.

Of the 115 articles that reported one or more DAG-implied adjustment set(s), 101 (88%) reported the causal effect estimate obtained from this adjustment set specifically (i.e. with no covariates added or removed) including 95 (76%) where these formed the primary results. Estimates from other (i.e. bespoke) or unclearly derived adjustment sets were reported in 171 (73%) of the total 234 articles, including 159 (68%) where these formed the primary results.

Finally, 42 (18%) studies applied additional statistical criteria or algorithms for covariate selection, including 28 (12%) that used change-in-estimate criteria and 14 (6%) that used p-value criteria.

## DISCUSSION

### Summary of findings

This review examined the use of DAGs in applied health research. Although they remain very rare within the wider literature, an increasing absolute number of observational studies in health, medicine, and related areas are using and presenting causal diagrams to aid confounder selection. There is however substantial variation in the way these diagrams are reported and used, with potential impacts on the study results and the contributions towards increasing transparency and scrutiny of the analytical assumptions.

DAGs were not always presented, and in some instances very little information was provided regarding their construction or use (see Supplementary Table 4 and Supplementary Table 5). Where DAGs were presented, they varied substantially in the number of variables and arcs included, the inclusion of unobserved variables, the inclusion of super-nodes, details on their design, and how they were used to inform the subsequent statistical modelling. Some studies presented simplified illustrations of the relationships between only a few variables considered key to the focal relationship (e.g. Wei et al 2016)^25^, others presented large schematics with multiple interconnected measured variables (e.g. Ng et al 2017A)^26^, but a minority incorporated many, if any, unobserved variables. In general, diagrams included relatively few arcs per node and very few were ‘saturated’ or ‘complete’, where each variable in turn is assumed to (potentially) cause all future variables.

Only half of the articles reported the adjustment set(s) implied by their DAG(s), and only a fifth stated their target estimands, with most instead referring to the ‘association’, ‘relationship’, or ‘effect’. Several studies used additional statistical criteria or algorithms to reduce the number of candidate variables, while others made bespoke modifications to their adjustment set(s) for other subjective reasons not implied by their DAG(s) (e.g. “*An additional set of covariates was considered important to include in the full model because of their well-established association with exposure and outcome*” (Weyde et al 2017A)^27^ (see Supplementary Table 4 and Supplementary Table 5).

### Observations and remarks

This review explicitly considered the use of DAGs in applied health research, where they were used not simply to discuss causal inference theory but to ‘*identify variables necessary for adjustment*’ (Upson et al 2013).^28^ Although such applications are widely marketed as a key benefit of DAGs, it is contested whether they actually lead to more accurate and/or reliable effect estimates.^15,16^

Harder to dispute are the potential benefits for transparency. Compared with traditional approaches to selecting candidate variables and adjustment sets, DAGs encourage observational data scientists to declare their assumptions about the data generating process for the data they are analysing, which clearly facilitates external scrutiny. Reporting the estimand(s) of interest and DAG-implied adjustment set(s) offer similar benefits to the clarity of the study aim(s) and the model interpretability.

Unfortunately, many of the articles reviewed did not report their DAG(s) and fewer still declared their estimand(s) of interest. Reluctance to share such details may reflect a lack of awareness of the benefits of these details for readers, lack of confidence, fear of criticism, cultural reticence, and/or a lack of encouragement or facilitation from journals, editors, and reviewers. These may also explain why some articles adopted hybrid approaches to variable selection that combined DAG-based approaches with traditional statistical approaches, or where investigators overruled the DAG-implied adjustment set(s) by adding or removing covariates. Such actions suggest a need for practical guidance and supporting materials that are accessible to data analysts, journal editors and reviewers. Regardless, several recommendations emerge from recognising the best practices among the studies reviewed (see below).

Further benefits may also be achieved by emphasising the different assumptions made from the inclusion and omission of arcs within DAGs. Although a small number of articles reported ‘saturated’ DAGs (i.e. where preceding variables are assumed to potentially cause *all* future variables) the majority of the available diagrams included less than half of all possible arcs. Since a confounder must cause both the exposure and outcome, inadvertently omitting either of these arcs may result in a confounder being mislabelled and omitted from the DAG-implied adjustment set, leading to unadjusted confounding. Such omissions may be explained by a cultural aversion to declaring potential causal relationships in observational data without a strong theoretical basis or definitive empirical evidence.^4^ This may also explain why some authors included citations to justify the inclusion of arcs within their diagrams. This is arguably unnecessary since *omitting* an arc invokes the greater assumption, because no arc between two variables declares there is *no* causal relationship between the two variables, regardless of direction, sign, strength, or parametric form.^11^ Increasing the number of arcs does however bring practical challenges that may also explain the absence of plausible arcs. In general, it is difficult to draw causal diagrams with multiple nodes and multiple arcs; both to ensure that all potential arcs have been included and that the diagram remains decipherable to the reader. We observed that the proportion of data extraction errors increased with the number of nodes and arcs, although this could also result from an increasing number of objects available for miscounting. Subjectively, we judged that DAGs that had been drawn so that their arcs flowed consistently from left-to-right or top-to-bottom were easier to interpret and critique. This appears to be supported by the lower proportion of data extraction errors among DAGs that had been arranged in either of these ways. Several algorithms exist to help with arranging a DAG into strict temporal ‘layers’,^29^ but they do not appear to have been widely adopted in applied settings. Alternatively, for ‘saturated’ or ‘complete’ DAGs, it may be reasonable to simplify several details (e.g. using paired arrow-heads indicating ‘arcs to all future nodes’)^30^ because the sufficient adjustment set for any exposure-outcome relationship will simply include all variables that temporally precede the exposure.

### Recommendations

#### 1) The focal relationship(s) and estimand(s) of interest should be stated in the study aims

Causal inference methods separate the process of identifying the quantity of interest (i.e. estimand) from the process of estimating that quantity, with the latter being informed by the former. To reflect this, all estimands of interest should be clearly stated in the study aims or at the beginning of the article’s methods section before details on how estimation was attempted (e.g. this study aimed to estimate the total causal effect of type-2 diabetes at aged 50-years on the ten-year risk of myocardial infarction).

#### 2) The causal diagram(s) for each focal relationship and estimand of interest should be available

Causal diagrams explicitly depict the investigators’ assumptions about the data generating process of their dataset. The accuracy of any ensuing effect estimate is fundamentally contingent on the extent to which the diagram (as specified) reflects the true data-generating process. The diagram used to inform the model for estimating every causal effect estimate should therefore be available to all potential readers. This may be achieved by reproducing the diagram in the manuscript directly, in supplementary material, or by providing functional weblinks to a well-established open-source platform such as *www.dagitty.net*.

#### 3) Causal diagrams should include all relevant variables, including those where direct measurements are unavailable

The causal diagram for a specific focal relationship should include all *potential* confounding variables (i.e. that may *potentially* cause both the exposure and the outcome), regardless of whether direct measurements are available or possible. Explicitly depicting unobserved variables helps to highlight potential sources of unobserved confounding.

#### 4) Variables should be arranged so that arcs flow in a consistent direction

Arcs depict causal processes that occur over time. Causal diagrams, and the relationships they symbolise, are therefore considerably easier to interpret when the constituent variables are arranged spatially in a way that clearly reflects the passage of time, with arcs flowing consistently from left-to-right or top-to-bottom.

#### 5) Arcs should generally be assumed to exist between any two variables

Omitting an arc between two variables implies that there is precisely *no* causal effect of one on the other. This is a much stronger statistical assumption than is implied when an arc is included, which allows a causal effect of any sign, magnitude, or parametric form (including a very small effect). Omitted arcs should therefore be carefully considered and ideally justified with theory and/or evidence.

#### 6) The DAG-implied adjustment set(s) for the estimand(s) of interest should be clearly stated

The sufficient adjustment set(s) implied by a causal diagram to estimate every estimand of interest should be stated explicitly, including variables for which measurements are not available for adjustment.

#### 7) The estimate(s) obtained from using the unmodified DAG-implied adjustment set(s) should be reported

The estimate(s) obtained from using the unmodified DAG-implied sufficient adjustment set(s) should always be reported, even if not considered or interpreted as central to the study findings.

#### 8) Alternative adjustment set(s) should be justified and their estimate(s) reported separately

If alternative adjustment set(s) are used, they should be clearly described and justified, and the ensuing estimates should be reported separately to those reported using the DAG-implied adjust set. Modifications to DAG-implied adjustment sets may comprise the inclusion of competing exposures, surrogate confounders, or variables with ambiguous causal roles for the purposes of sensitivity analyses. Where alternatives DAGs are hypothesised, it may be helpful to evaluate their consistency with the observed data.^19^ Where direct causal effects are sought, these should be clearly stated in the aims as effects of interest, and their adjustment sets derived accordingly.

### Strength and weaknesses

This is the largest and most comprehensive review of the use of DAGs in applied research, whether in health, medicine, or otherwise. We identified and examined in excess of 200 articles published over several years across a diverse range of fields. The sample is however somewhat ill-defined and should not be considered to reflect the population of observational health research studies that have used DAGs. It is not clear what proportion of such studies mention their use of DAGs in their abstract or keywords, but we suspect many do not. A large proportion of the sample was therefore identified from having cited the DAGitty.net software package. Studies that used DAGitty.net for drawing their DAG(s) or determining their adjustment set(s) are likely over-represented. A lower proportion of data extraction errors occurred among DAGs that had been drawn using DAGitty, which may indicate some feature differences, but it may also indicate a training effect due to increased reviewer familiarity. We believe that the issues we describe, and their implications, are likely to be applicable to the use and reporting of causal diagrams irrespective of the tools used to construct and scrutinise these DAG(s).

Extracting data for the 234 articles included in this review was laborious and a substantial time therefore passed between the sample was identified and extraction was complete. We nevertheless decided not to update our search to include more recent articles (i.e. published from 2018 onwards) because we do not believe there will have been any substantial changes in practice in the intervening time.

Because of the quantity of information sought and the diversity of the studies examined, it was impossible to develop a data-extraction form that was entirely compatible with all the included articles. Some data items therefore required subjective judgement, inter-rater discussion, and/or further simplification, and others simply could not be synthesised due to a lack of standardisation in reporting. Even when using a reduced set of items for data extraction, clarity and transparency varied substantially between studies making it challenging to accurately identify all relevant information. All data was therefore extracted in triplicate, with the first extraction used to evaluate the ‘readability’ of the most objective diagram features. Occasional data extraction errors or discordances between the authors intended message and our interpretation are nevertheless still inevitable; but these should not materially alter the review results, messages, and/or recommendations.

We offer several recommendations for improving the reporting, specification, and application of DAGs in applied health research where causal effect estimates are sought from observational data. Studies that have not followed these practices should not however be considered necessarily less rigorous, less accurate, or less valuable. DAGs represent the investigators’ assumptions and hypotheses about the data generating process for a specific dataset. Where the DAG does not accurately reflect the true data generating mechanism, the ensuing estimates are likely to be unreliable. However, since the true process can never be known, a DAG can arguably only be wrong if it fails to correctly represent those assumptions and hypotheses. We therefore did not attempt to evaluate the plausibility of individual DAGs and instead focussed on identifying those areas where the implications of the DAG may not have met the investigators’ intentions. We offer no negative judgements on the intentions of individual authors or the veracity of individual studies. On the contrary, we welcome the large and growing number of applied health researchers who have used DAGs to assist with estimating causal effects in observational data and explored their benefits for declaring their assumptions, identifying potential sources of bias, identifying data for collection, and improving statistical analyses. These ‘early adopters’ have not only helped to reveal some potential pitfalls in the use of DAGs but have provided a growing wealth of innovative exemplars that will inspire future developments in this evolving field.

## Conclusion

DAGs are increasingly popular in applied health research as a transparent means of identifying confounding variables that require adjustment to estimate causal effects. This review examined their use in over 200 empirical studies of health, medicine, and related disciplines, and found substantial variation in their size, structure, complexity, availability, and implementation. While such variety partly reflects the inherent flexibility and subjectivity of DAGs, it also helps to highlight several potential pitfalls and aspirational practices. Consequently, we offer a list of simple recommendations for improving both the transparency and benefits of DAGs in observational research that we hope will help towards the ongoing development of these techniques.

## Data Availability

Data provided directly in supplementary material

## Author Contributions

GTHE conceived the project and with PWGT, MSG, and JT designed the study. PWGT, KFA, MSG, WJH, LB, SCG, CK, GDT, and GTHE were involved in the data extraction. All authors had access to the data and were involved in the interpretation and analysis. PWGT was chiefly responsible for summarising and reporting the data and accepts full responsibility for the conduct and accuracy of the work. PWGT drafted the manuscript and all authors contributed to critically reviewing and revising the draft. All authors read and approved the final manuscript before submission and agreed with the decision to submit to MedRXiv.

## Funding

This study received no specific funding. KFA and SCG are grateful for PhD funding from the Economic and Social Research Council, UK (ES/J500215/1 and ES/P000746/1, respectively). LB is grateful for PhD funding from the Medical Research Council, UK (MR/K501402/1). GDT is grateful for PhD funding from The Alan Turing Institute [EP/N510129/1]. MSG and PWGT are both supported by The Alan Turing Institute [EP/N510129/1].

## SUPPLEMENTARY FIGURES AND TABLES

**Supplementary Figure 1**

*Example 1 (Boyle et al 2015)*

Showing a DAG where several unobserved variables have been included; here denoted with the letter U. This helps to highlight potentially important sources of unobserved confounding.

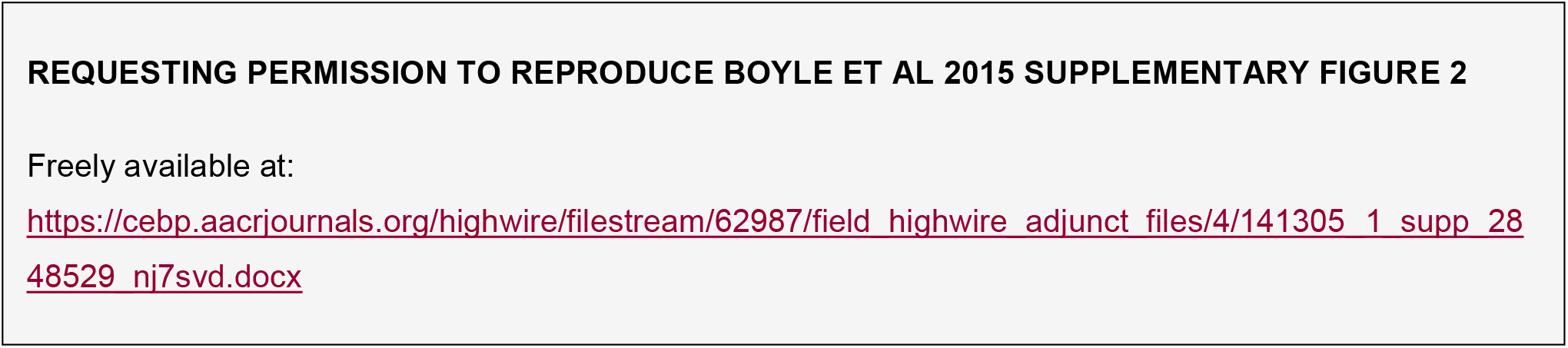

*Example 2 (Hinkle et al 2013)*

Showing a DAG where a (single) generic variable has been included to represent a cluster of unobserved variables; here denoted using Structural Equation Modelling notation with an ellipse. This helps to emphasise the collective influence of these unobserved variables but does less to indicate which of the underlying variables are thought most important.

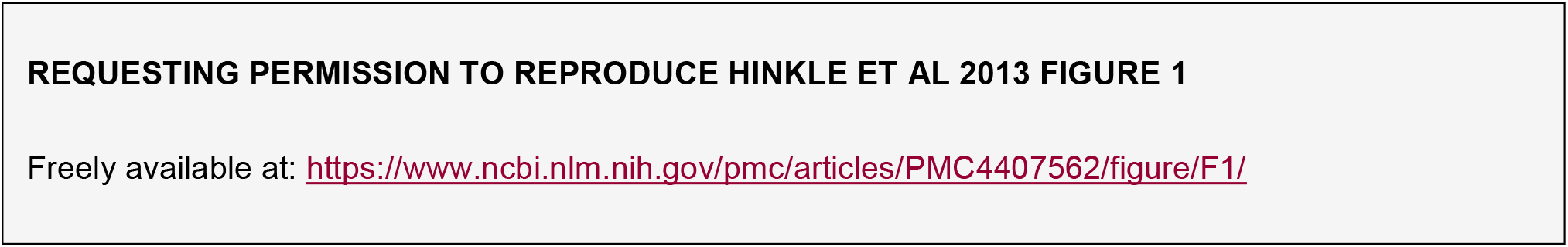

*Example 3 (Asgari et al 2011)*

Showing a DAG that includes super-nodes, i.e. nodes that contain more than one variable (e.g. in this diagram the Demographics node contains Gender, Age, and Race/ethnicity). This makes the diagram easier to interpret but increases the potential risk of misspecification.

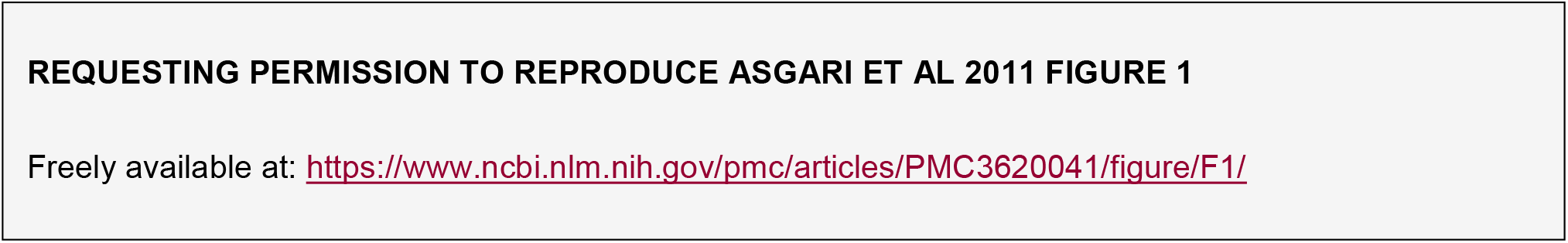

*Example 4 (Reiner et al 2016)*

Showing a DAG, which has been arranged so the constituent arcs flow consistently from top-to-bottom. Diagrams arranged in this manner are easier to interpret.

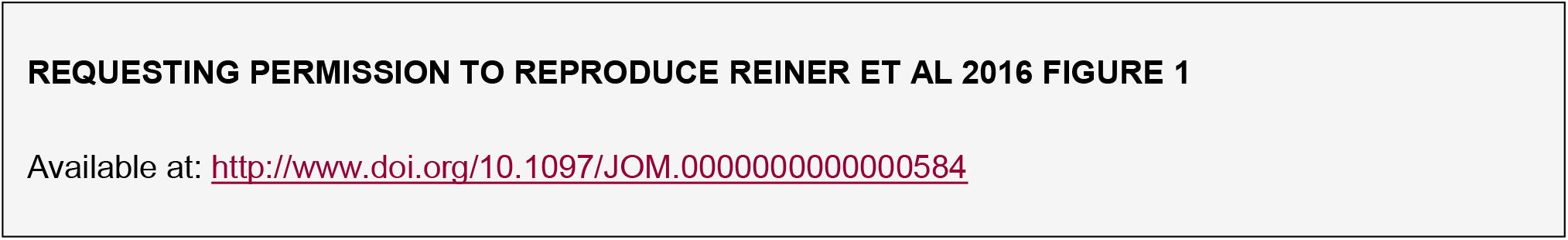

*Example 5 (Gocke et al 2014)*

Showing a DAG where the arcs do not flow in a consistent direction. This has no impact on the plausibility or validity of the diagram, but it does make it harder to interpret.

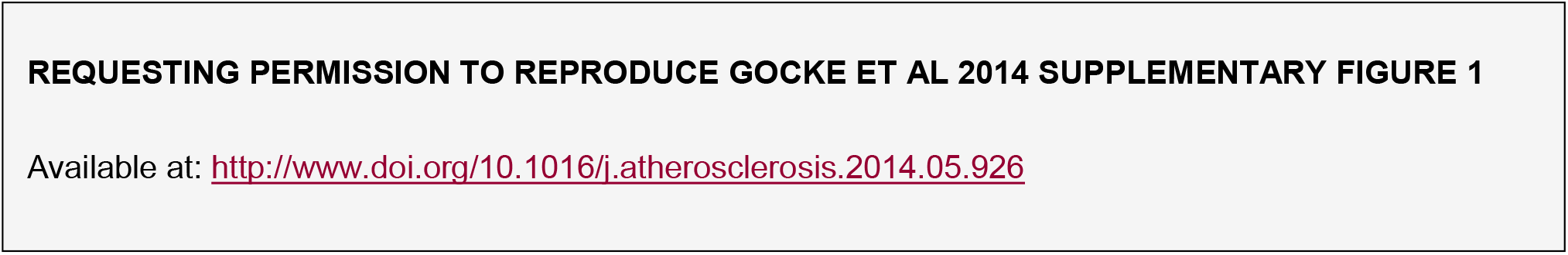

*Example 6 (Grill et al 2014)*

Showing a DAG where references to previous empirical research have been added to individual arcs to justify their inclusion.

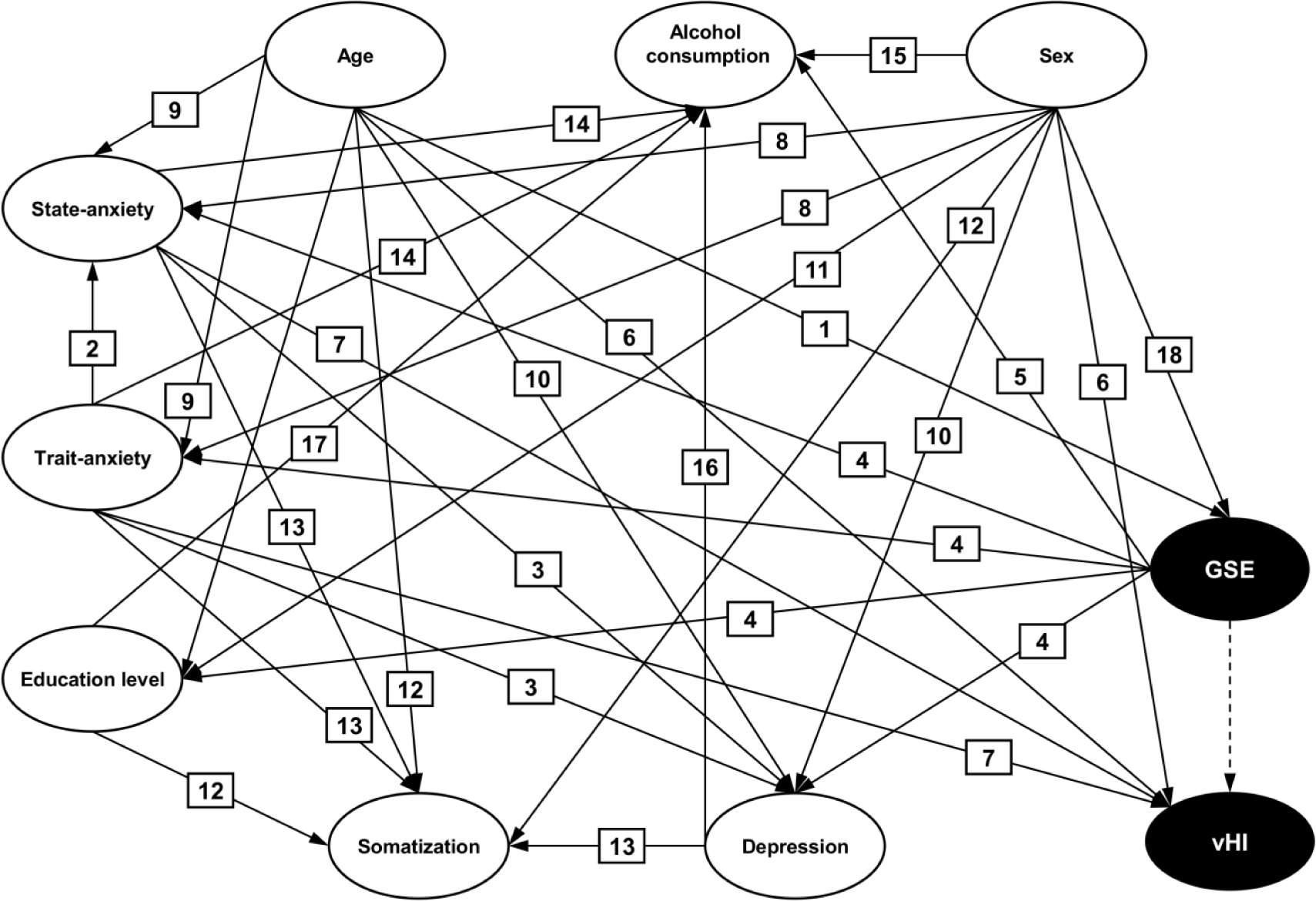

**REPRODUCED FROM:** Grill E, Schäffler F, Huppert D, Müller M, Kapfhammer H-P, Brandt T (2014) Self-Efficacy Beliefs Are Associated with Visual Height Intolerance: A Cross-Sectional Survey. PLoS ONE 9(12): e116220. https://doi.org/10.1371/journal.pone.0116220

**Supplementary Table 1.**
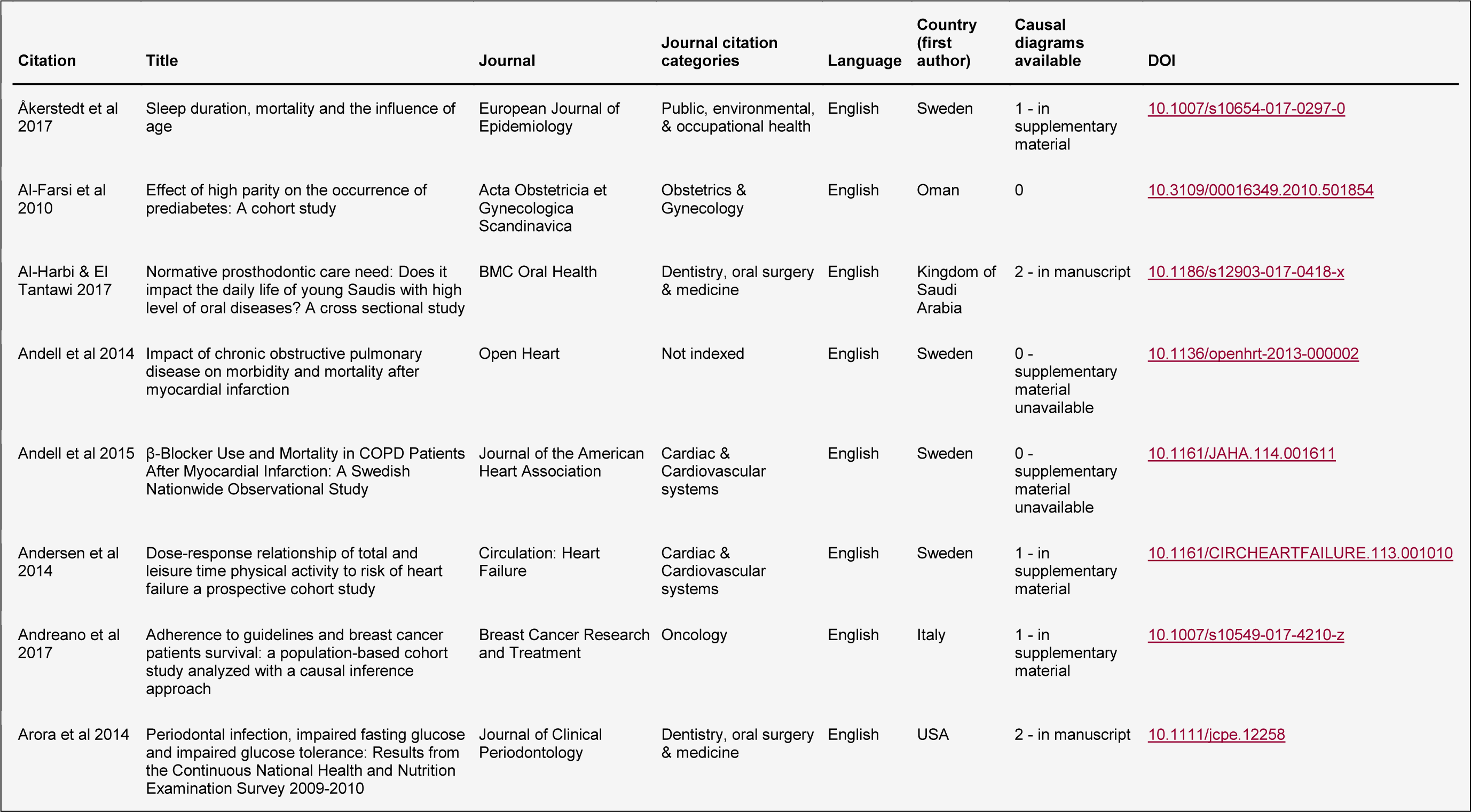
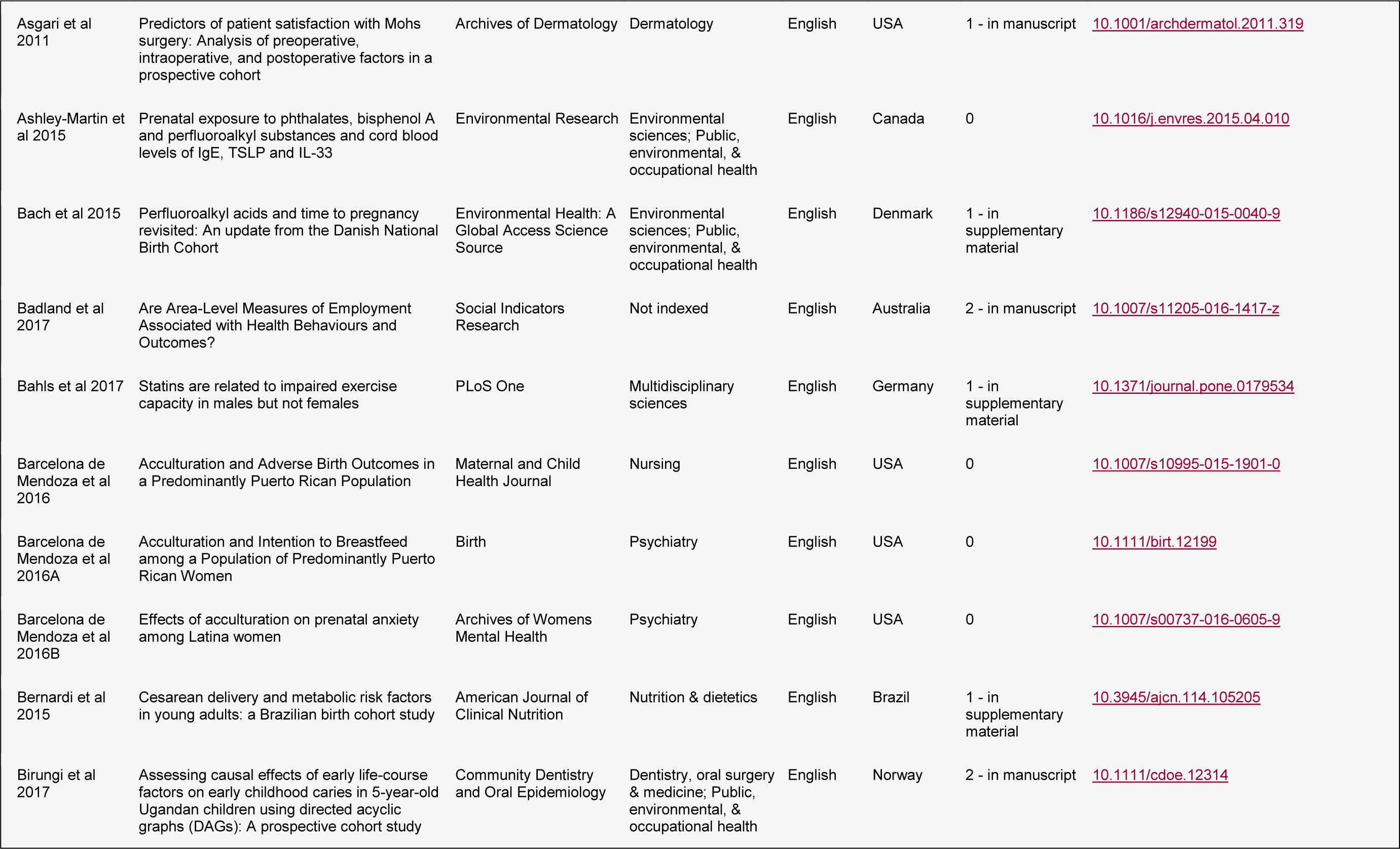
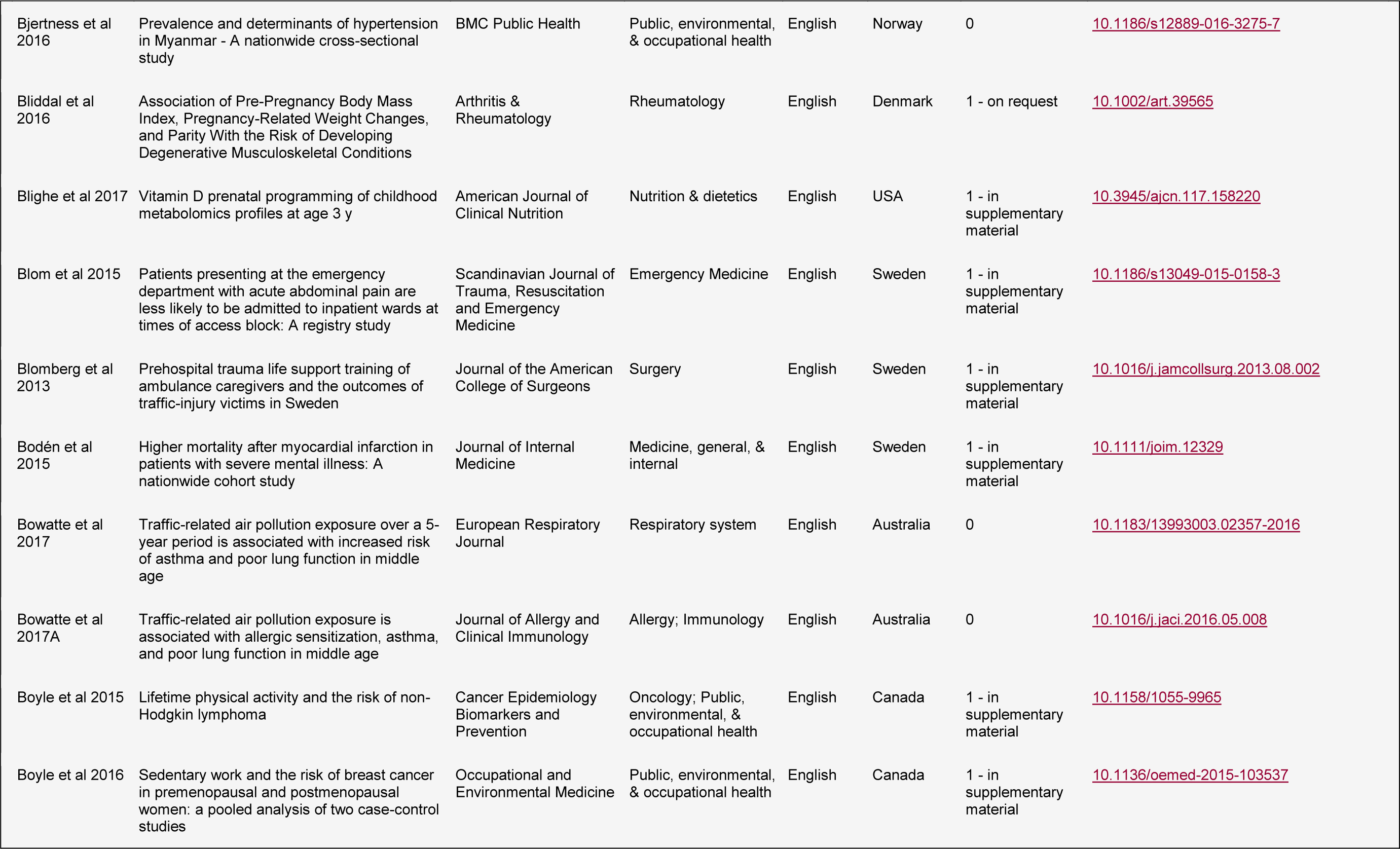
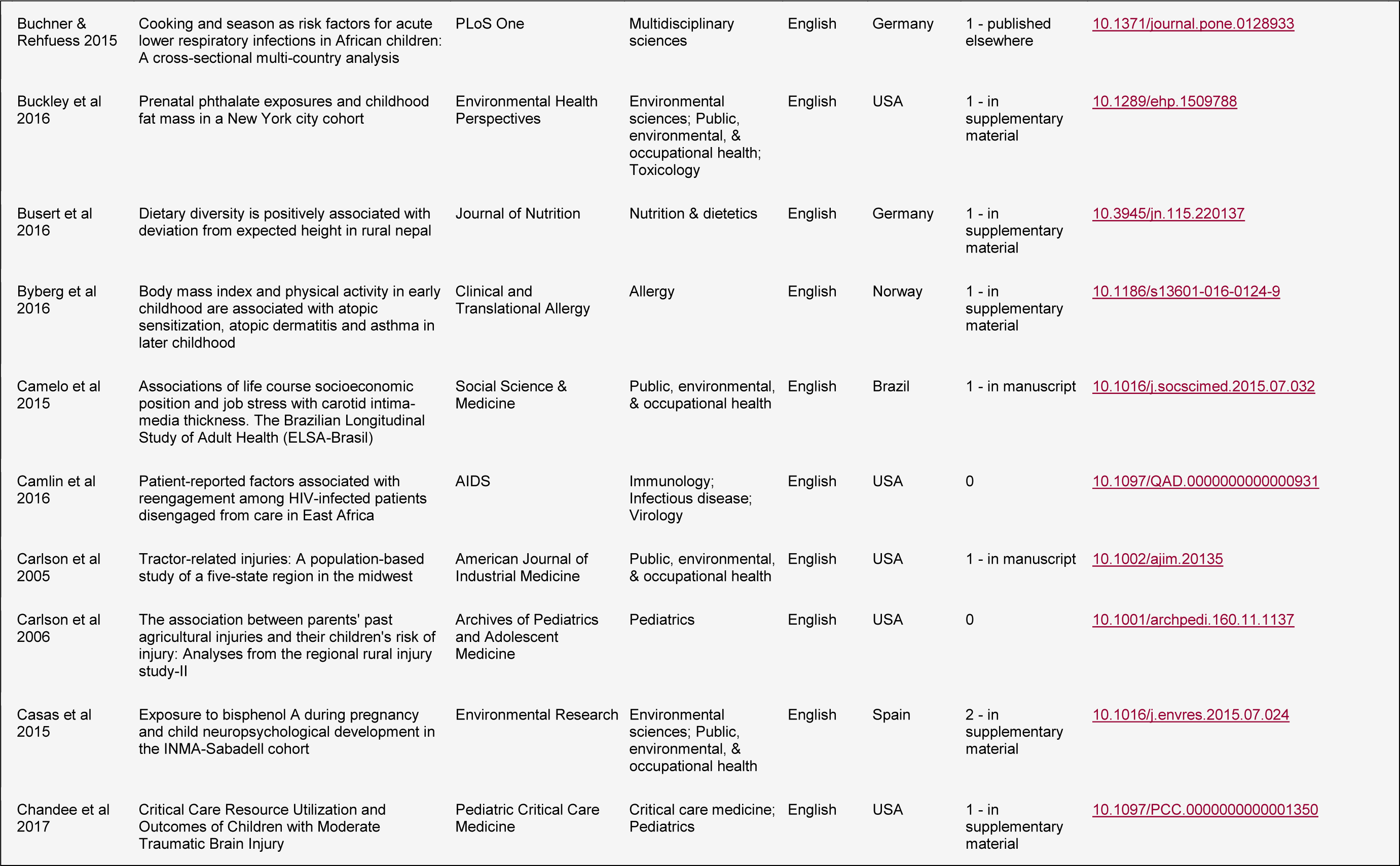
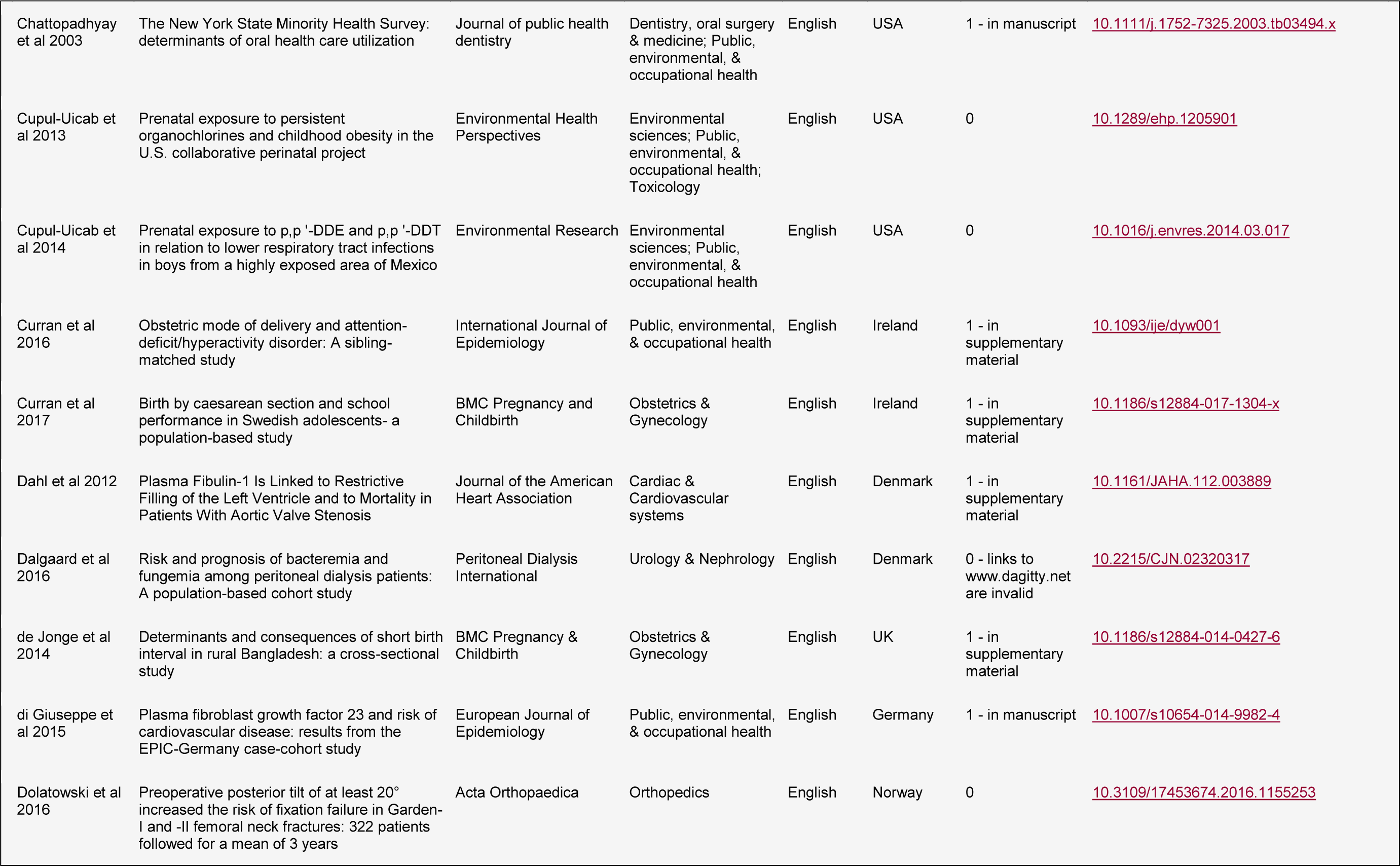
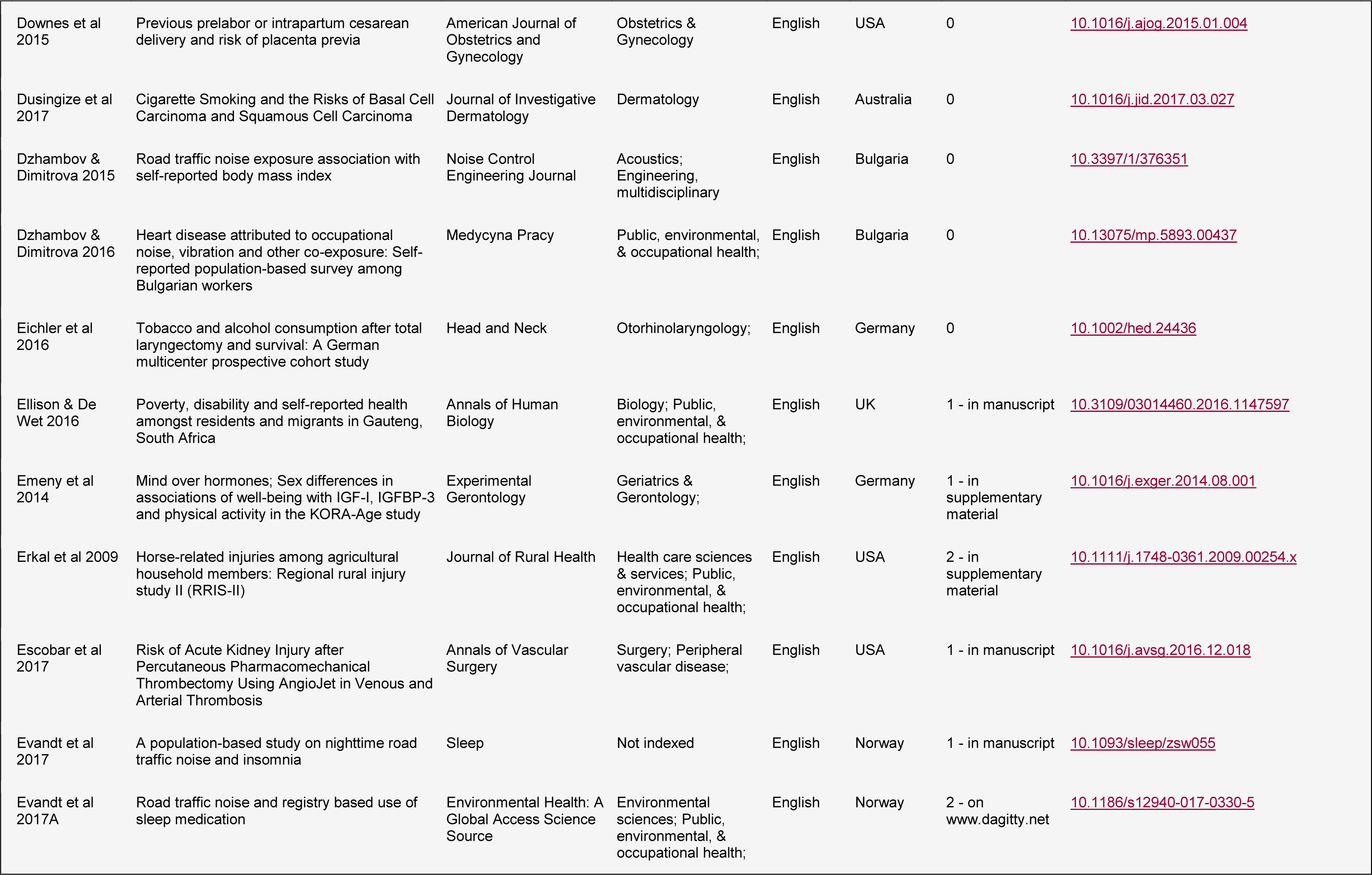
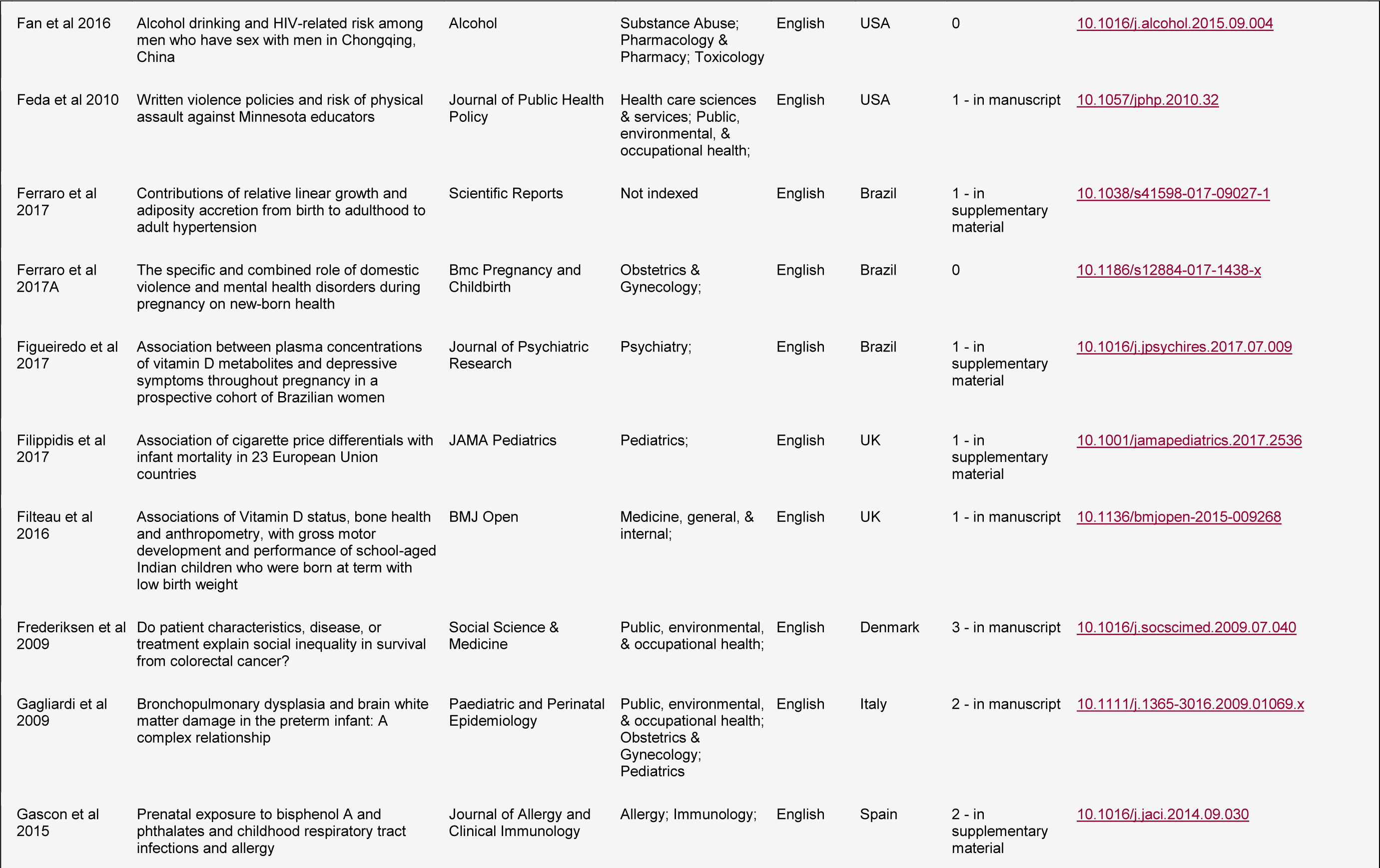
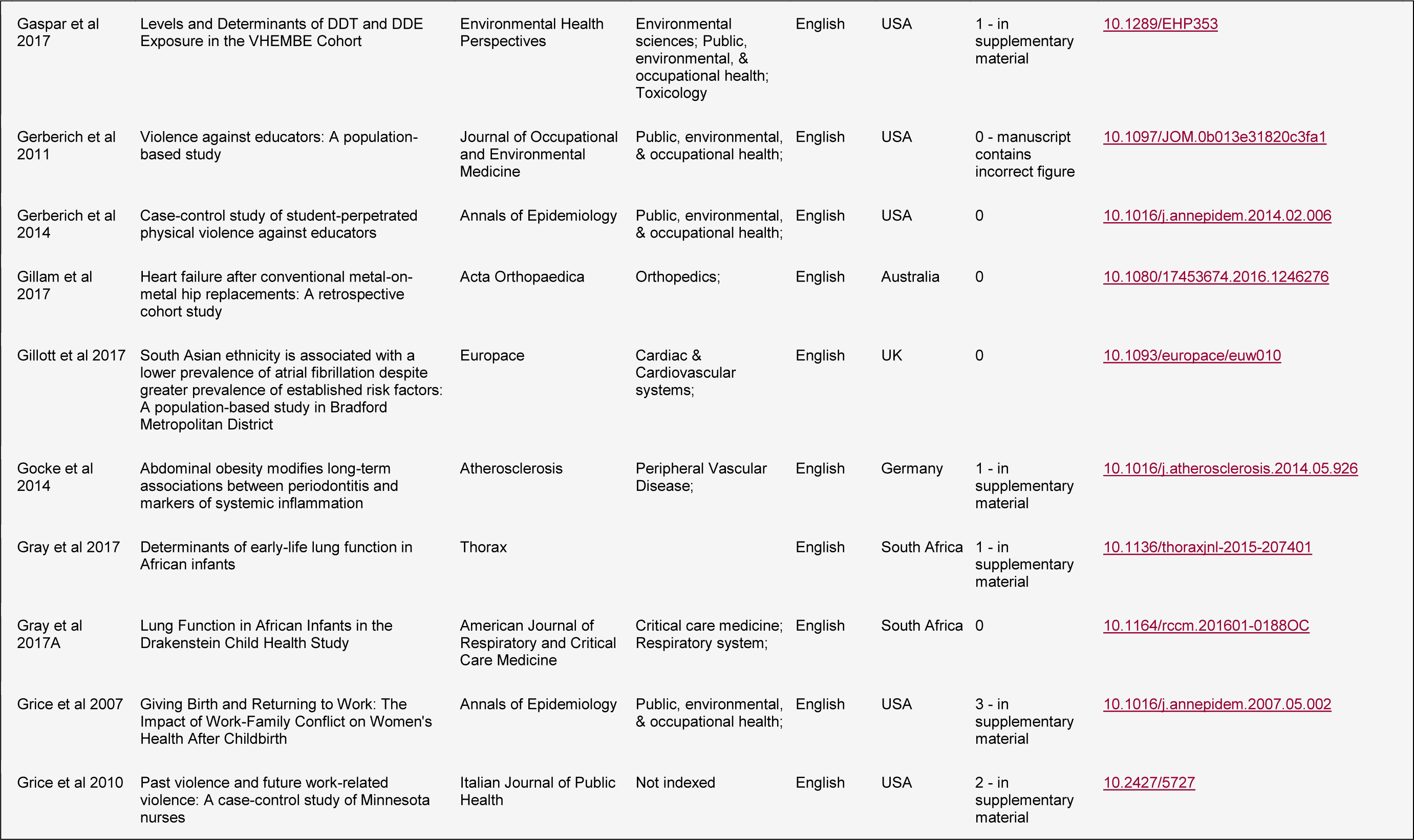
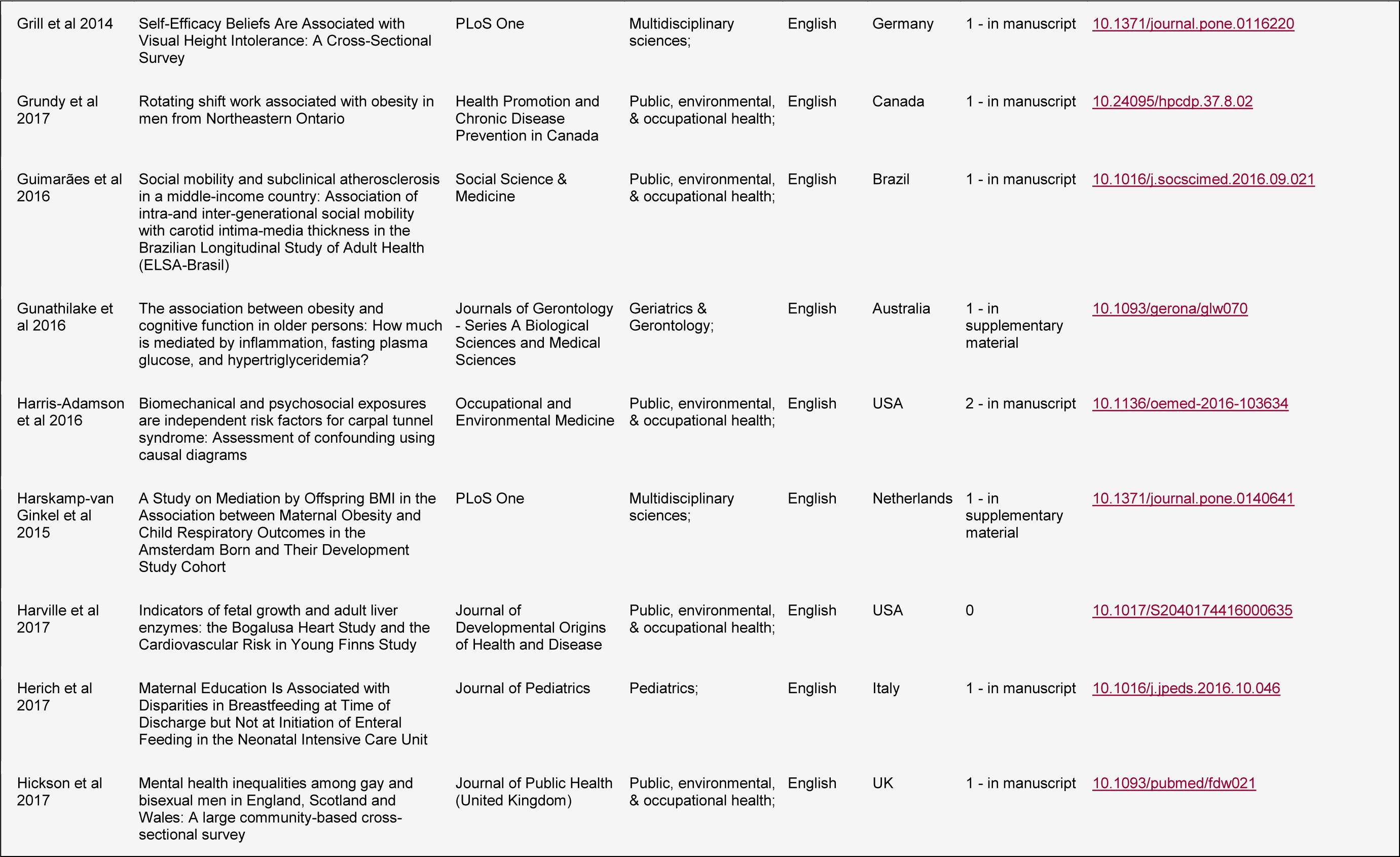
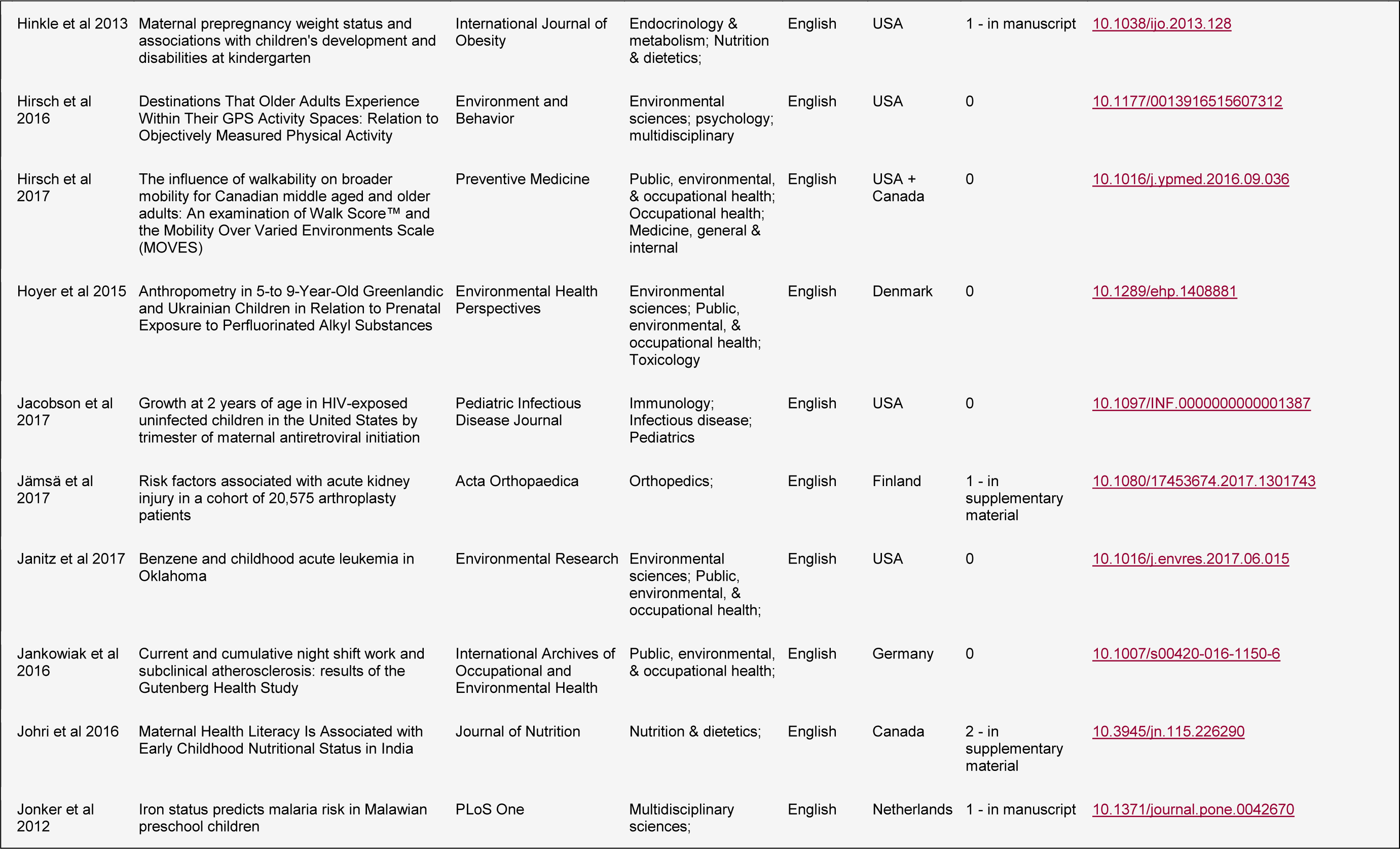
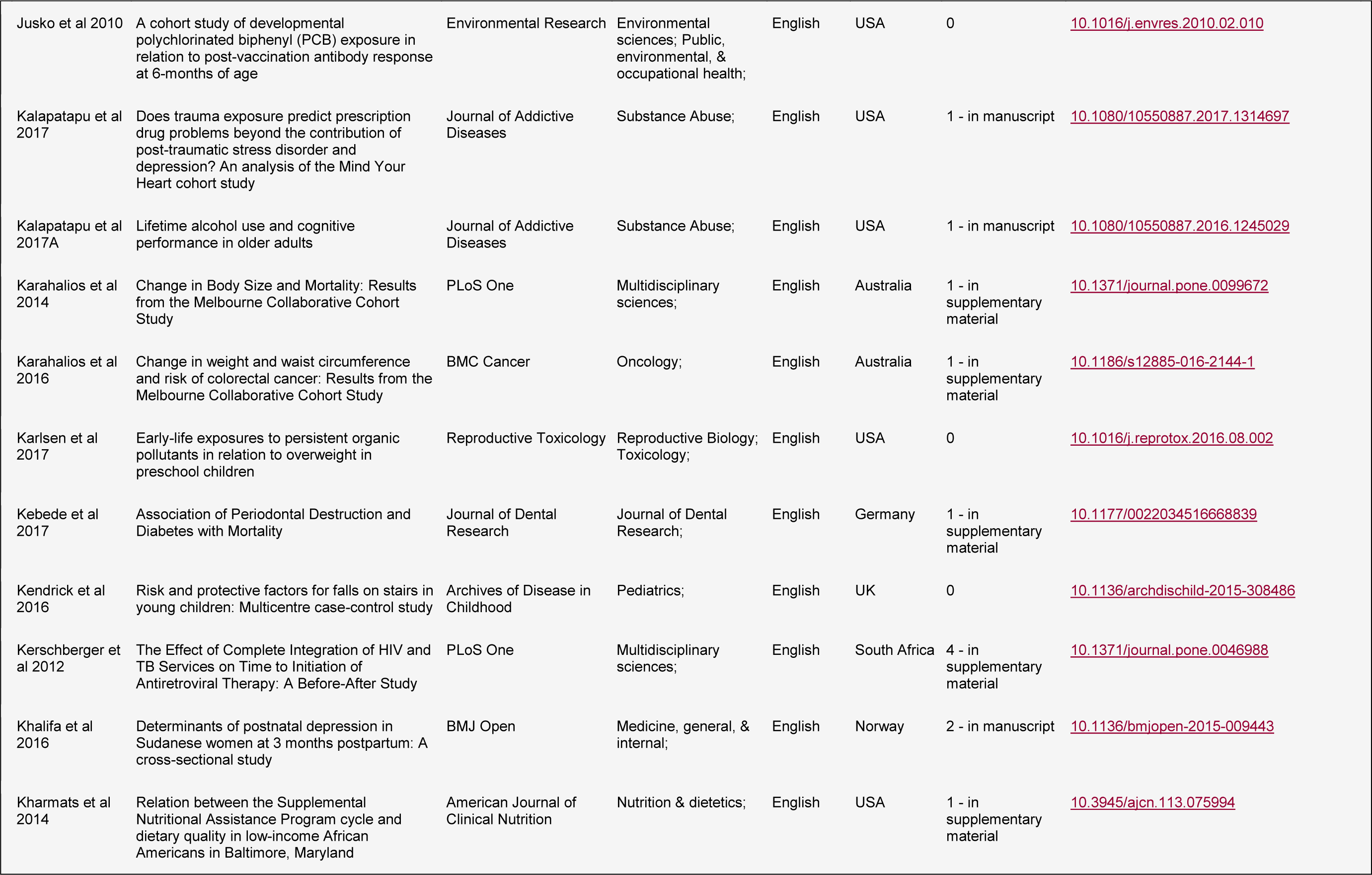
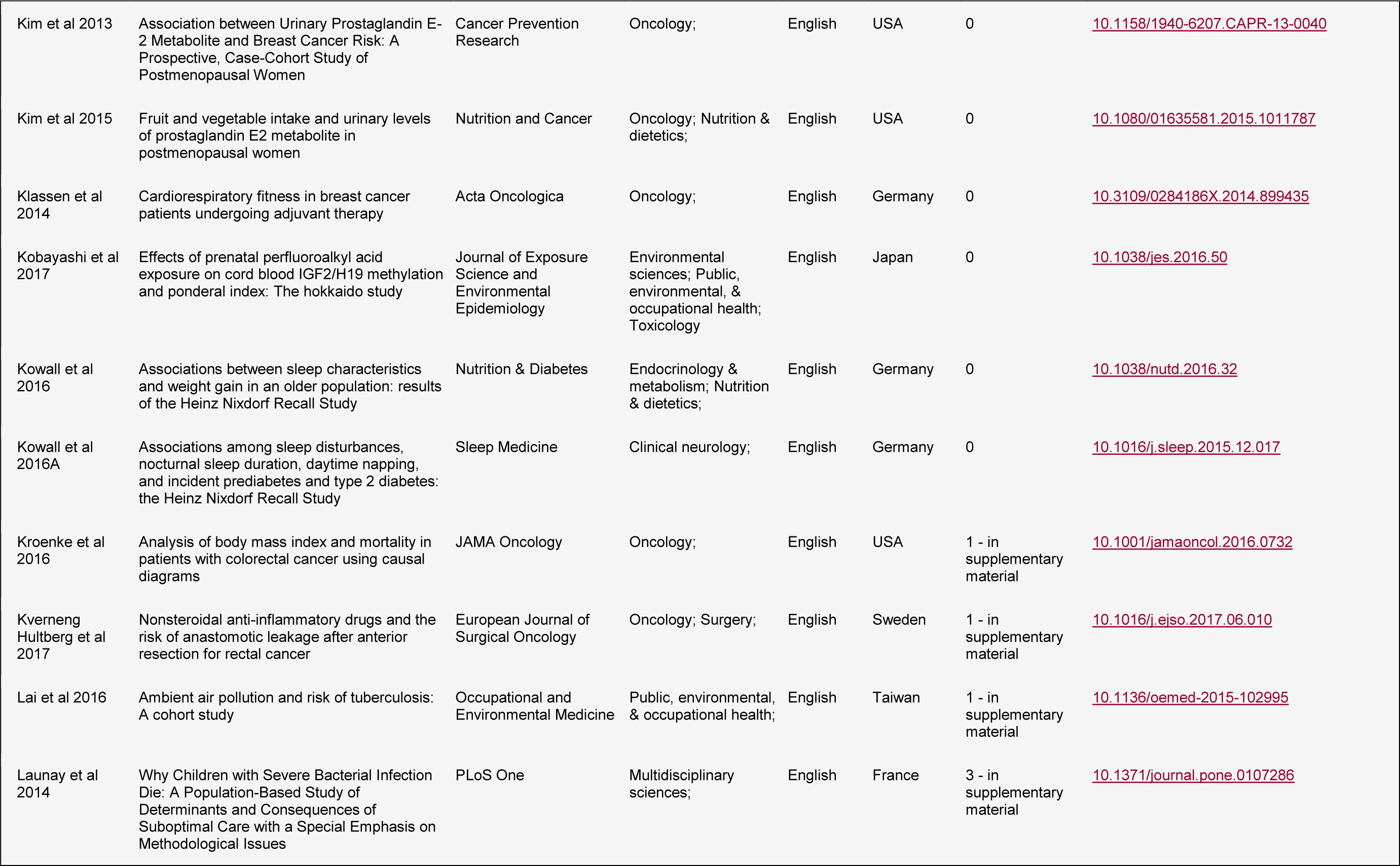
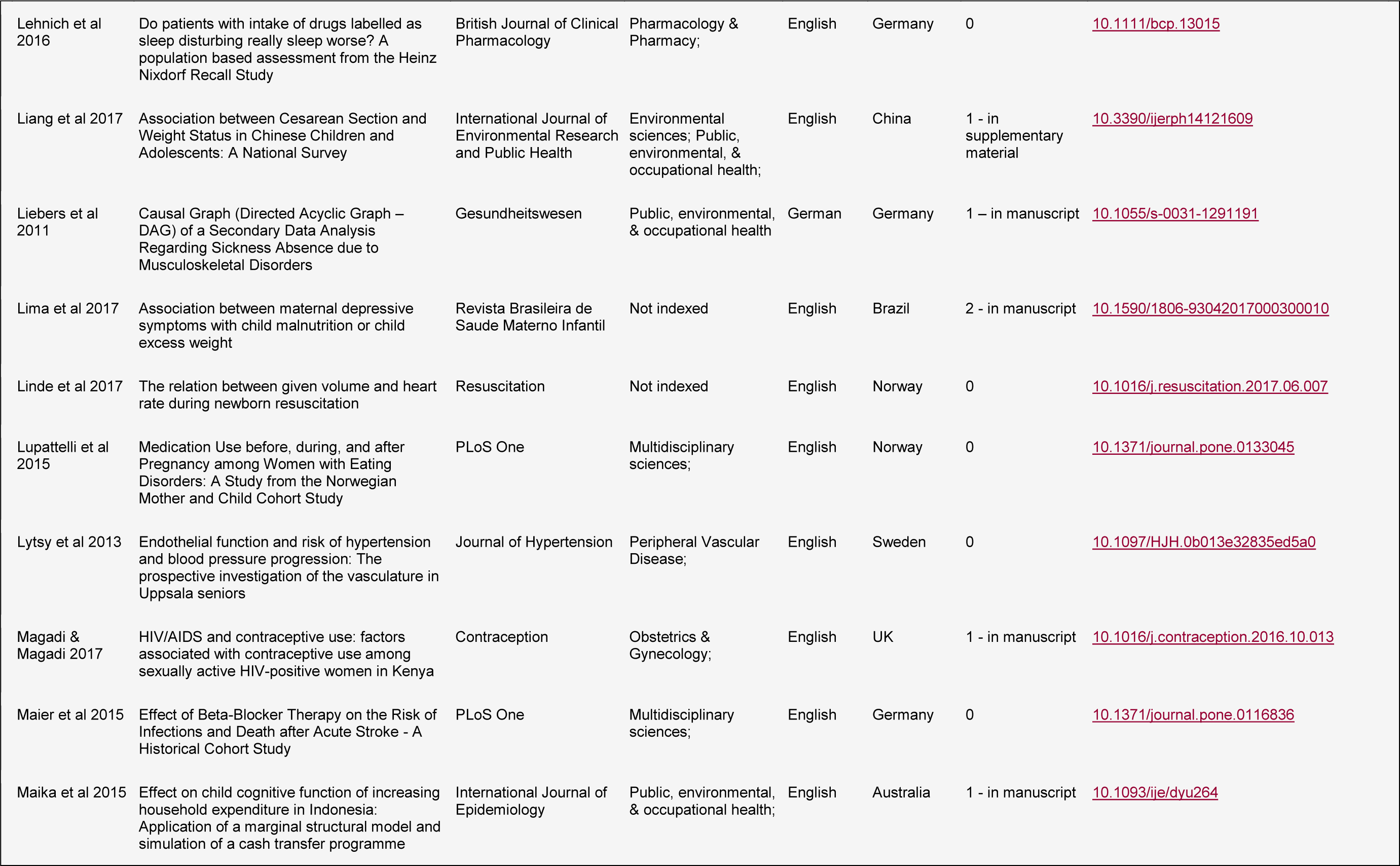
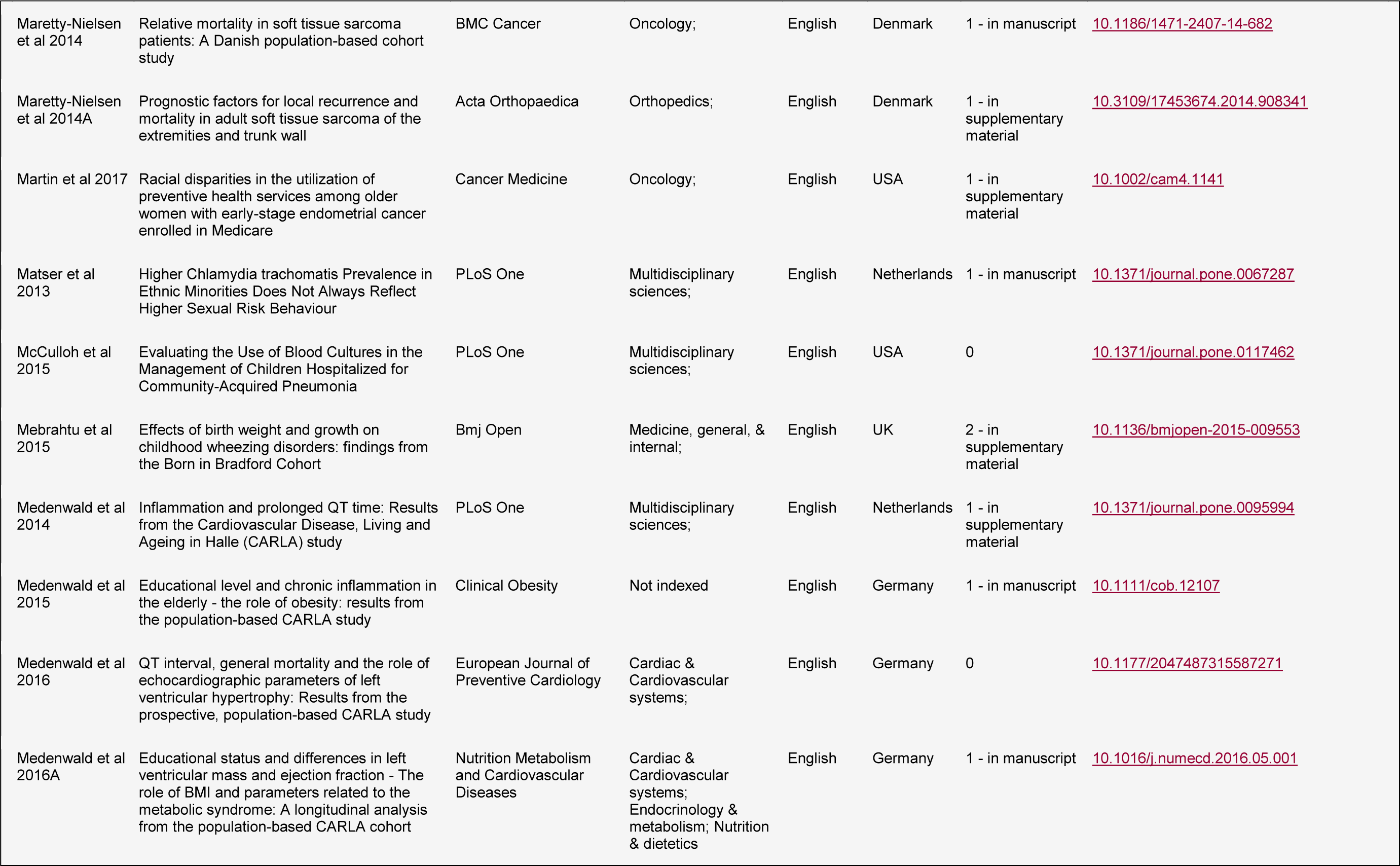
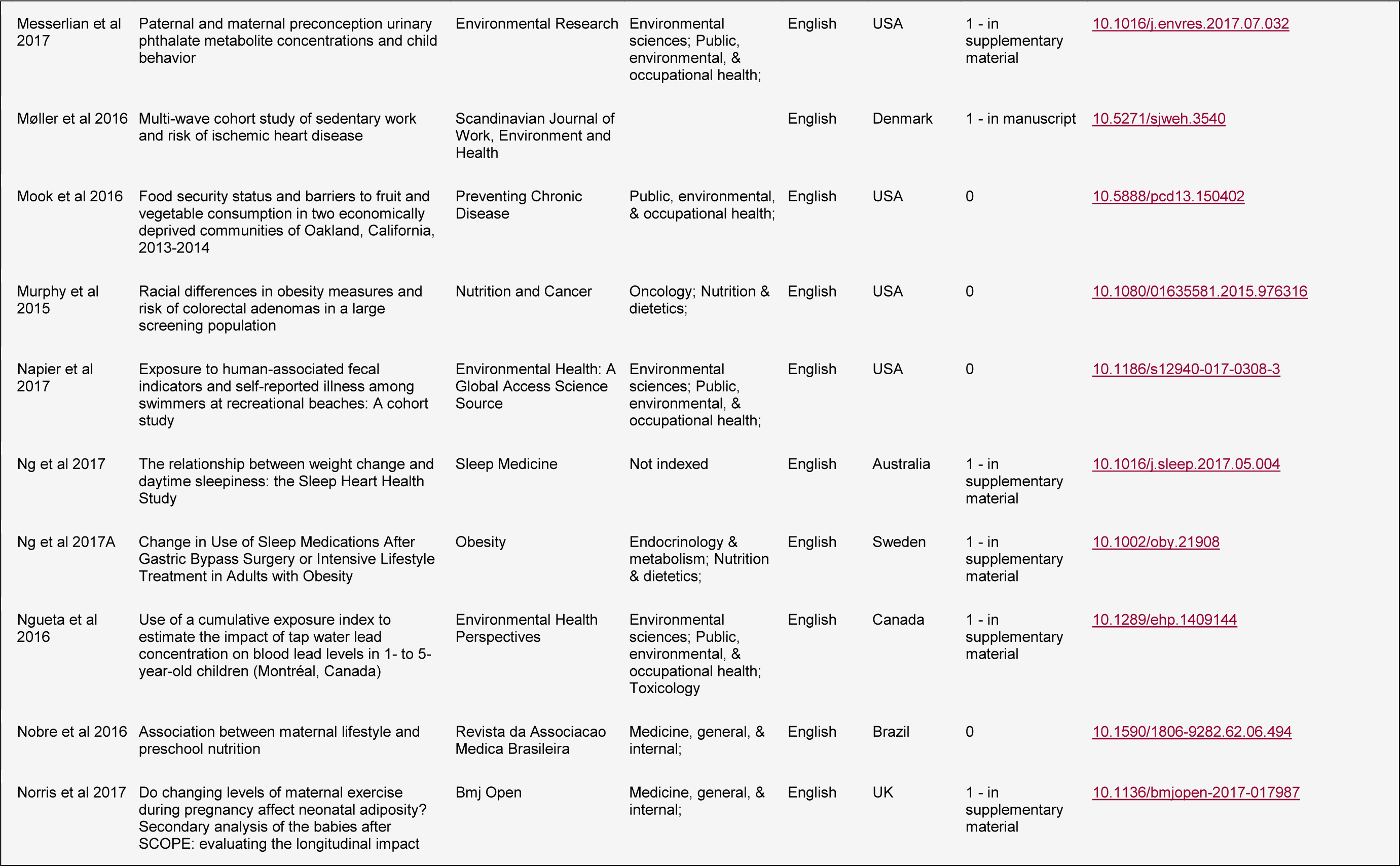
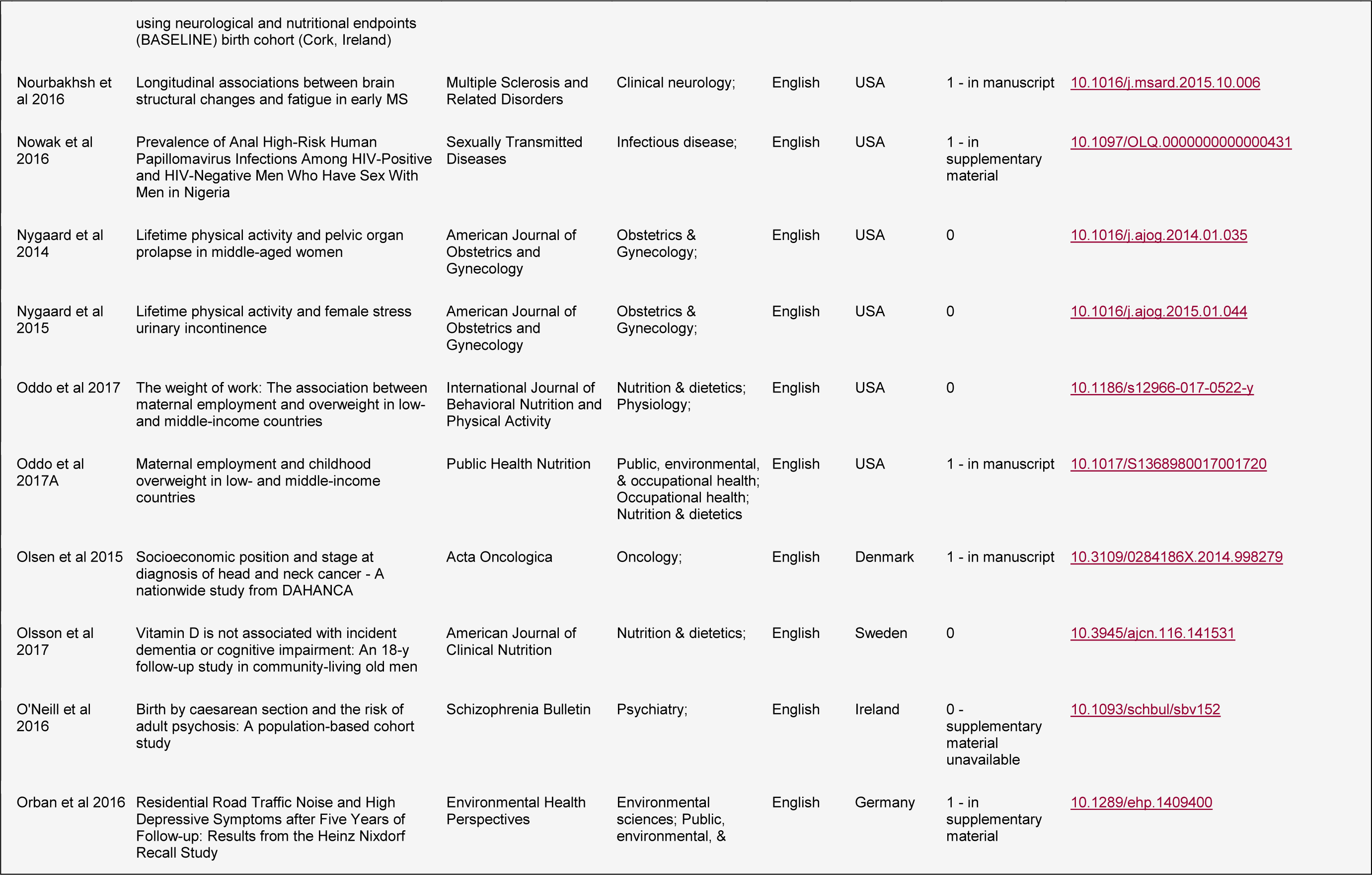
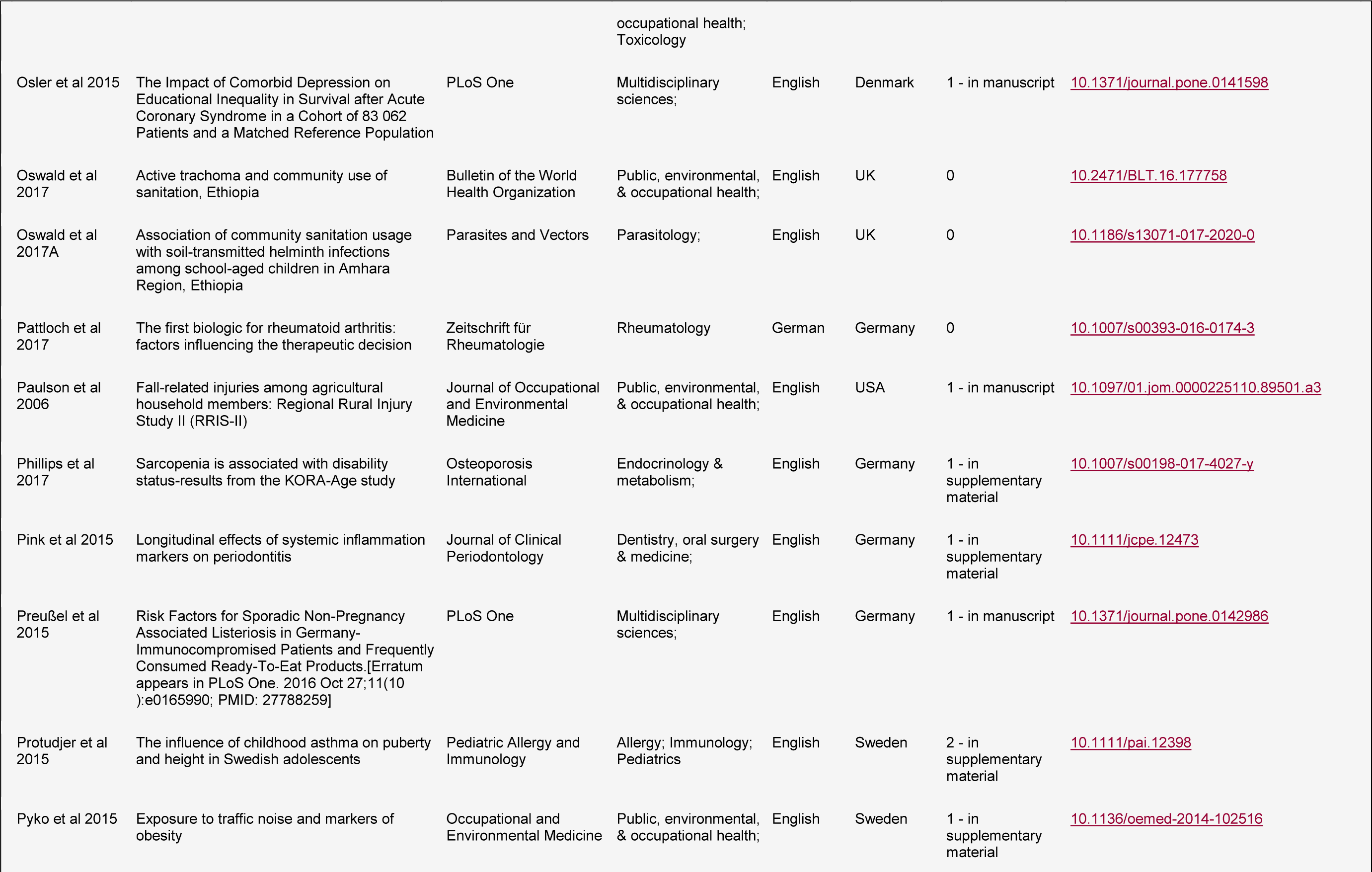
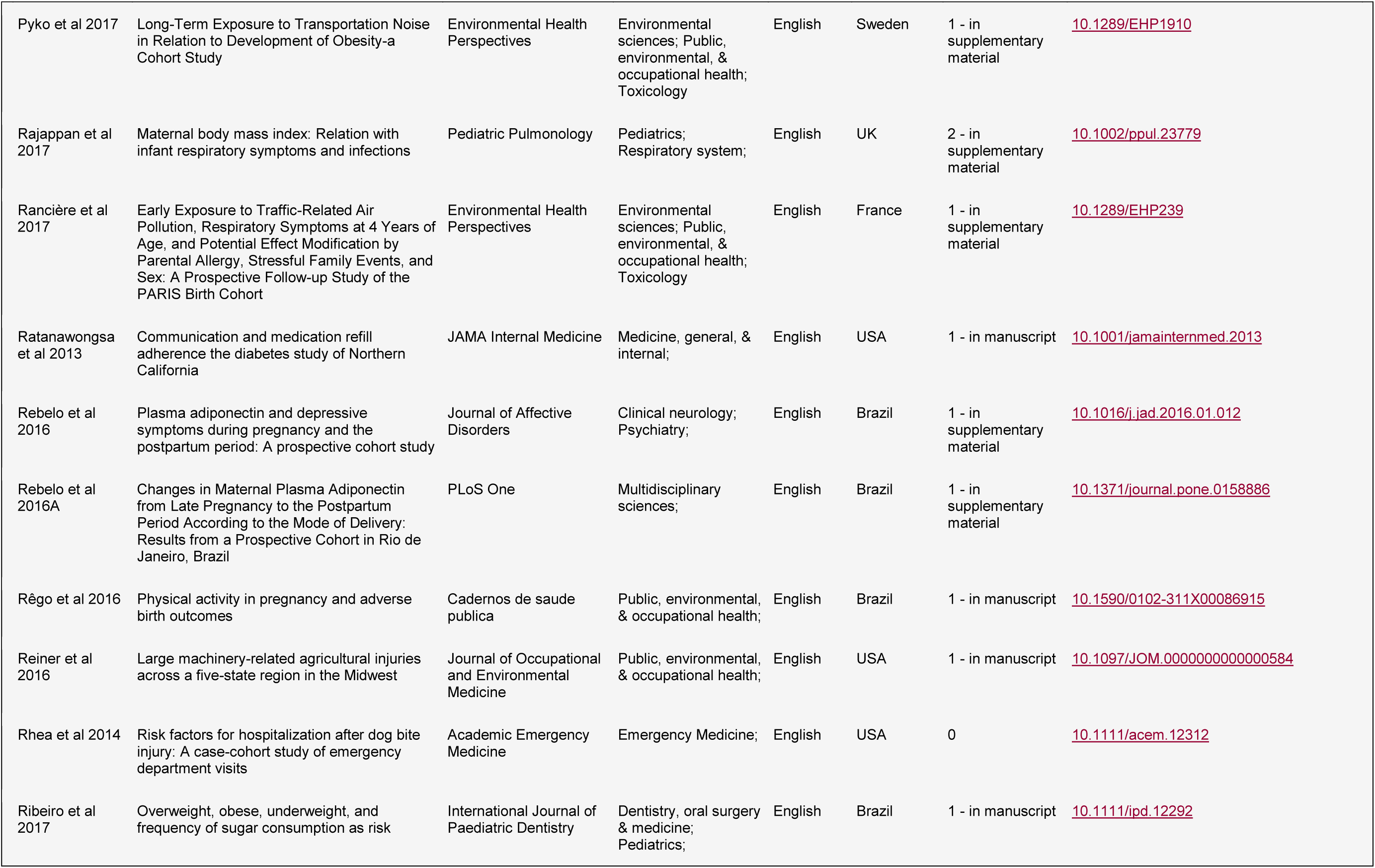
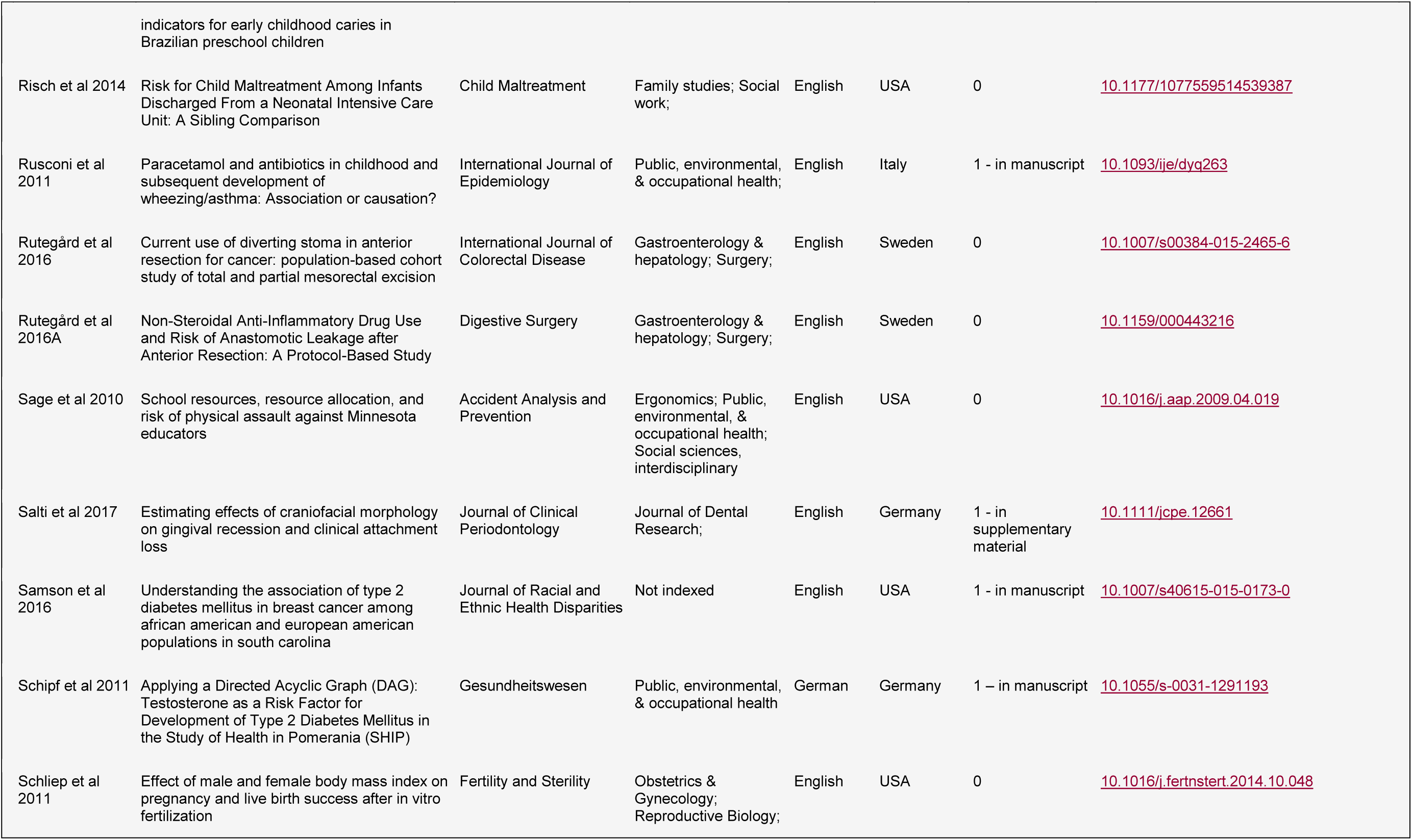
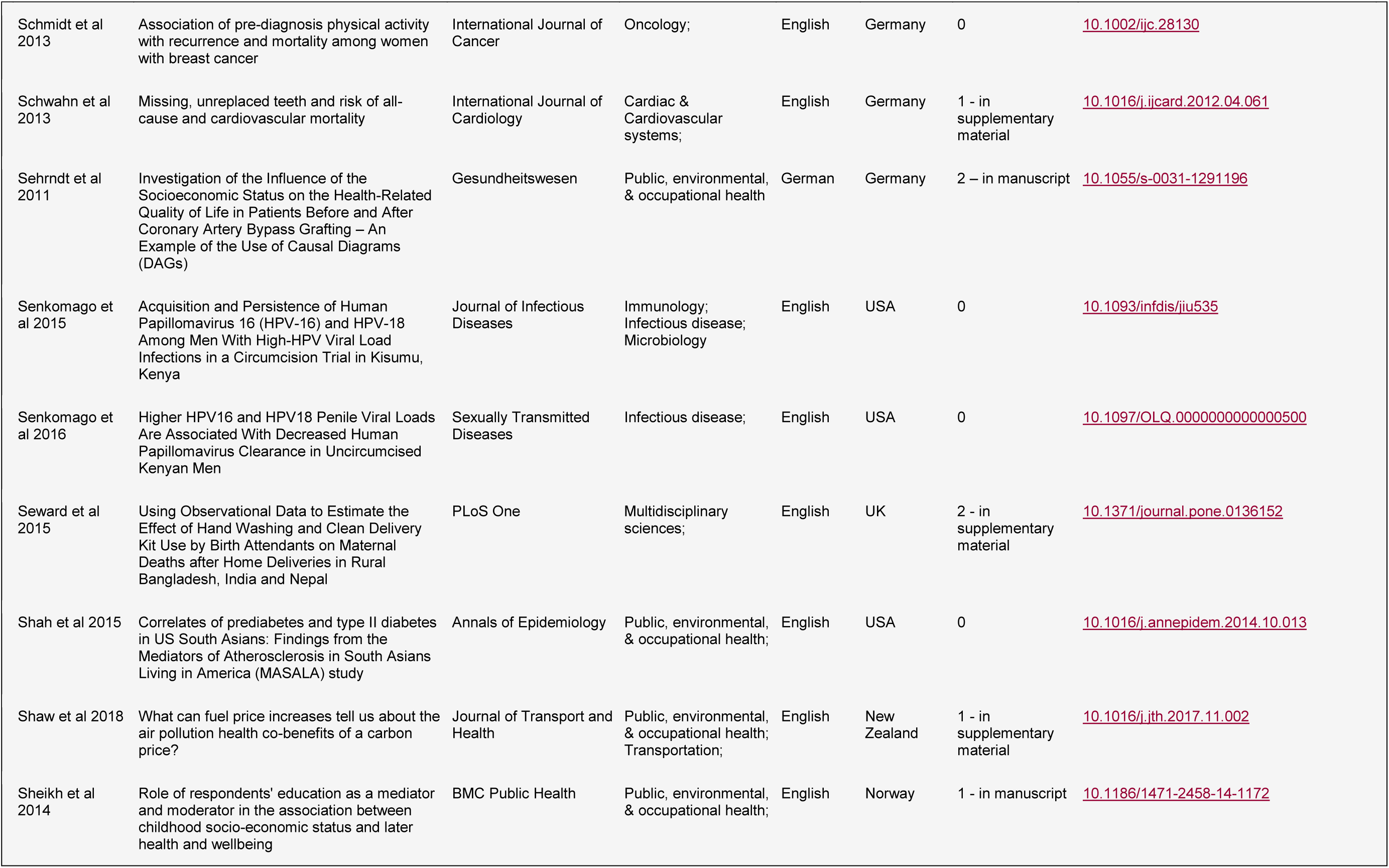
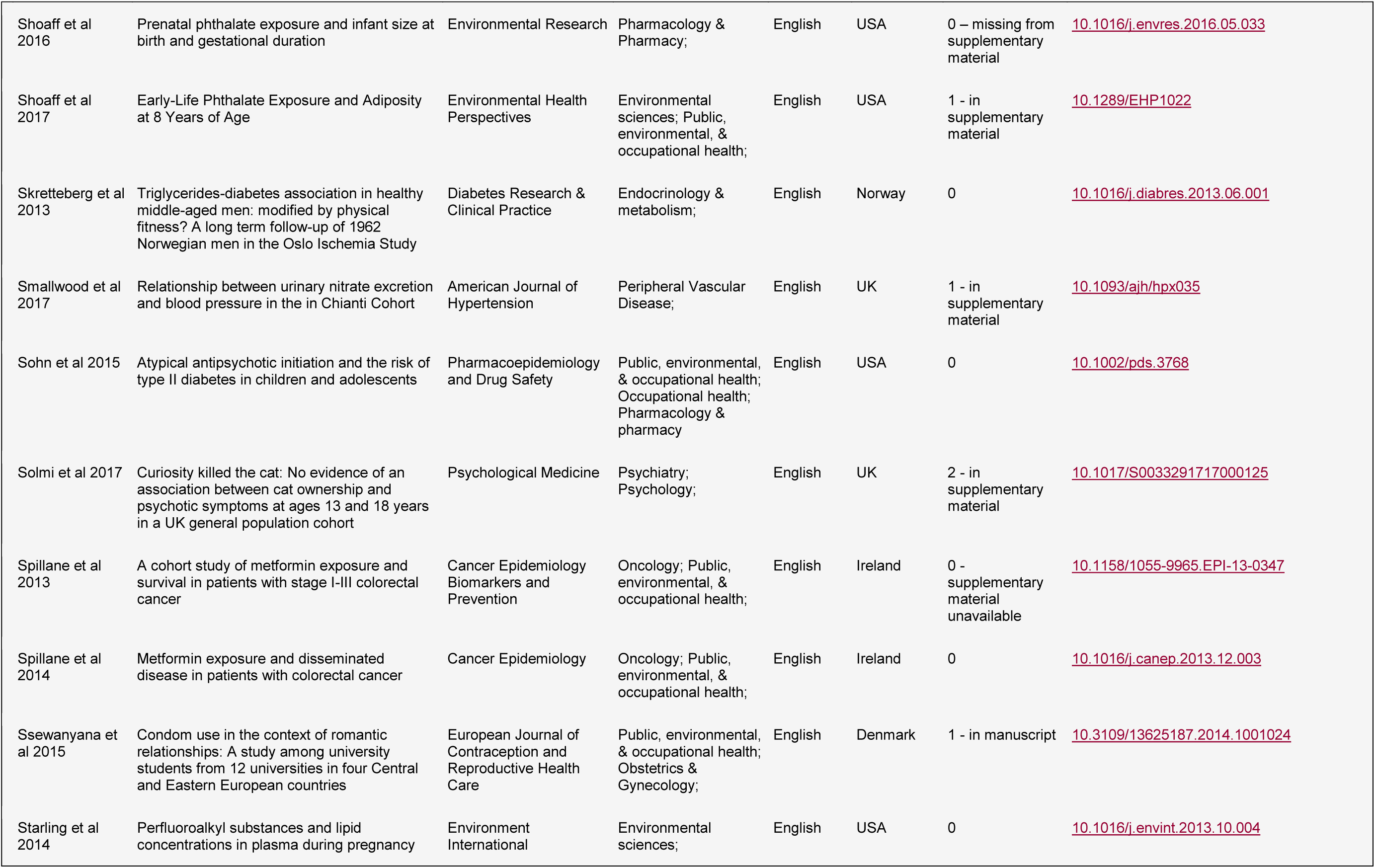
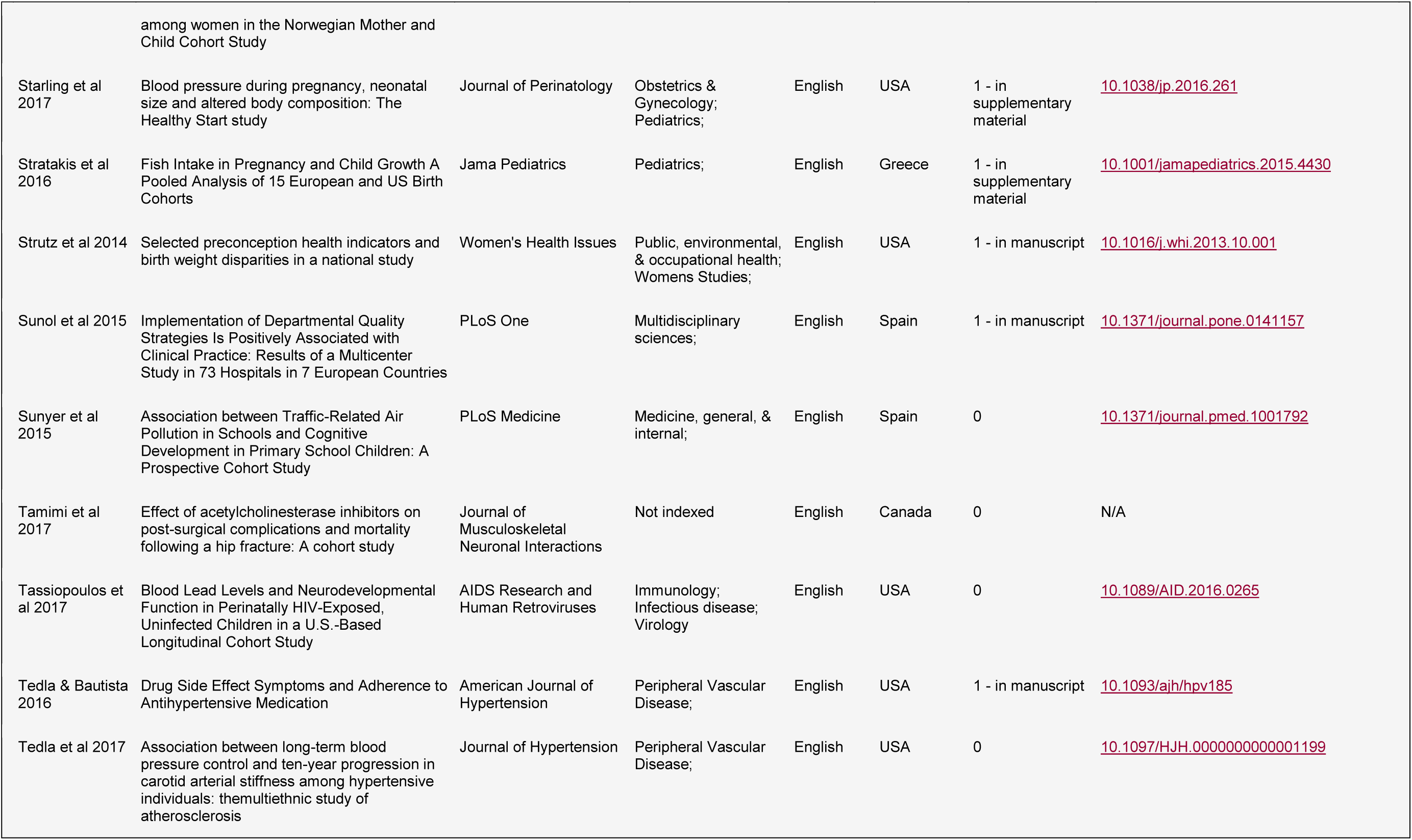
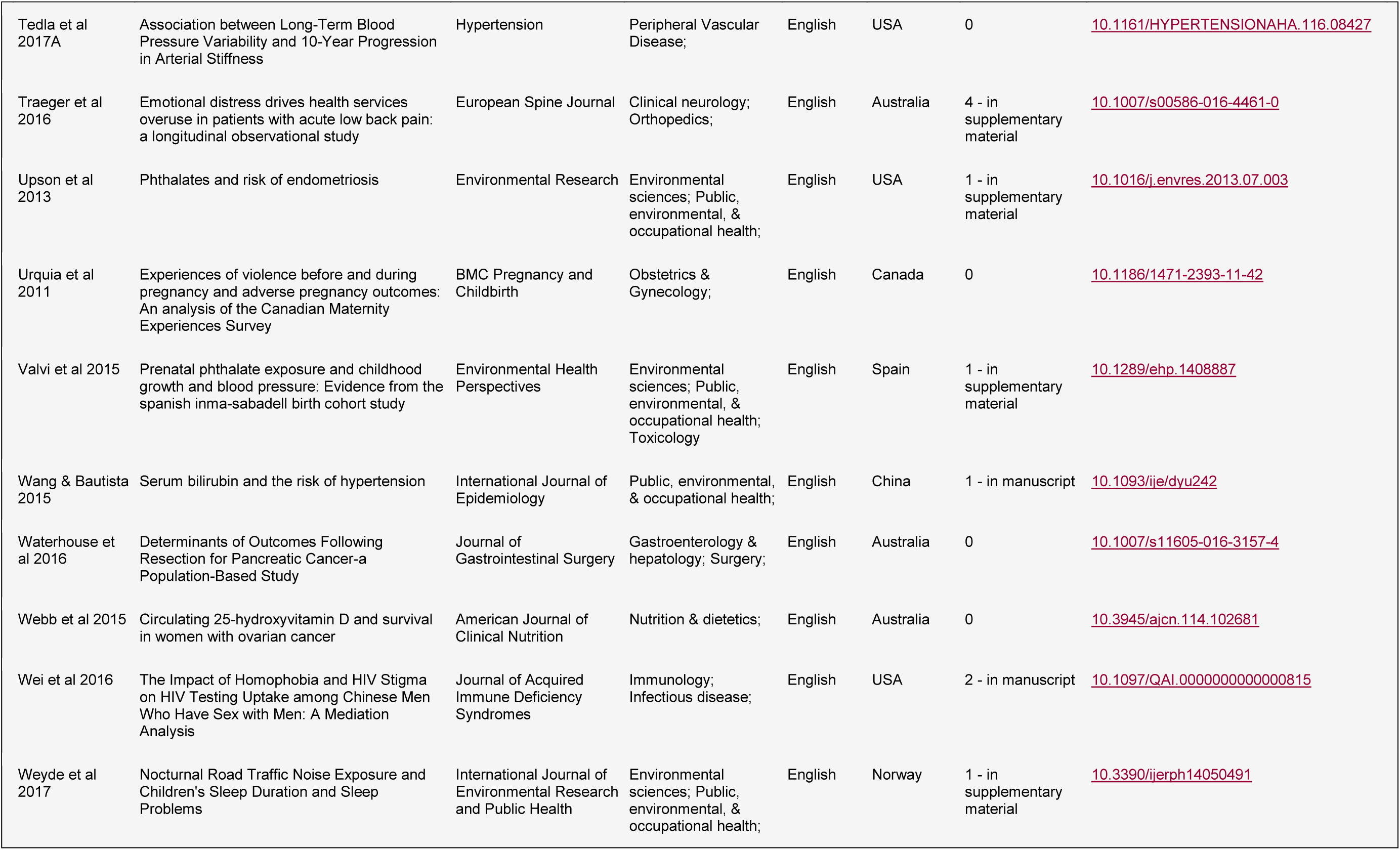
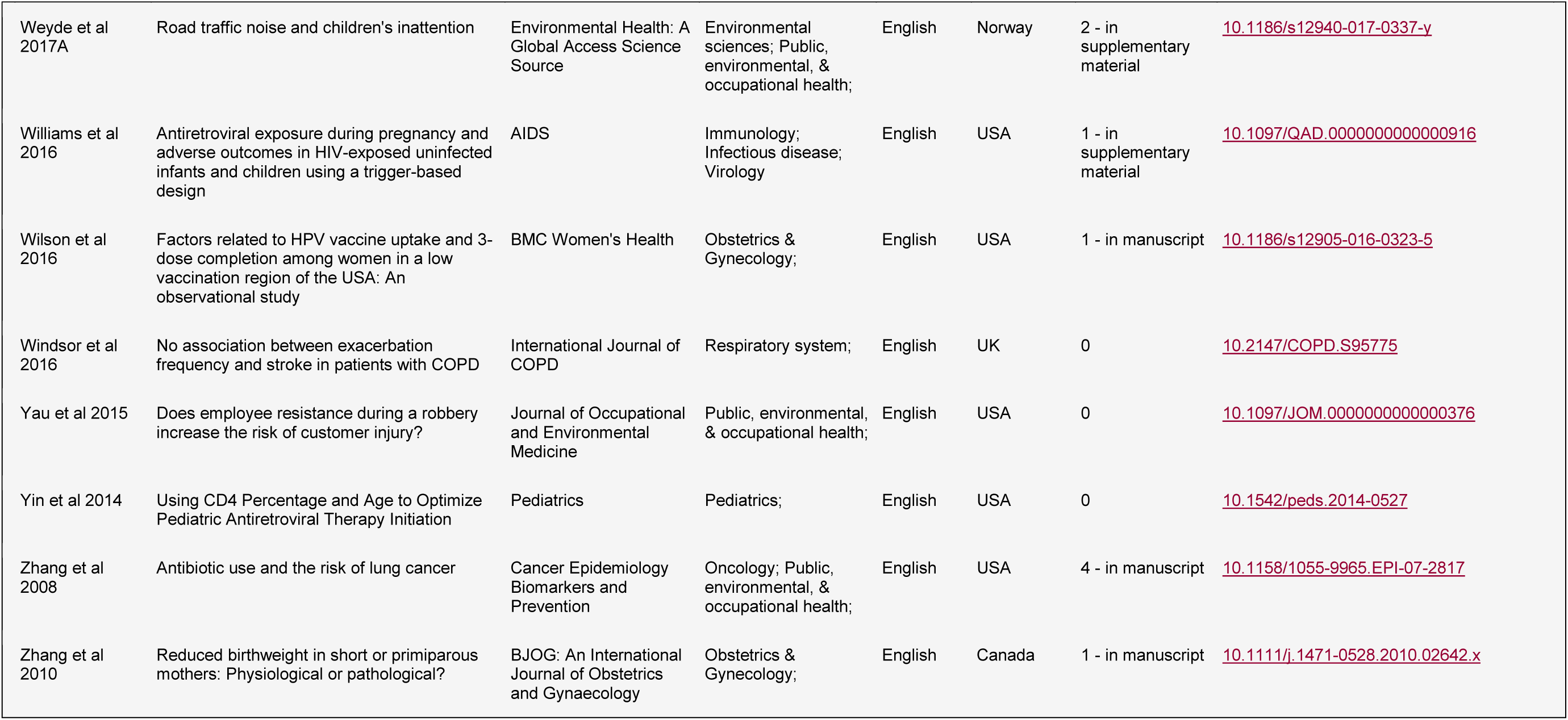
Summary details of the 234 articles included in the review

**Supplementary Table 2.**
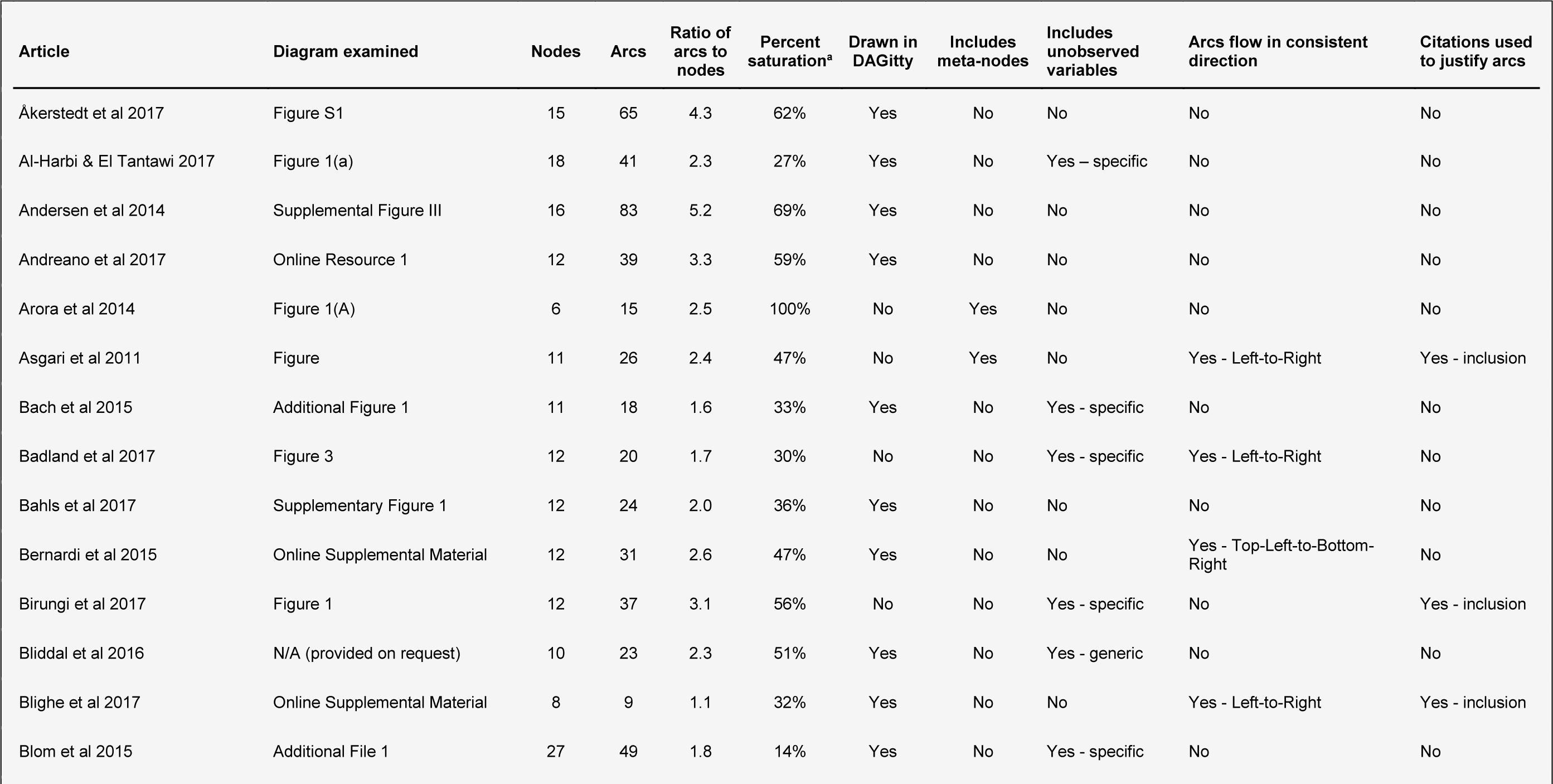
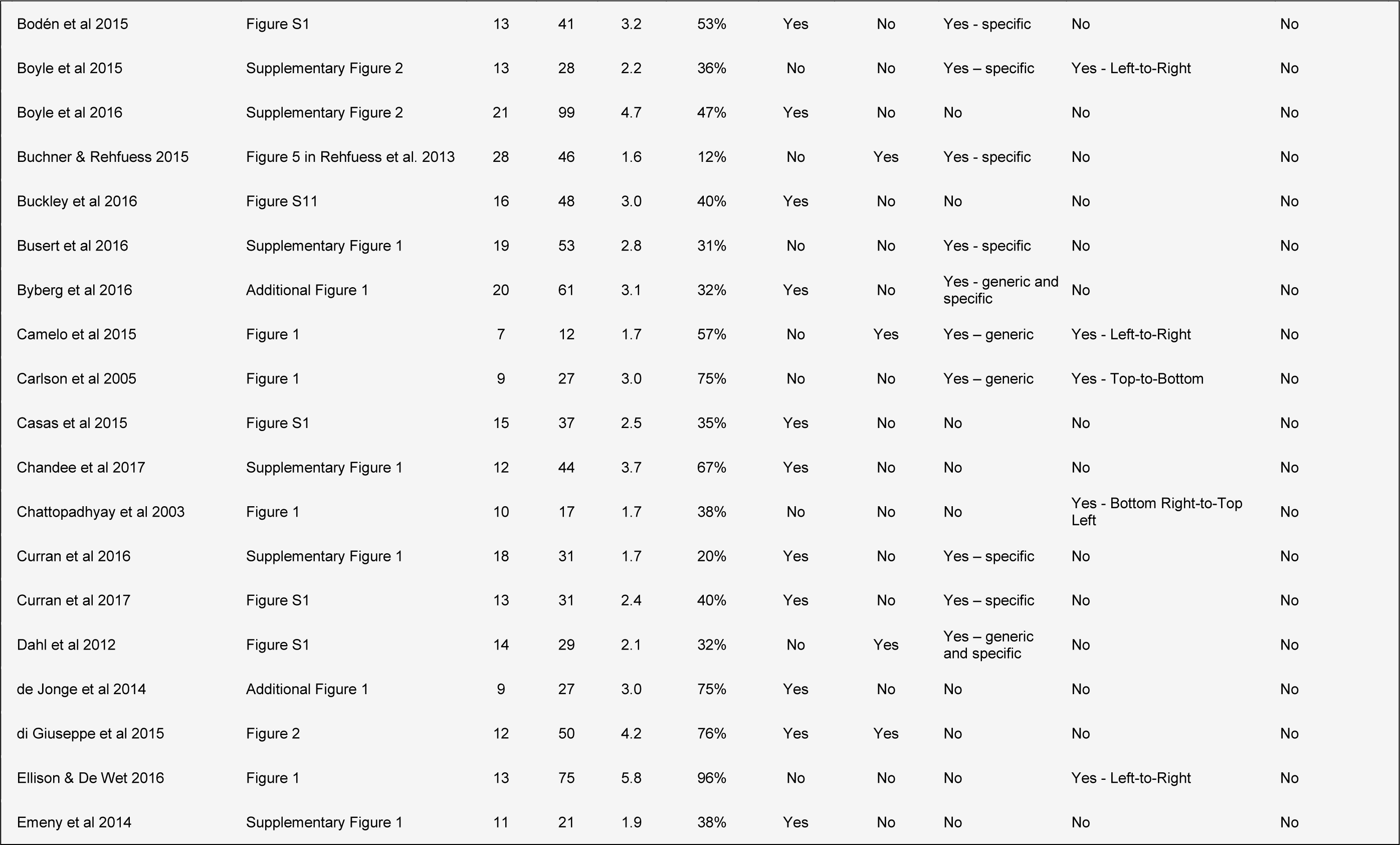
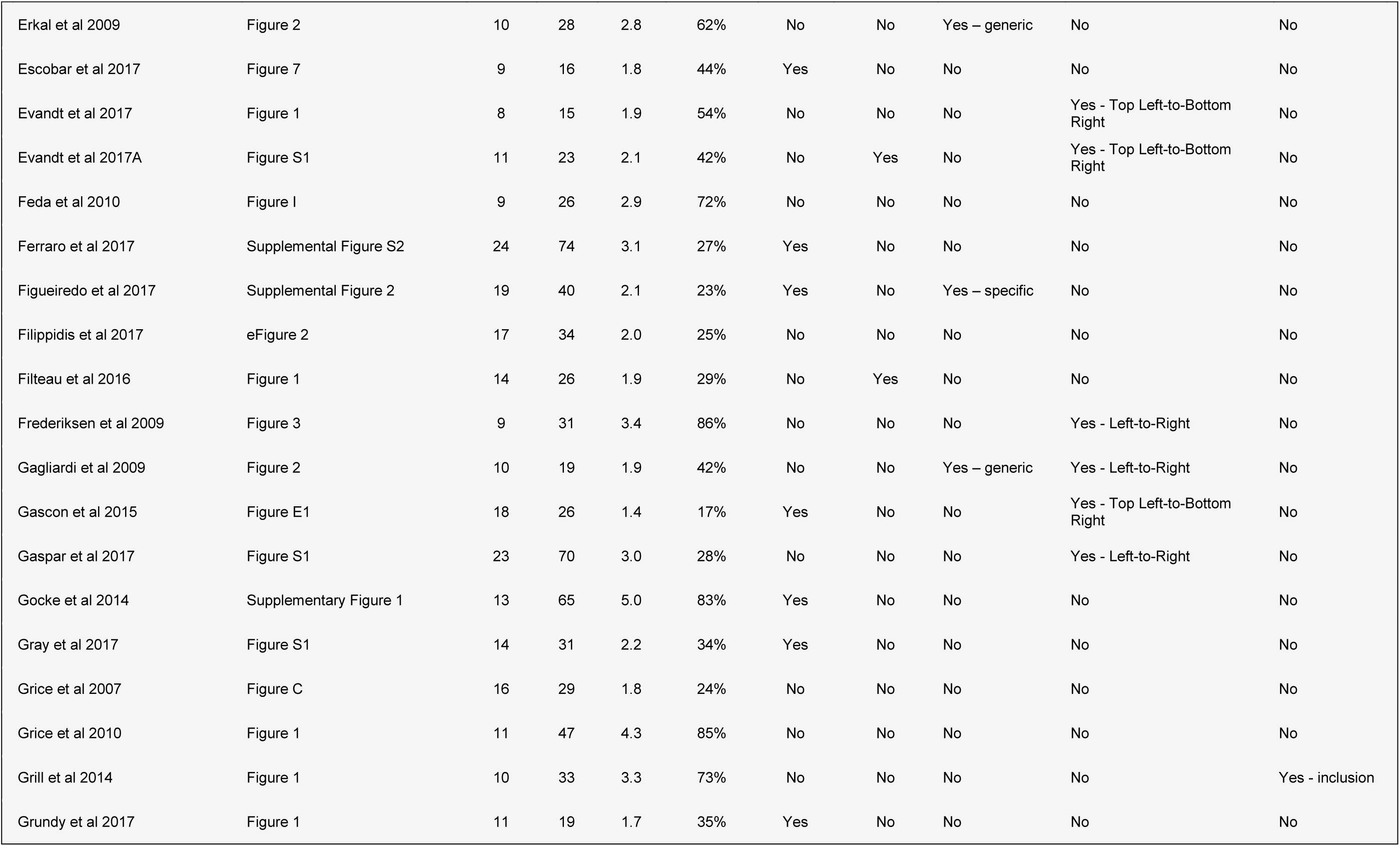
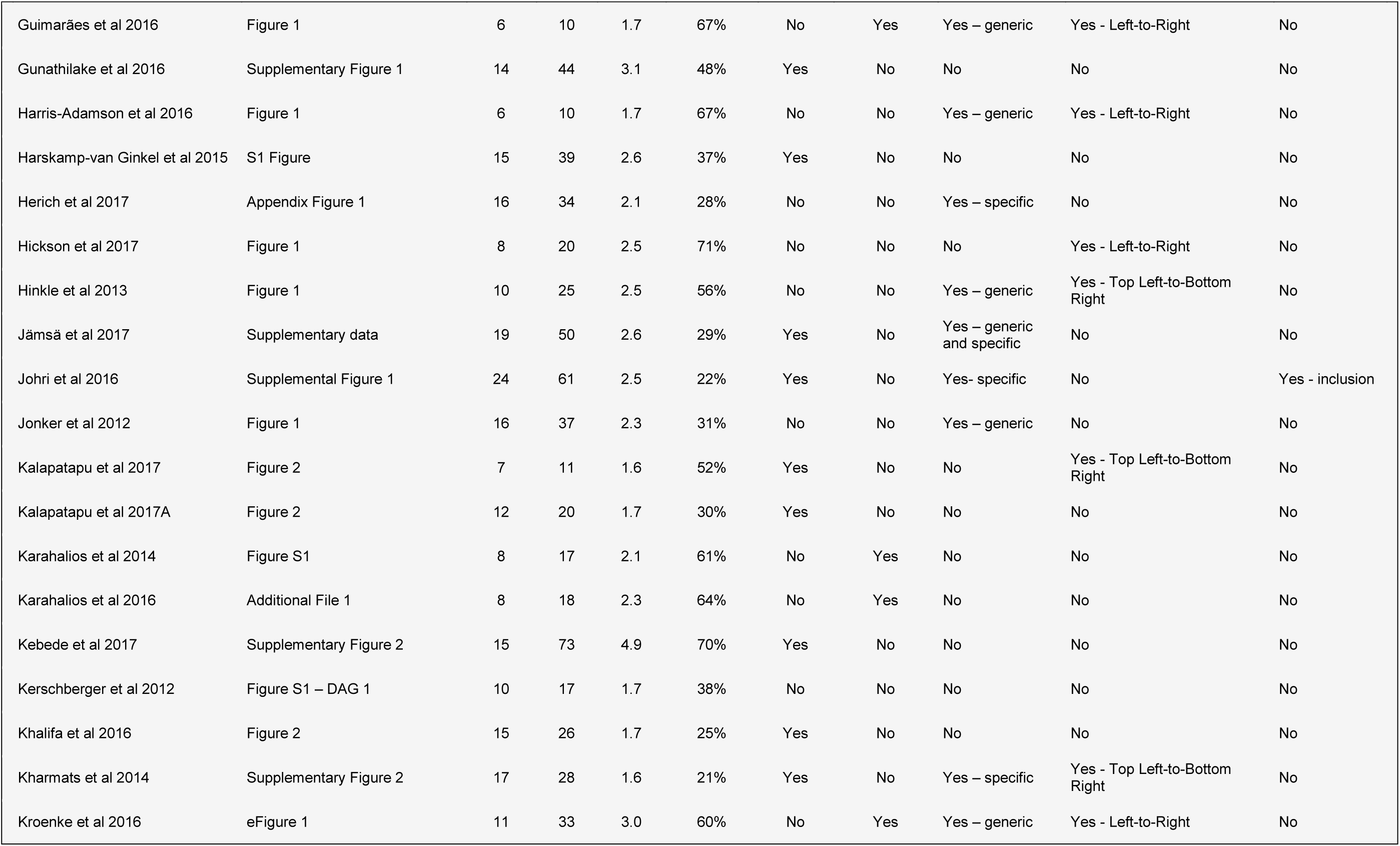
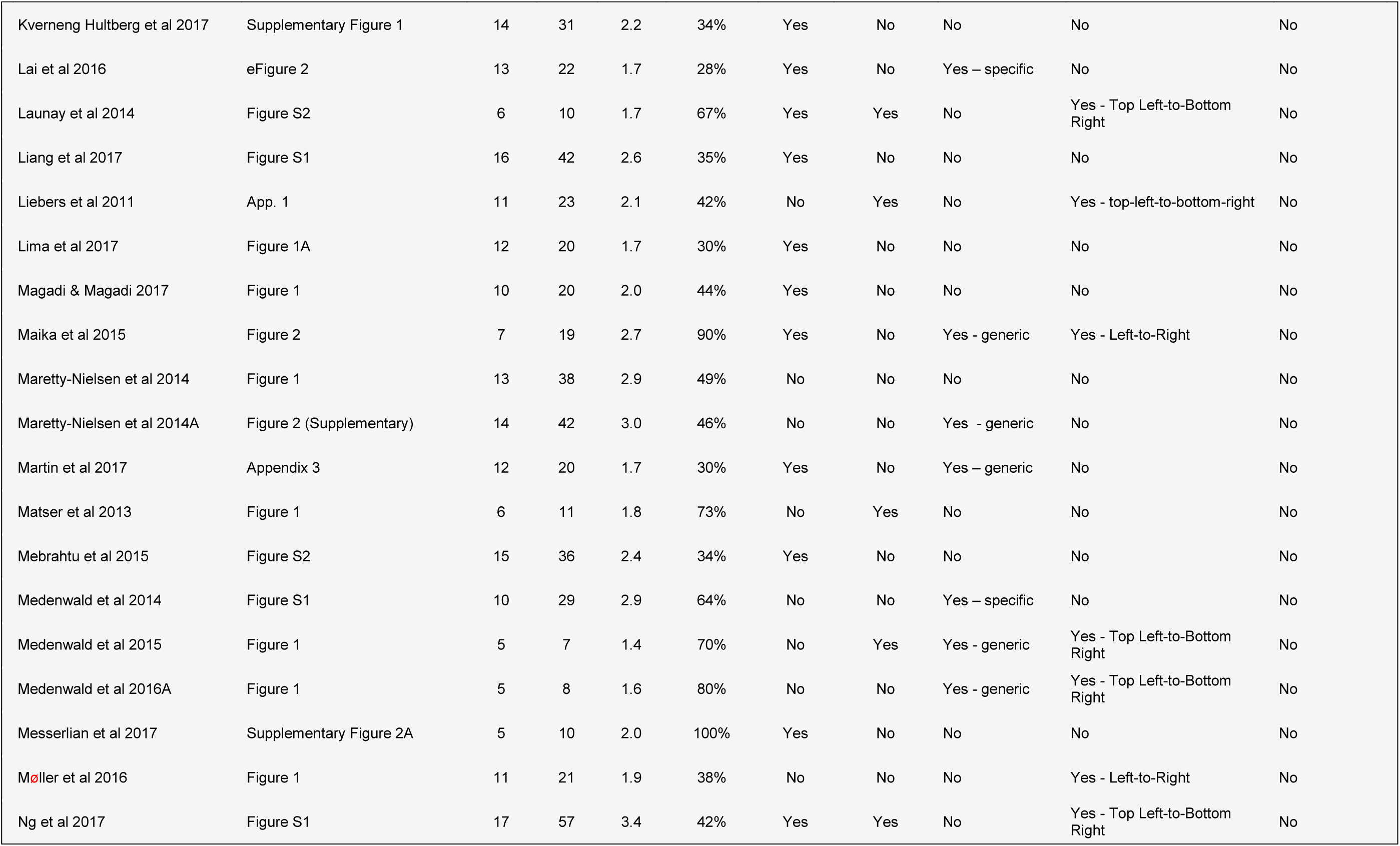
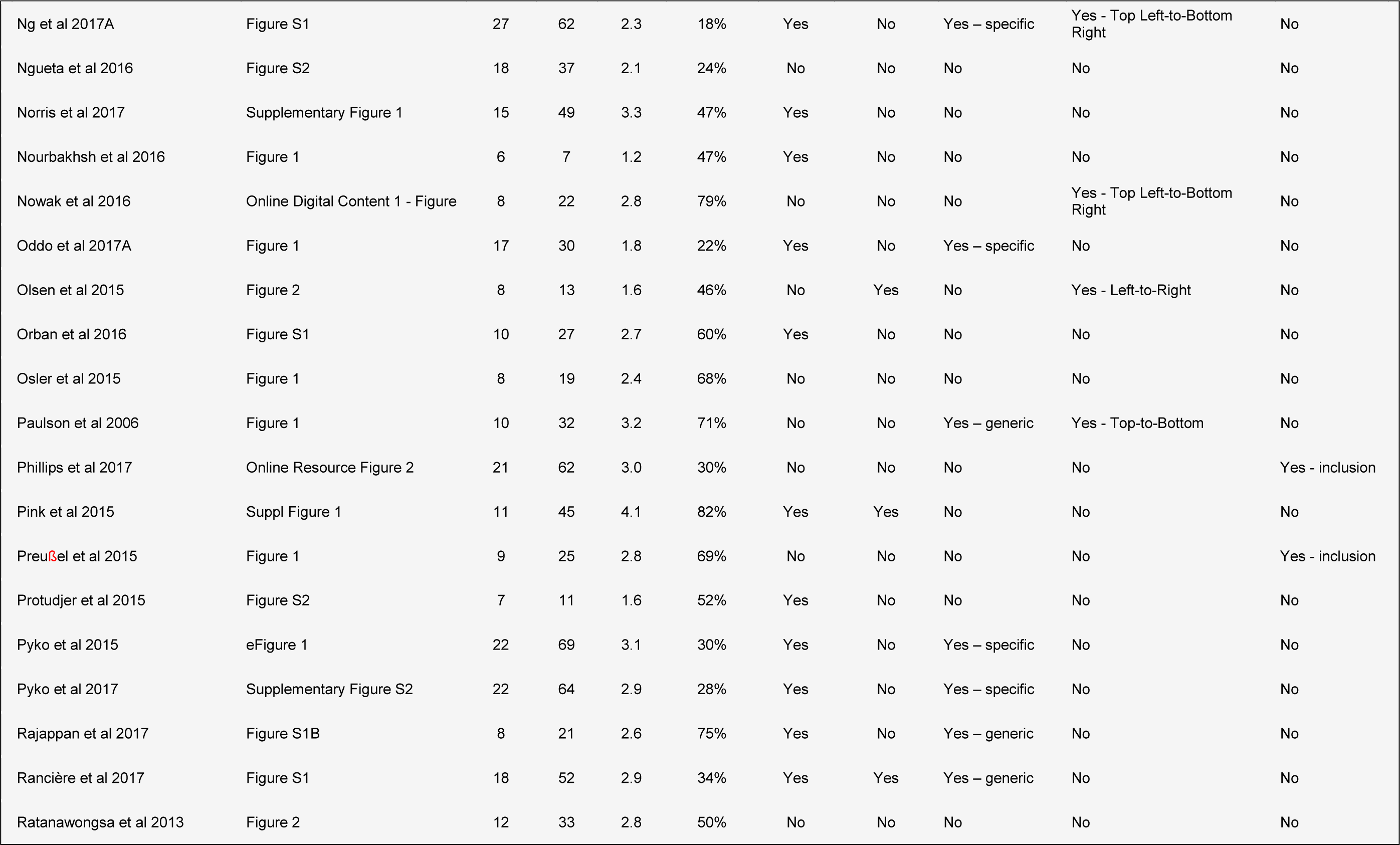
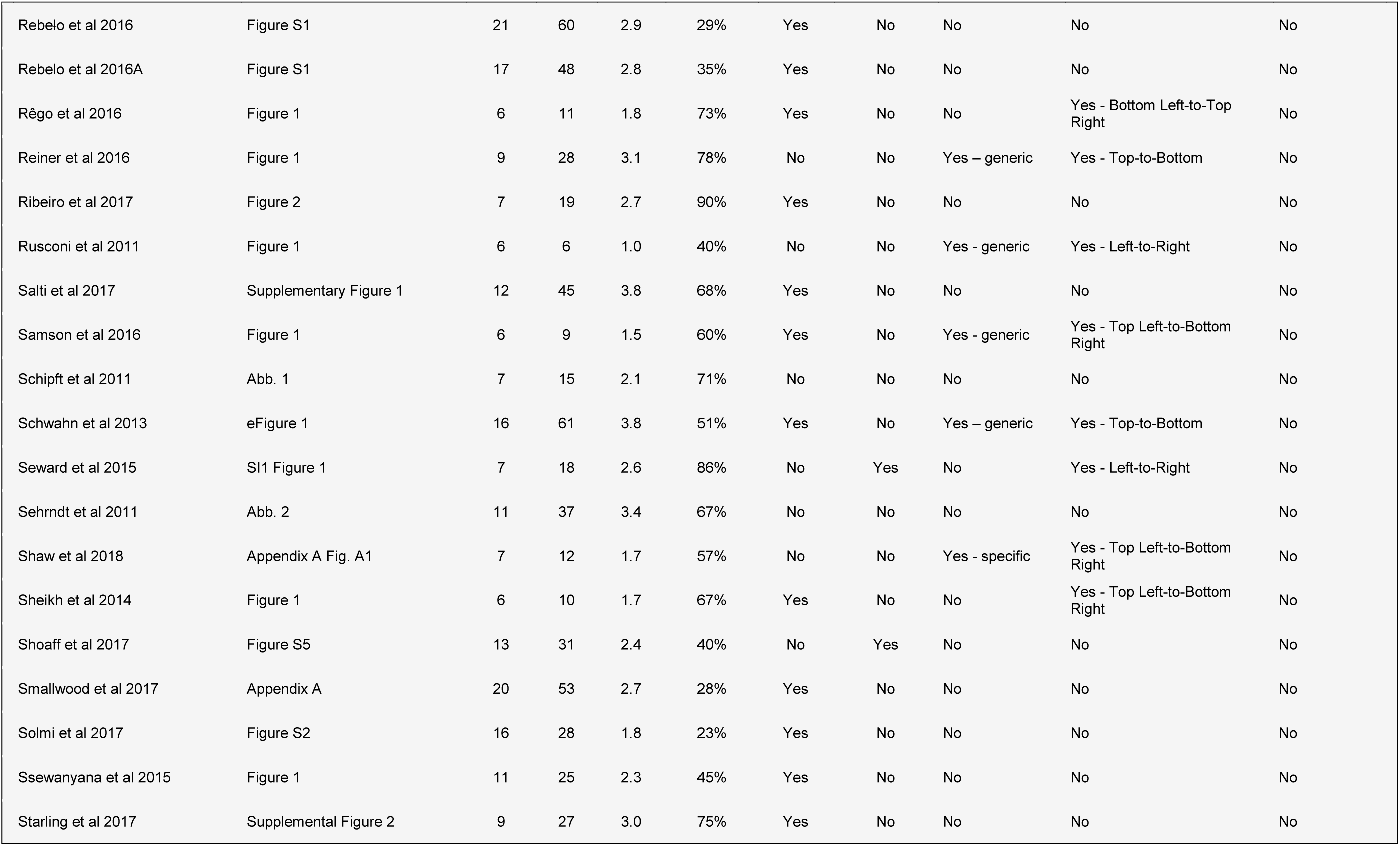
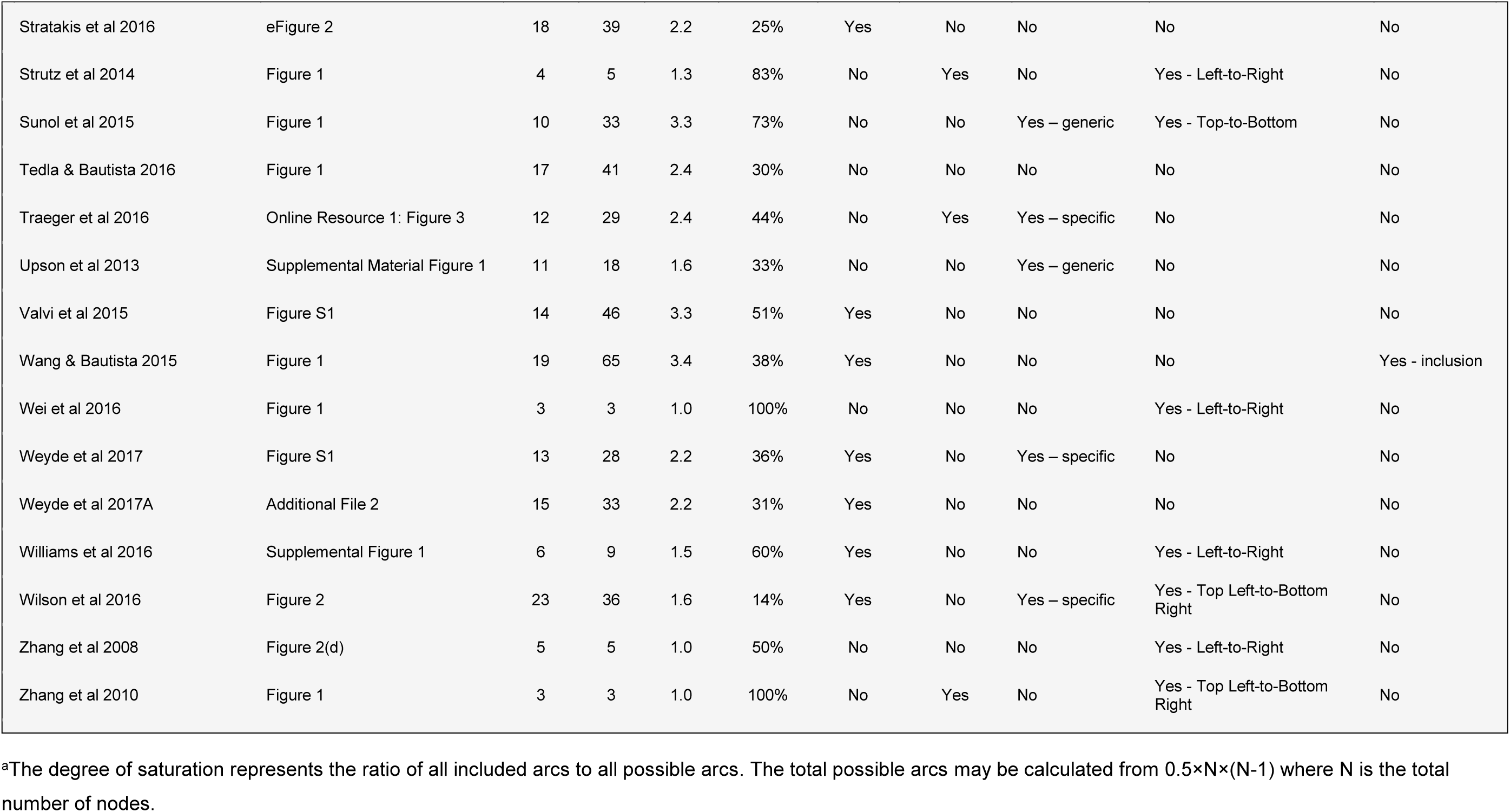
Summary details of the largest causal diagram reported in the 144 studies with at least one diagram available

**Supplementary Table 3.** Relationship between common diagram features and probability of data extraction error by the first data extractor.

| **Continuous variable** | **Unit** | **Relative probability of error (95% CI)** |
| --- | --- | --- |
| Total arcs | (per arc) | 1.016 (1.008, 1.024) |
| Total nodes | (per node) | 1.042 (1.011, 1.075) |
| **Categorical variable** | **Category** | **Model marginal probability (%) of error<sup>a</sup> (95% CI)** |
| Drawn in DAGitty | No | 50.1 (37.7, 62.6) |
|  | Yes | 32.4 (23.2, 41.8) |
| Arrangement of arcs | No consistent direction | 40.7 (31.1, 50.2) |
|  | Top-to-bottom or left-to-right | 29.5 (11.1, 48.0) |
|  | Corner-to-corner | 43.9 (19.7, 68.1) |
| Includes unobserved variables | No | 37.5 (27.9, 47.1) |
|  | Yes | 42.4 (29.1, 55.8) |
| Includes super-nodes<sup>b</sup> | No | 39.3 (30.9, 47.8) |
|  | Yes | 38.8 (17.8, 59.7) |

<sup>a</sup>Adjusted for total number of nodes and arcs <sup>b</sup>Nodes containing more than one variable

**Supplementary Table 4.**
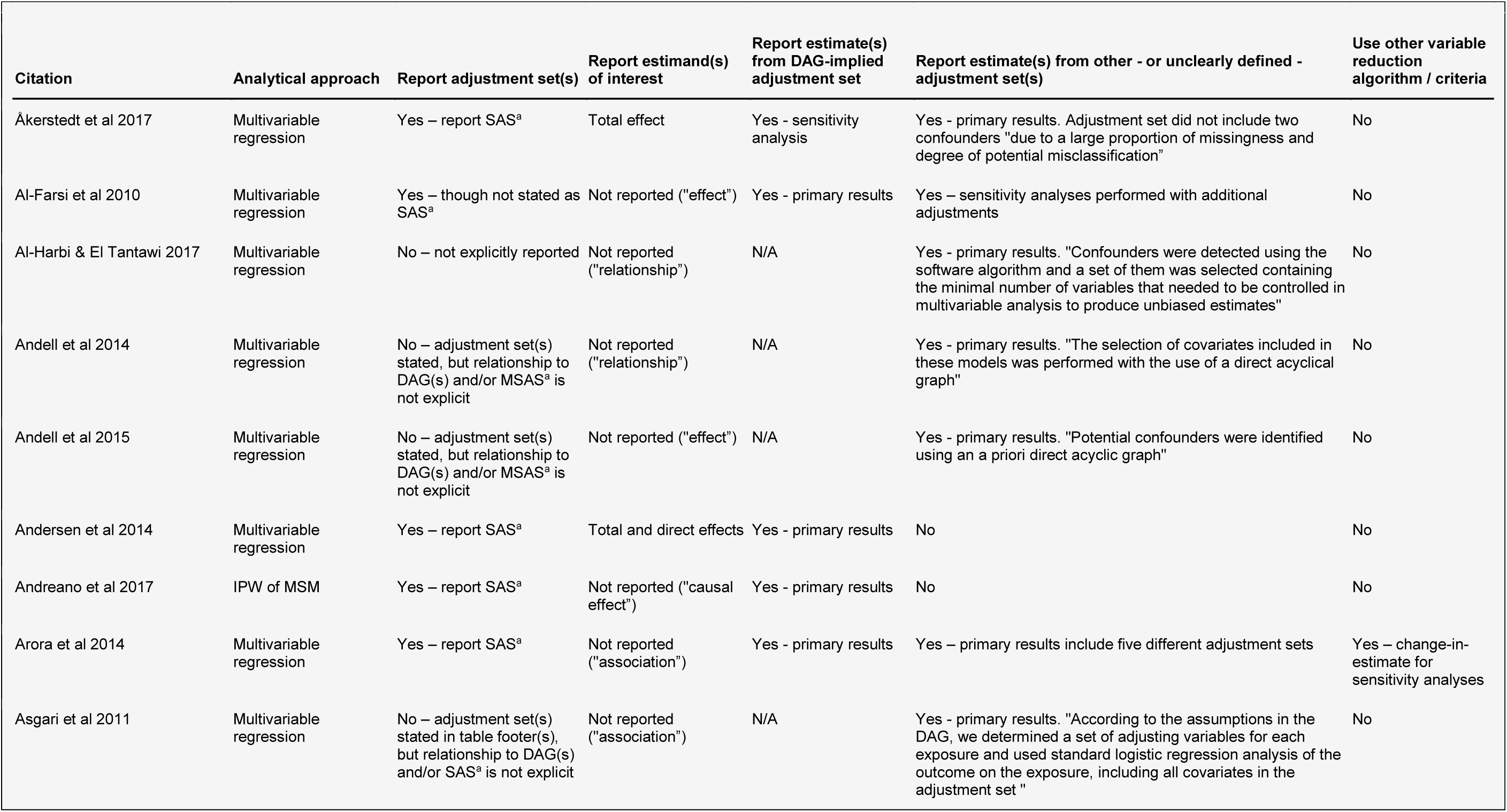
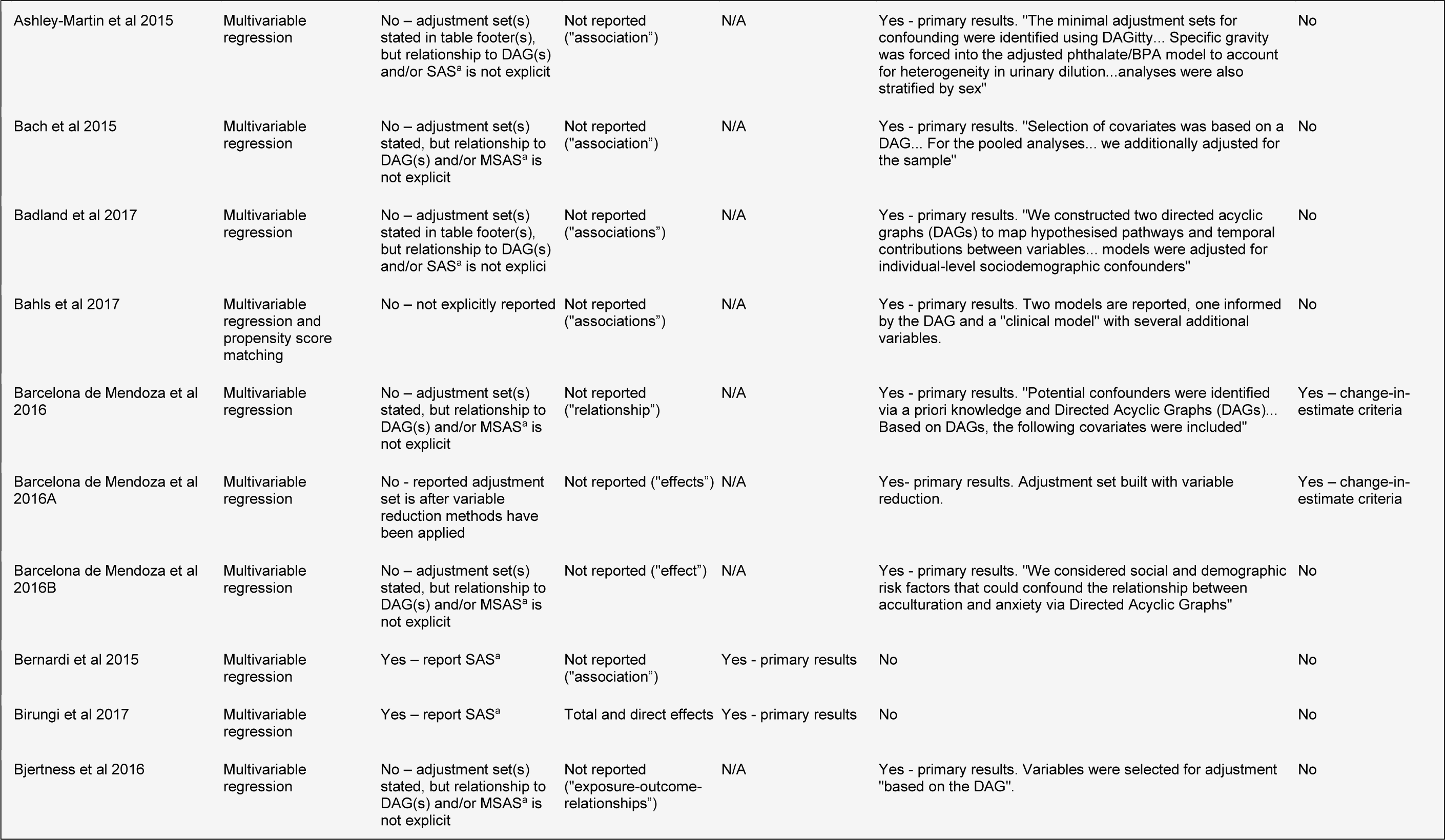
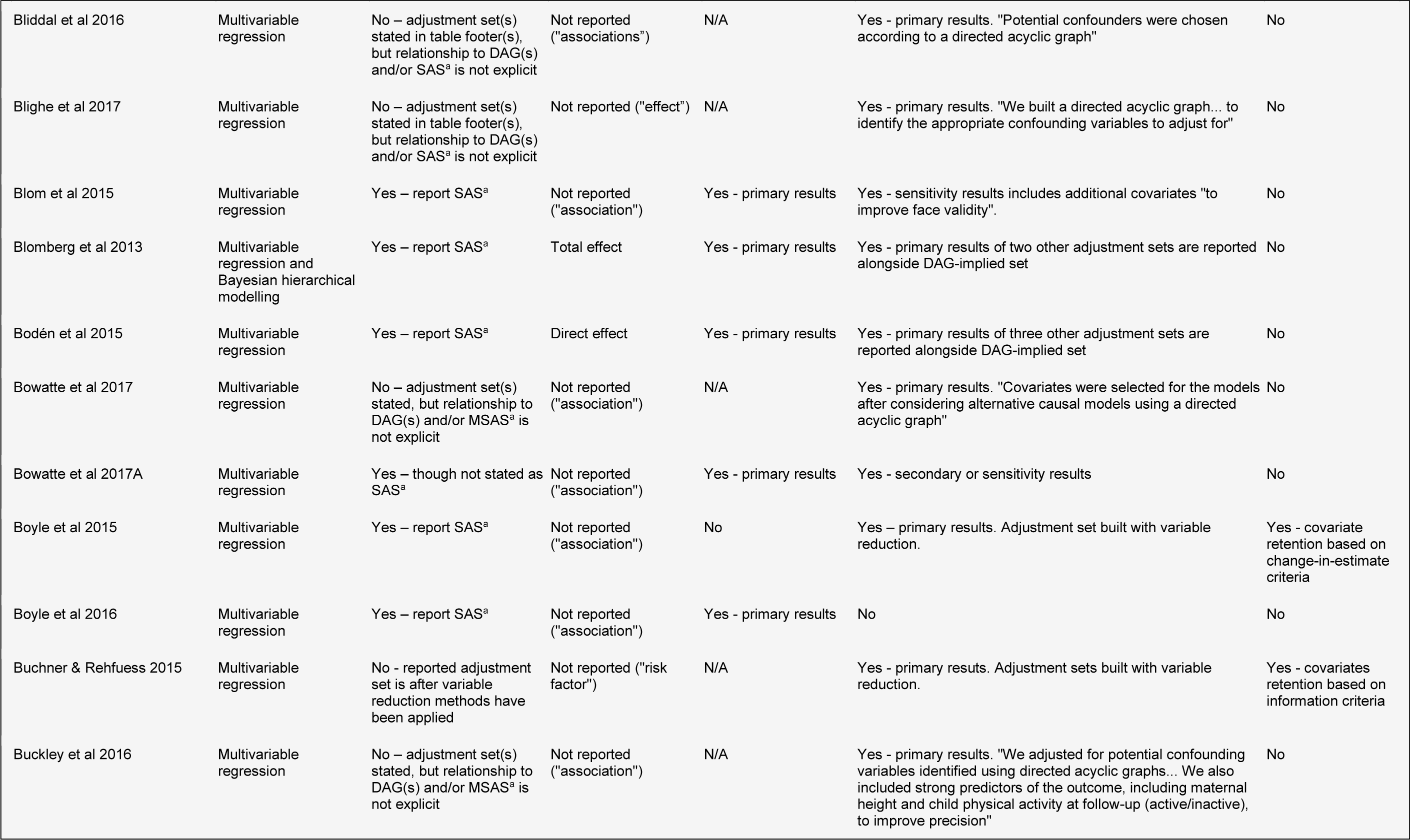
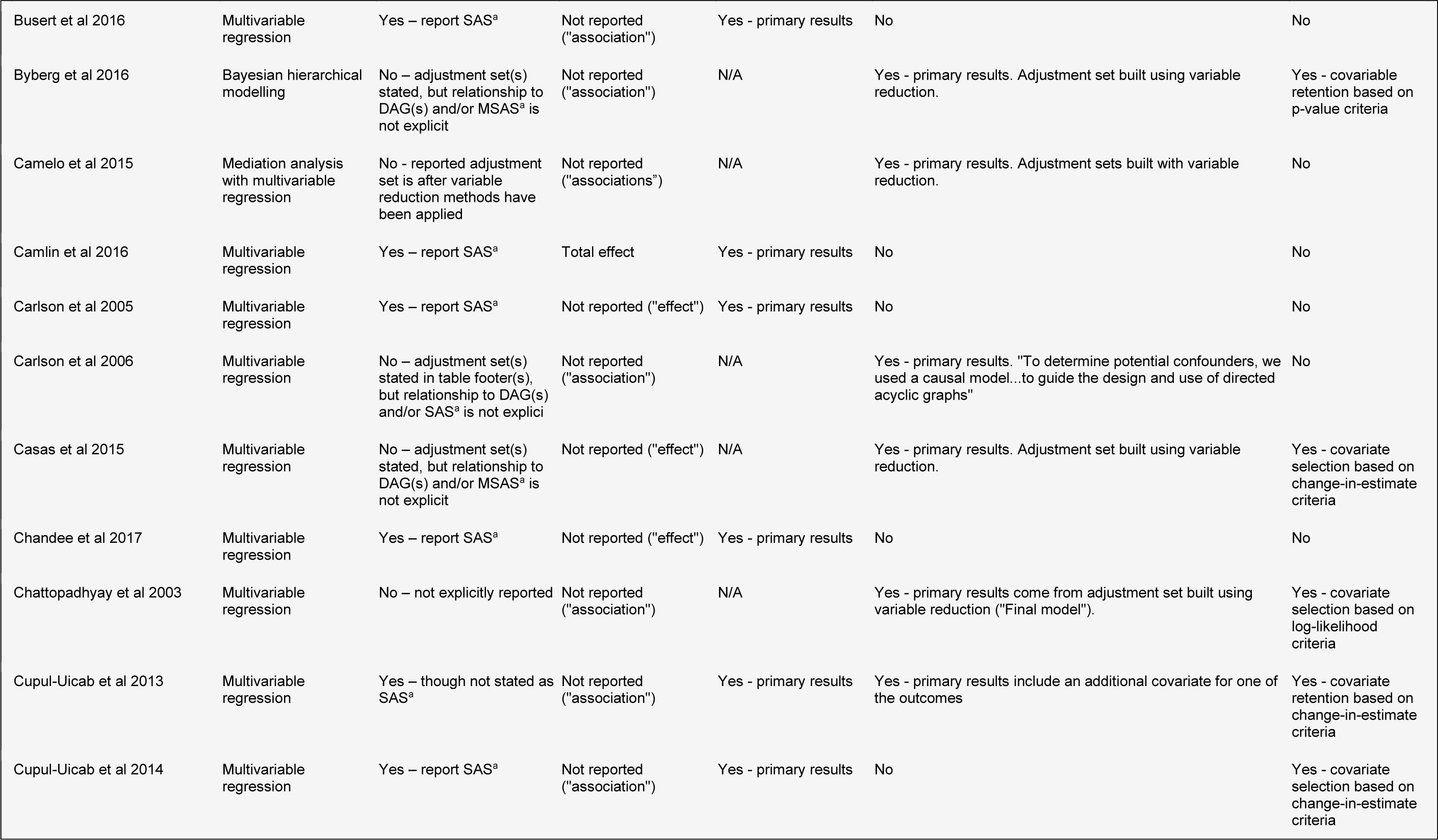
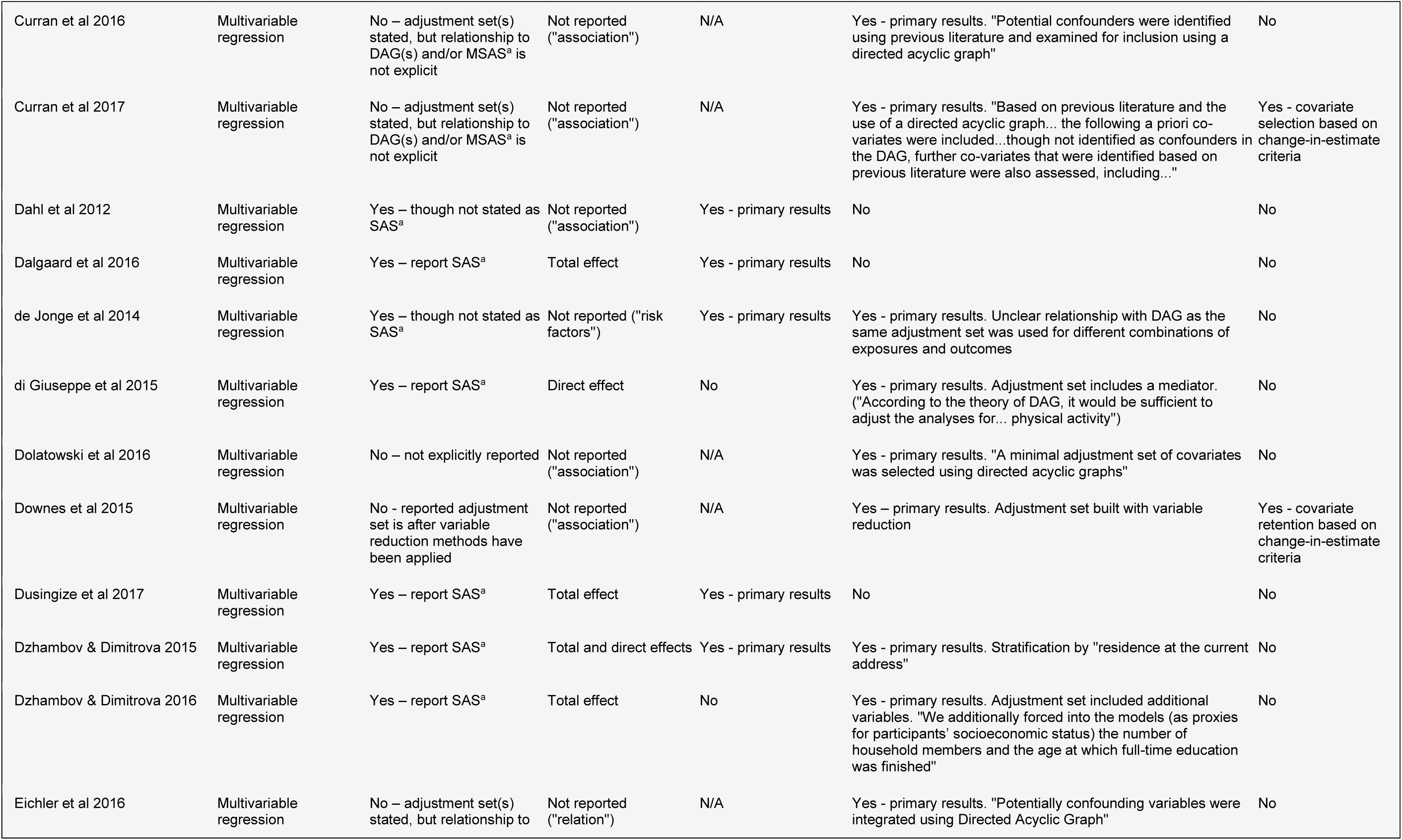
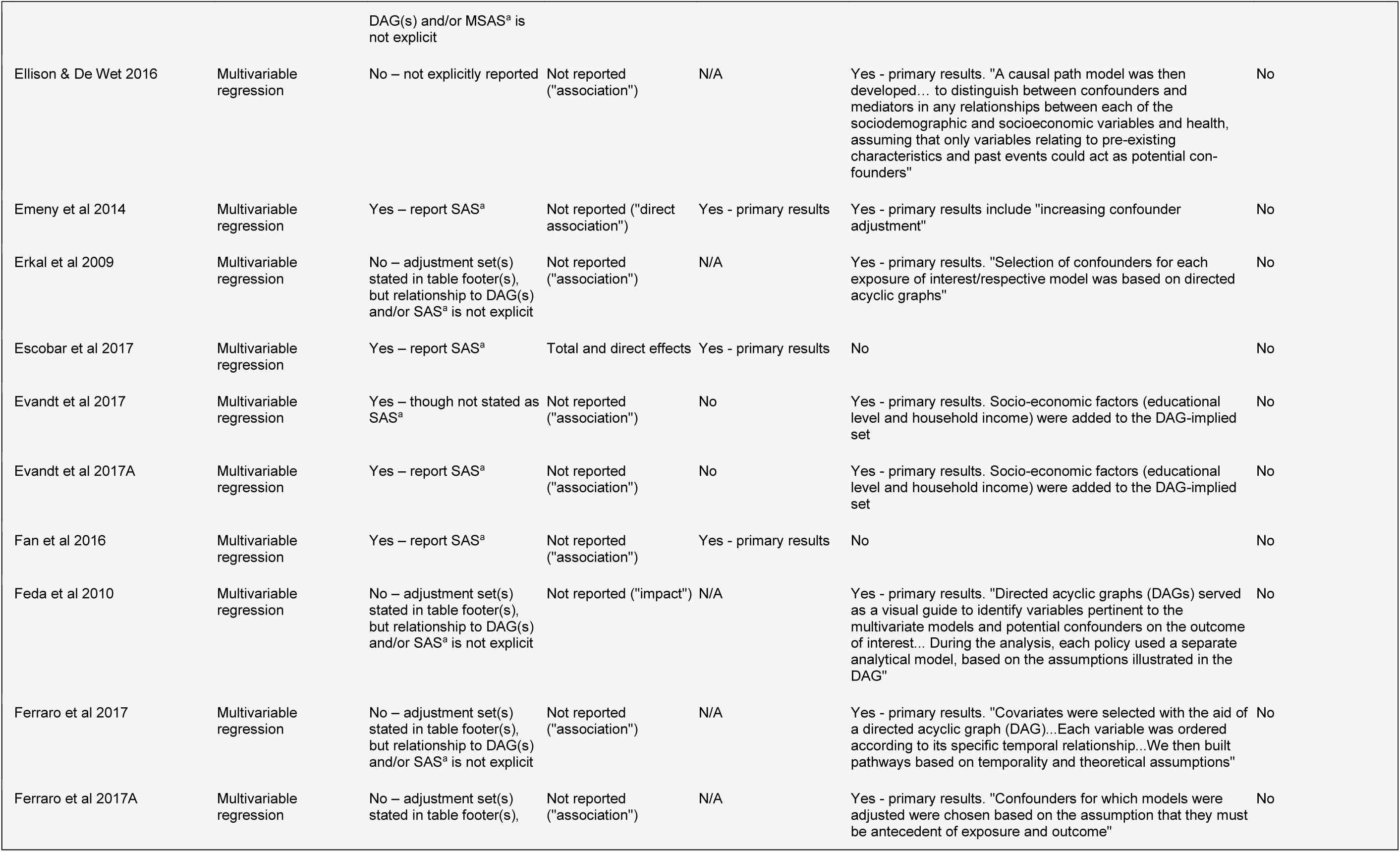
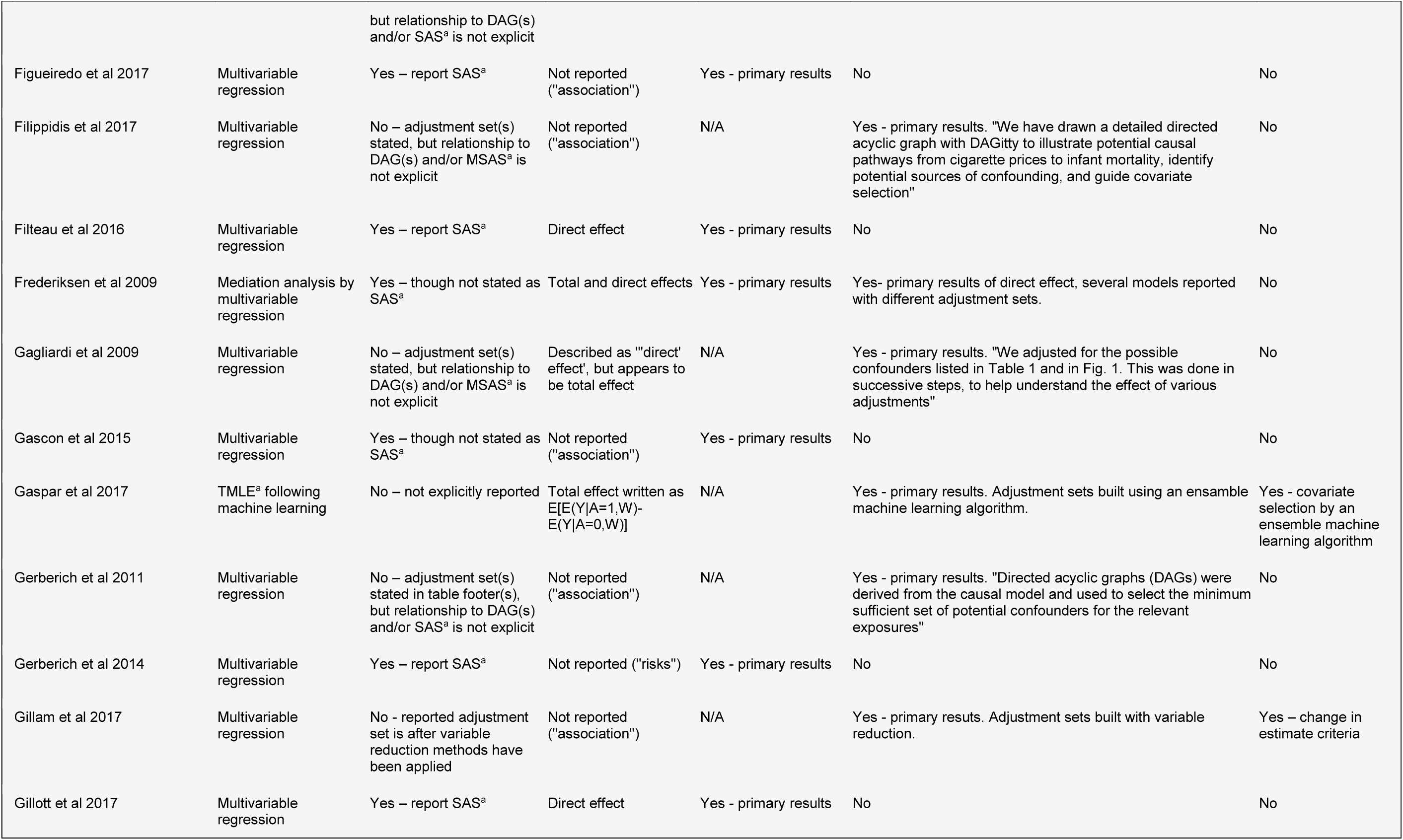
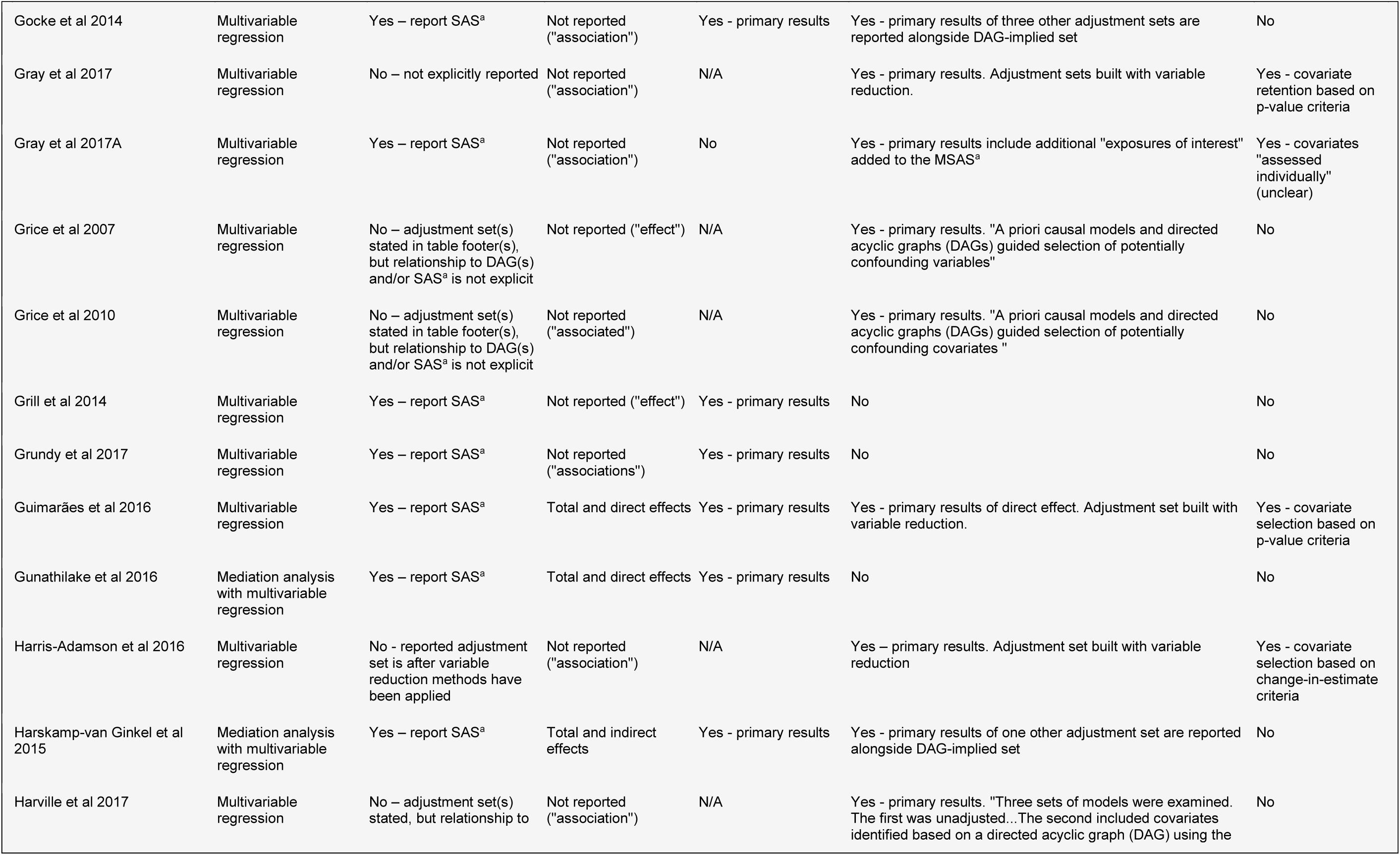
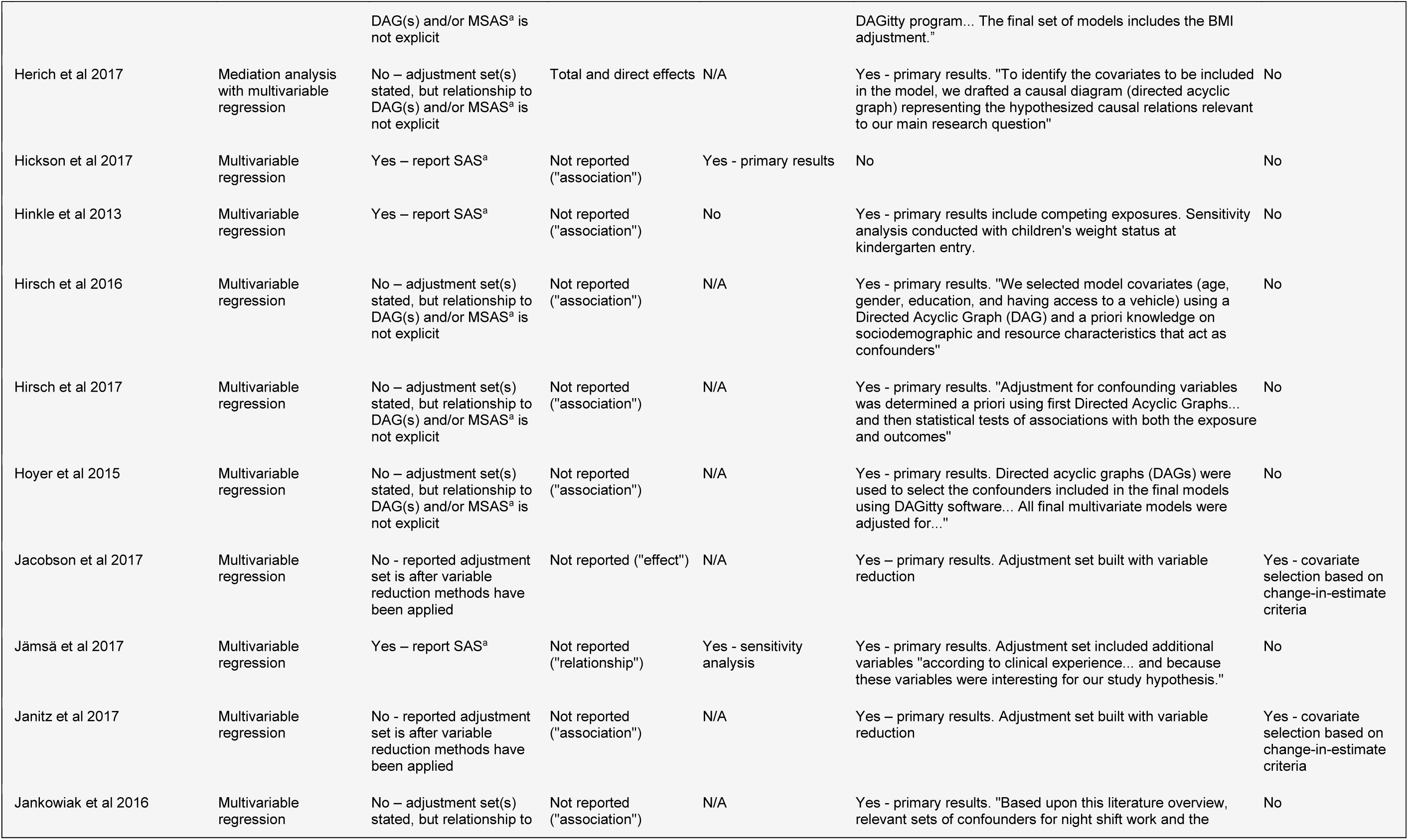
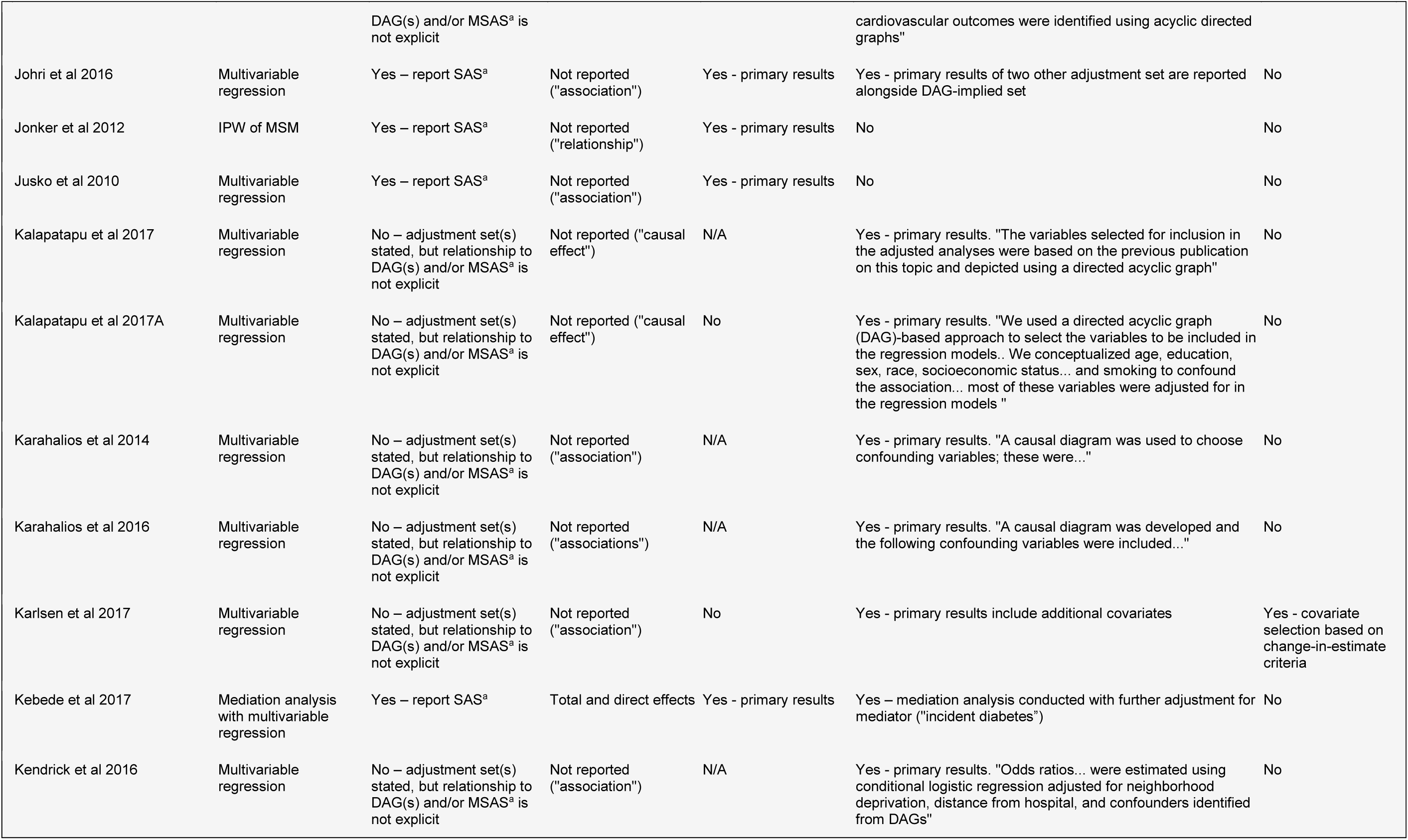
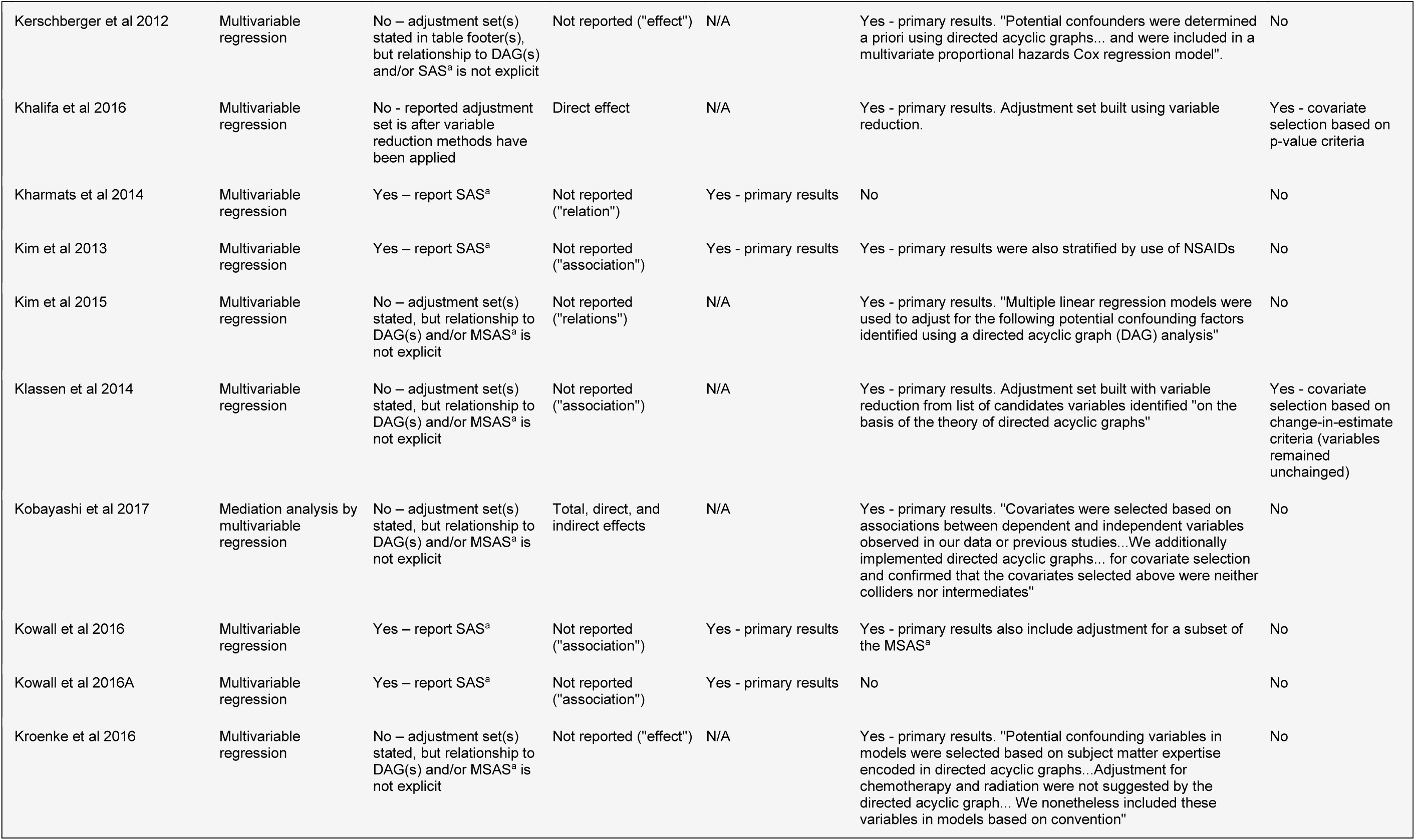
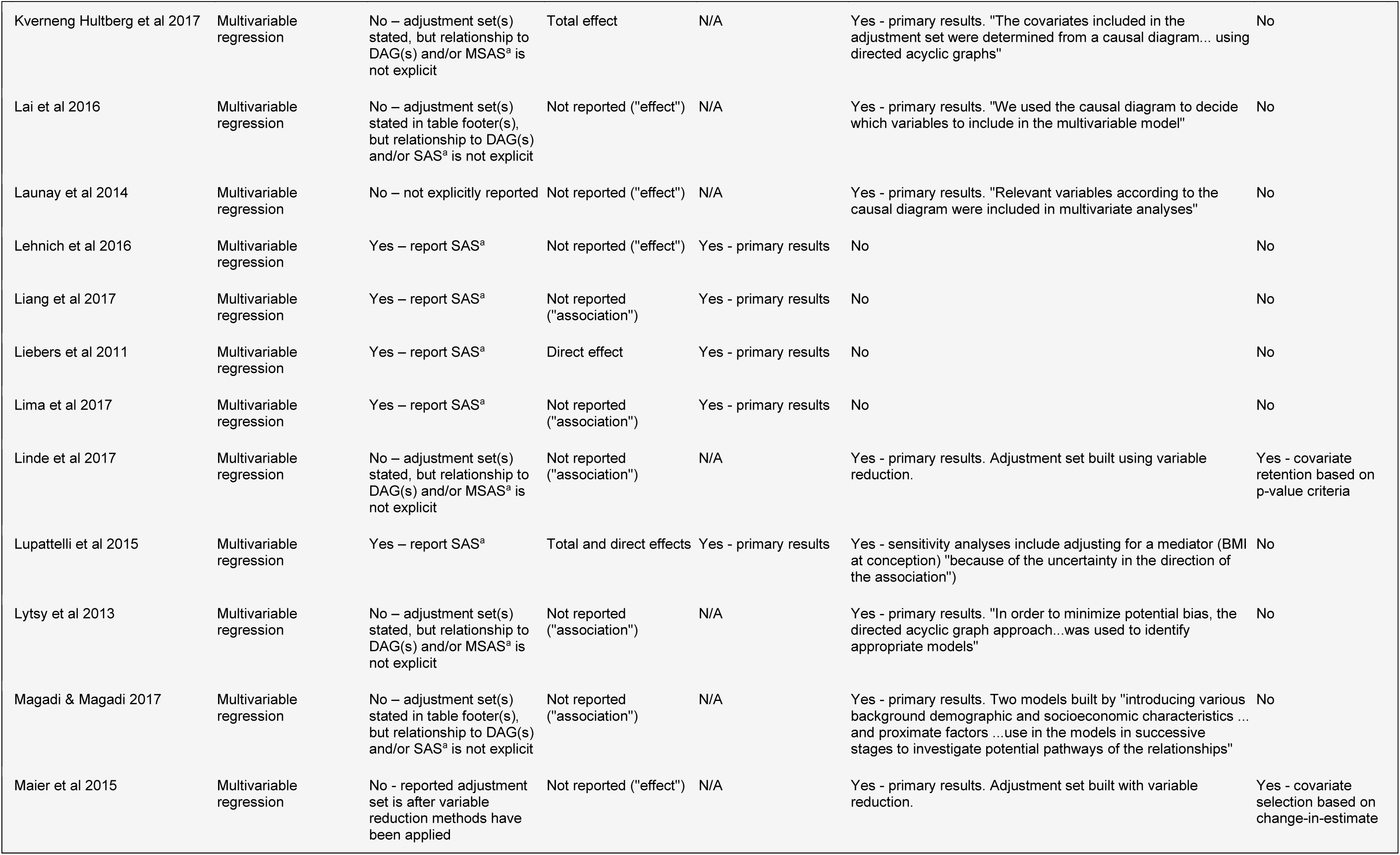
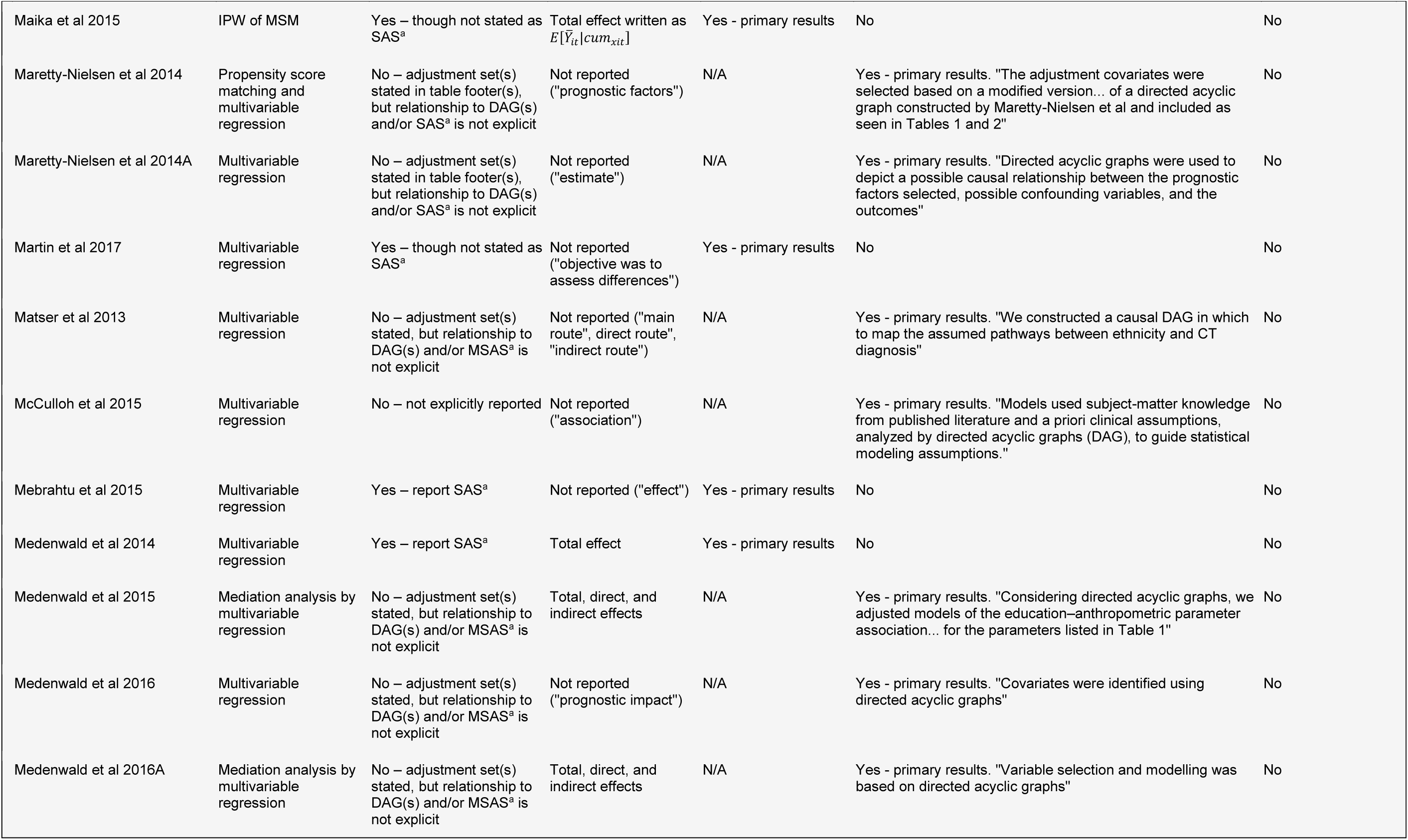
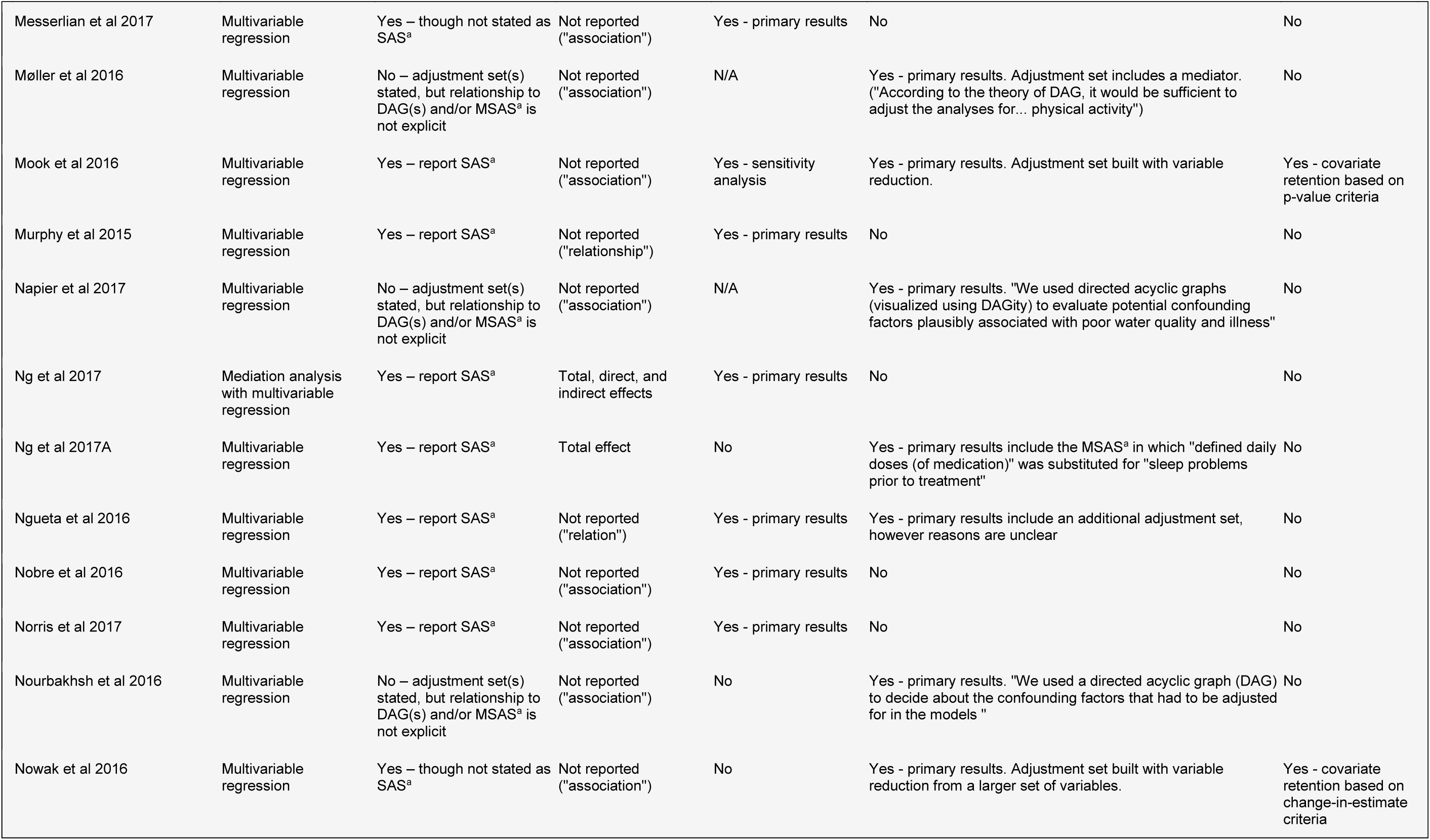
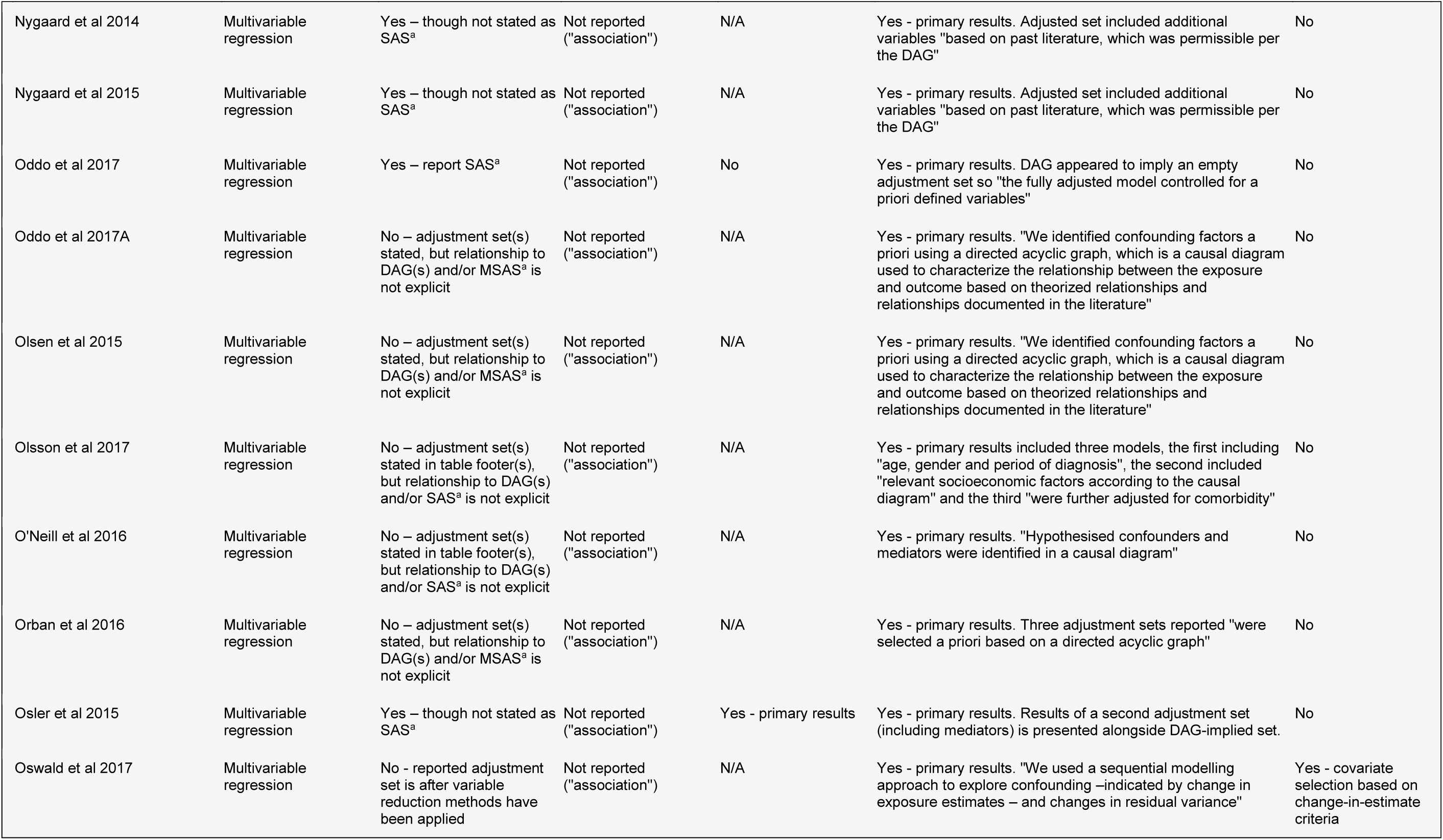
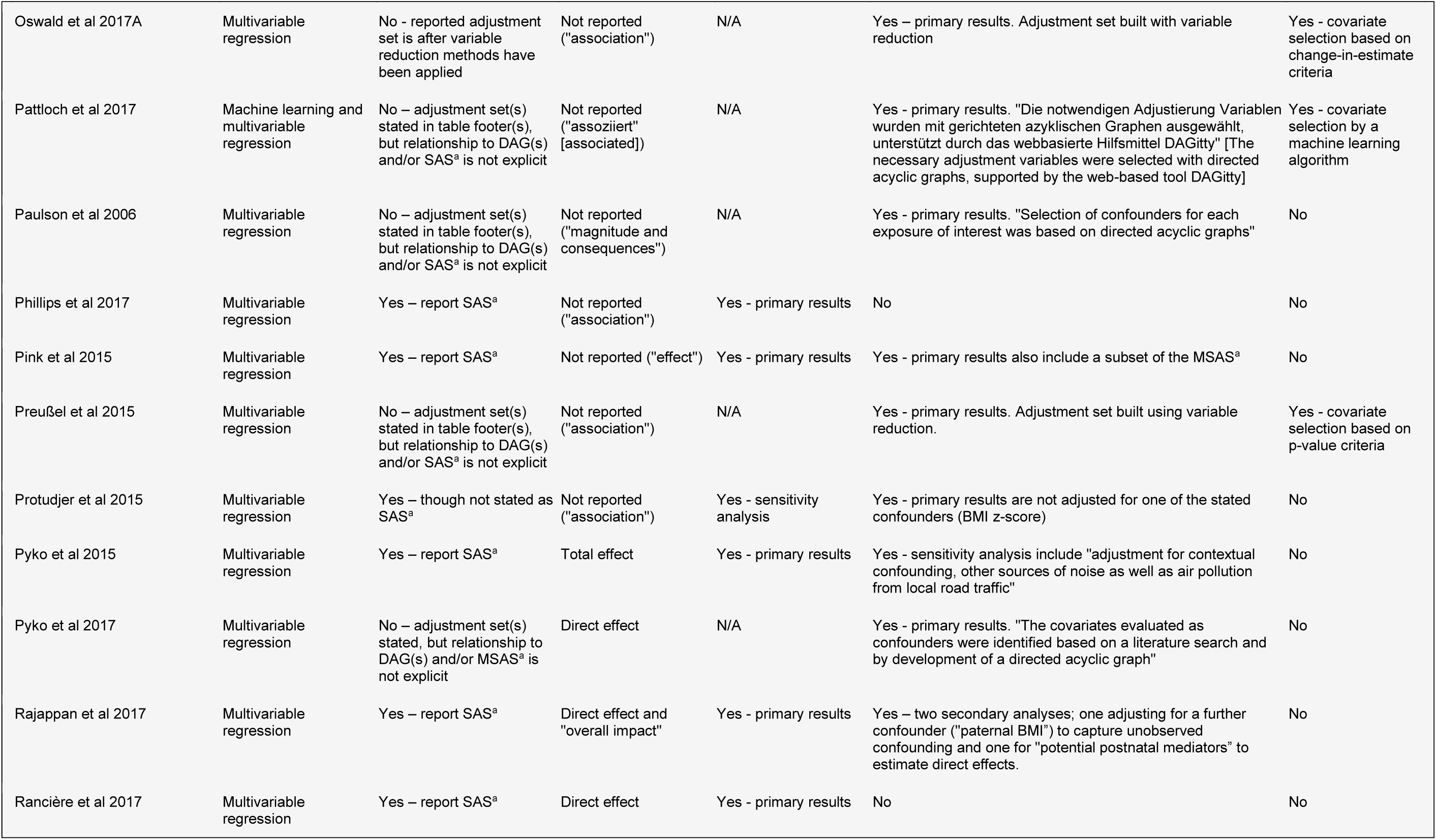
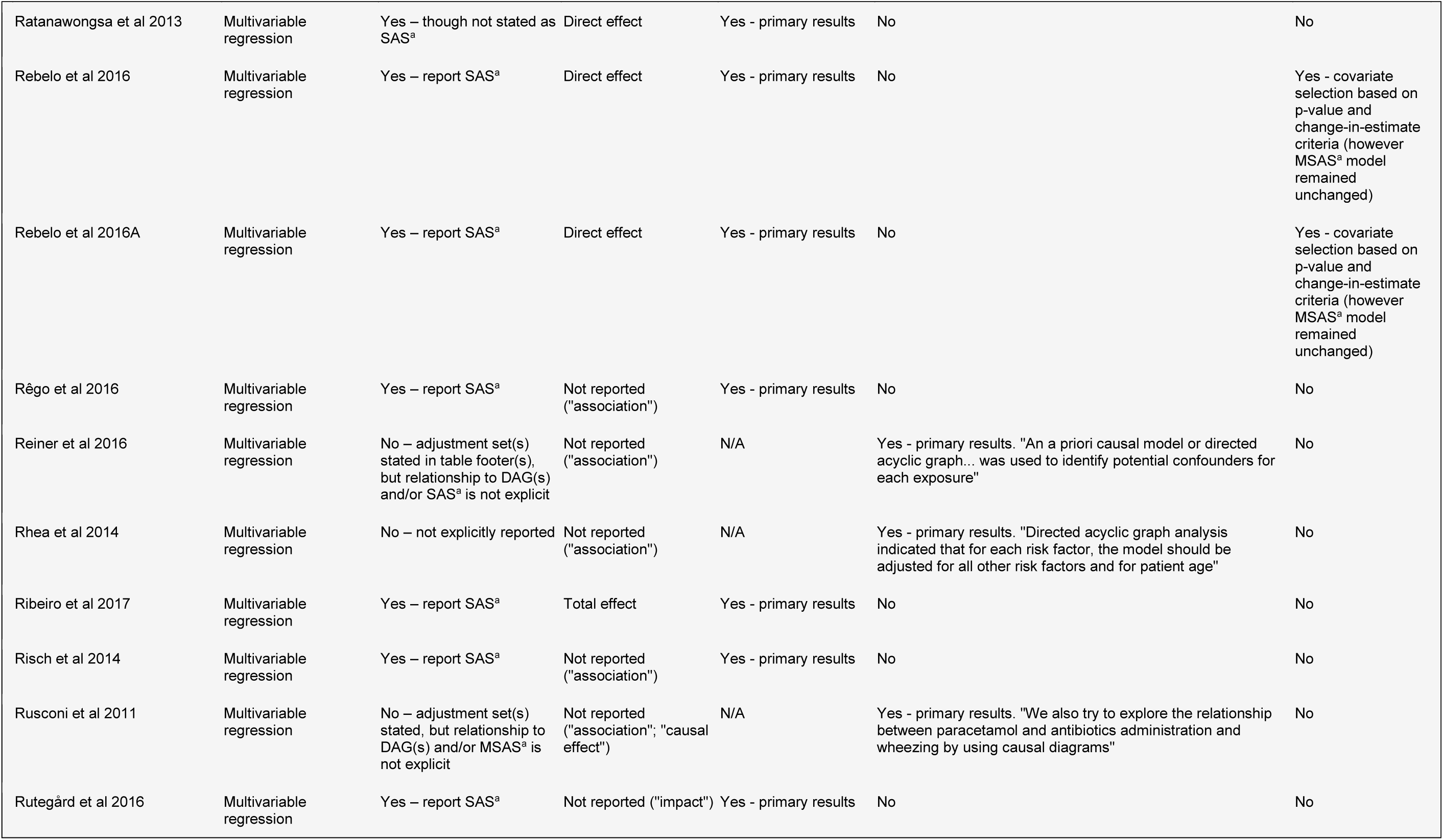
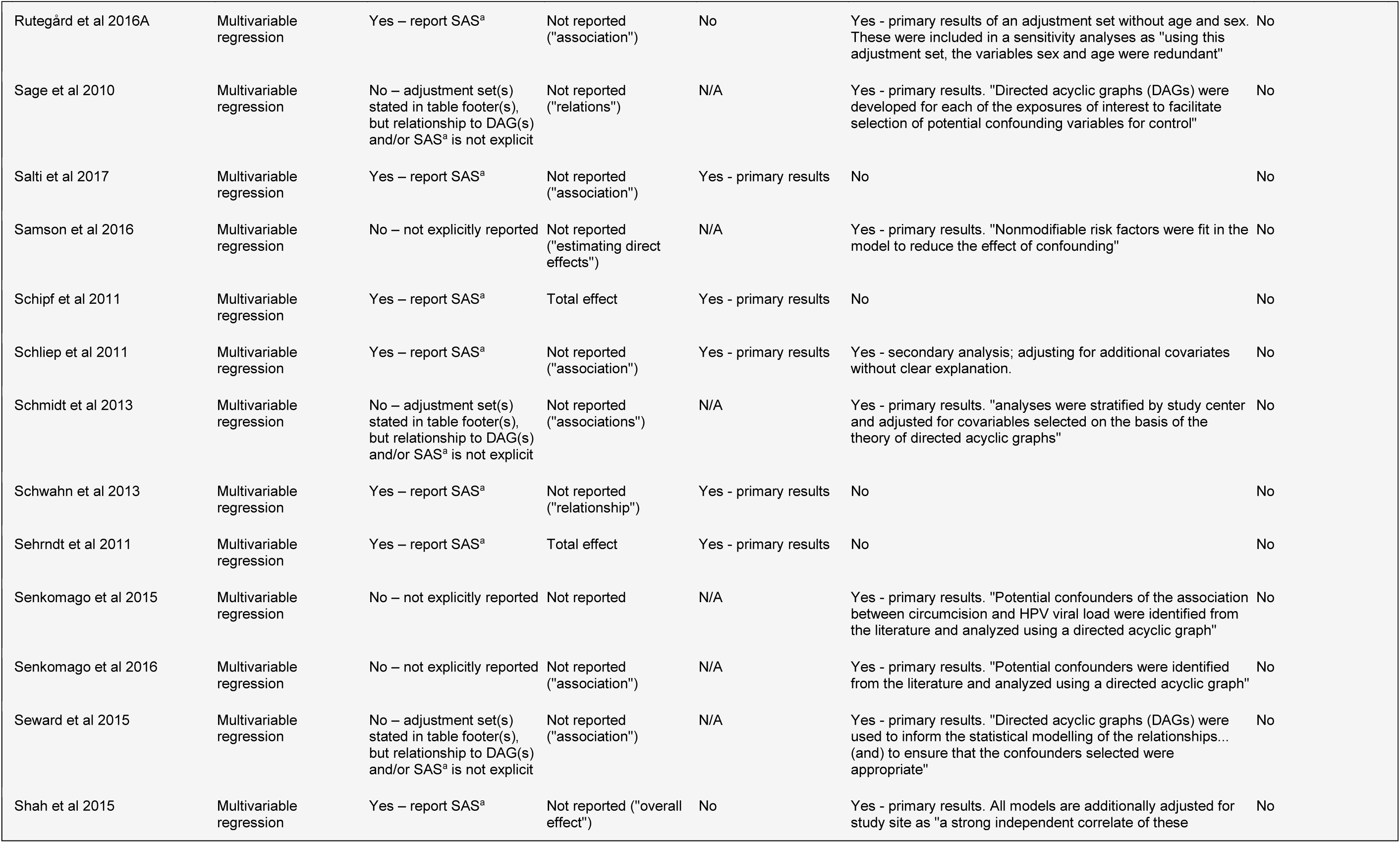
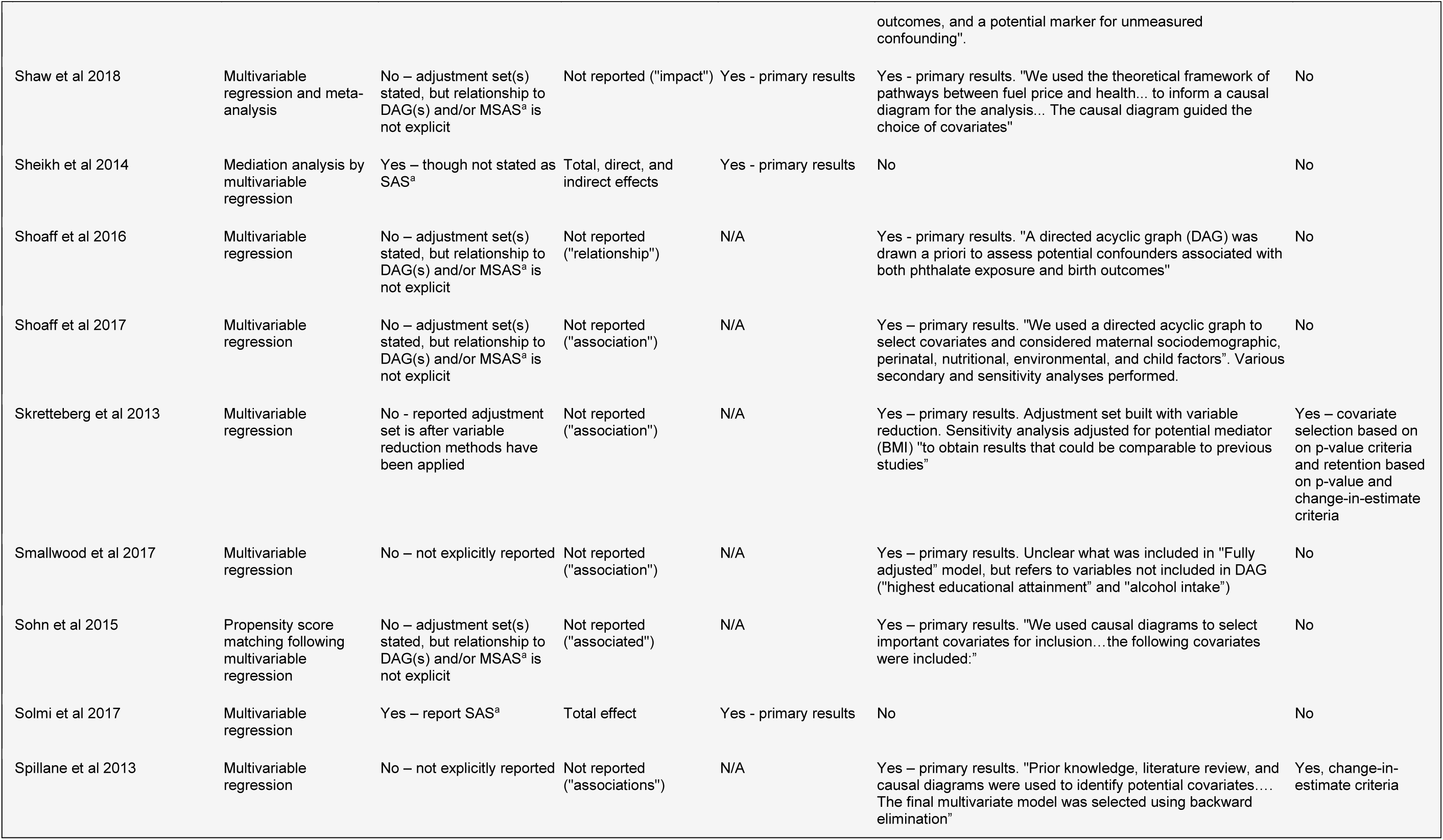
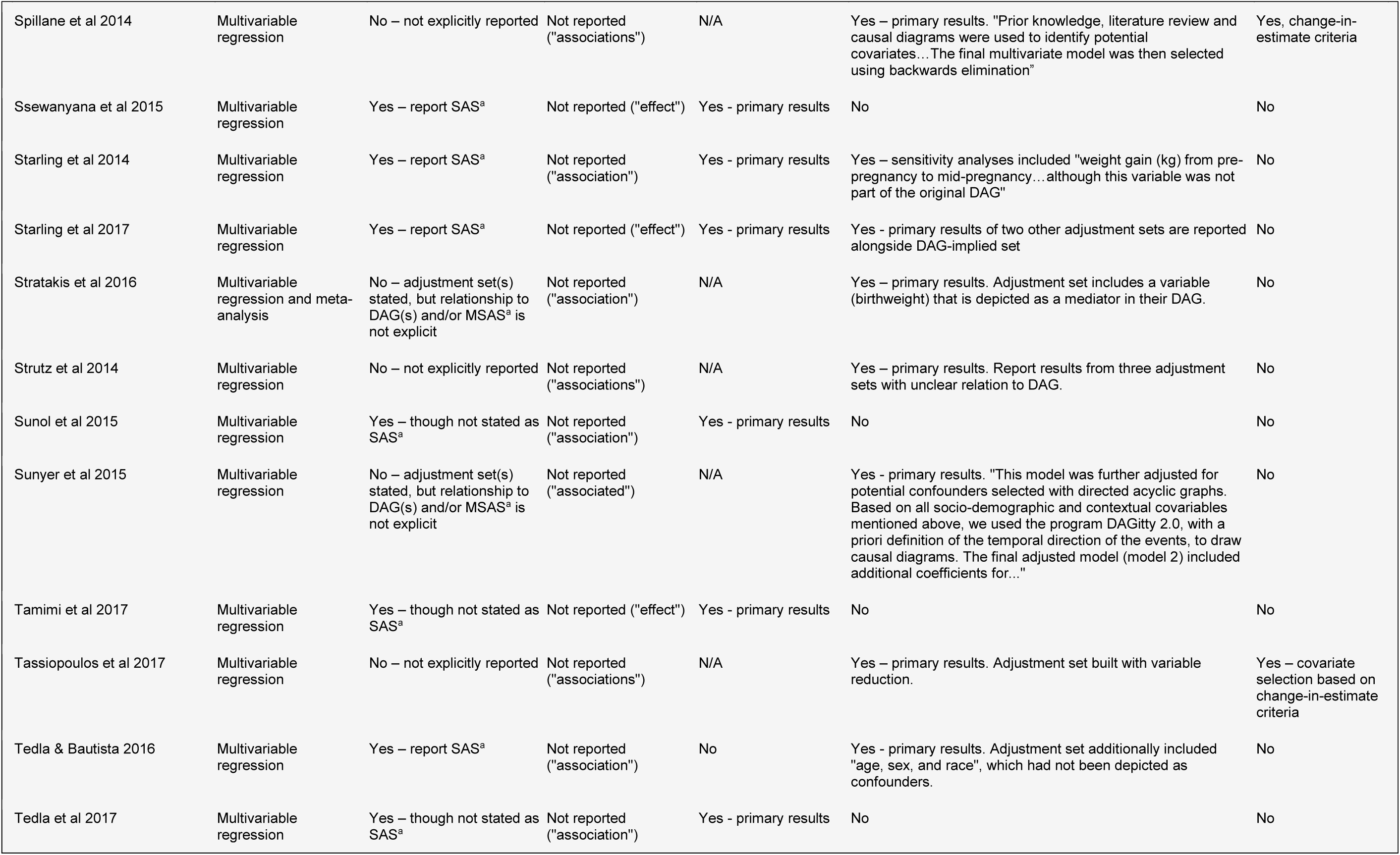
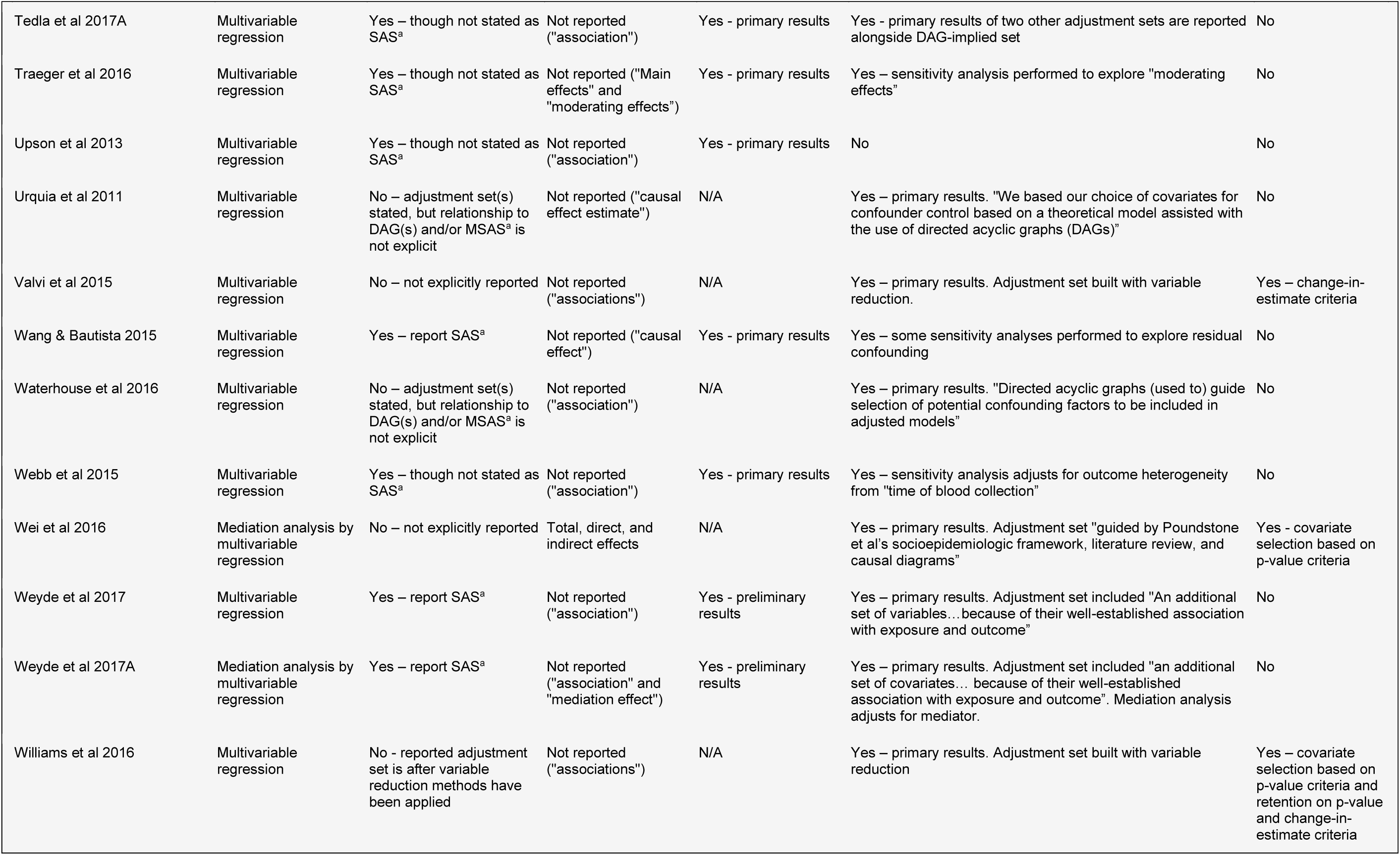
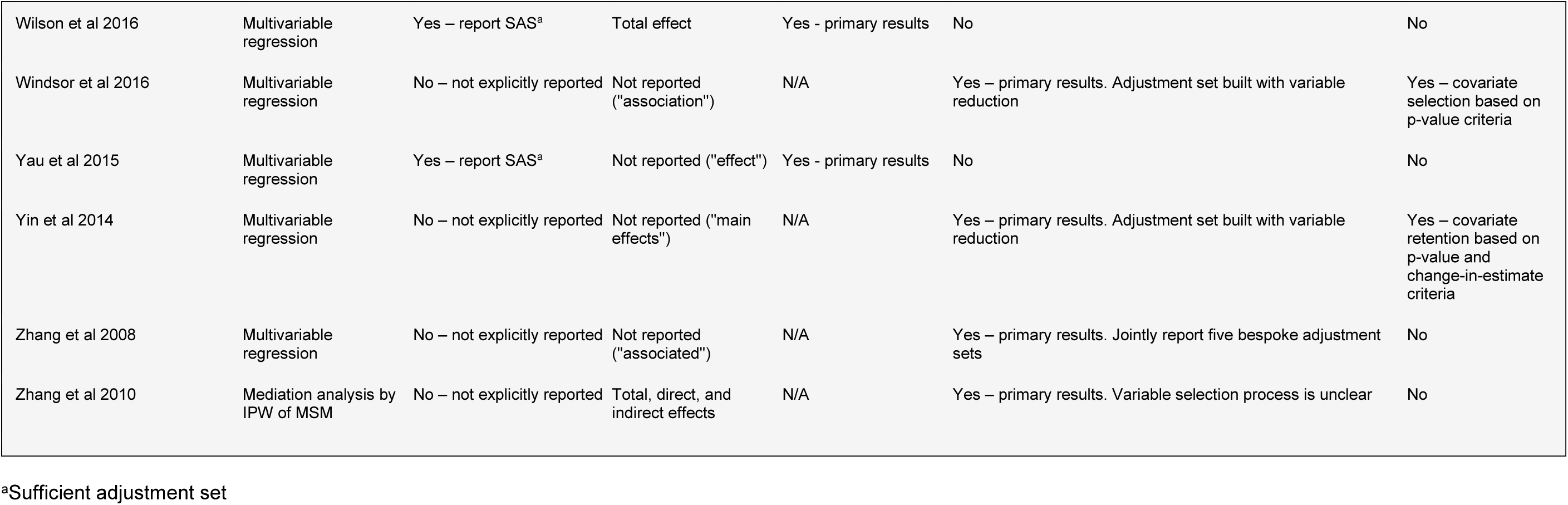
Summary details of the analytical approach, estimand(s) of interest, and adjustment set(s) used by the 234 articles included in the review

**Supplementary Table 5.**
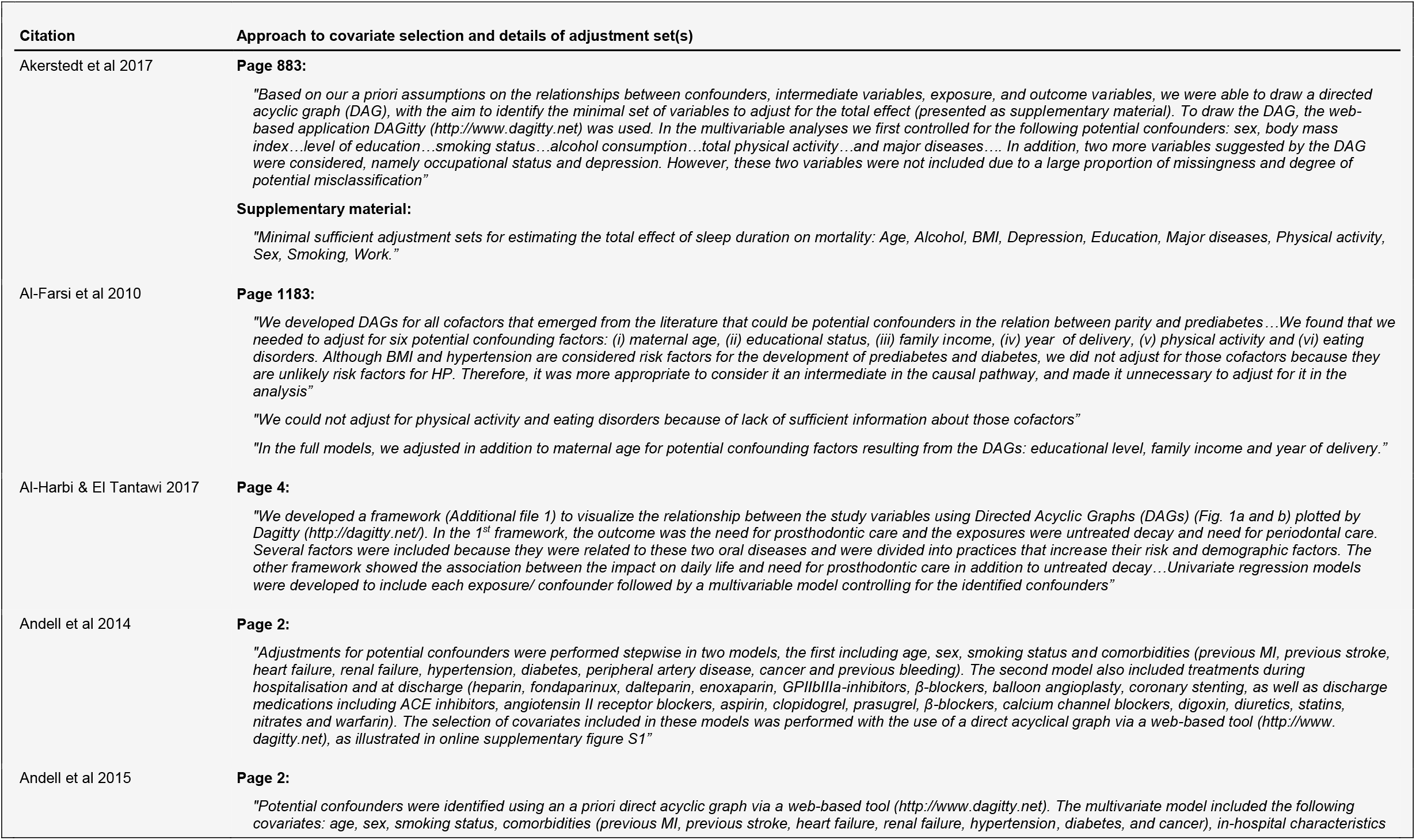
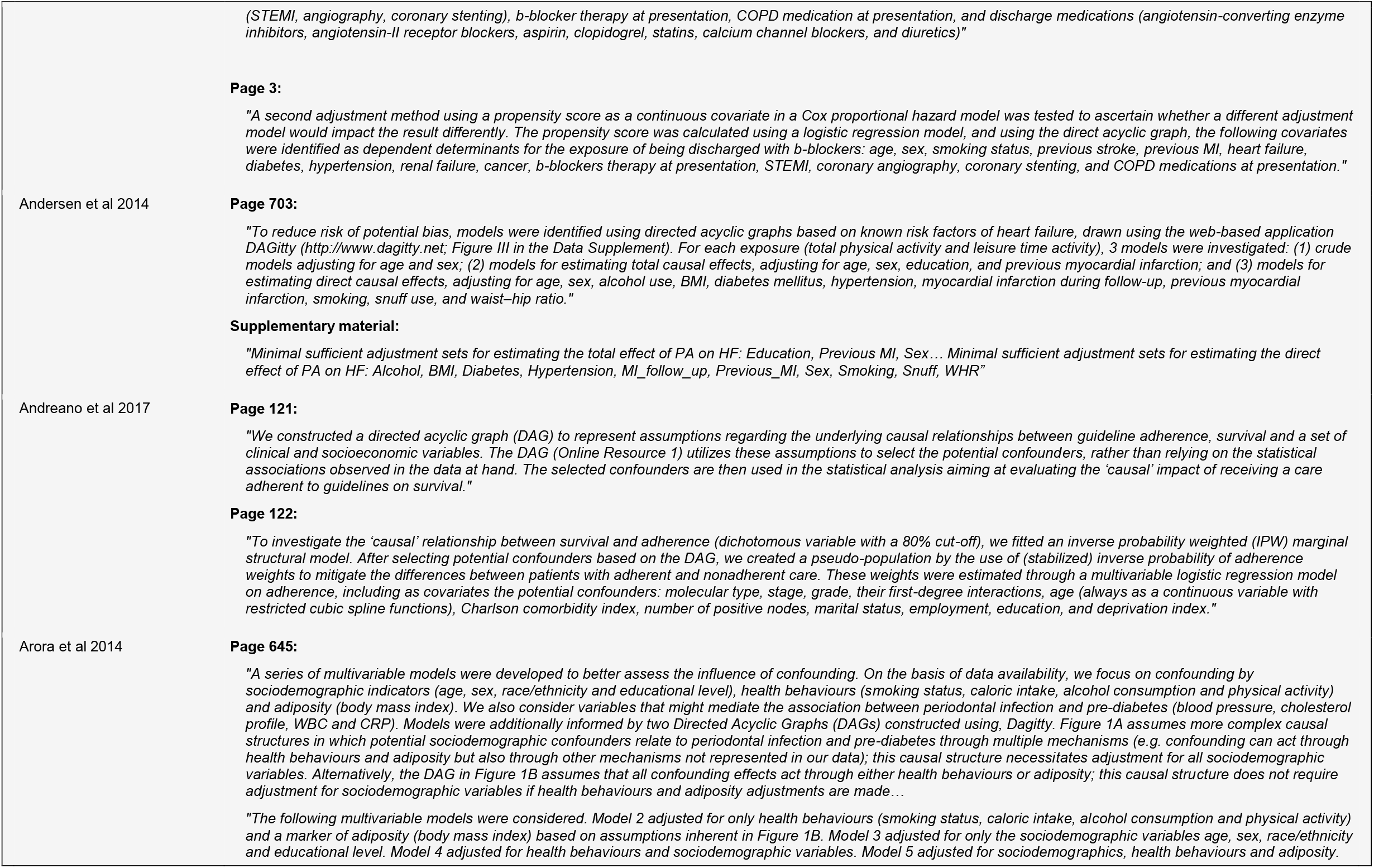
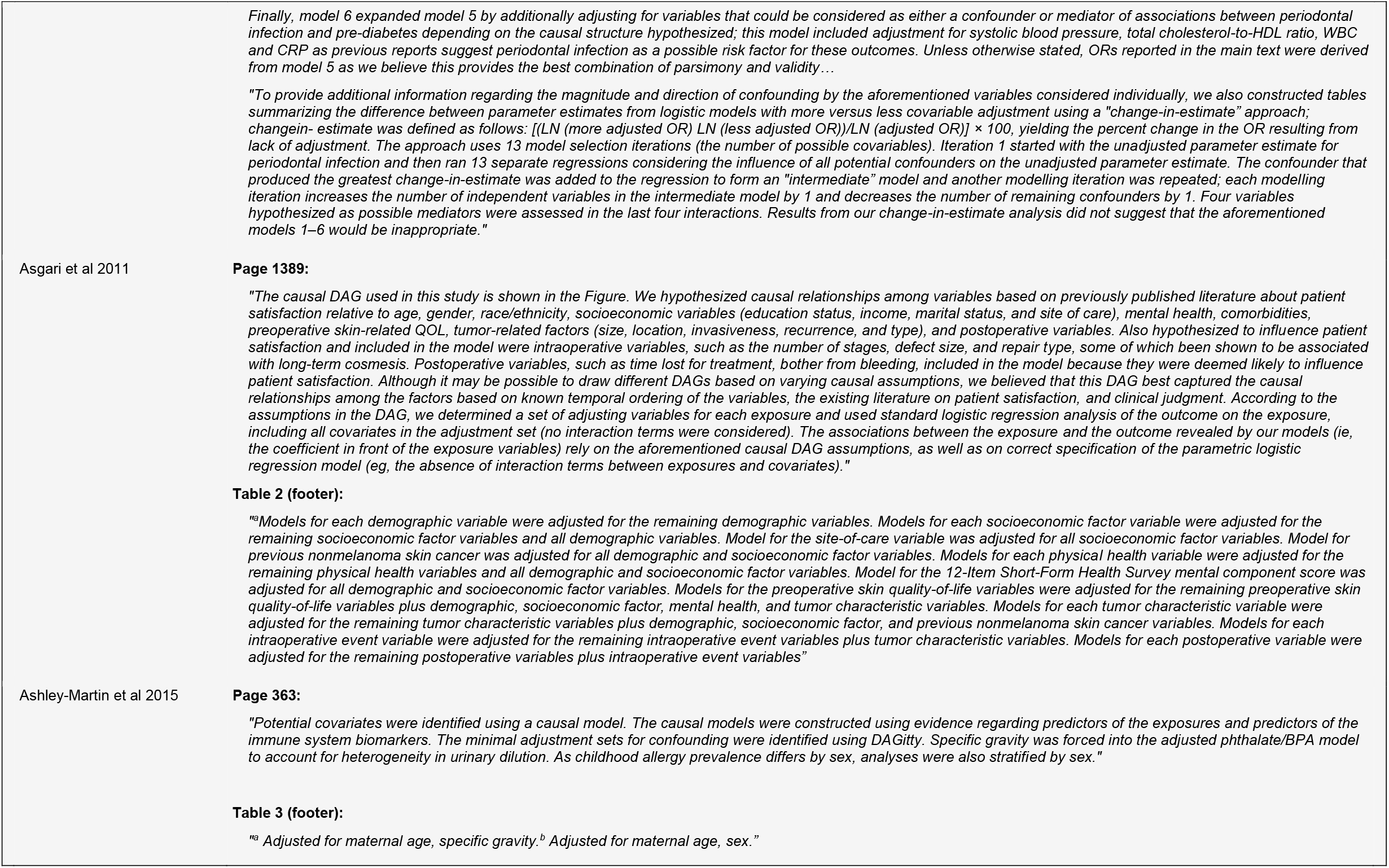
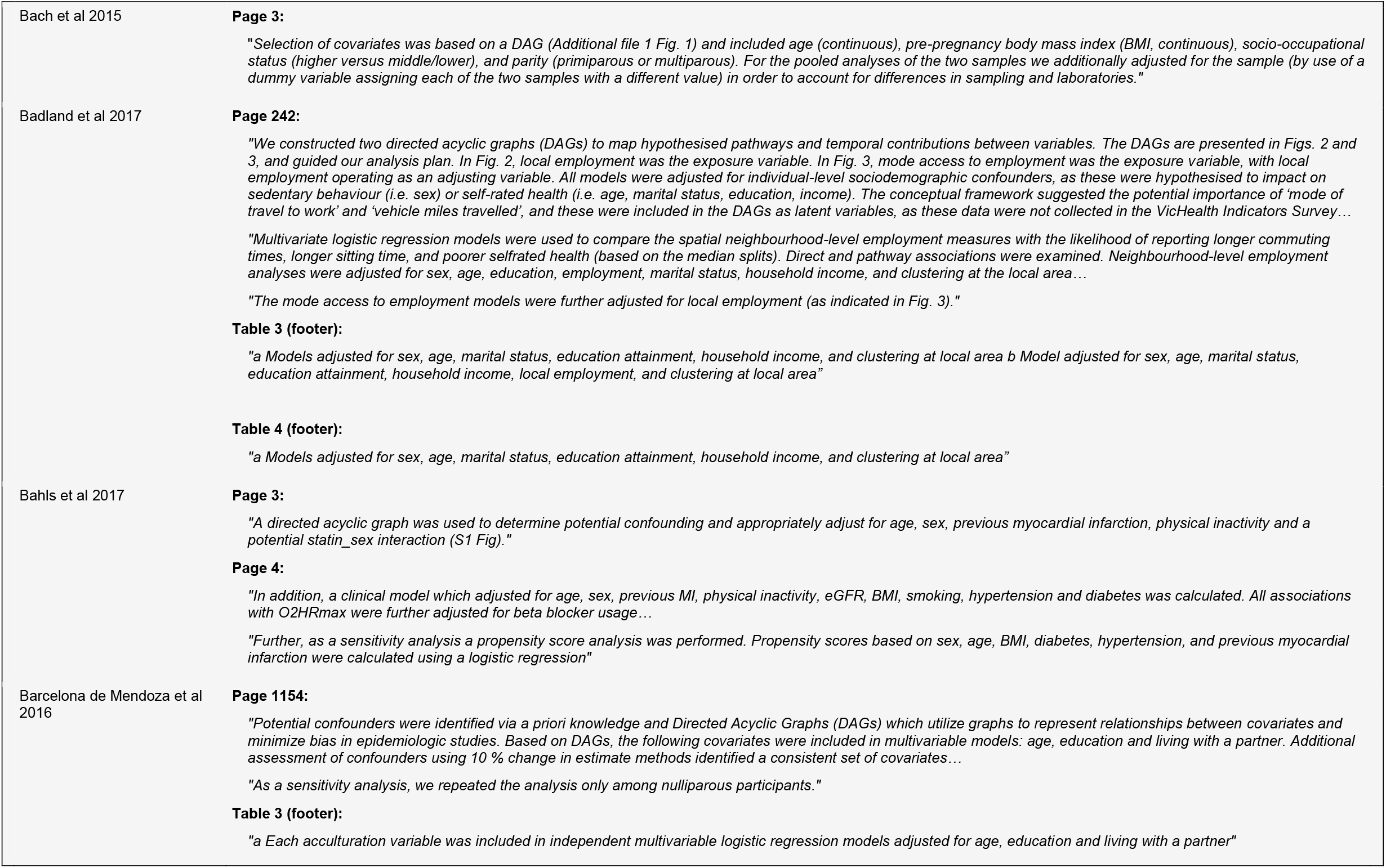
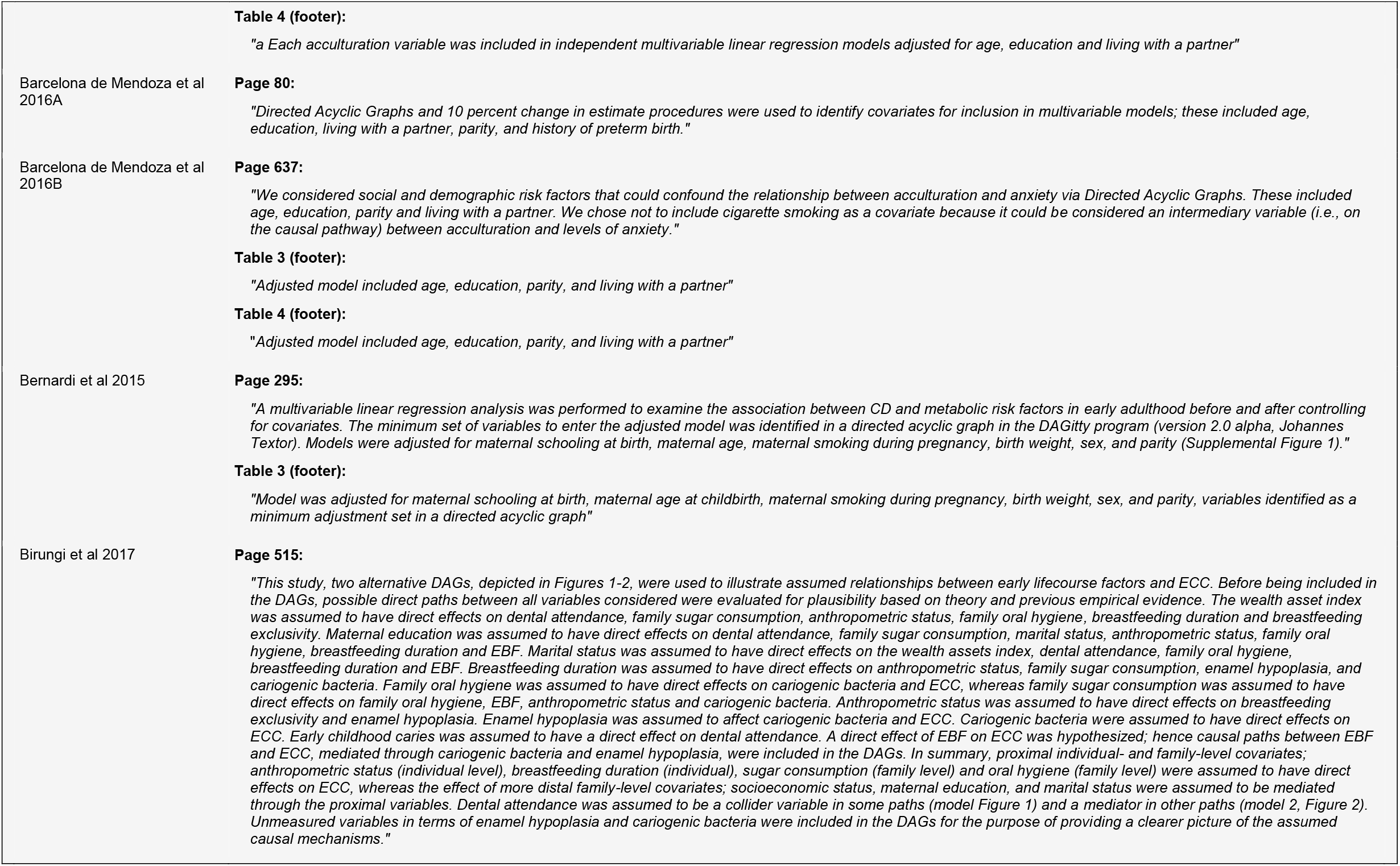
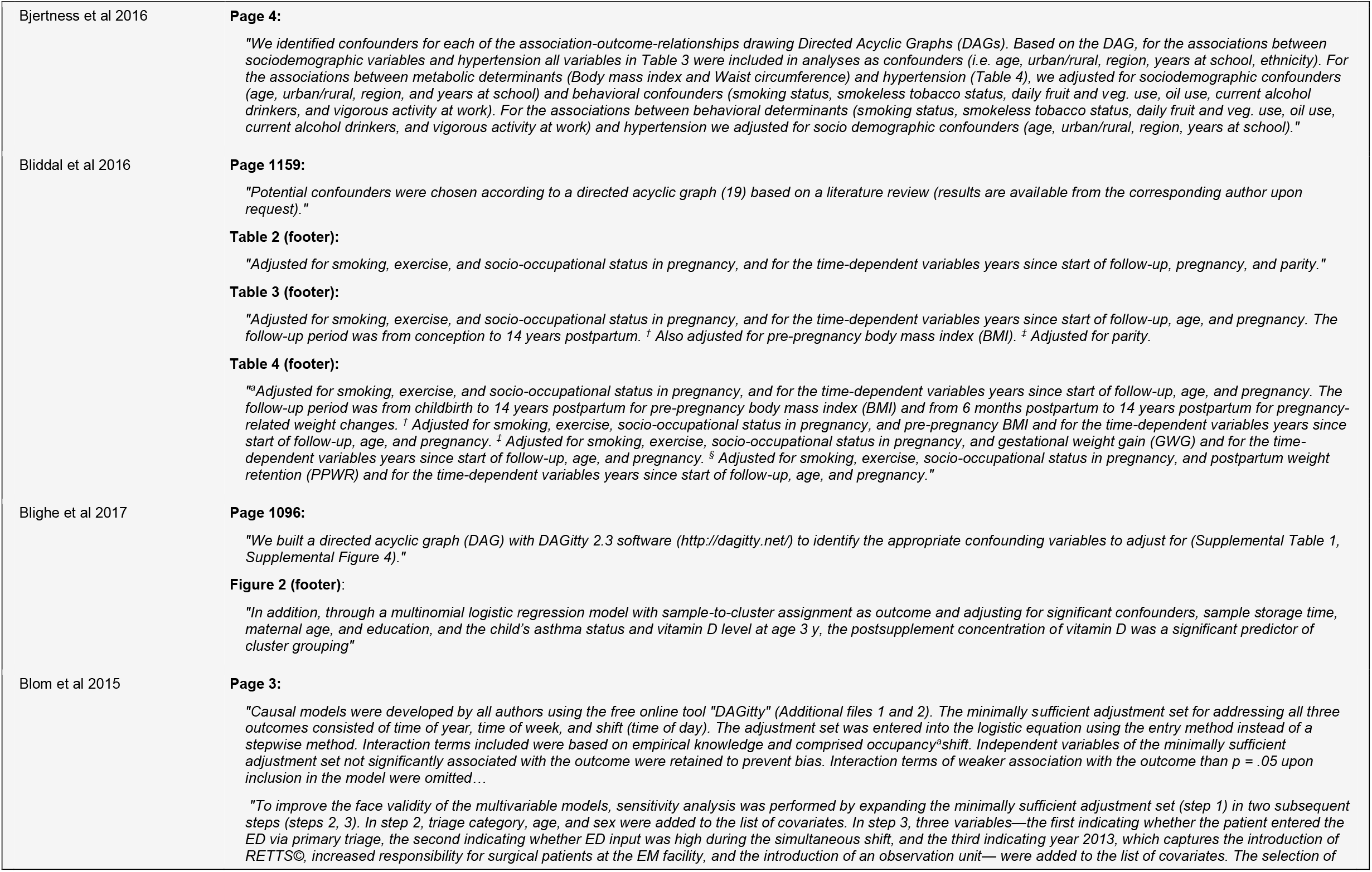
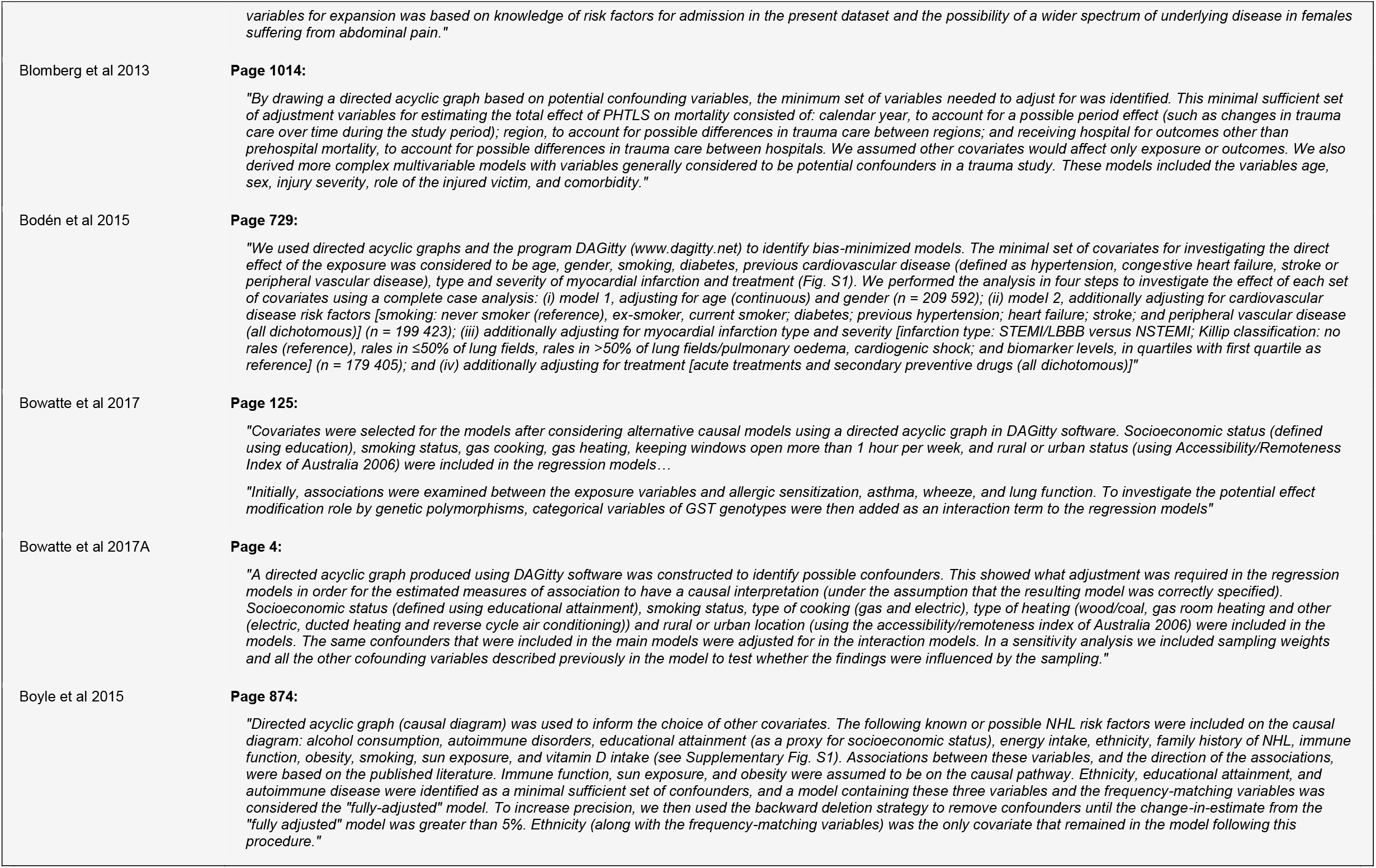
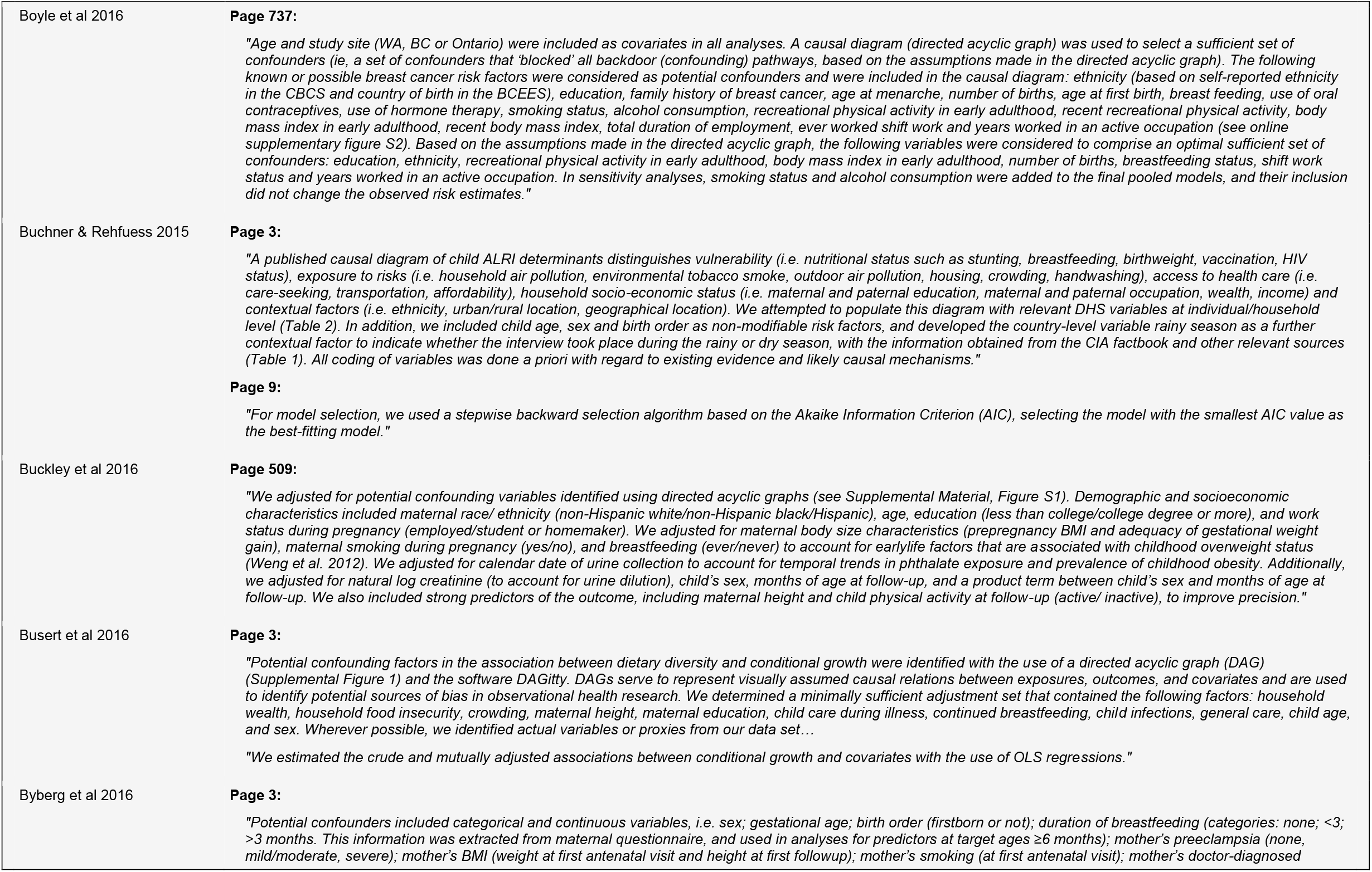
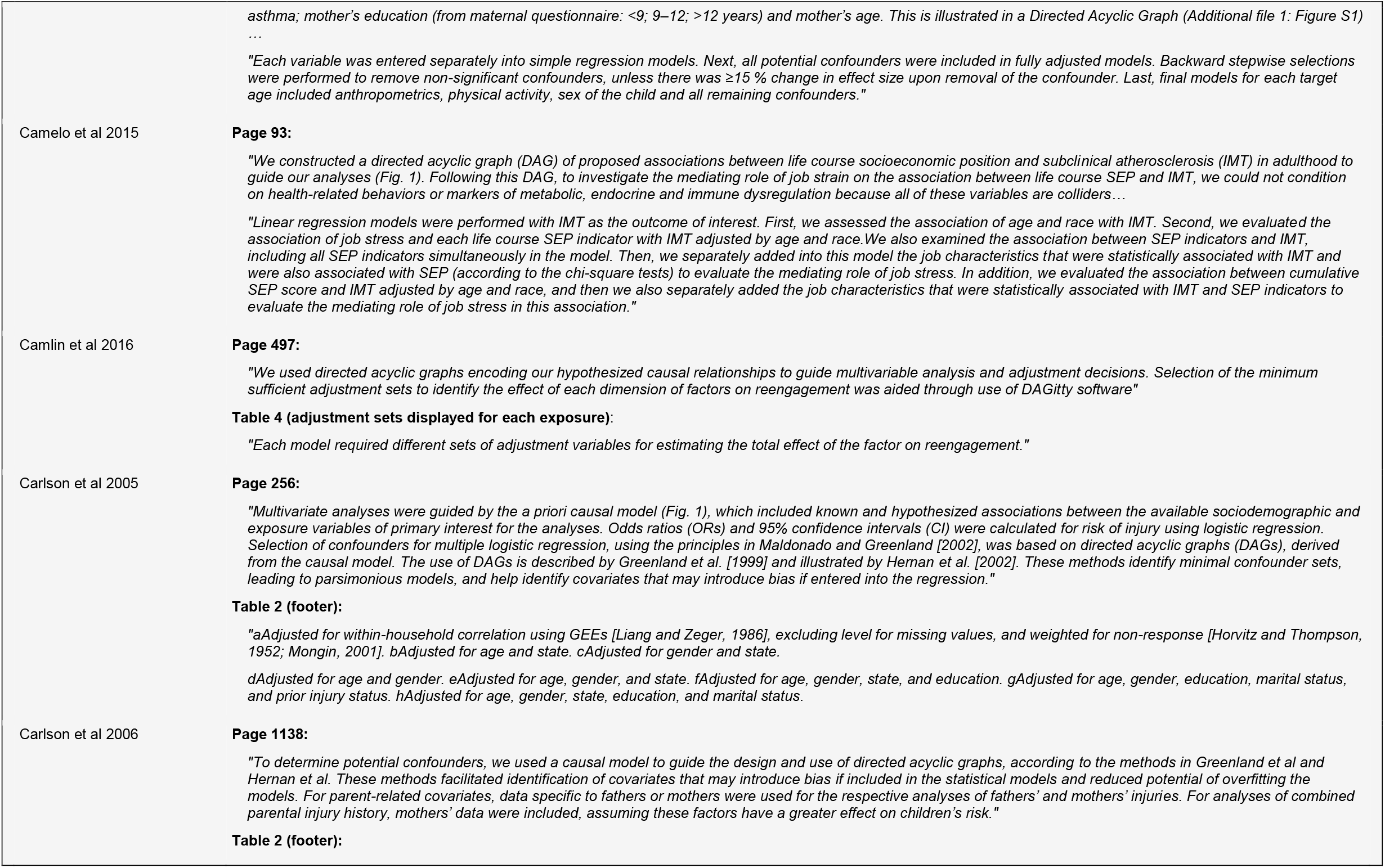
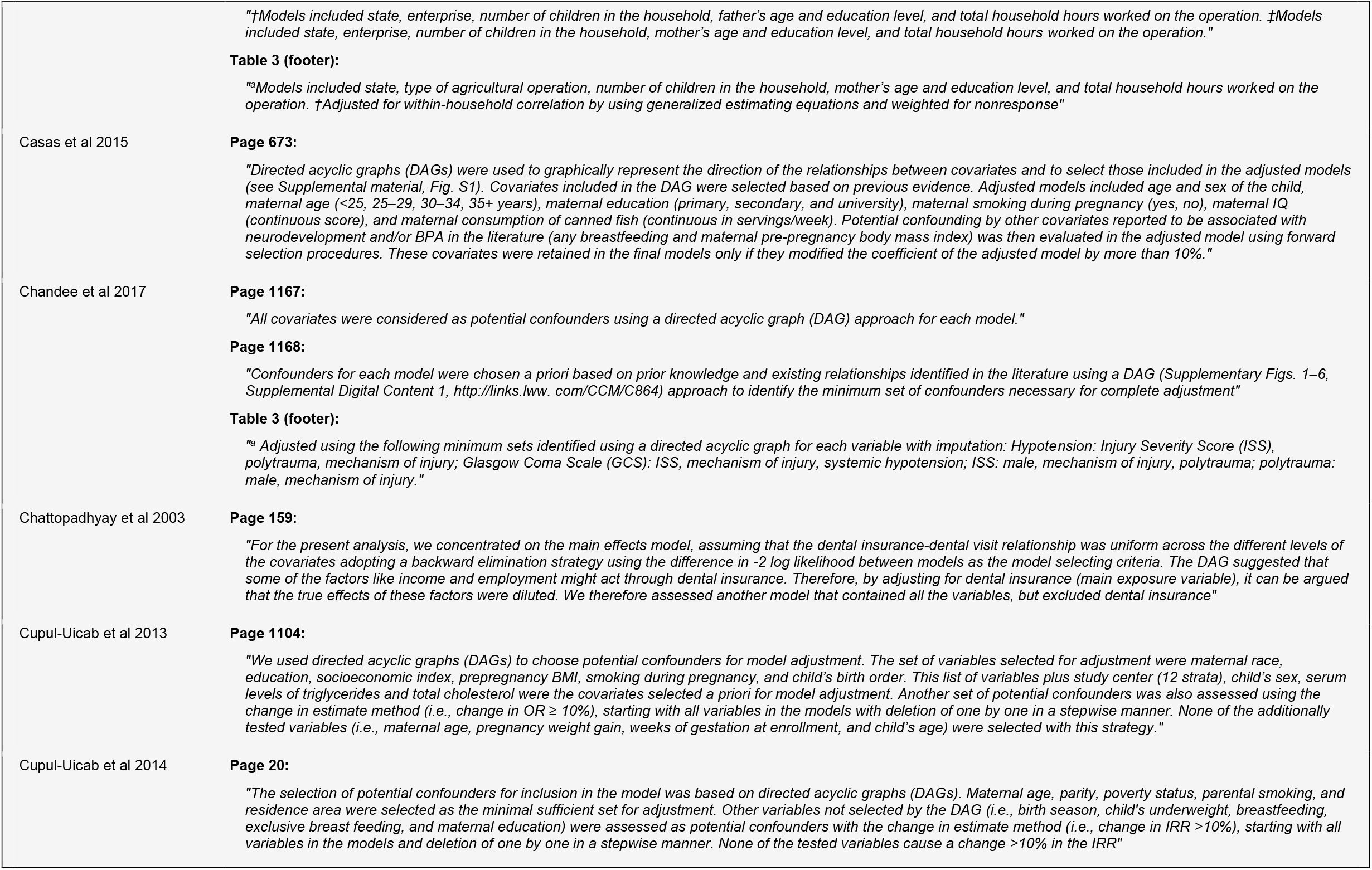
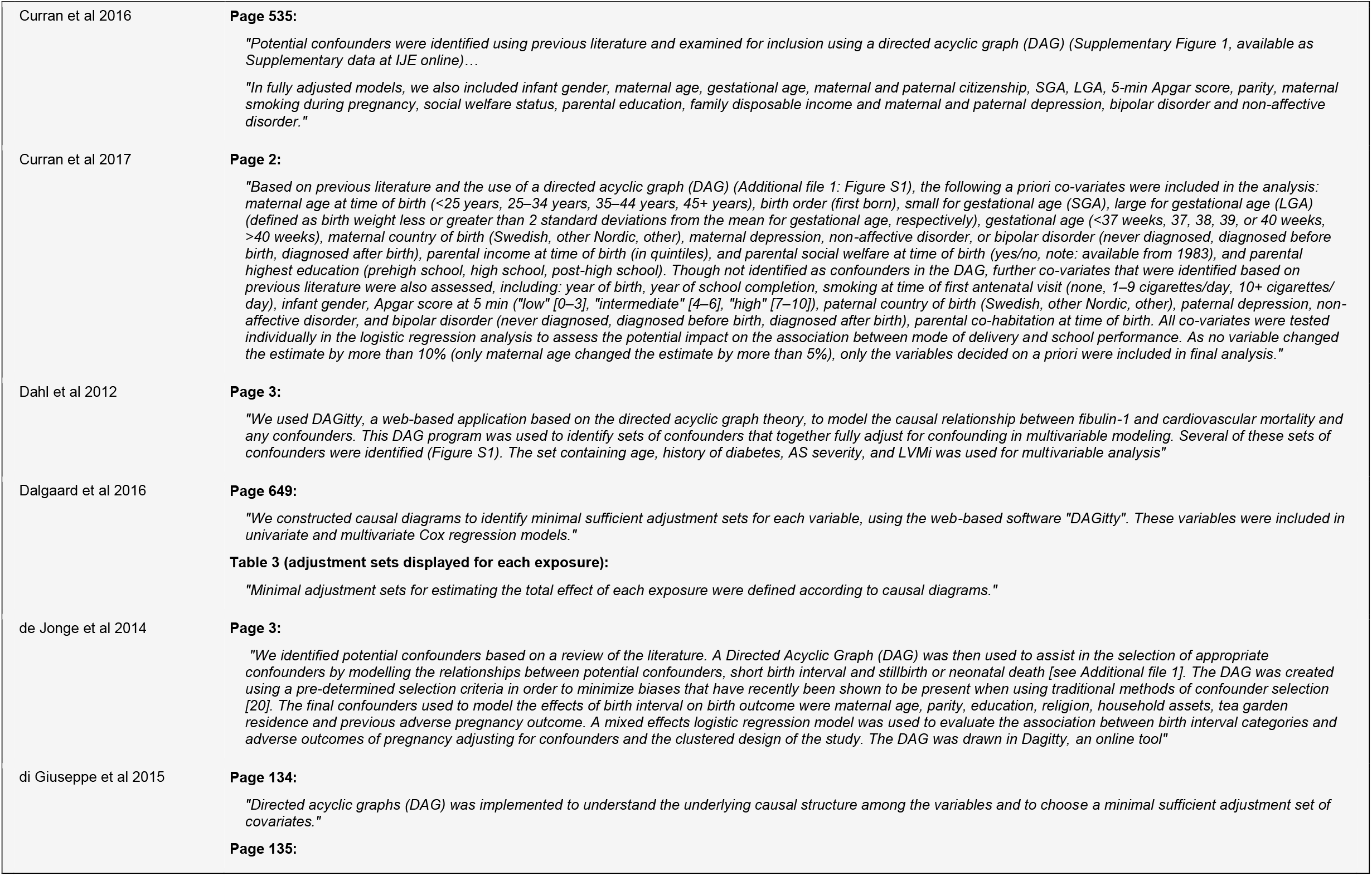
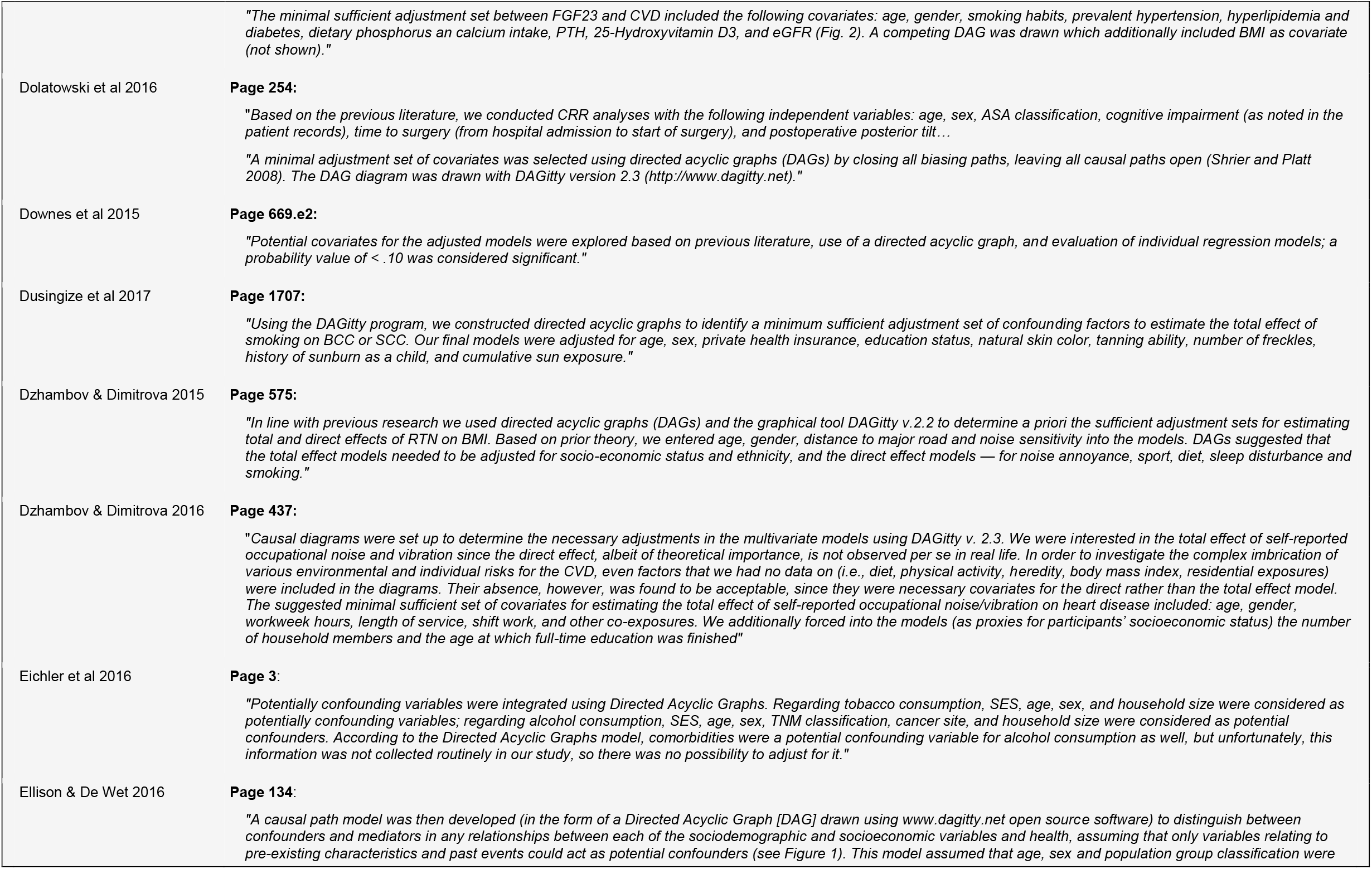
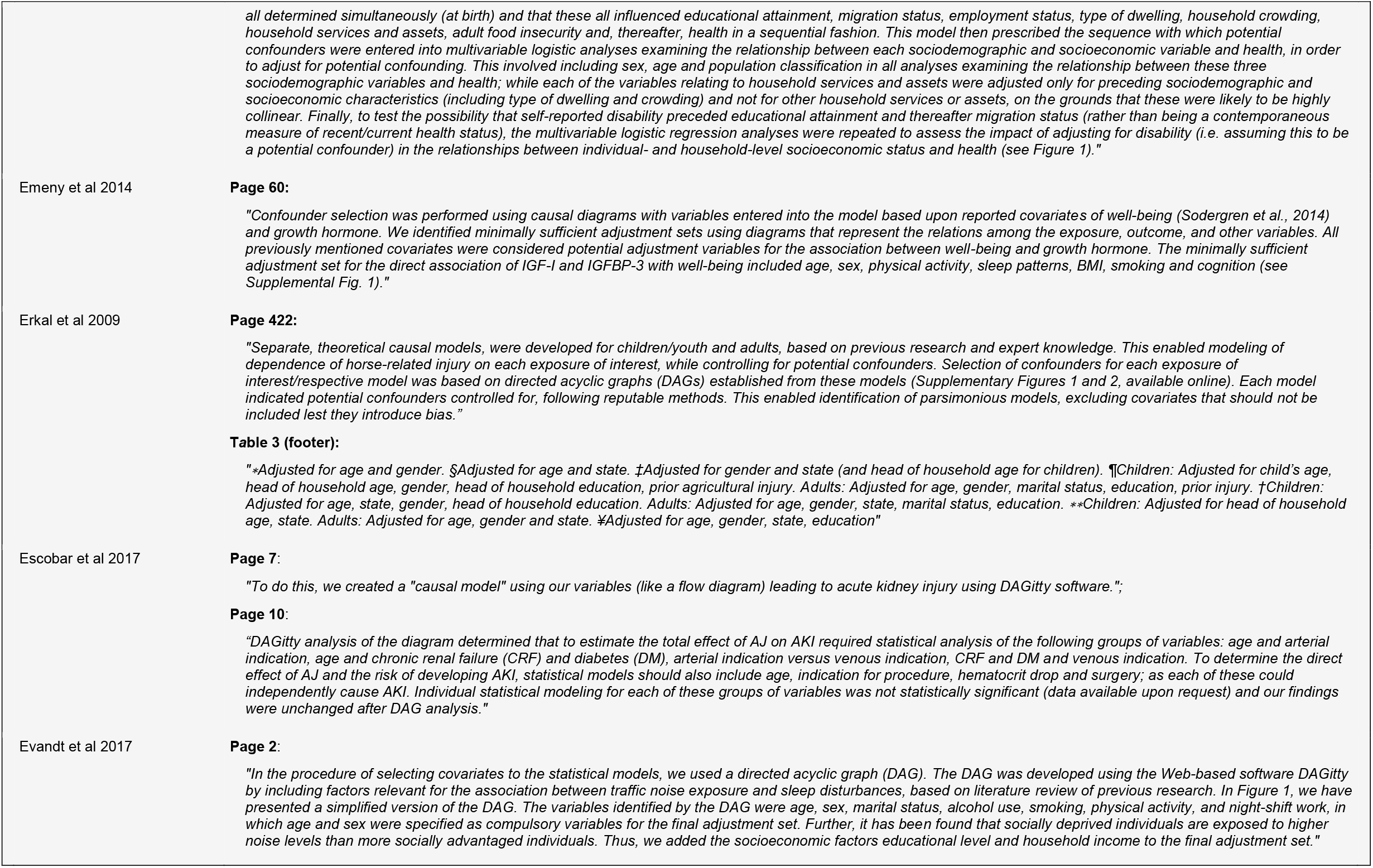
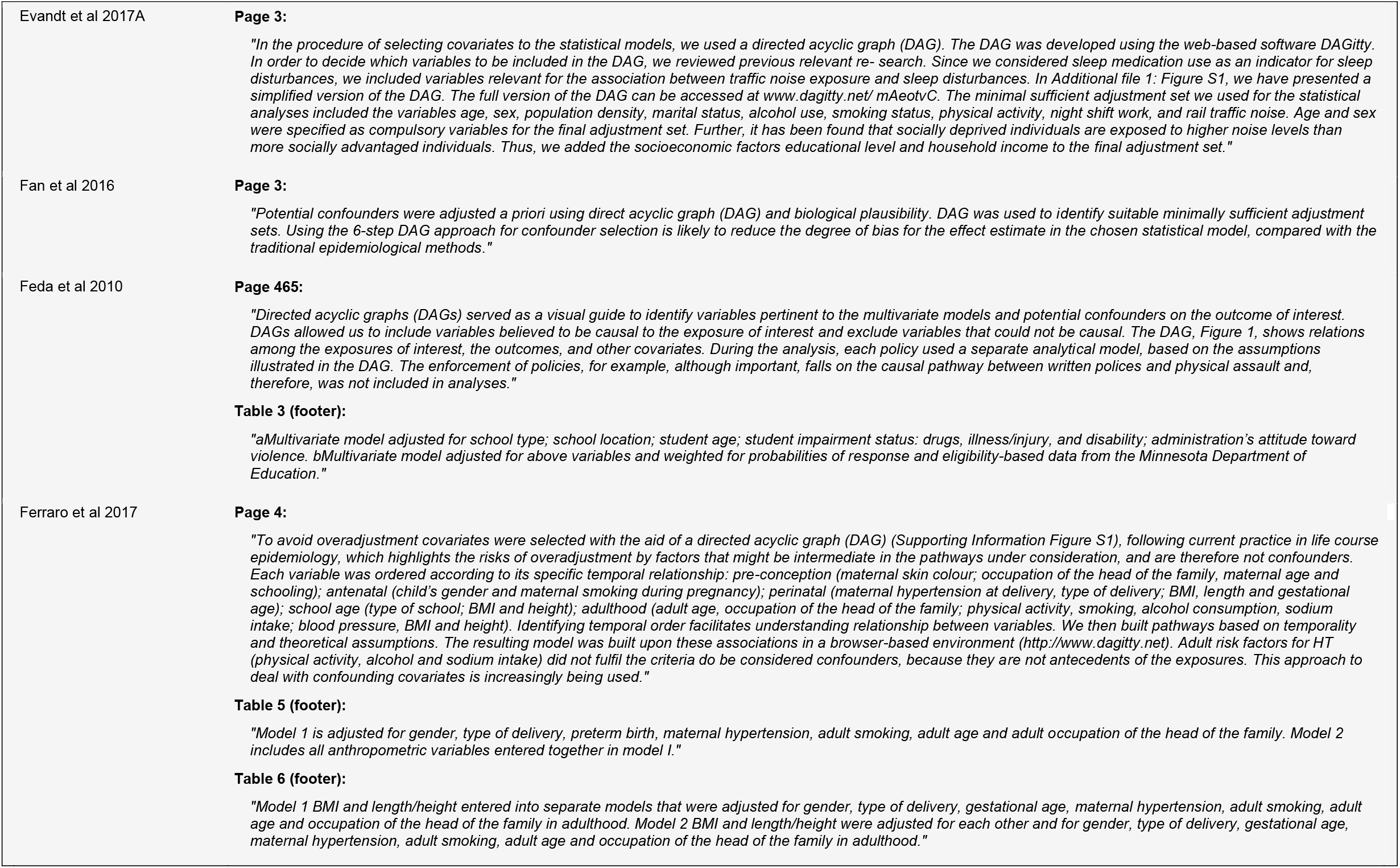
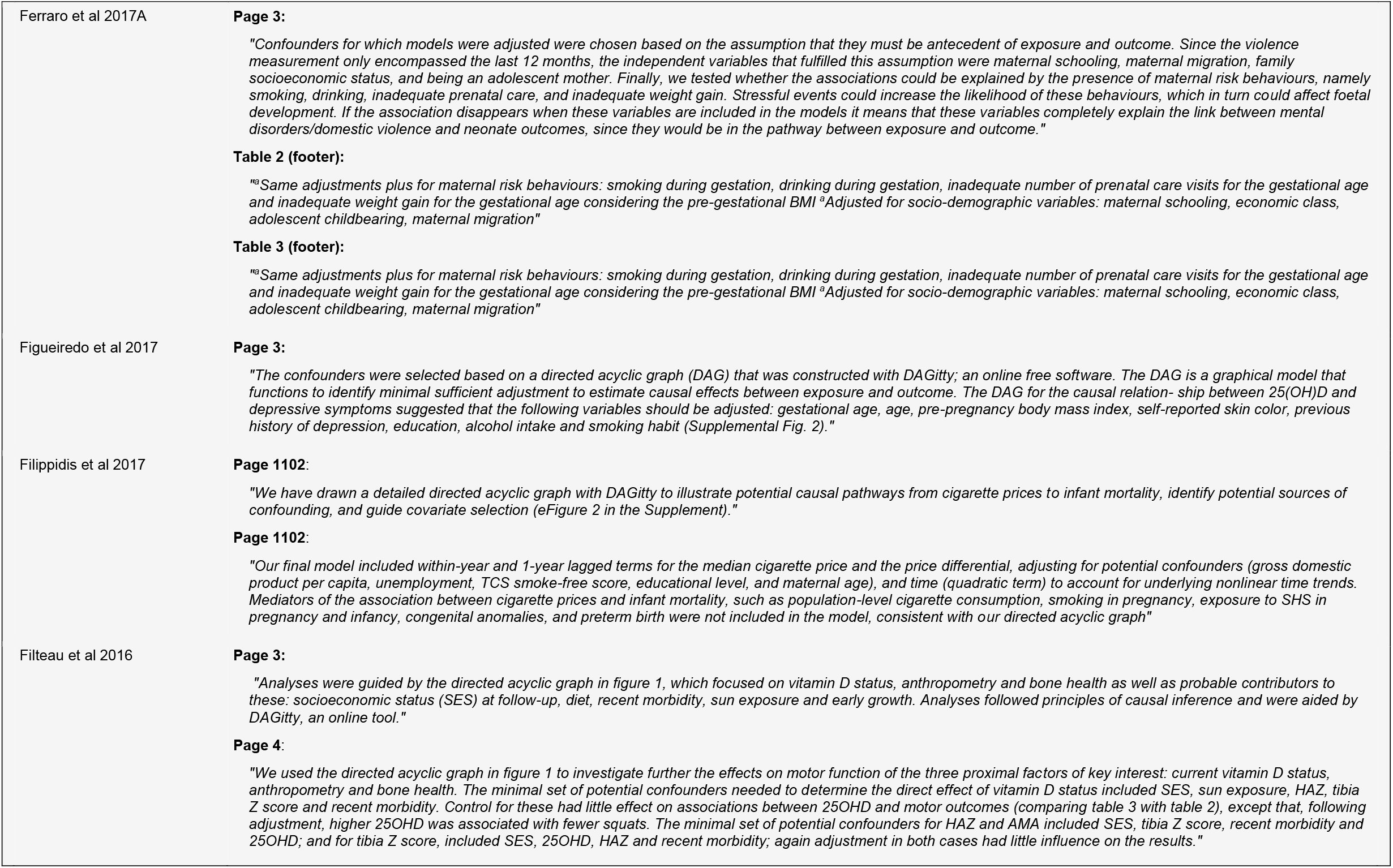
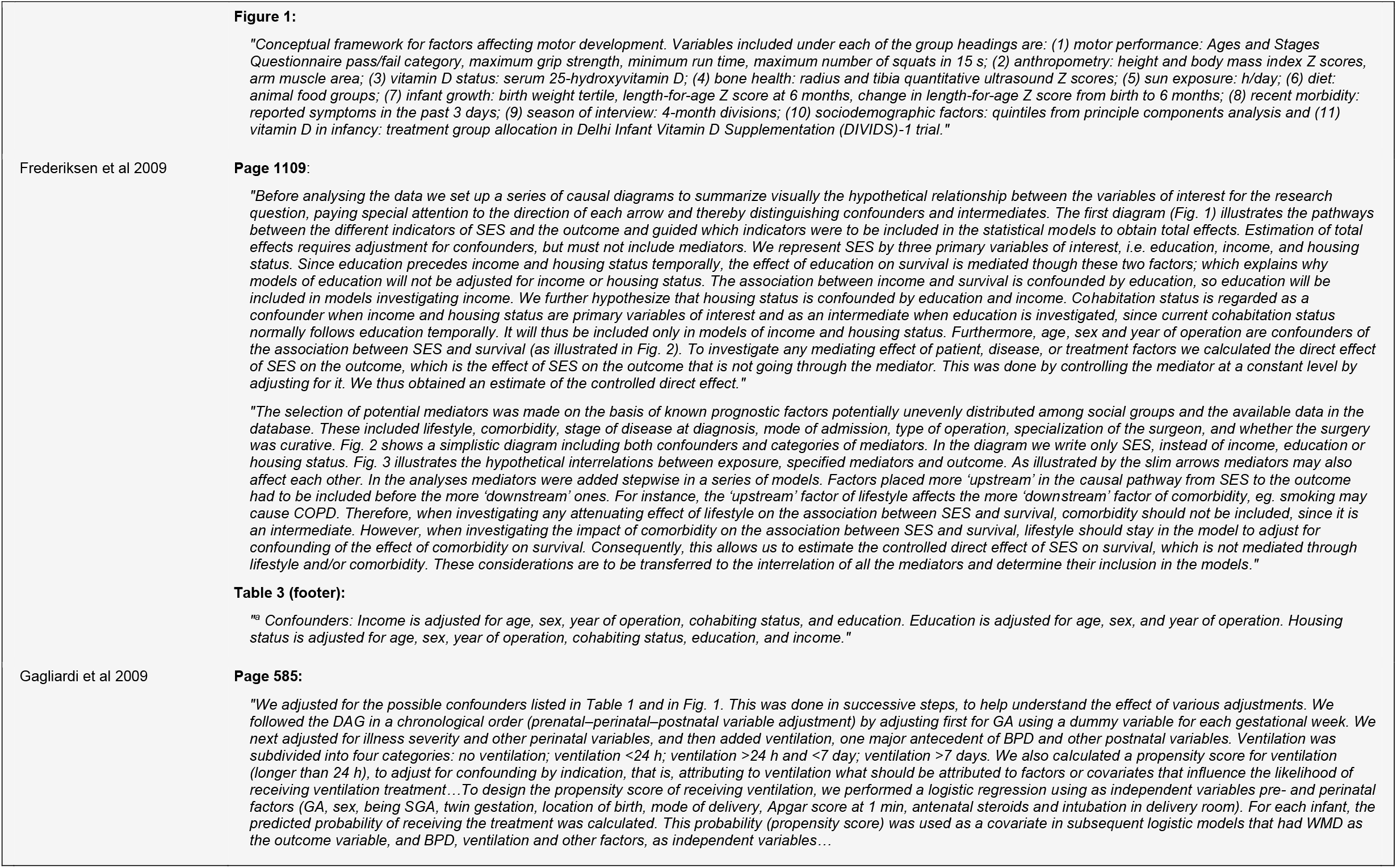
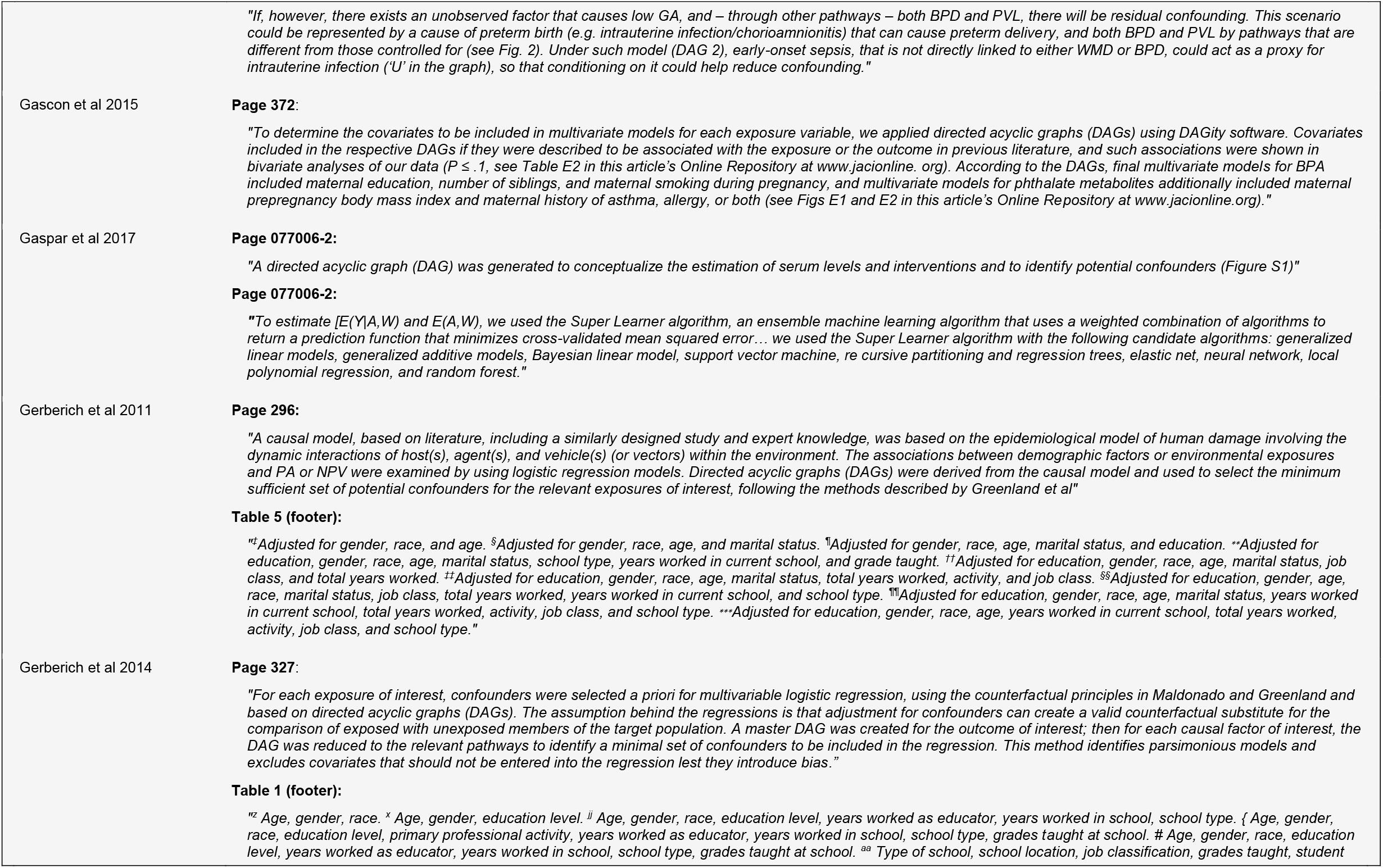
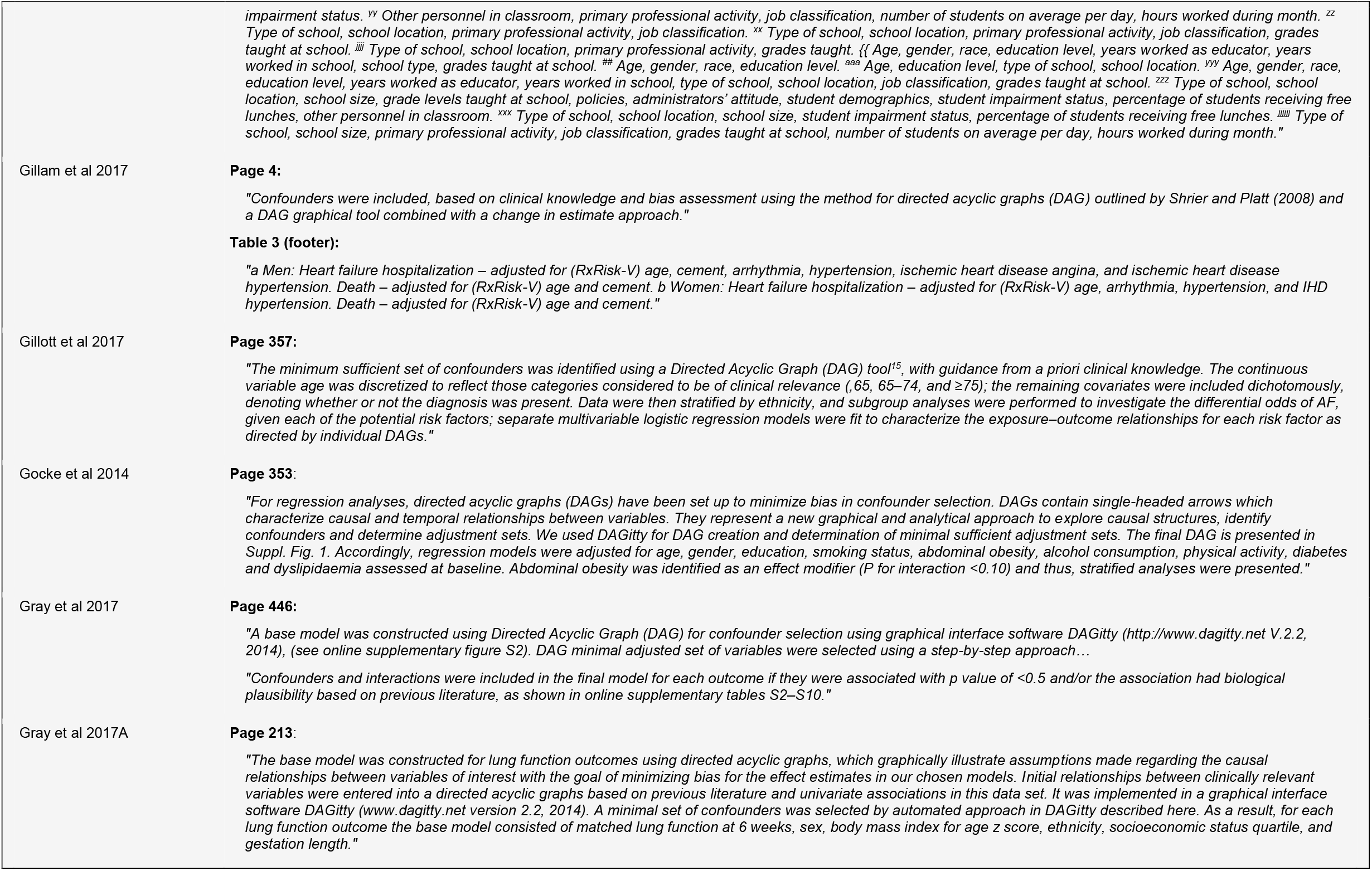
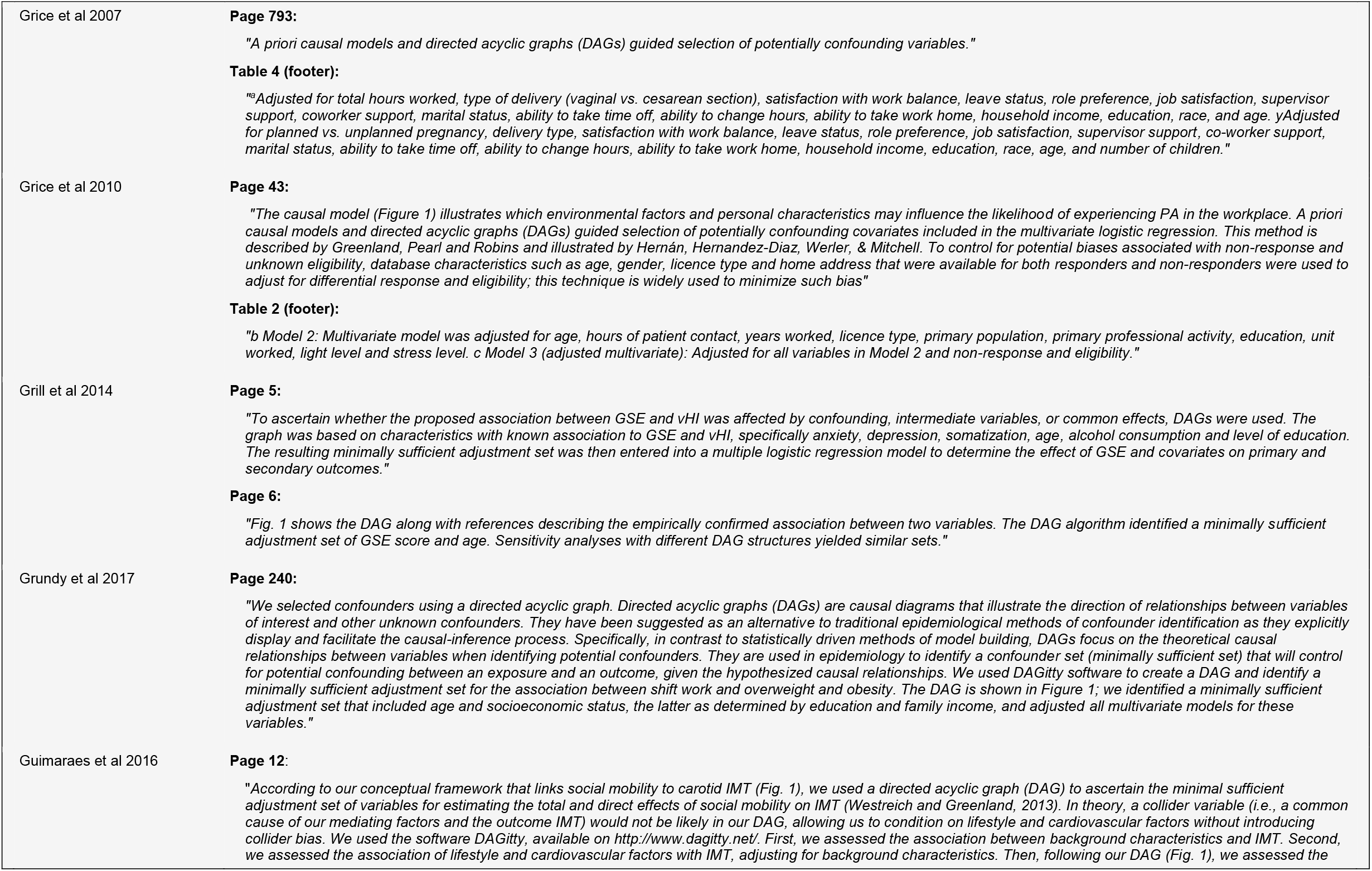
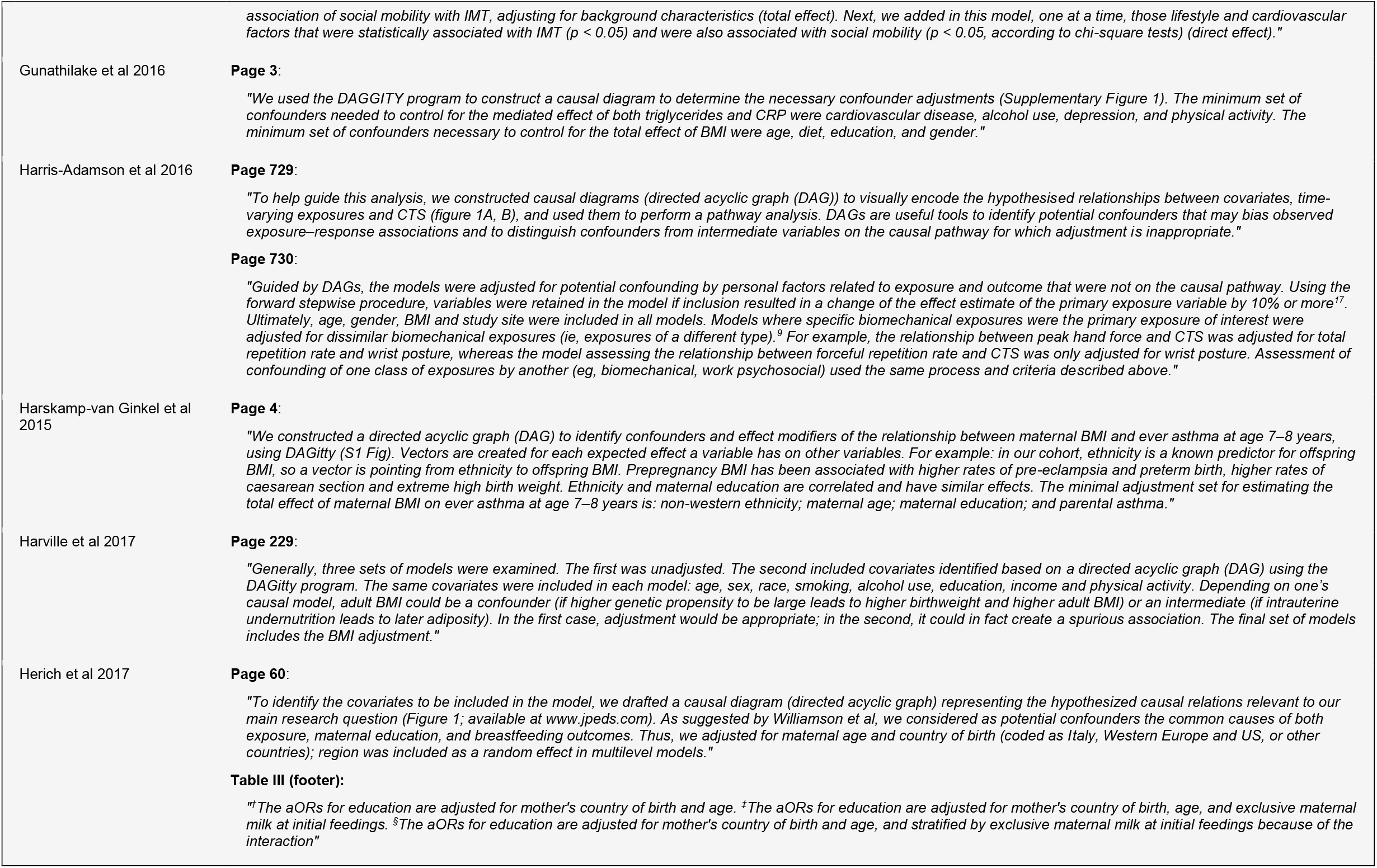
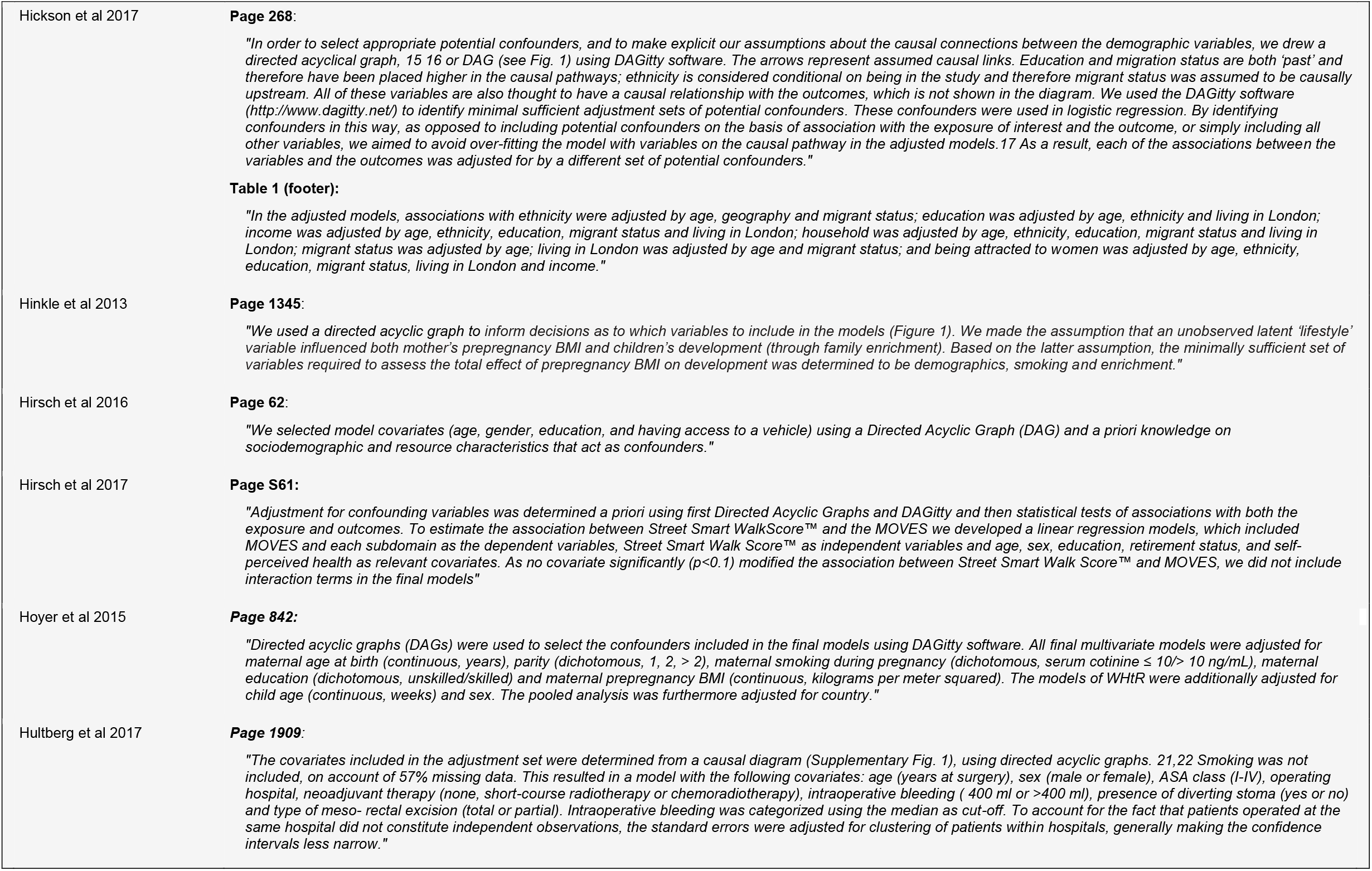
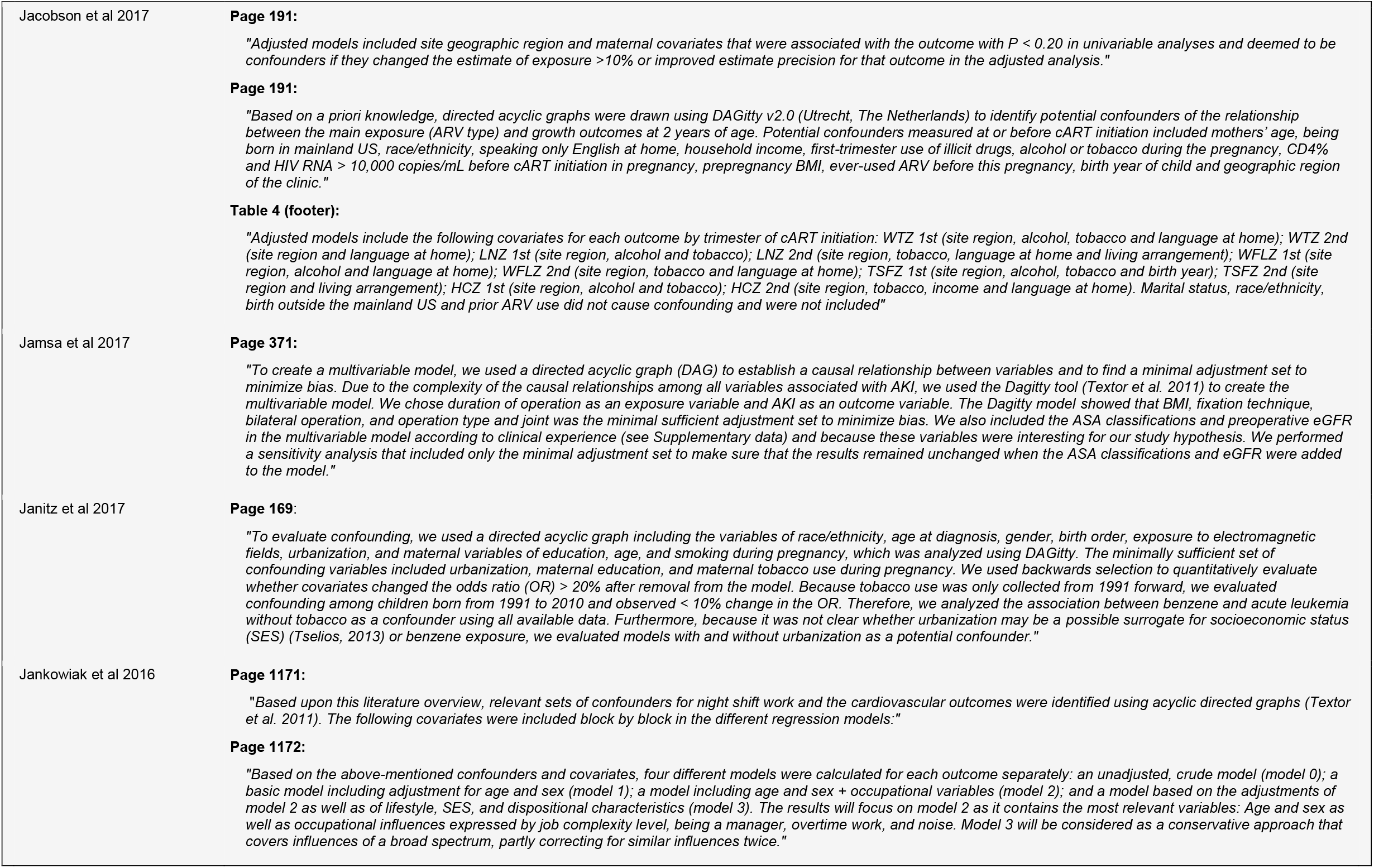
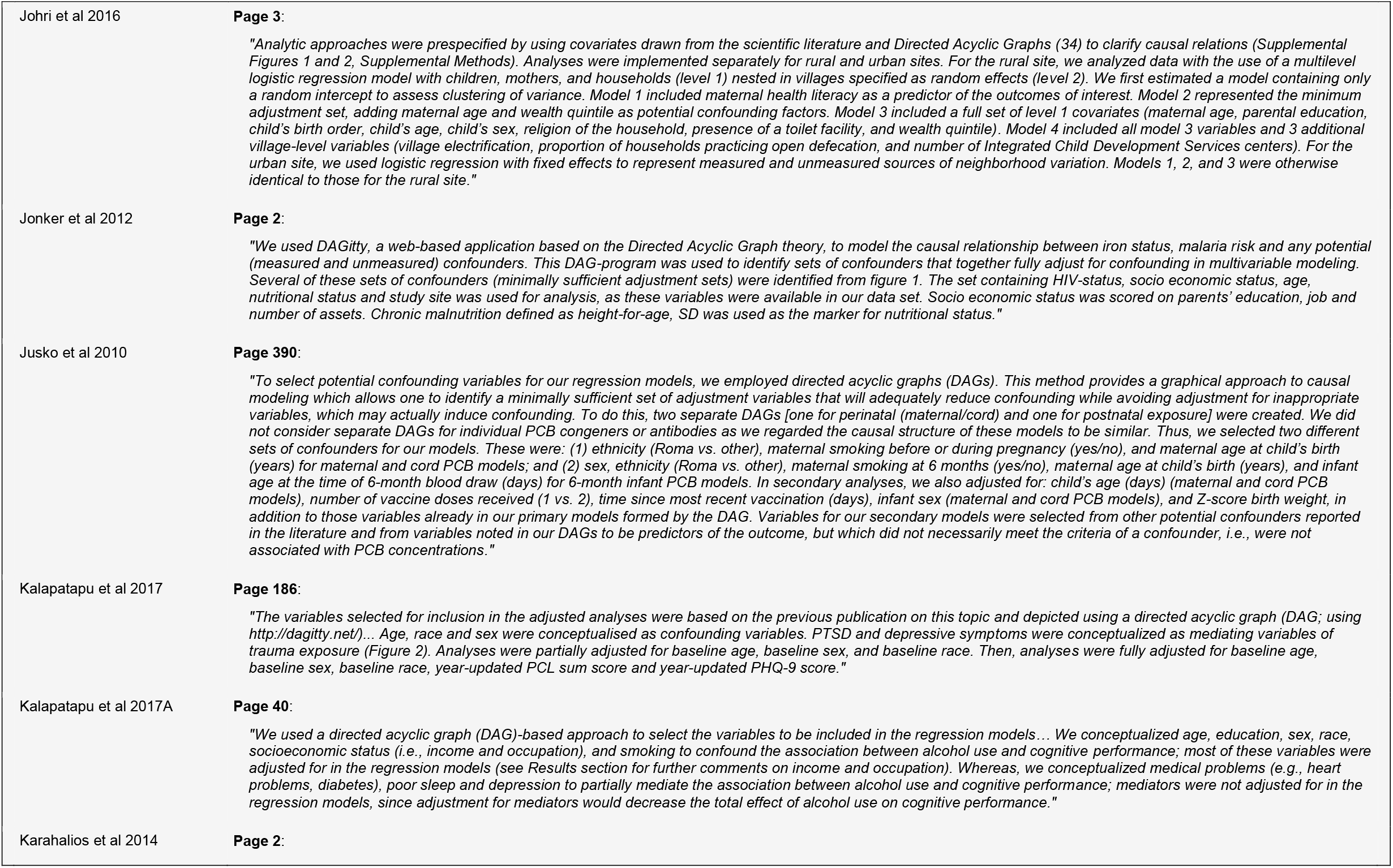
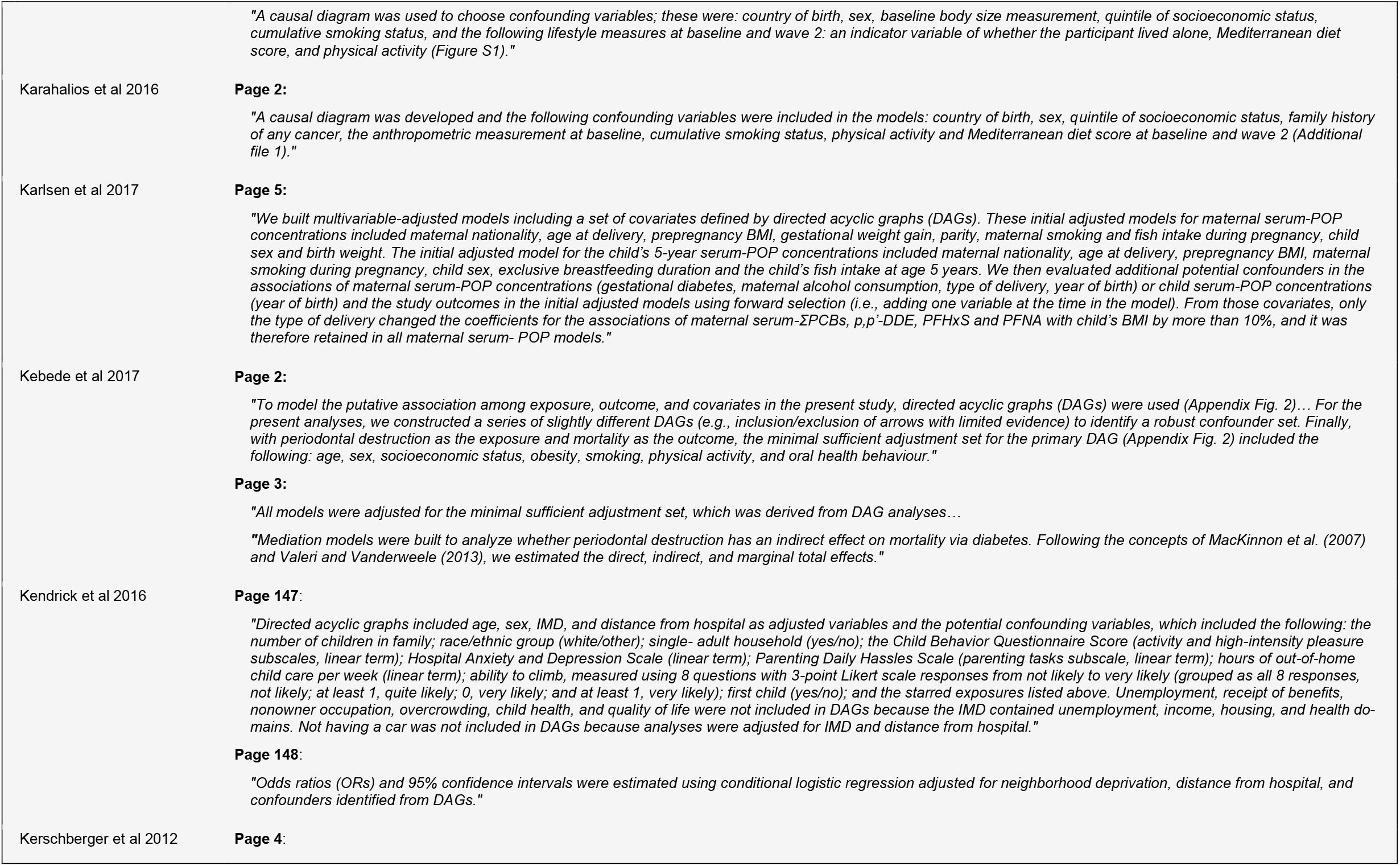
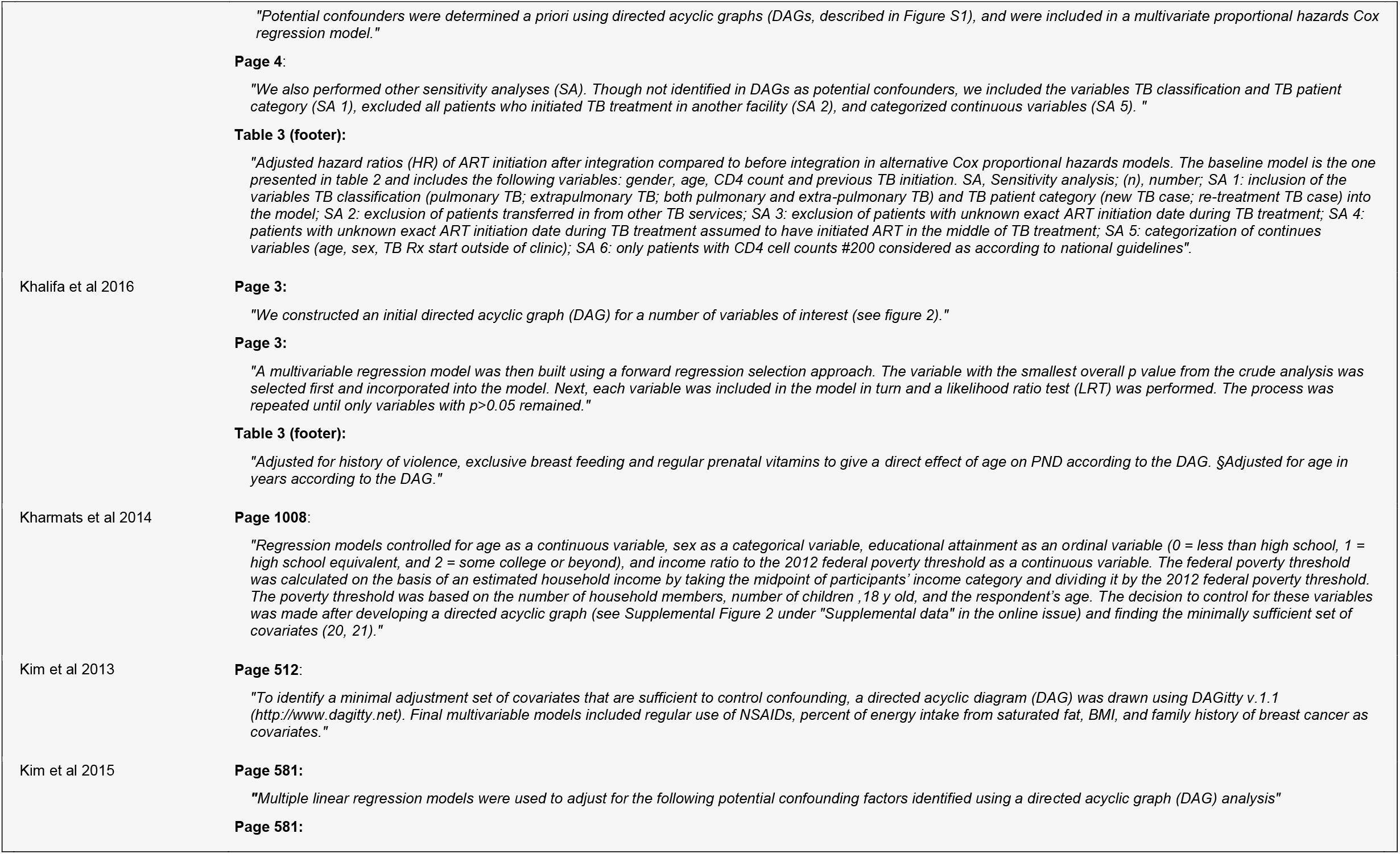
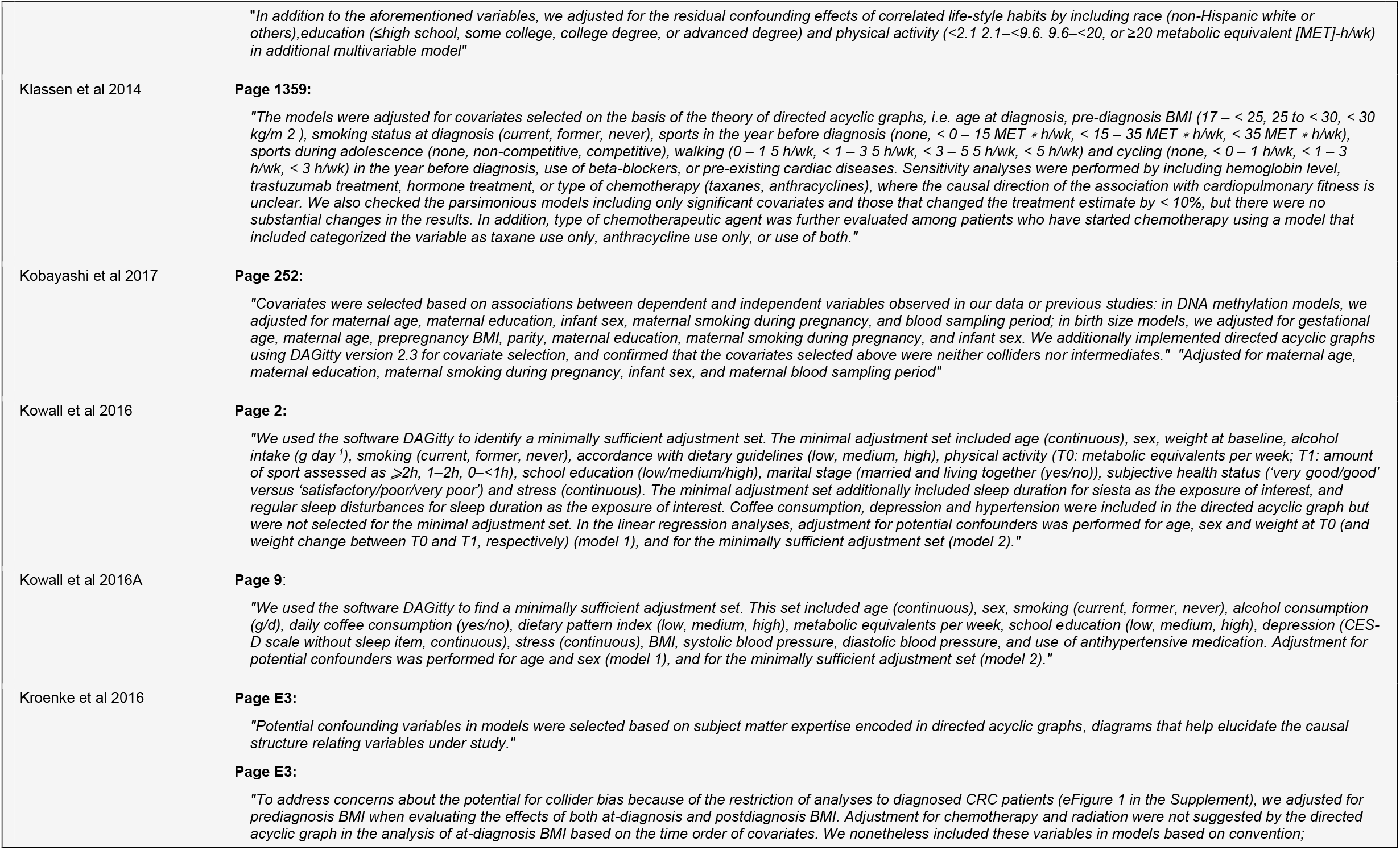
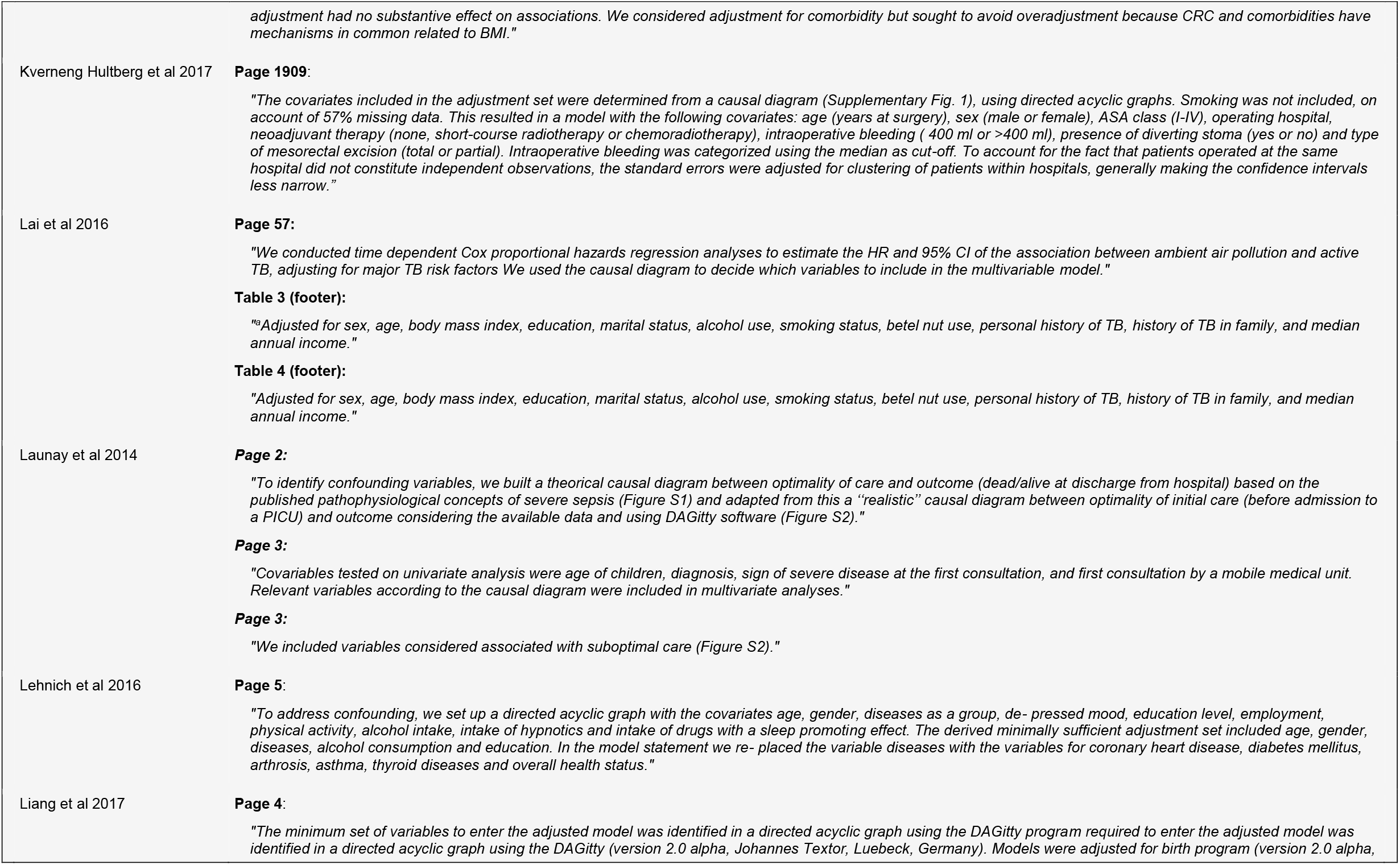
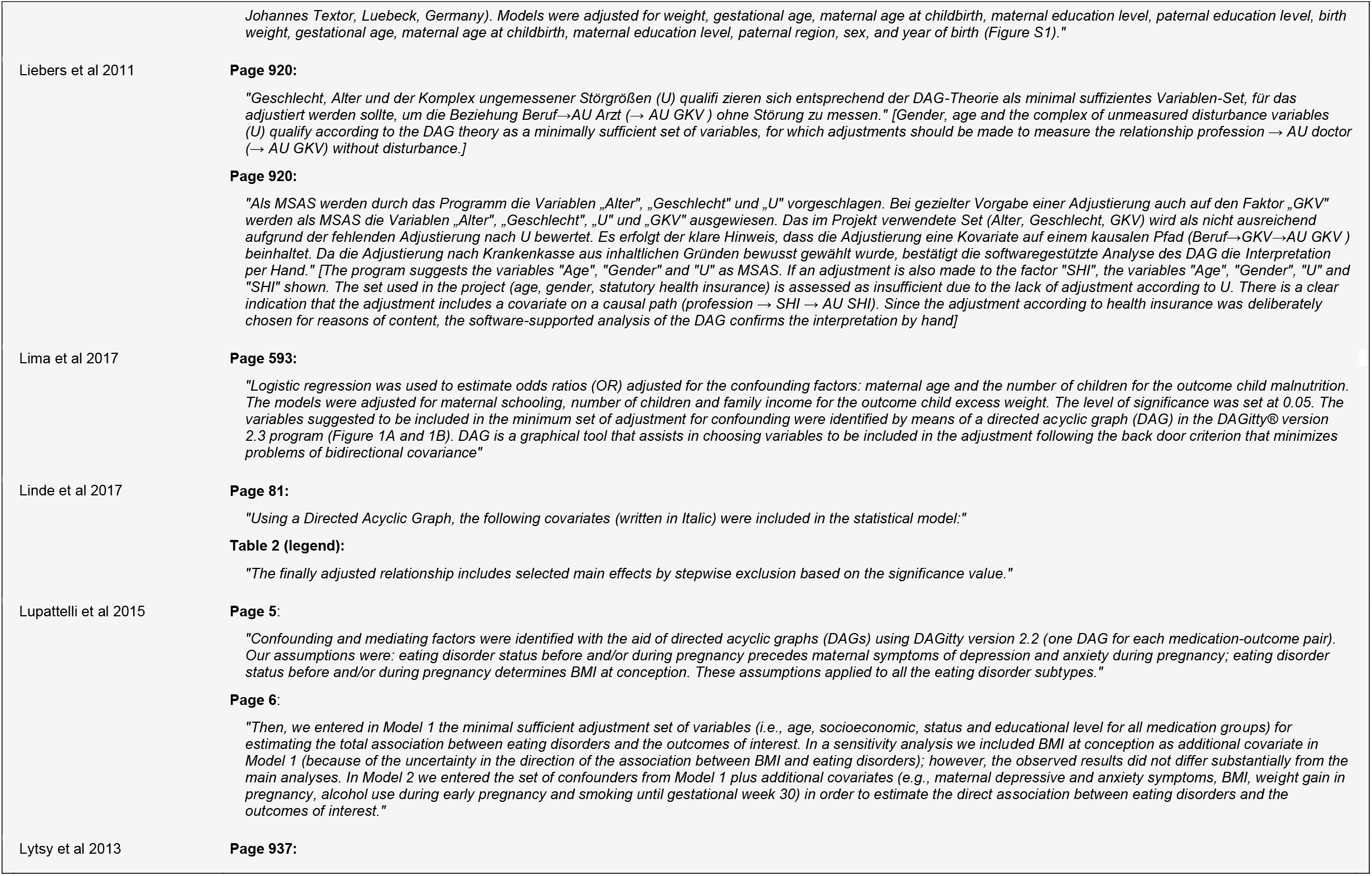
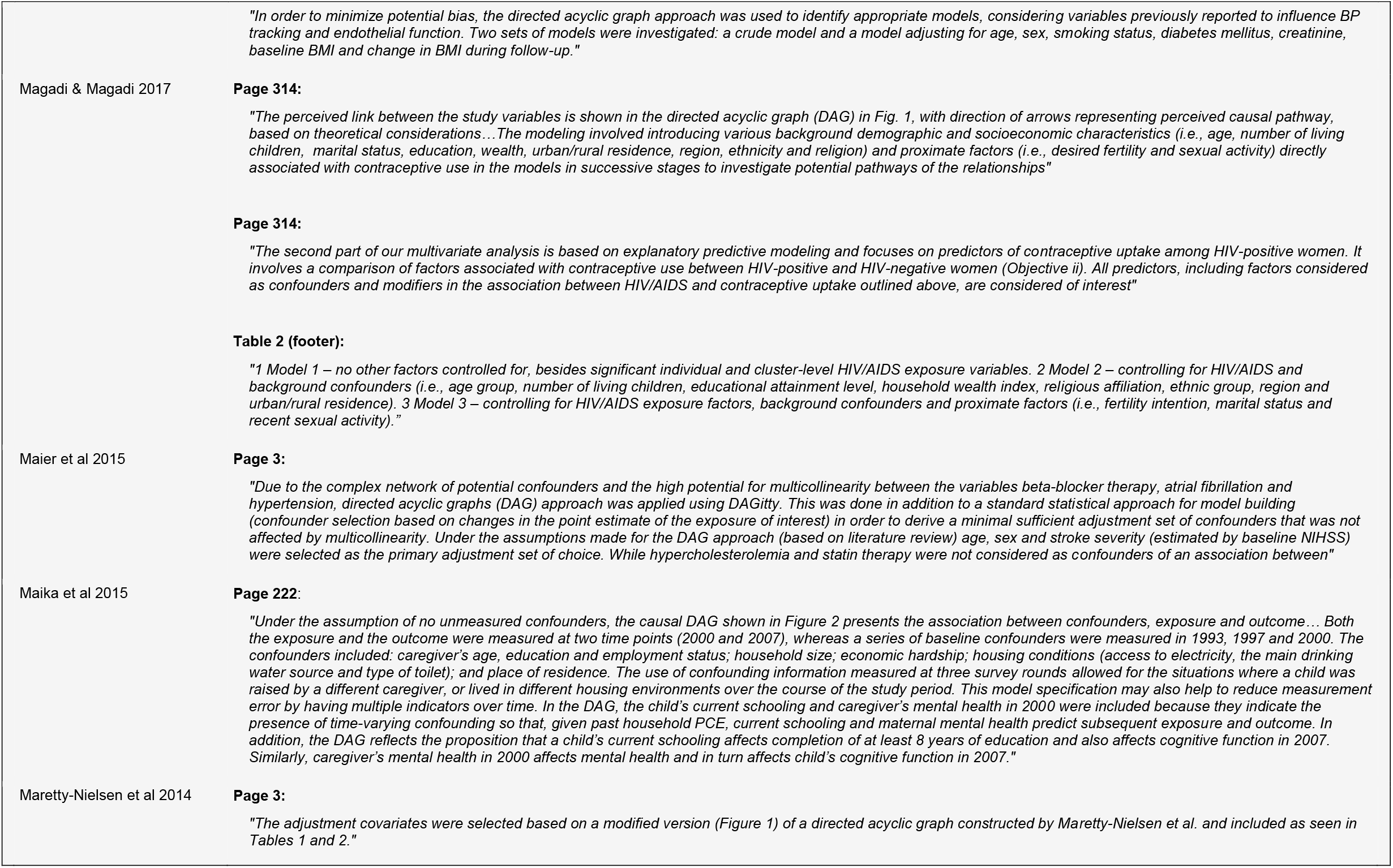
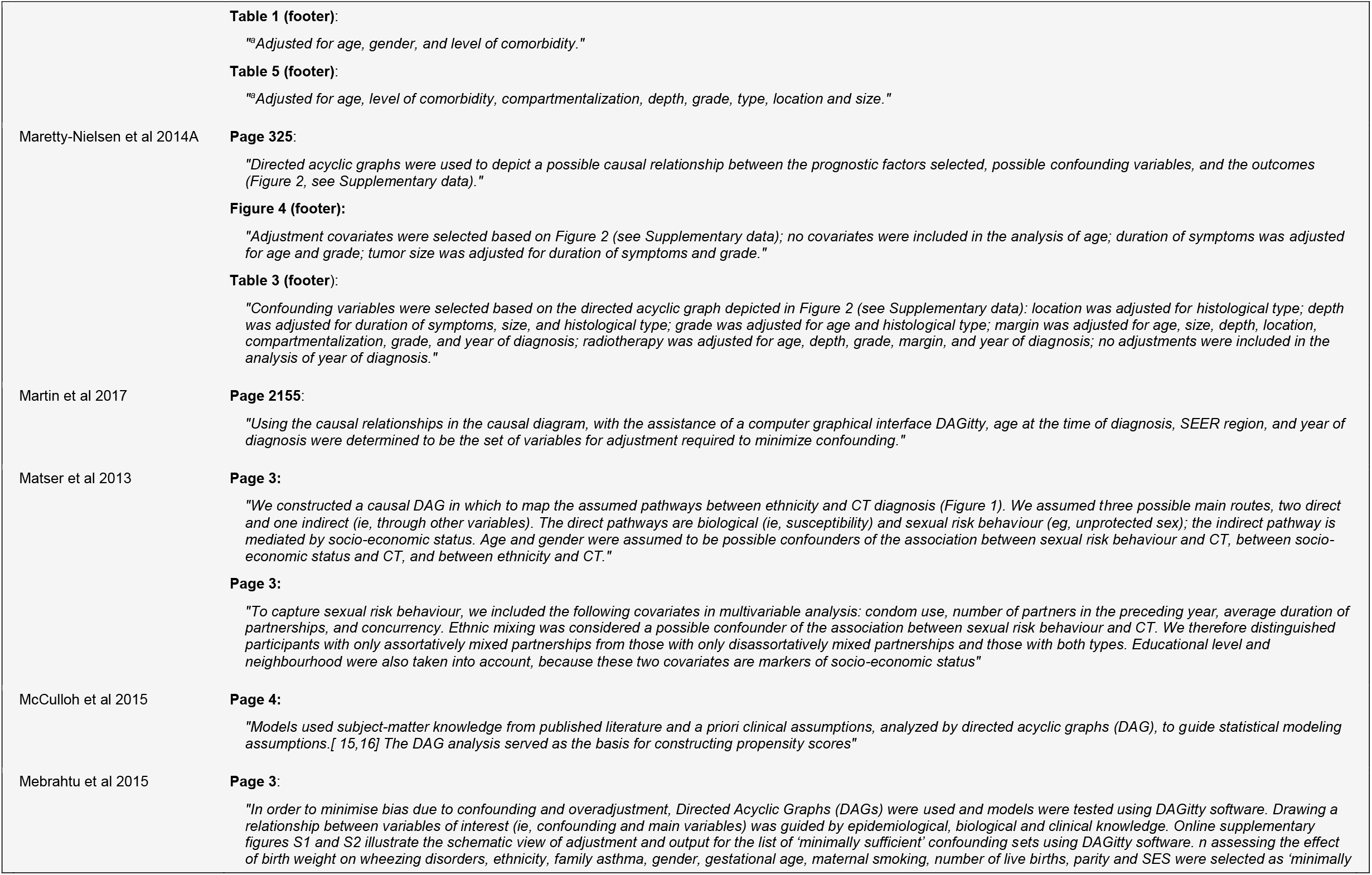
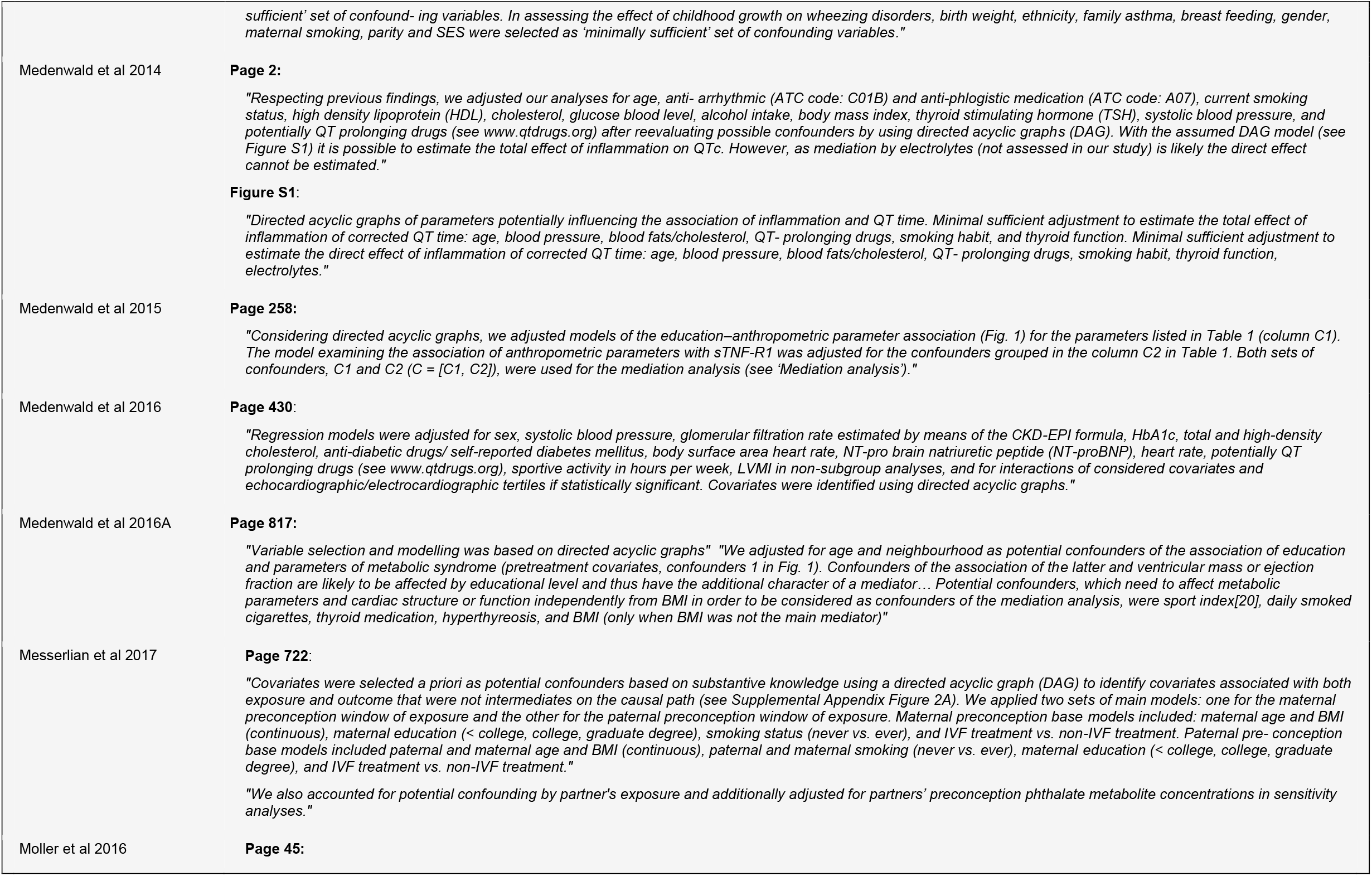
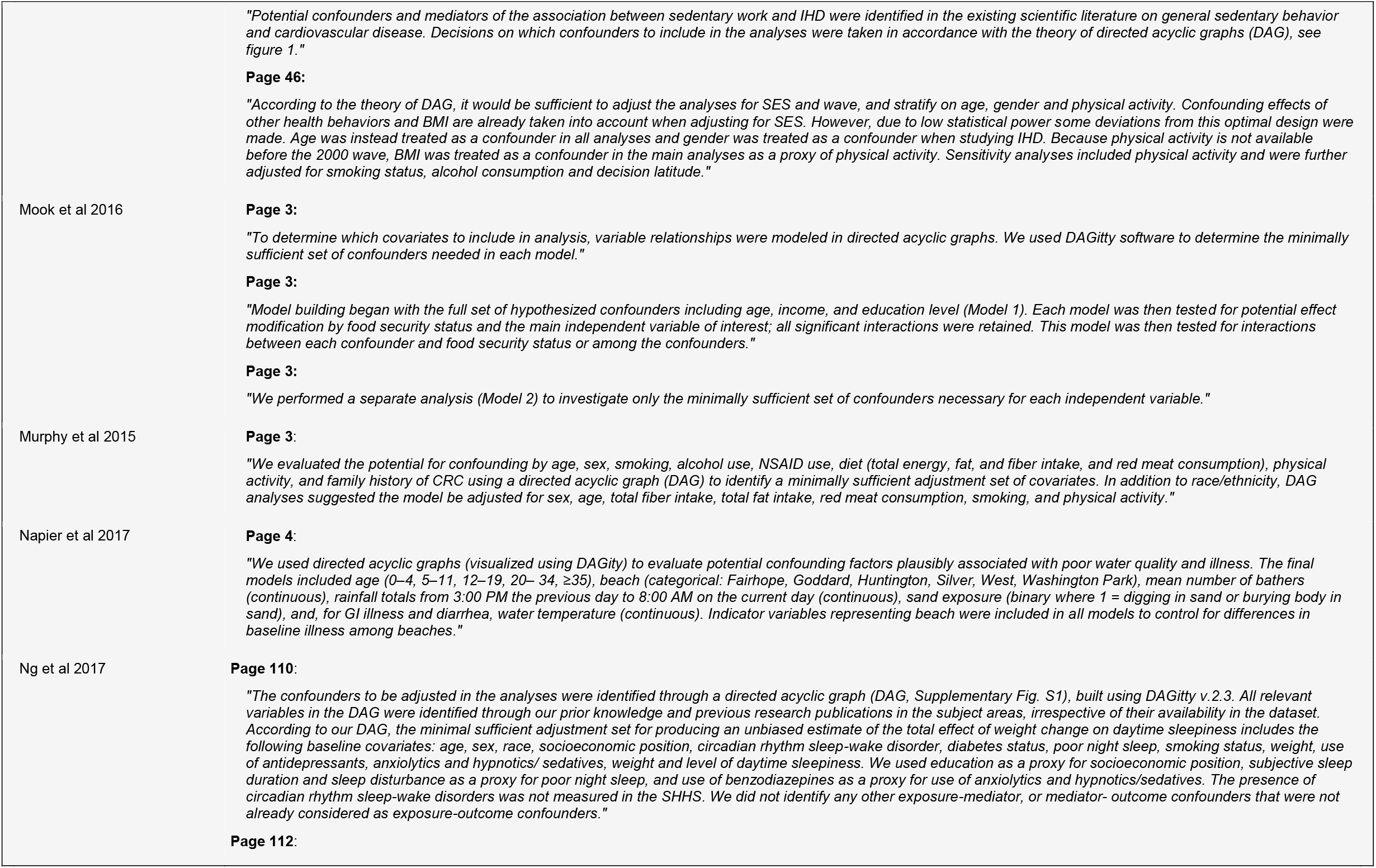
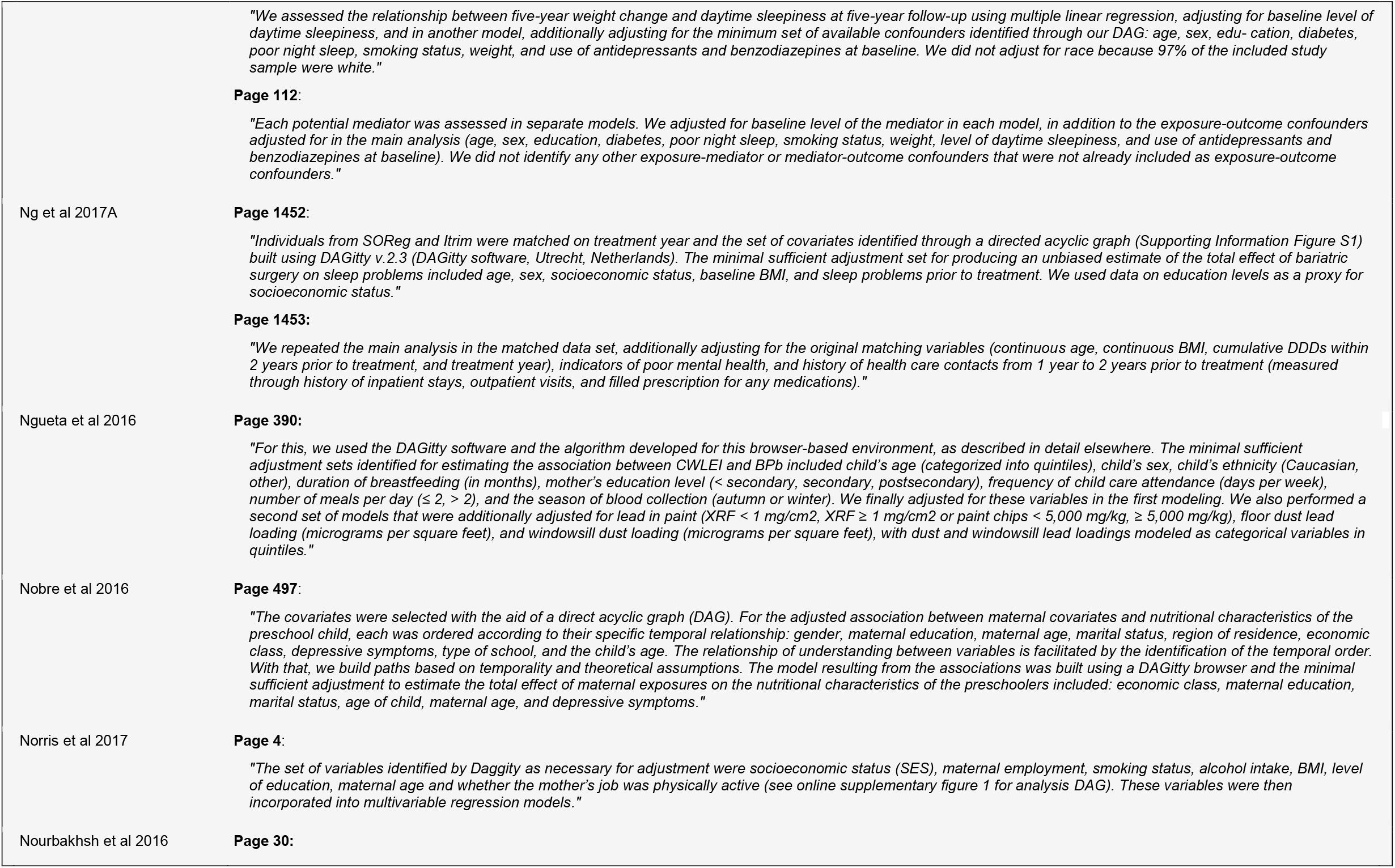
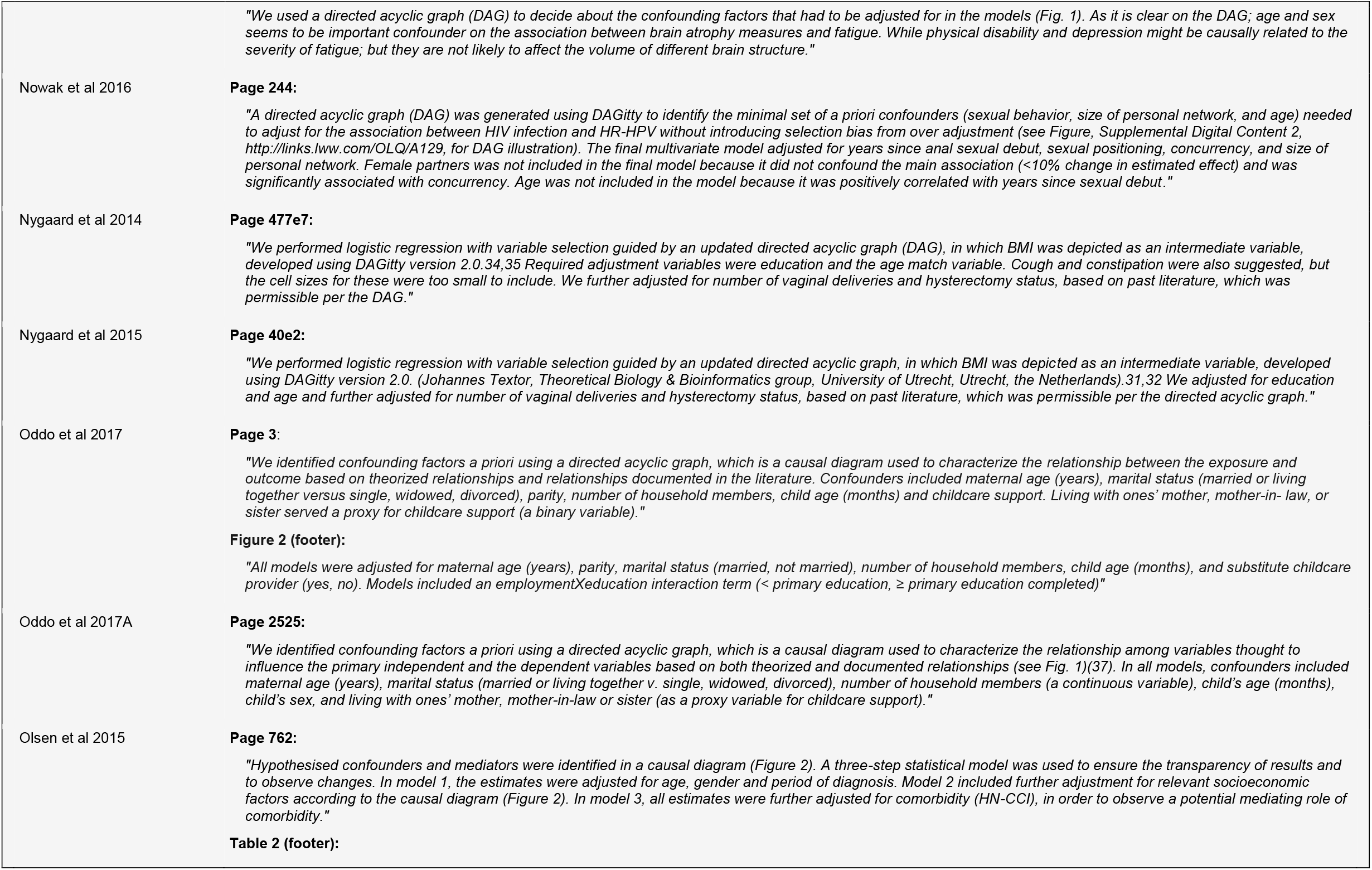
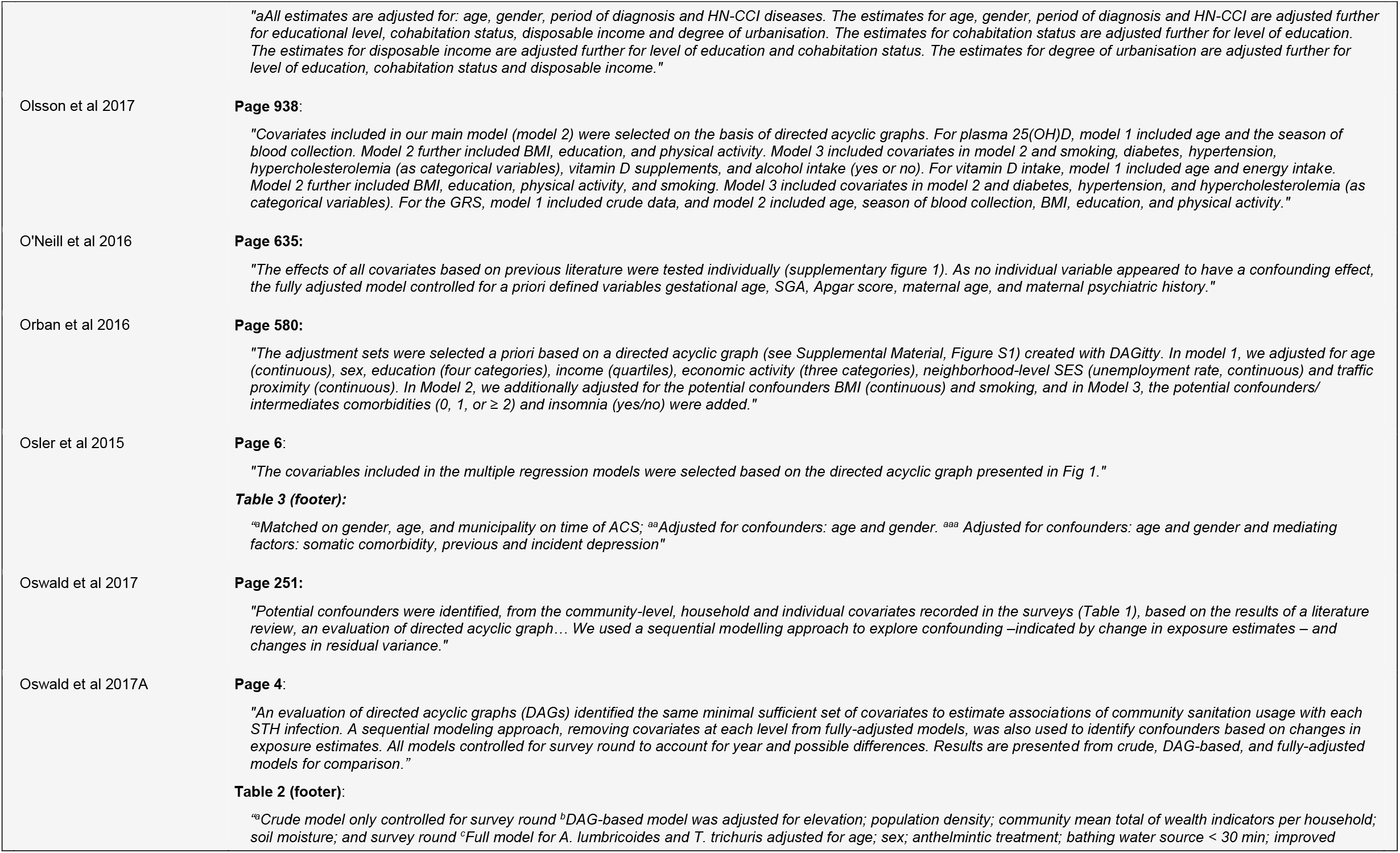
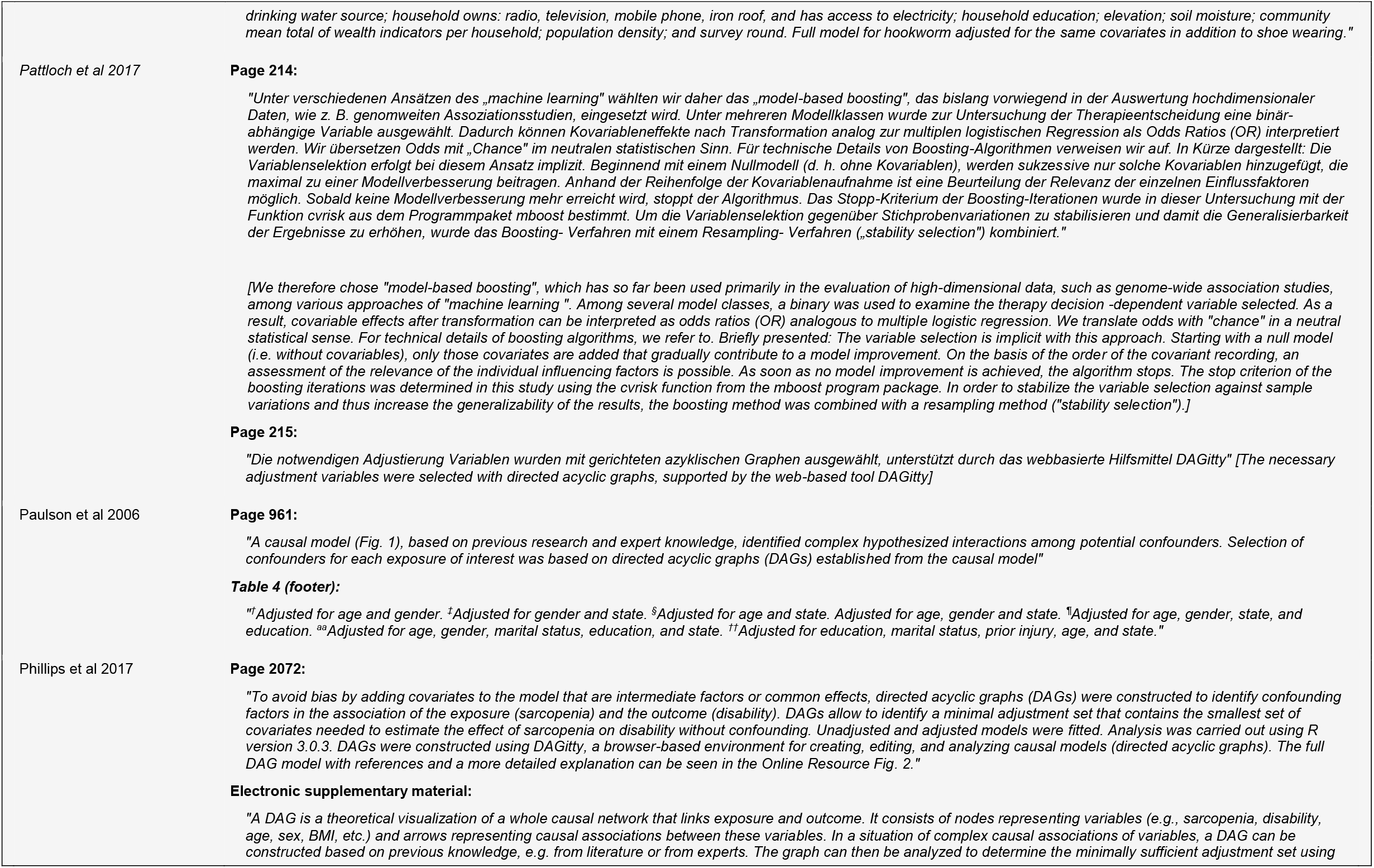
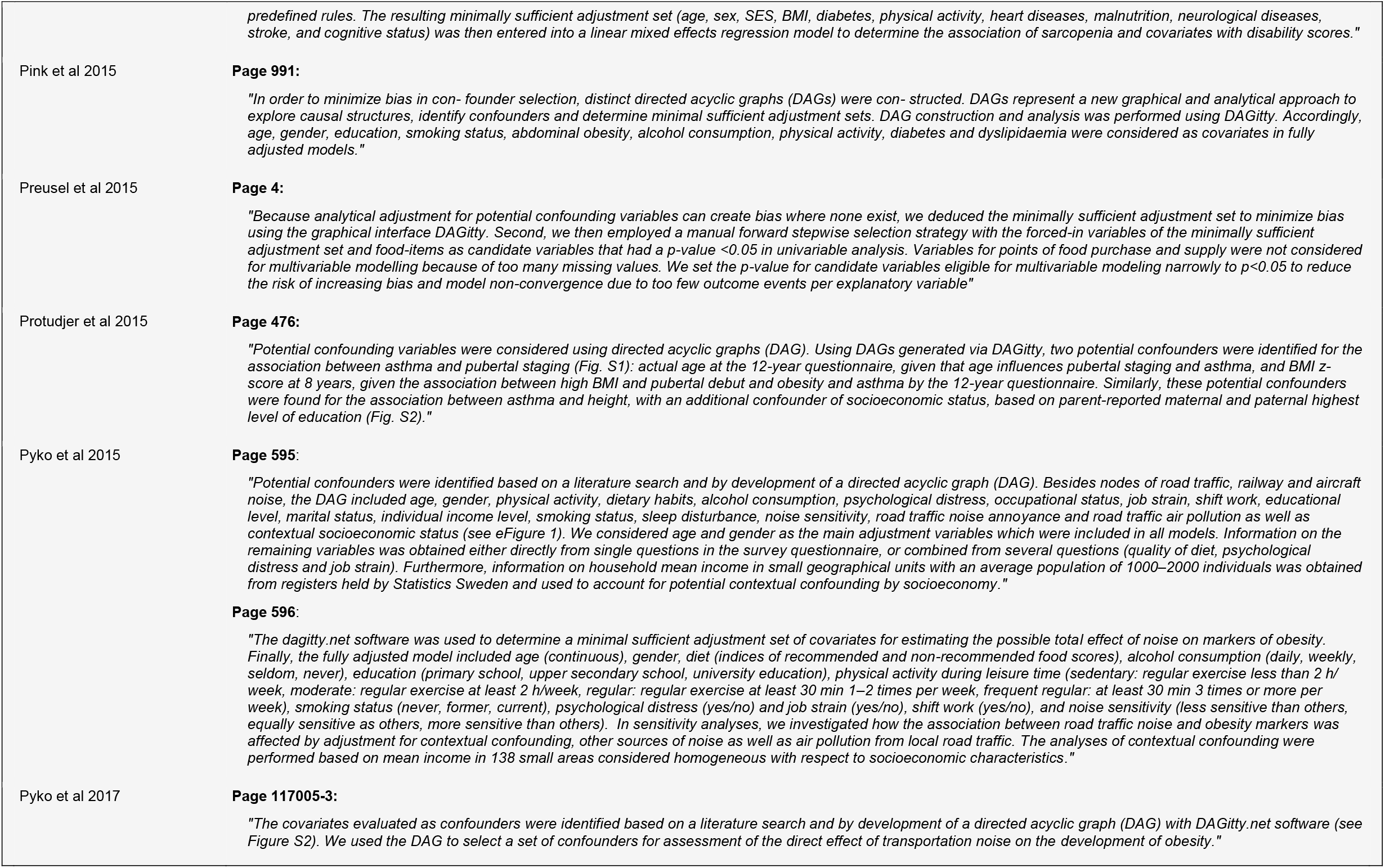
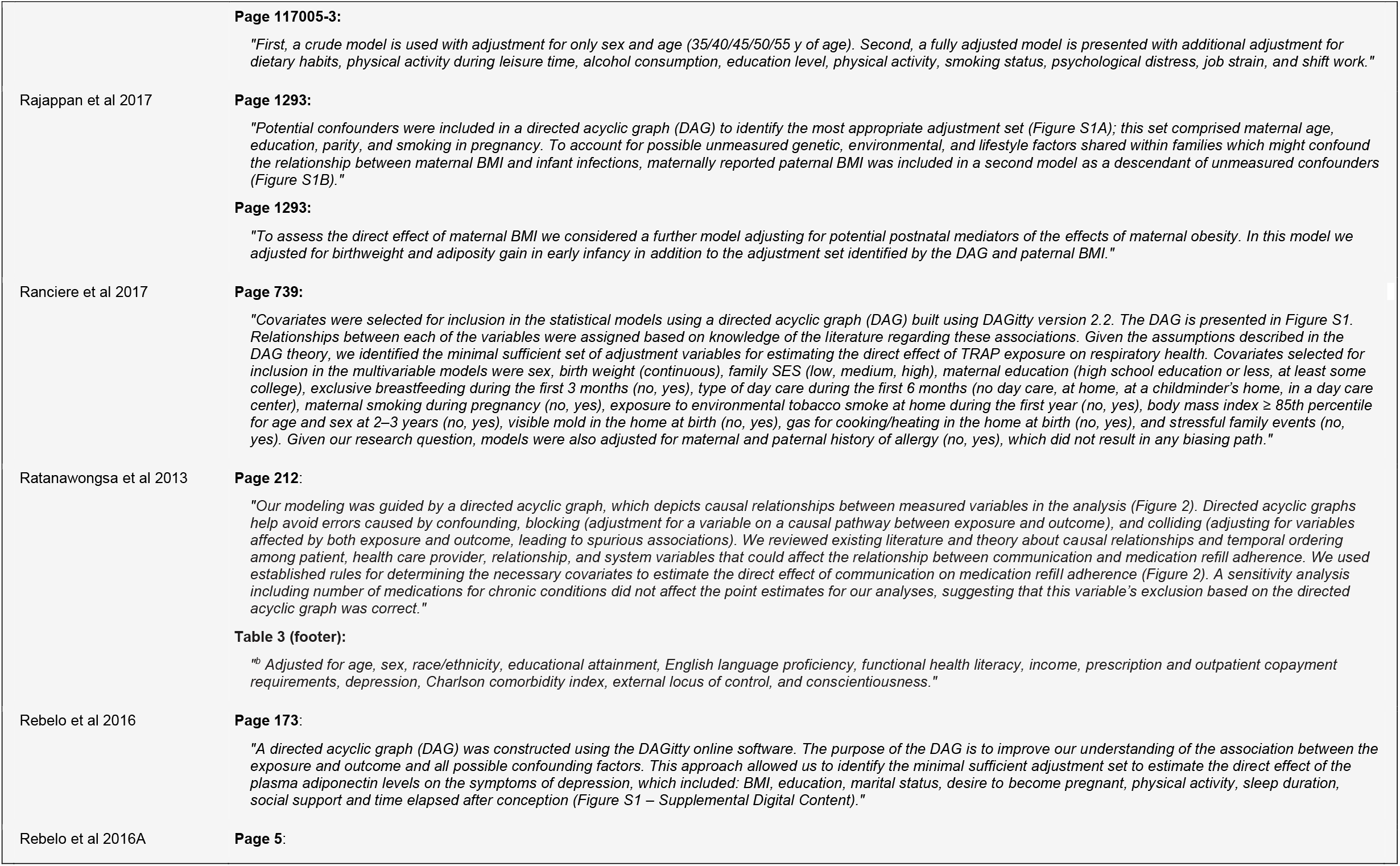
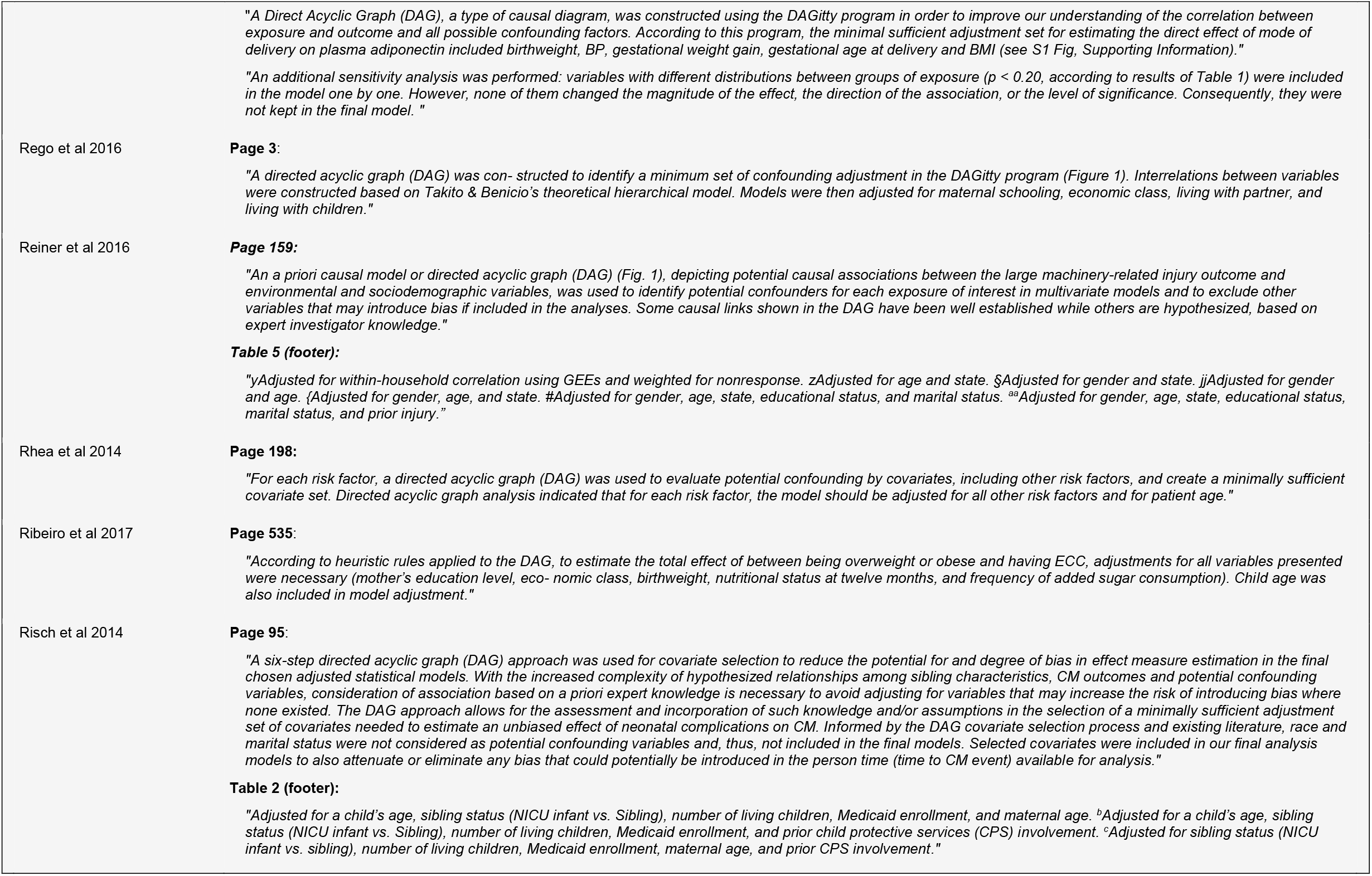
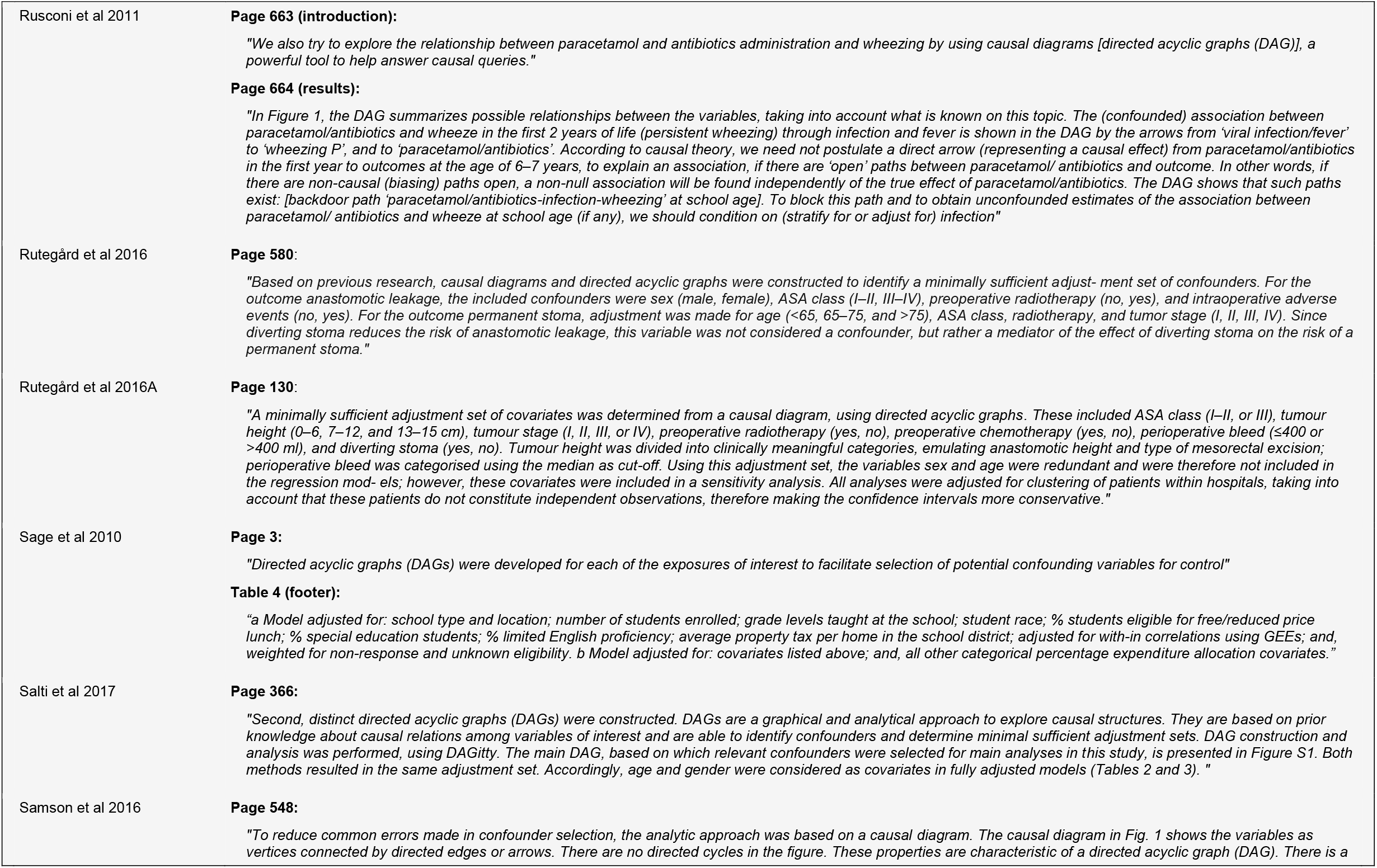
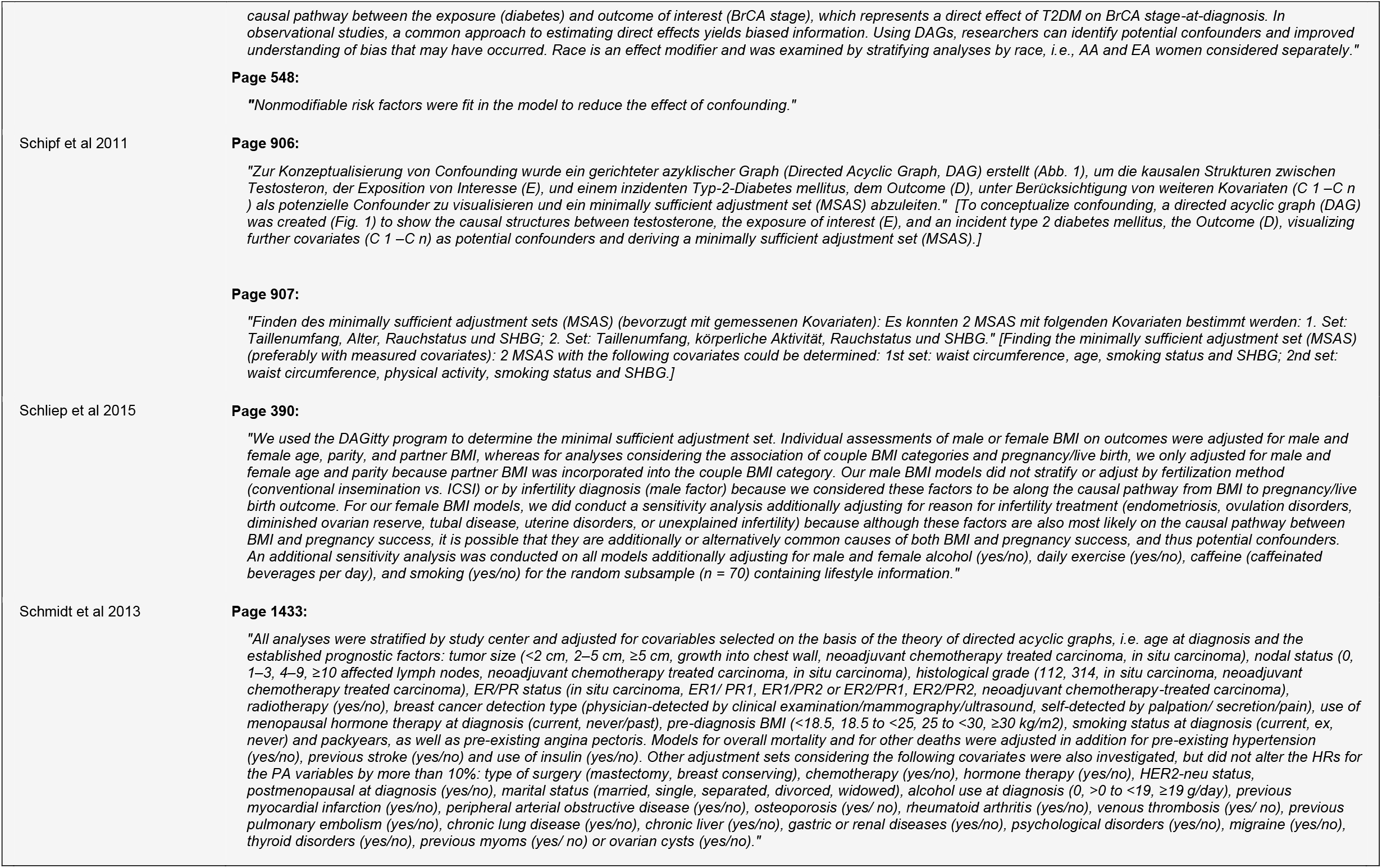
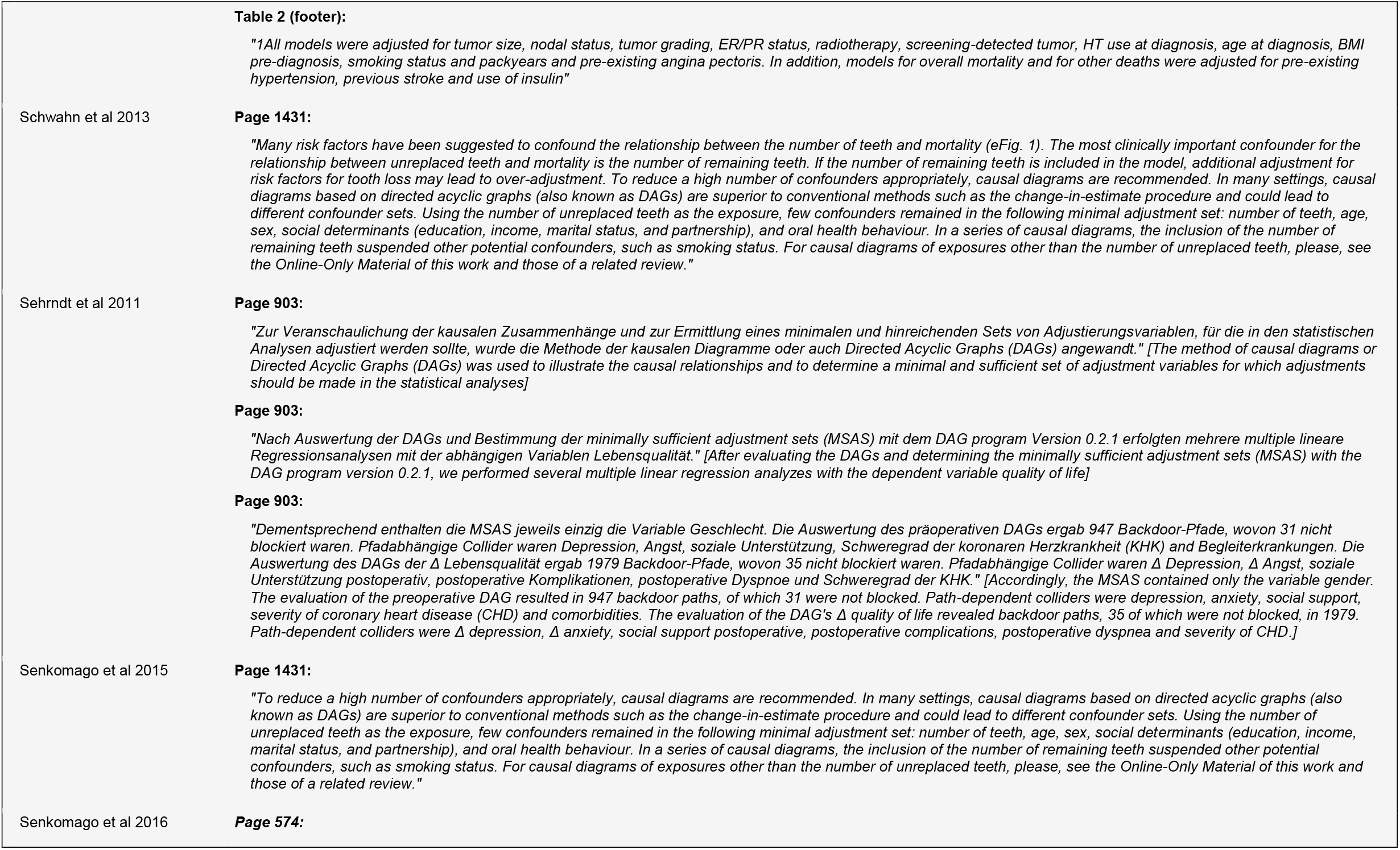
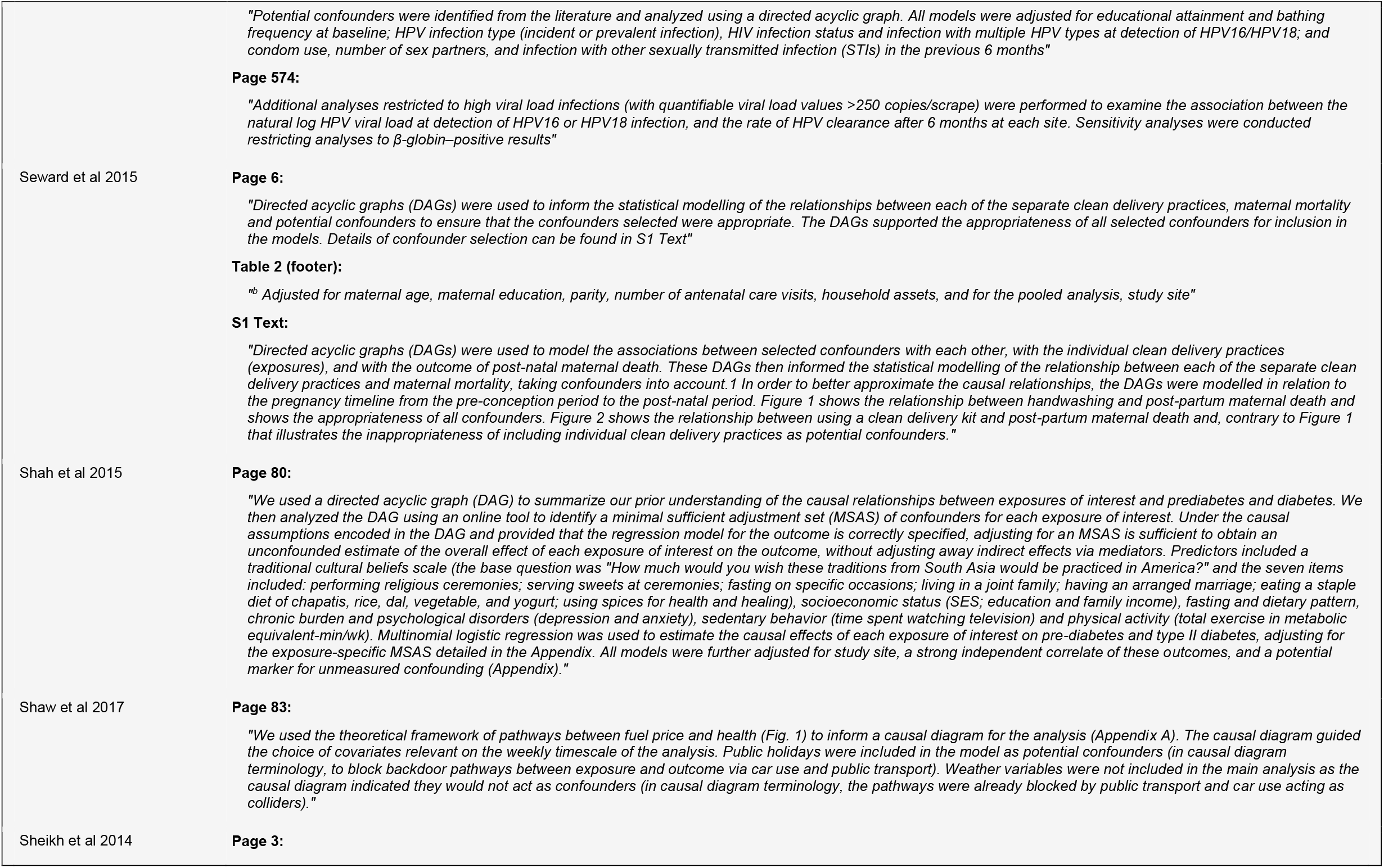
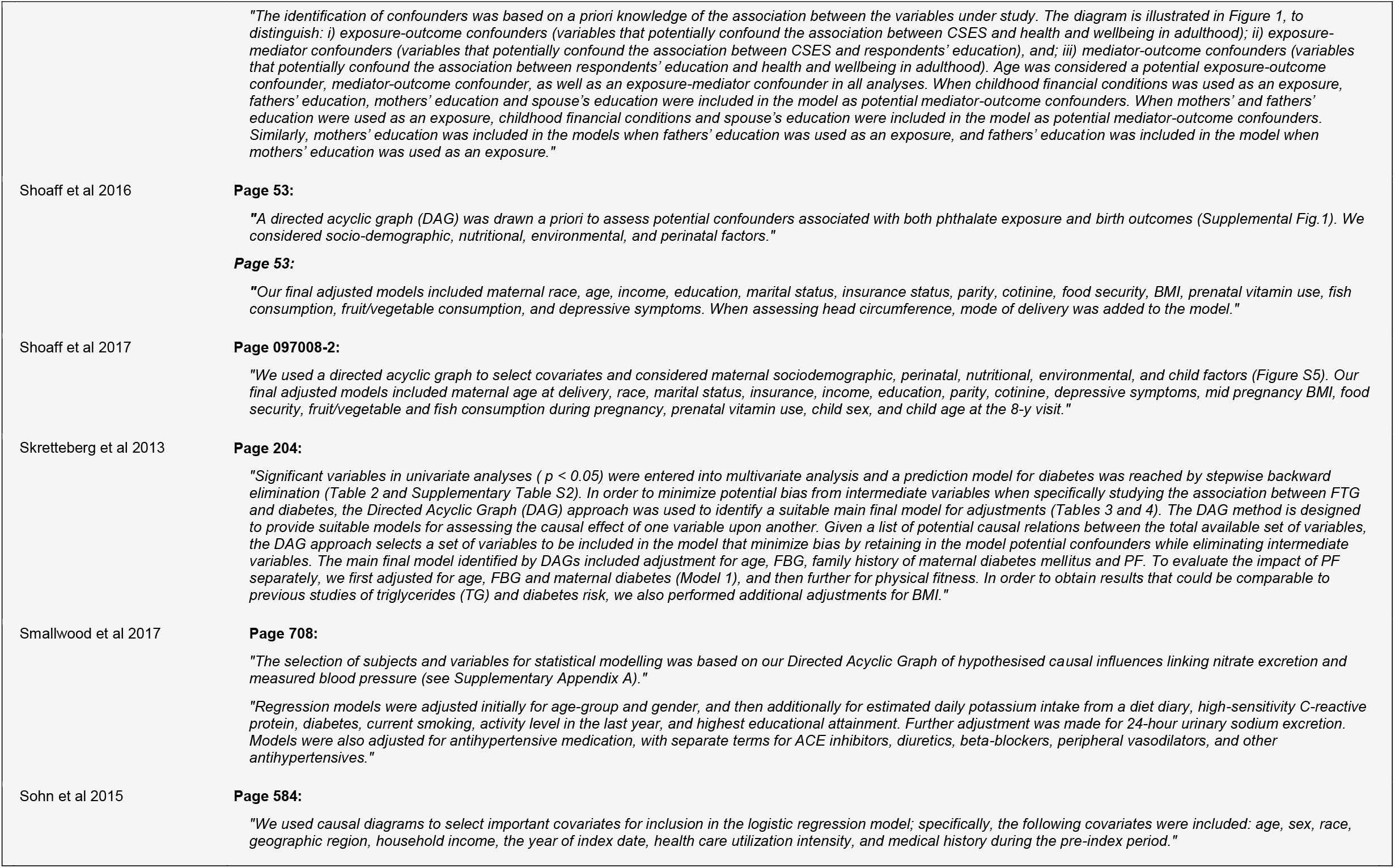
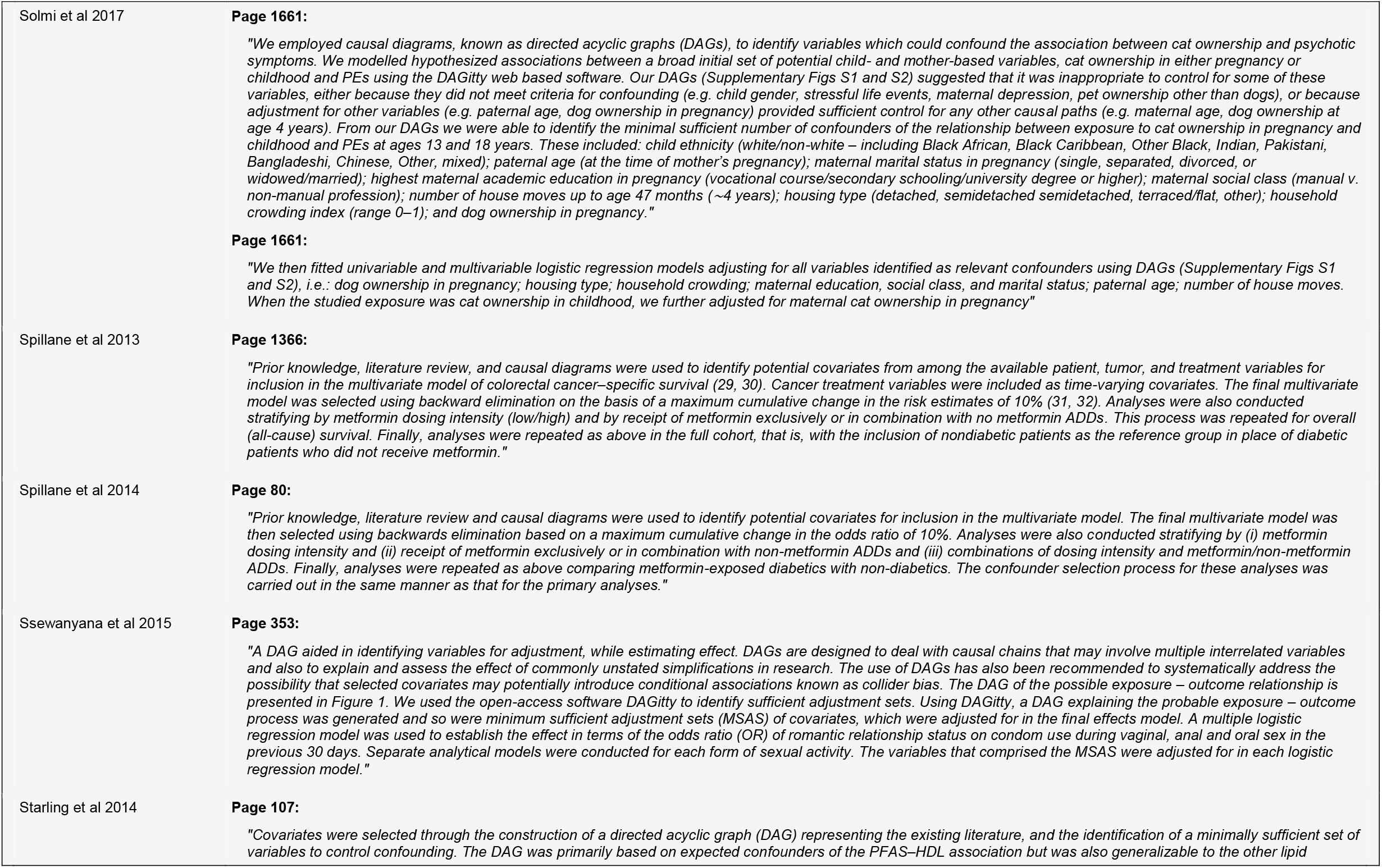
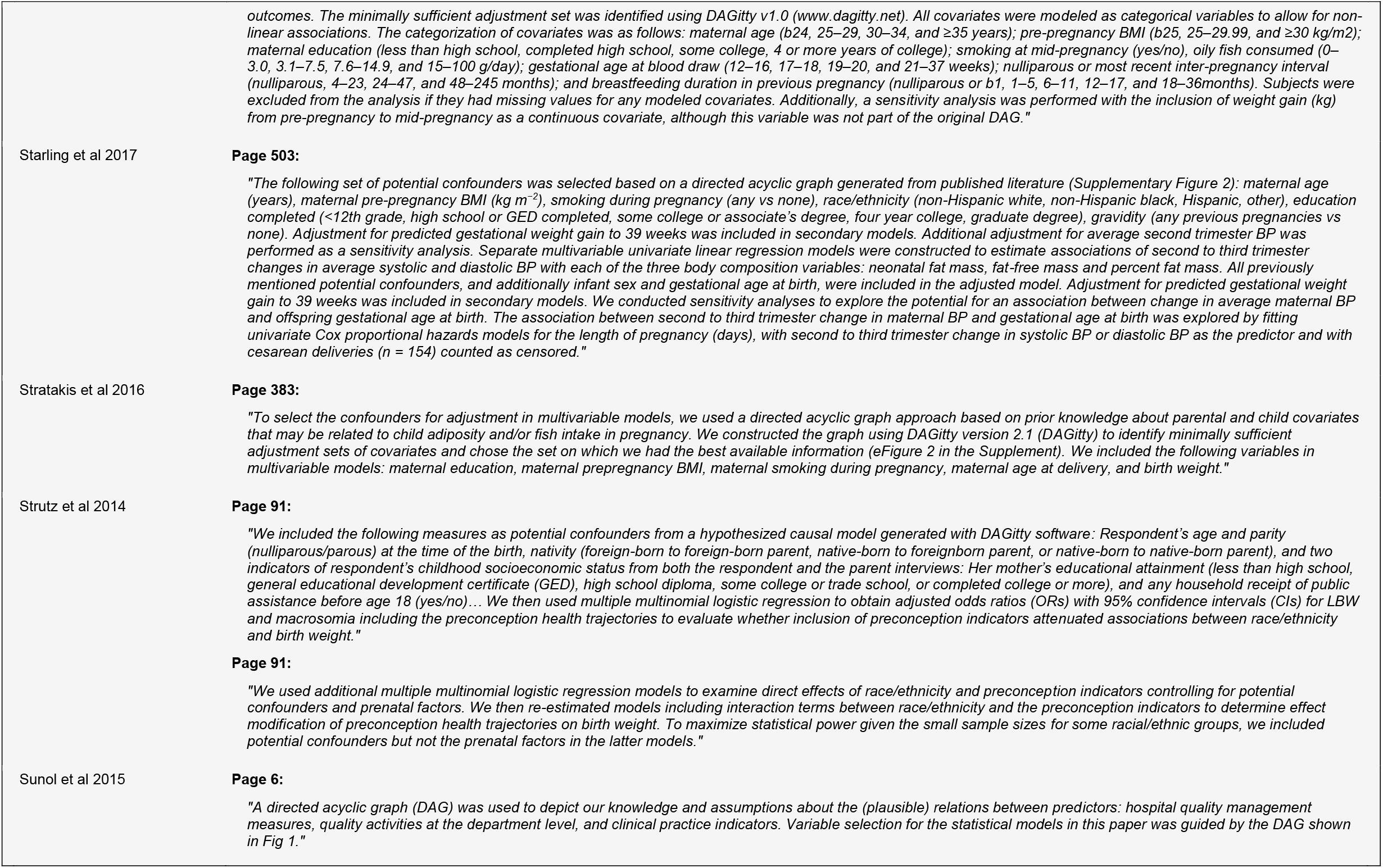
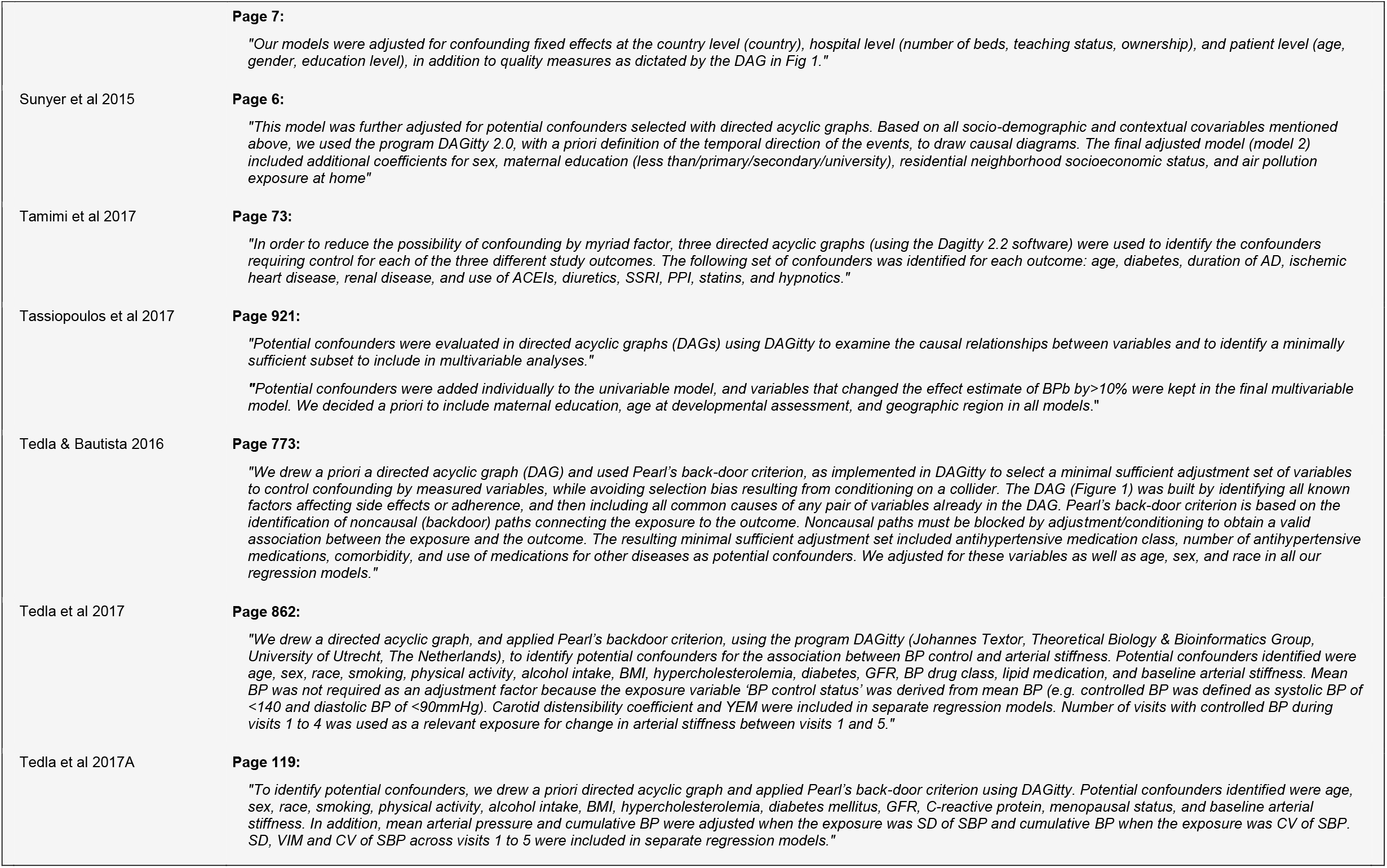
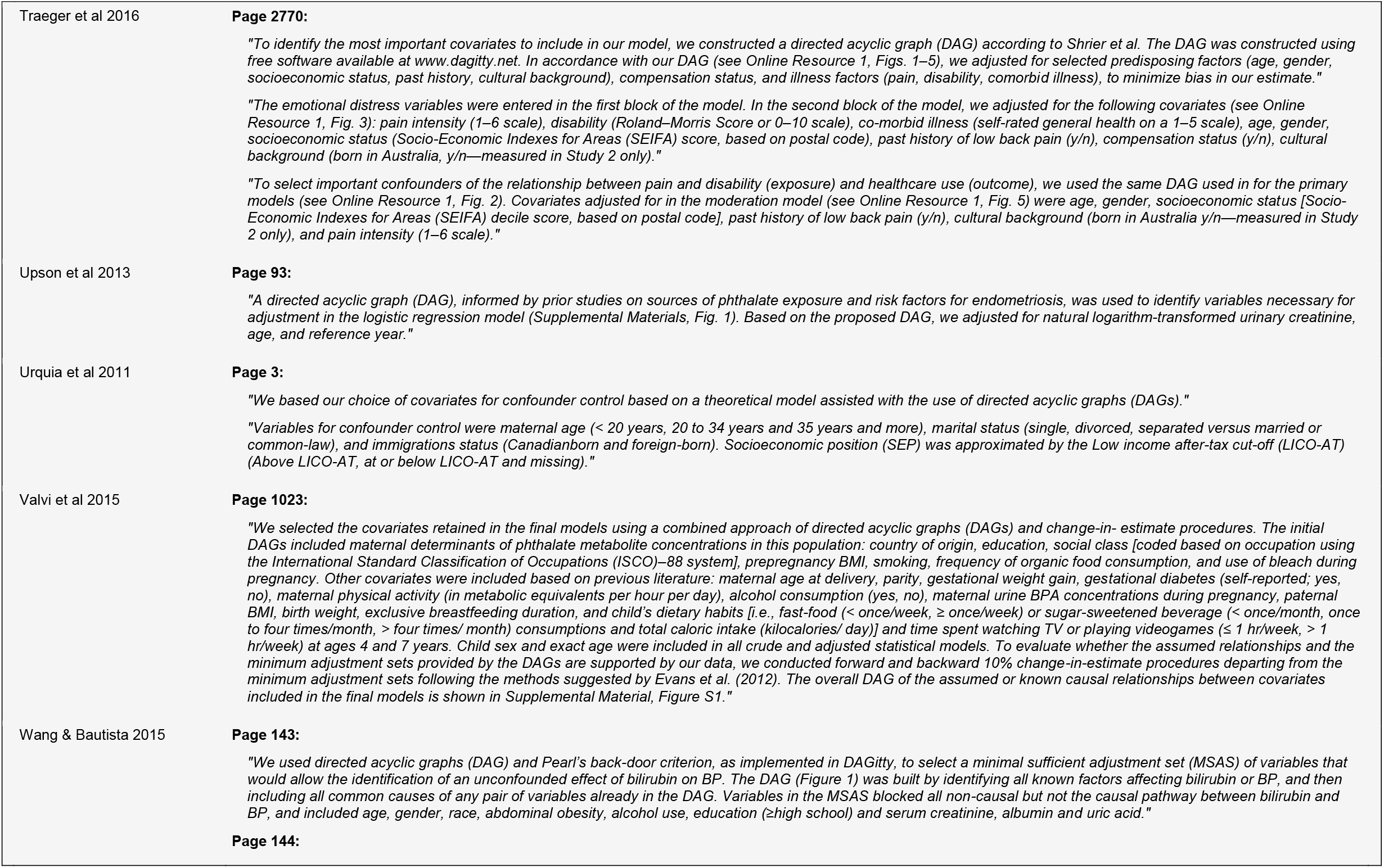
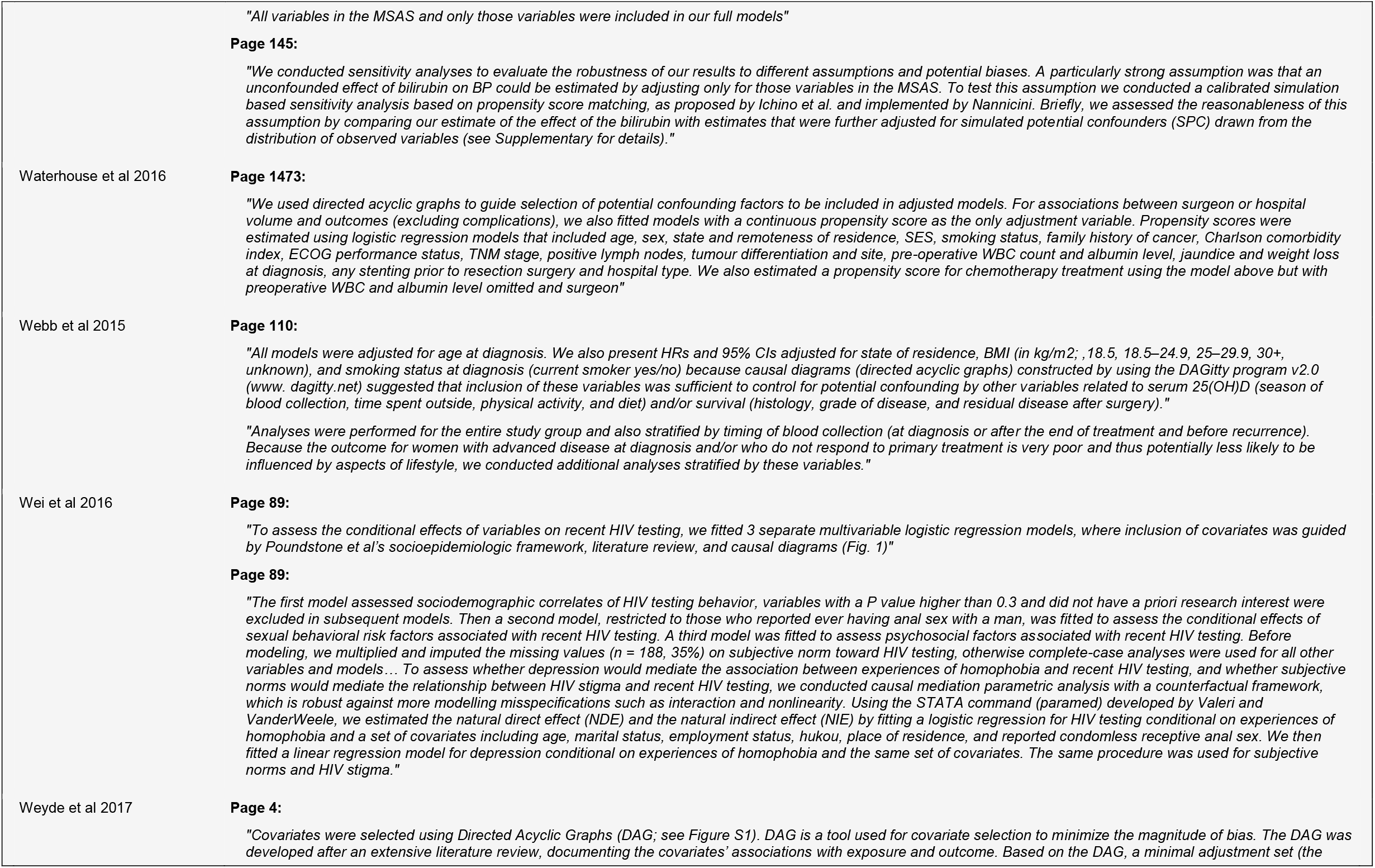
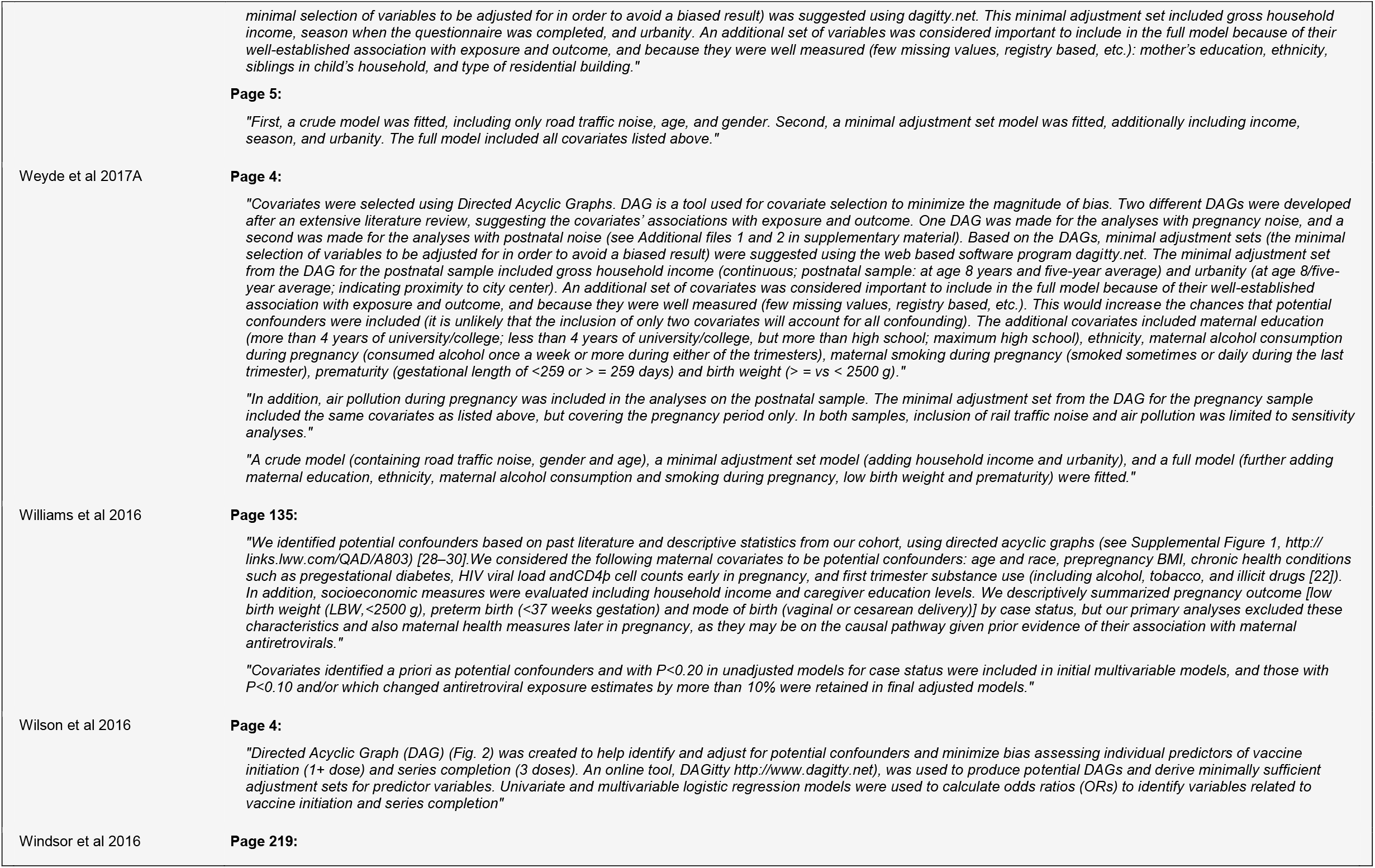
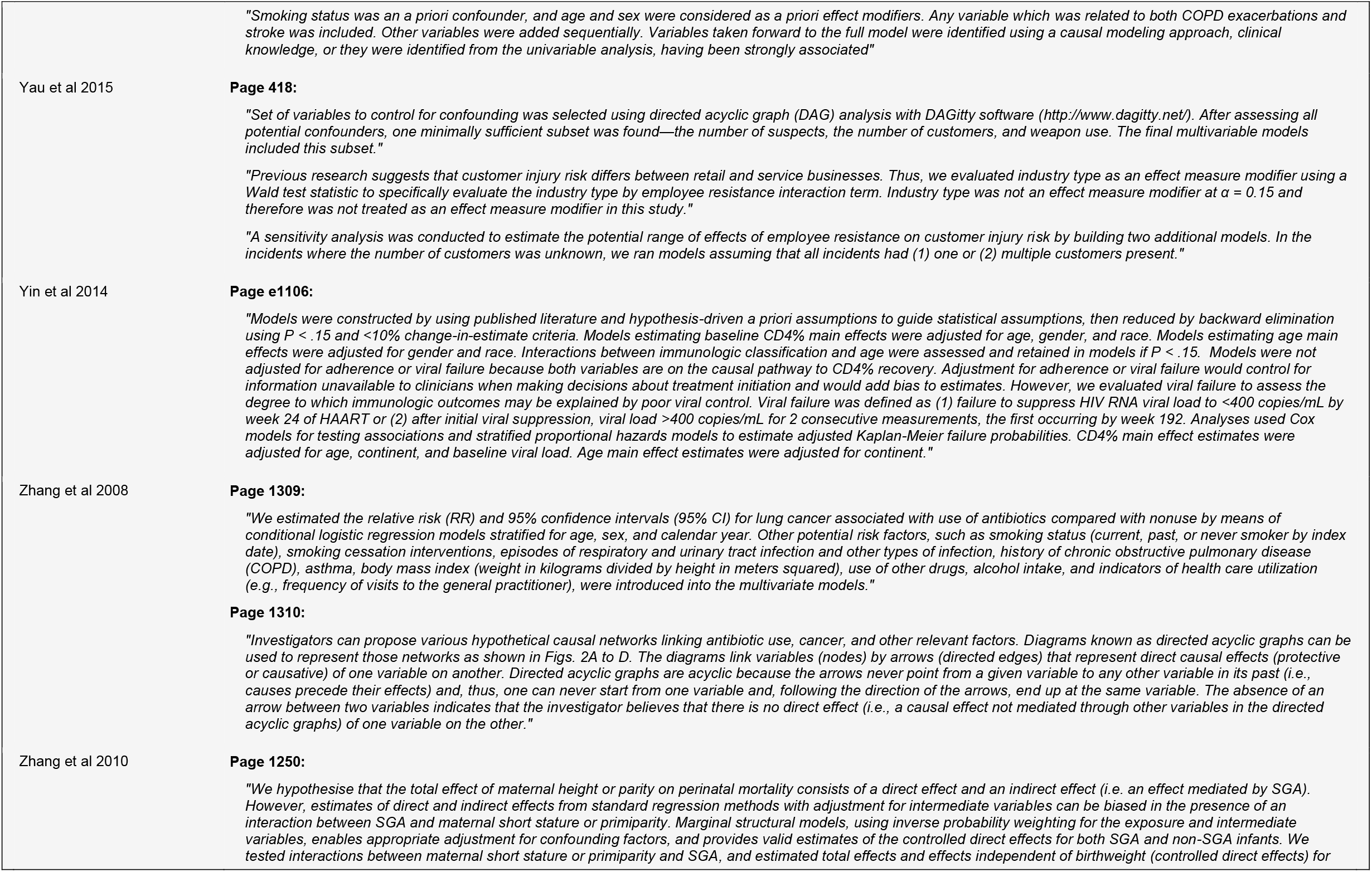
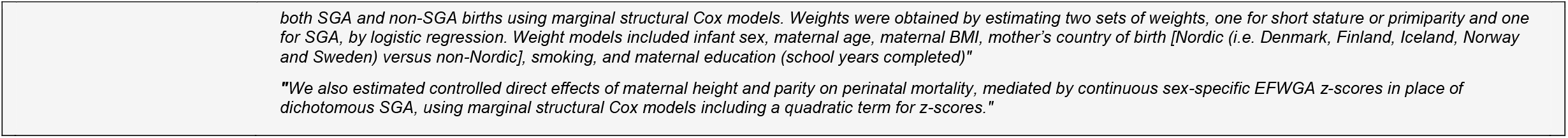

## Notes

### Competing Interest Statement

The authors have declared no competing interest.

